# Pharmacist-involved services for cardiometabolic conditions in China: a systematic review and meta-analysis

**DOI:** 10.1101/2024.07.25.24310734

**Authors:** Zhijie Deng, Fanglu Chen, Lin Gui, Yangjin Huang, Jiahui Chen, Yansheng Liu, Yueqiu Gong, Lu Zeng, Anhua Wei

## Abstract

**Background:** Pharmacist roles in chronic care are expanding, but clinical evidence is often separated from the implementation and cost data commissioners need. We assessed pharmacist-involved services for cardiometabolic conditions in China.

**Methods:** We searched PubMed, Embase, CENTRAL, CNKI, WanFang, and SinoMed from inception to July 20, 2026 for randomised trials comparing pharmacist-involved services with usual care in Chinese patients with cardiometabolic conditions (PROSPERO CRD42023414180). Chinese-language reports were eligible only from three core-index journal lists, defining evidence scope rather than quality. Two reviewers screened and extracted data independently. Random-effects models used restricted maximum likelihood with Hartung-Knapp inference; nine outcomes were rated with GRADE.

**Findings:** Of 22,609 records, 266 trials with 37,066 participants were included. Unplanned readmission (RR 0.52, 95% CI 0.46-0.60; I²=10.0%; 147 fewer per 1,000) and reported adverse drug reactions (RR 0.41, 0.37-0.45; 139 fewer per 1,000) both decreased, with low certainty; 15 of 22 readmission trials (72.6% of weight) involved heart failure, myocardial infarction, or coronary disease. Reported adherence increased (RR 1.37, 1.32-1.43; I²=80.2%; 255 more per 1,000; very low certainty), but only nine of 228 adherence-reporting trials used objective measures. Restricting to low-risk results left the four readmission results imprecise (RR 0.56, 0.30-1.05). No cost-reporting trial captured the service inputs needed to estimate net costs.

**Interpretation:** In patients admitted with heart failure or coronary disease, pharmacist involvement was associated with fewer unplanned readmissions, the finding best suited to prospective evaluation, although certainty was low. Evidence for the other outcomes was less directly applicable to decision making, and adherence gains reflect reported rather than verified medicine taking. Pilots should use fixed 30-day and 90-day endpoints, dispensing-based adherence measures, and prospective delivery-cost data.

**Funding National Natural Science Foundation of China (grant number 81803841).**

**Research in context:** *Evidence before this study:* Systematic reviews have reported improvements in adherence and selected clinical outcomes after pharmacist involvement, but conclusions differ by disease, service model, and endpoint. Economic reviews repeatedly note incomplete measurement of delivery costs and health consequences.

*Added value of this study:* Reviews of pharmacist services, including our own of medication therapy management, have reported favourable pooled effects alongside marked heterogeneity and have concluded that stronger evidence is needed. This review tested that conclusion against 266 randomised trials from a single health system, the largest such body assembled. Effects were larger, not smaller, yet no outcome reached moderate certainty: adherence was measured objectively in nine of 228 reporting trials, follow-up duration was unreported in 108 of 266, and none of the 13 trials reporting expenditure recorded the service inputs needed to cost delivery. Adding trials did not raise certainty; what constrained the evidence was how little of it was measured in a way that supports a decision. The audit of all 228 trials reporting any adherence information was intentionally broader than the quantitative syntheses, which included sufficiently comparable binary adherence rates and a separate MMAS-8 analysis. The non-core validation supported the directions of the principal outcomes; detailed eligibility, contribution, and effect estimates are provided in the Supplementary Material.

*Implications of all the available evidence:* For practice, the evidence points most clearly to patients admitted with heart failure or coronary disease, where pharmacist involvement was associated with fewer unplanned readmissions on an objectively ascertainable endpoint. Reported adherence should be treated as a measurement outcome rather than a direct measure of observed medicine taking, and the pooled estimates should not be assumed to transfer across settings. For research, the main limitation is measurement quality rather than a shortage of trials: objective adherence measurement, fixed 30-day and 90-day endpoints, complete follow-up reporting, and prospective collection of delivery inputs and costs would do more for decision-making than further effectiveness trials of the same design.

## Introduction

Health systems are broadening pharmacists’ work from medicine supply to medication optimisation, chronic-care management, and coordination between care settings.^1^ This shift responds to multimorbidity and medication-related harm, but clinical roles also depend on regulation, financing, information access, and collaboration.^2^ What a pharmacist is permitted to do, and who pays for it, therefore varies more than the shared vocabulary of pharmaceutical care suggests.

The clinical literature is substantial but uneven. Reviews suggest benefits for adherence, cardiovascular risk factors, and selected patient outcomes, whereas mortality and healthcare use are less consistent.^3–7^ Service labels cover different activities and levels of clinical authority, and payment arrangements remain fragmented.^8^ Two problems follow from this. Pooled estimates rest on outcomes that trials measure in very different ways, so a single summary figure can conceal what was actually observed. And the information a payer needs, namely what the service consumes and what it costs to deliver, is rarely collected alongside the clinical result.

China combines a large cardiovascular and diabetes burden with rapid expansion of clinical pharmacy.^9,10^ National policy promotes patient-facing services, but provision and professional authority vary, while diagnosis-related group (DRG) and diagnosis-intervention packet (DIP) reform increases pressure to demonstrate value.^11–13^ Payment rules are being set while this evidence base is still being assembled. Effectiveness evidence therefore has to align with service definitions, implementation requirements, and payment design.

We had reached a version of this question before: our earlier synthesis of medication therapy management services in the international literature found favourable clinical effects alongside marked heterogeneity and inconclusive economic outcomes, and concluded that stronger evidence was needed.^14^ This review asks what happens when that evidence arrives. This evidence base is relevant beyond China because many health systems face the same transition from expanded pharmacist roles to reproducible delivery and sustainable payment. We assessed the clinical effects of pharmacist-involved services and examined which outcomes were sufficiently measurable, consistent, and clinically interpretable to inform implementation and payment decisions. Prediction intervals, the Grading of Recommendations Assessment, Development and Evaluation (GRADE) approach, and moderator analyses were used to separate average effects from evidence carrying greater credibility and applicability. The standard applied throughout is decision relevance.

## Methods

### Protocol and reporting

We conducted this systematic review and meta-analysis in accordance with the Preferred Reporting Items for Systematic Reviews and Meta-Analyses (PRISMA) 2020 statement.^15^ The review was registered in PROSPERO on April 14, 2023 (CRD42023414180). Three principal amendments bear on interpretation. A Chinese-language journal-scope criterion based on the union of three core-index lists was introduced during conduct of the review to define the primary evidence scope. The analytical framework was expanded to 21 outcomes, adding prediction intervals, the GRADE approach, risk of bias due to missing evidence (ROB-ME) assessment, follow-up and disease-spectrum analyses, and INSPECT-SR. The registered protocol had specified excluding trials at high risk of bias; we retained them in the primary synthesis and addressed risk of bias through outcome-specific low-risk sensitivity analyses instead, which preserves the full evidence base. These decisions preceded final data lock, but they were entered in the register on August 9-10, 2026, after the search closed on July 20, 2026. Each amendment and its inferential status is listed in Supplementary Table S16. The sampled non-core-journal validation and its leave-one-out checks were conducted after final data lock and are labelled post hoc or exploratory. We therefore present outcome-specific syntheses as a structured evidence map rather than a single confirmatory test.

### Search strategy and eligibility

We searched PubMed, Embase, the Cochrane Central Register of Controlled Trials (CENTRAL), the China National Knowledge Infrastructure (CNKI), WanFang, and SinoMed from inception to July 20, 2026. The database-specific strategies combined pharmaceutical-service, cardiometabolic-disease, and China-region concepts; no study-design or other database filter was appended. Chinese-language reports were restricted to journals included in the union of three core-index lists (Supplementary Table S14A). This journal-scope criterion was introduced during conduct of the review to define the primary evidence scope and was not used as a proxy for trial quality. The locked analytic dataset and PRISMA counts derive from the final search run completed through July 20, 2026; Appendix 1 reproduces the database-specific strategies used in that search.

After duplicate removal, journal titles of Chinese-language reports were matched against the union of three core-index lists used to define the Chinese-language evidence scope. Titles were normalised for punctuation, spacing, and common variants, and unresolved matches were checked manually. Trial quality was assessed separately at the trial level. Two reviewers then screened all remaining records and made every full-text decision.

Eligible reports were randomised controlled trials in the Chinese healthcare system among patients with the prespecified cardiometabolic conditions. Interventions were structured, patient-facing pharmacist services, and comparators were usual care or care without the structured service. Head-to-head services and pharmacist co-interventions in both arms were excluded.

A sampled post-data-lock validation of non-core-journal reports used the same population, intervention, comparator, and randomised-study-design criteria as the primary review. Reports contributing at least one validation outcome were analysed using the primary effect measures and random-effects model, with core-versus-non-core differences estimated using a study-level subgroup indicator. The 8-item Morisky Medication Adherence Scale (MMAS-8) and mortality were summarised separately, and follow-up availability was recorded. INSPECT-SR was not reapplied because the validation assessed directional robustness rather than the full trustworthiness framework.

### Study selection, extraction, and classification

Two reviewers independently screened records and extracted data with a piloted form, resolving disagreements by discussion and third-reviewer adjudication. Reports were checked for overlap and linked at trial level, so the dataset contained no shared control groups and no repeated time points or subgroups for the same outcome within a trial. When more than one eligible follow-up assessment was reported for a continuous outcome, the final reported follow-up value was selected. The locked analytical datasets contained no missing outcome values requiring imputation. Extracted fields are listed in the Supplementary Material.

Each trial received one principal disease category, and we kept the author-assigned service label as a descriptor rather than a description of content. Follow-up was converted to months and left missing when unreported. Province names were standardised and mainland provinces grouped into four regions using the National Bureau of Statistics classification.^16^ Coding rules are in the Supplementary Material.

### Outcomes and intervention reporting

Outcomes comprised reported adherence rate, MMAS-8, satisfaction, adverse drug reactions, unplanned readmission, mortality, disease-control measures, length of stay, and reported expenditures. The adherence measurement audit covered every trial reporting any adherence information and was intentionally broader than the quantitative syntheses. Sufficiently comparable binary adherence rates and MMAS-8 scores were pooled separately; bespoke continuous scores with incompatible ranges, definitions, or directions were retained in the evidence map but not quantitatively combined. Measurement was classified as named self-report, investigator-developed, dispensing-based objective, physiological-marker inference, or not reported.^17^ Each trial contributed at most one estimate to any single outcome; a trial reporting both a binary adherence rate and an MMAS-8 score contributed to two different outcomes rather than twice to one. No included trial randomised more than two groups.

Intervention reports were coded for eight activities, each classified as explicitly reported or not reported, with the latter treated as unknown rather than absent; the reporting-status analysis was therefore exploratory.^18^

### Risk of bias and certainty

Two reviewers independently appraised design and reporting features across all 266 included trials using the five domains of the Cochrane risk-of-bias tool for randomised trials, version 2 (RoB 2), with third-reviewer adjudication.^19^ This appraisal preceded and was independent of any outcome result, and identified 60 trials at low risk of bias overall. Outcome-specific RoB 2 was then completed for every result that these 60 trials contributed to the three core outcomes, giving 52 results (28 for reported adherence, 20 for reported adverse drug reactions, and four for readmission); all 52 were confirmed low risk across the five domains and overall. This subset was used to stress-test the principal findings. GRADE was applied to nine outcomes selected to represent the principal medication-use, patient-important, disease-control, and resource-use domains, considering risk of bias, inconsistency, indirectness, imprecision, and publication bias.^20^

### Trial trustworthiness screening

Two reviewers independently applied all 21 checks in the INSPECT-SR trustworthiness tool to trials contributing the three principal outcomes; unclear items remained unresolved and serious concerns required strong evidence of an integrity problem.^21^

### Outcome-specific and robustness analyses

For unplanned readmission we grouped reported follow-up as 3 months or less, more than 3 to 6 months, and more than 6 months. The prespecified primary moderator was the continuous change per 6 months, reported without adjustment; the window-based omnibus comparison was exploratory. Follow-up duration was unavailable for 1 of the 22 estimates, which did not enter these models. We also stratified readmission trials as heart failure, myocardial infarction, or coronary disease; diabetes or hypertension; and mixed cardiometabolic conditions.

We used ROB-ME to structure the assessment of bias due to missing evidence for readmission, considering how many eligible trials did not contribute the outcome, whether protocols were available, whether non-reporting appeared related to result direction, and small-study-effect evidence. Non-reporting alone was not treated as evidence that a measured result had been suppressed.^22^

For blood pressure, glucose, glycated haemoglobin, and lipids we recalculated each mean difference and variance from group summaries and identified influential estimates using externally studentised residuals, Cook’s distance, and Baujat contributions. For reported adherence, the RR was primary, with the OR and risk difference as effect-scale sensitivities.

### Statistical analysis

Binary outcomes were analysed as RRs, with ORs used for selected exploratory outcomes and sensitivity analyses. Continuous outcomes were mean differences (MDs) after unit harmonisation. Random-effects models used restricted maximum likelihood (REML) and Hartung-Knapp 95% confidence intervals (CIs), with τ², Q, I², and prediction intervals reported.^19,23^ A 0.5 continuity correction was applied only to trials with a zero cell, and trials with no events in either arm were removed in a prespecified sensitivity analysis (Supplementary Table S5).^19^

We applied pooled RRs for reported adverse drug reactions, readmission, and mortality to the median control risk, taking the number needed to treat for benefit (NNTB) as the reciprocal of the absolute risk reduction. Because NNTBs for event outcomes depend on baseline risk and follow-up, the full scenario set is reported in Supplementary Table S7.^19,24^

Exploratory meta-regression examined disease, service label, mean age, publication year, follow-up, region, and intervention reporting, with cluster-robust variance estimation (CR2) and Benjamini-Hochberg control of the false discovery rate (FDR) within each moderator family. Small-study effects were assessed with Peters’ test for binary outcomes and Egger regression for continuous outcomes when at least ten estimates were available. Full moderator, sensitivity, and small-study results are reported in Supplementary Tables S2A, S21A, and S21B.

Low-risk and other results were compared formally in an interaction model. Because this restriction sharply reduces precision, especially for readmission, the restricted estimates were interpreted alongside the full evidence base rather than as a pass-fail test.

### Expenditure analysis

Hospitalisation and medication expenditures were treated as unpaired direct-cost components, not an economic evaluation. Amounts were standardised with the national healthcare consumer price index (CPI), with overall CPI as a sensitivity analysis.^25^ Completeness was assessed according to the Consolidated Health Economic Evaluation Reporting Standards (CHEERS) 2022, covering perspective, horizon, delivery inputs, price year, incremental costs, consequences, and uncertainty; service-delivery costs were treated as missing rather than zero.^26^ The annual indices and official source pages used for standardisation are reported in Supplementary Table S17B.^27^

Analyses used R 4.5.2 with metafor, meta, and clubSandwich; package versions are given in the Supplementary Material.

## Results

The searches identified 22,609 records. After removing 4,371 duplicates and 13,694 non-core-index records, 4,544 titles and abstracts were screened. We assessed 648 full texts and excluded 382, leaving 266 trials with 37,066 participants and 1,107 estimates across 21 outcomes (Figure 1; Table 1; Supplementary Table S8A).

**Figure 1.**
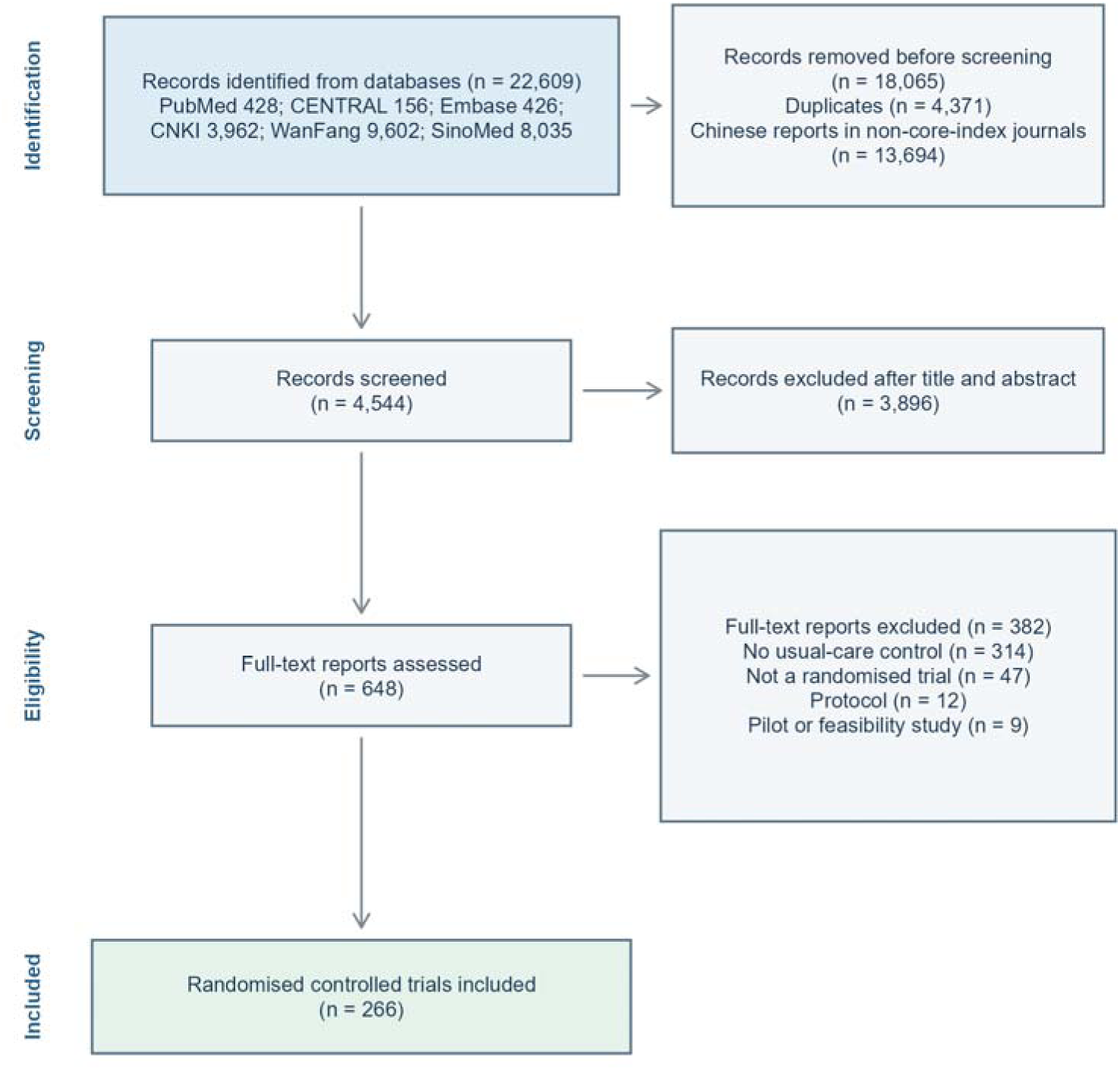
Study-selection flow. After duplicate removal, journal titles of Chinese-language reports were matched against the union of three core-index lists used to define the Chinese-language evidence scope. The remaining records underwent independent title-and-abstract and full-text screening.

**Table 1.**
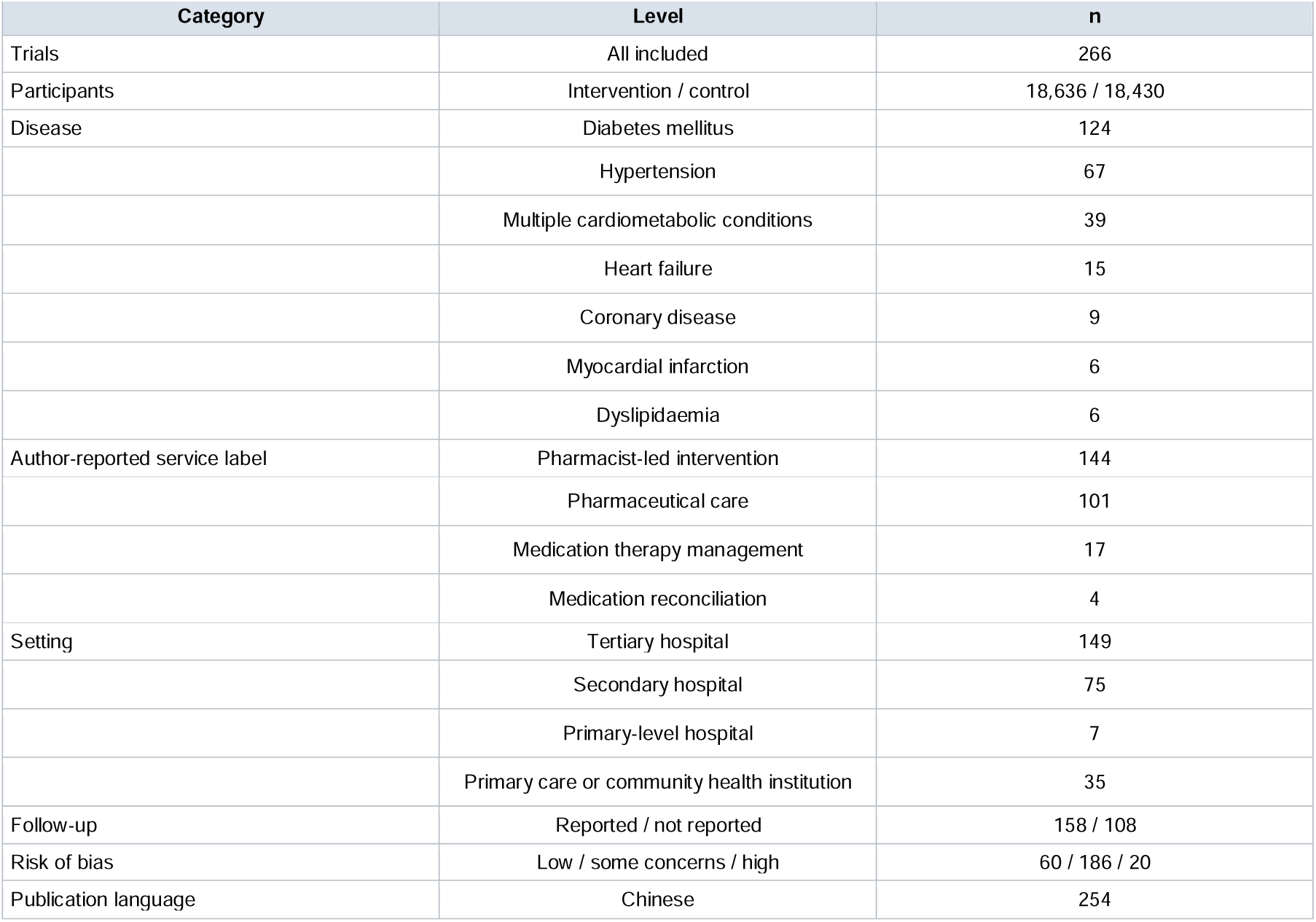
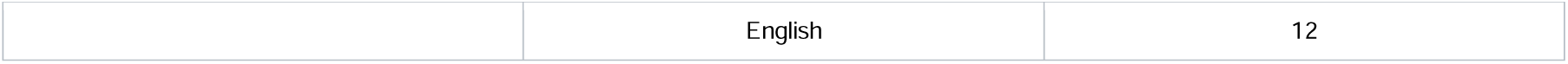
Study characteristics.

The evidence base comprised 124 diabetes, 67 hypertension, 39 mixed-condition, 15 heart-failure, nine coronary-disease, six myocardial-infarction, and six dyslipidaemia trials; 254 reports were in Chinese and 12 in English. Authors labelled 144 services pharmacist-led intervention, 101 pharmaceutical care, 17 medication therapy management, and four medication reconciliation. Risk of bias was low in 60 trials, raised some concerns in 186, and was high in 20 (Supplementary Tables S8A, S10A, S10B, and S14B). Care settings were 149 tertiary hospitals, 75 secondary, seven primary-level, and 35 primary care or community institutions, with hospital grade taken as at the publication year (Table 1; Supplementary Table S8B).

Adherence information was reported in 228 of 266 trials. The evidence-map audit classified 115 as using named self-report instruments, 104 as using investigator-developed definitions, and nine as using objective measures; it covered all trials reporting adherence information, not only the 163 binary estimates. The quantitatively comparable adherence-rate synthesis was directionally favourable (RR 1.37, 95% CI 1.32-1.43; I²=80.2%; prediction interval 0.98-1.93; very low certainty). The reduction in reported adverse drug reactions was the most statistically consistent principal finding (RR 0.41, 95% CI 0.37-0.45; low certainty), whereas readmission was comparatively consistent but concentrated in high-risk cardiovascular populations (RR 0.52, 95% CI 0.46-0.60; low certainty) (Figures 2-3; Tables 2-3; Supplementary Table S13B; Supplementary Figures S5-S13).

**Figure 2.**
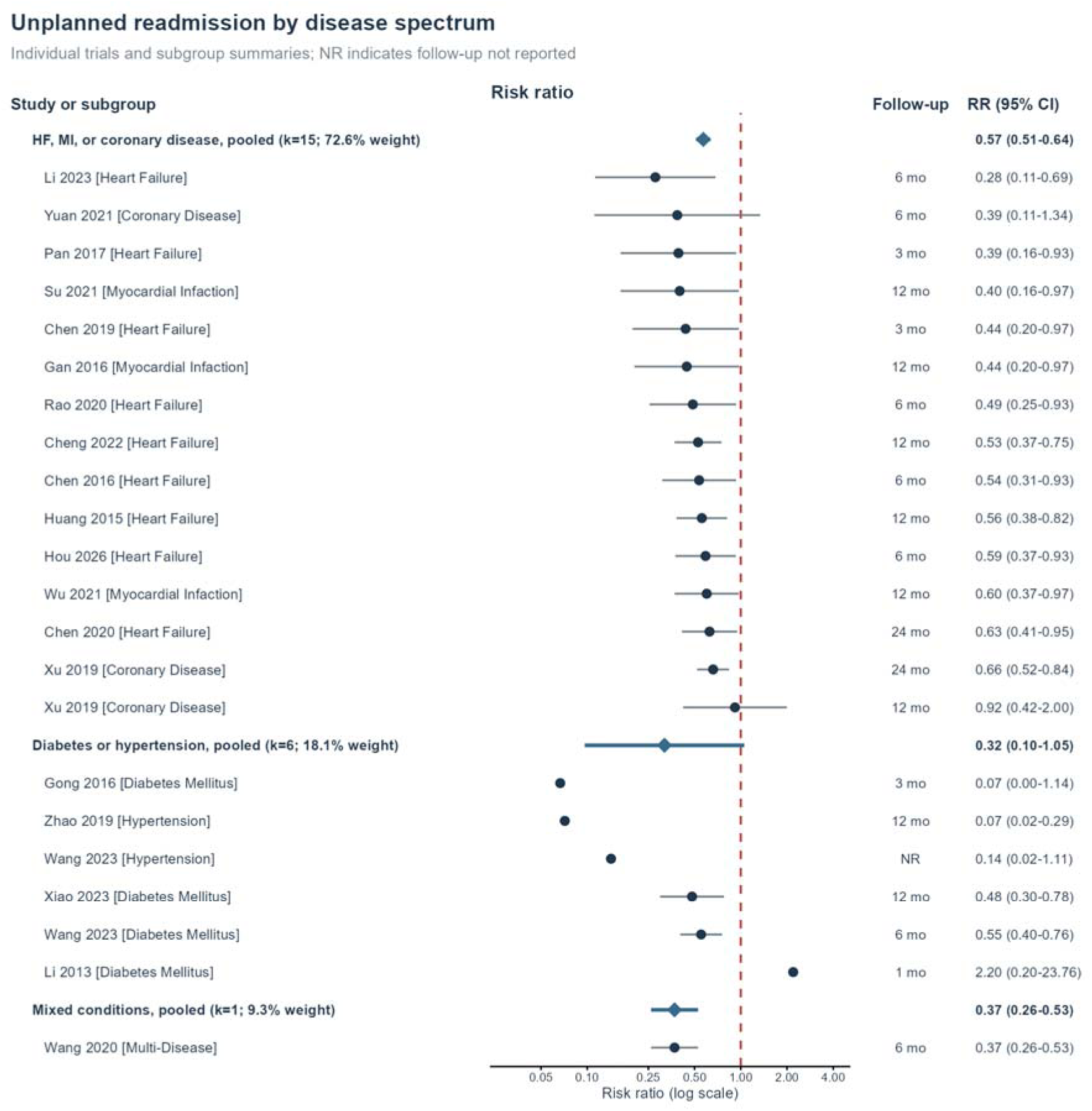
Individual-trial and subgroup risk ratios for unplanned readmission by disease spectrum. Diamonds show subgroup pooled estimates; study and subgroup columns report follow-up, effect estimates, subgroup k, and random-effects weight. The red dashed line is the null, and NR denotes follow-up not reported.

**Figure 3.**
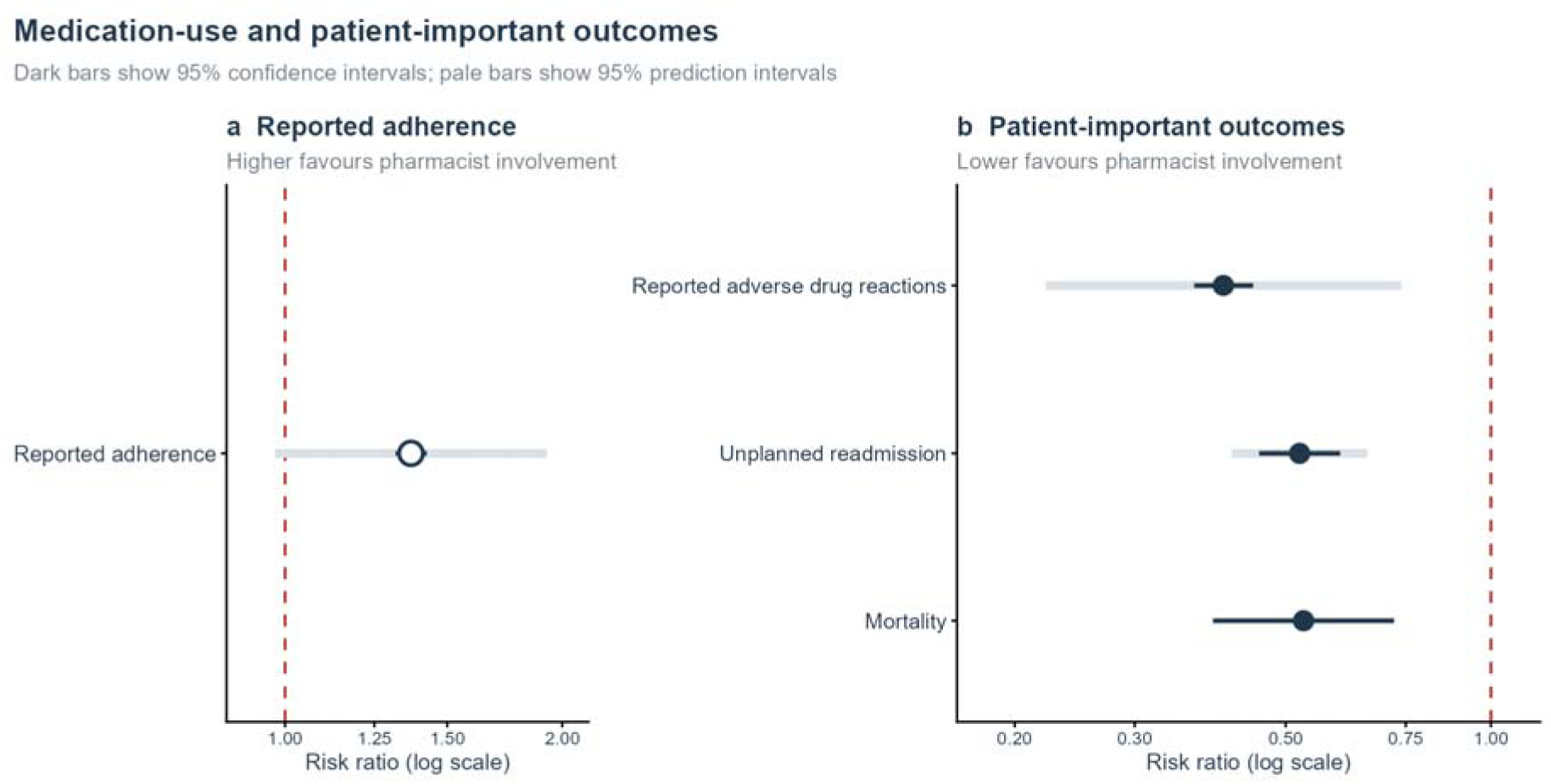
Medication-use and patient-important outcomes. Panel A shows reported adherence, for which higher values favour pharmacist involvement. Panel B shows adverse drug reactions, unplanned readmission, and mortality, for which lower values favour pharmacist involvement. Dark bars show 95% confidence intervals and pale bars show 95% prediction intervals where robustly estimable. Open symbols denote very-low-certainty evidence and filled symbols low-certainty evidence; the mortality prediction interval was not robustly estimable.

**Table 2.** Main meta-analysis results grouped by evidence certainty.

| Outcome | k | Pooled effect (95% CI) | 95% prediction interval | I<sup>2</sup> (%) |
| --- | --- | --- | --- | --- |
| **Low-certainty patient-important outcomes** |  |  |  |  |
| Reported adverse drug reactions | 94 | RR 0.41 (0.37 to 0.45) | 0.22 to 0.74 | 38.3 |
| Unplanned readmission* | 22 | RR 0.52 (0.46 to 0.60) | 0.42 to 0.66 | 10.0 |
| Mortality | 9 | RR 0.53 (0.39 to 0.72) | Not estimable | 0.0 |
| **Very-low-certainty outcomes** |  |  |  |  |
| Reported adherence rate | 163 | RR 1.37 (1.32 to 1.43) | 0.98 to 1.93 | 80.2 |
| Length of stay | 13 | MD -1.77 (-2.96 to -0.58) | -5.71 to 2.17 | 92.6 |
| Systolic blood pressure | 91 | MD -8.83 (-10.08 to -7.58) | -20.27 to 2.61 | 97.7 |
| Glycated haemoglobin | 110 | MD -1.09 (-1.24 to -0.93) | -2.63 to 0.45 | 97.9 |
| Hospitalisation expenditure | 9 | MD -1258 (-2011 to -504) | -3293 to 778 | 80.2 |
| Medication expenditure | 12 | MD -1519 (-2699 to -339) | -5325 to 2288 | 99.3 |
| **Disease-control outcomes not formally graded. Pooled estimates are descriptive averages; prediction intervals are wide and heterogeneity is extreme.** |  |  |  |  |
| Diastolic blood pressure | 87 | MD -6.62 (-7.47 to -5.76) | -14.24 to 1.01 | 97.2 |
| Fasting plasma glucose | 137 | MD -1.24 (-1.38 to -1.10) | -2.81 to 0.33 | 97.7 |
| 2-h postprandial glucose | 105 | MD -1.79 (-2.02 to -1.56) | -4.11 to 0.53 | 97.6 |
| Total cholesterol | 28 | MD -0.66 (-0.86 to -0.45) | -1.74 to 0.43 | 96.7 |
| Triglycerides | 29 | MD -0.44 (-0.65 to -0.23) | -1.55 to 0.67 | 99.3 |
| LDL cholesterol | 35 | MD -0.64 (-0.89 to -0.40) | -2.05 to 0.76 | 99.5 |
| HDL cholesterol | 27 | MD 0.13 (0.01 to 0.25) | -0.46 to 0.71 | 98.9 |
| **Other outcomes not formally graded** |  |  |  |  |
| MMAS-8 score | 28 | MD 1.21 (0.92 to 1.50) | -0.30 to 2.73 | 94.7 |
| Patient satisfaction | 12 | OR 5.39 (2.91 to 9.99) | 0.84 to 34.51 | 76.5 |
| **Exploratory investigator-defined outcomes** |  |  |  |  |
| Investigator-defined glycaemic effective rate | 31 | OR 4.54 (3.43 to 6.00) | 1.18 to 17.43 | 70.6 |
| Investigator-defined blood-pressure effective rate | 56 | OR 3.46 (2.89 to 4.14) | 1.25 to 9.57 | 64.9 |
| Investigator-defined overall effective rate | 9 | OR 2.63 (1.83 to 3.77) | Not estimable | 0.0 |

RR is the primary scale for headline binary outcomes. For outcomes with extreme heterogeneity, pooled estimates are descriptive averages only. Prediction intervals were not reported when fewer than ten estimates produced $\tau^2$ equal to zero. Investigator-defined effective rates were exploratory.

**Table 3.** Core-outcome robustness analyses.

| Outcome | Analysis | k | Effect (95% CI) | 95% prediction interval | Interpretation |
| --- | --- | --- | --- | --- | --- |
| Reported adherence | RR, all eligible trials (primary) | 163 | RR 1.37 (1.32 to 1.43) | 0.98 to 1.93 | Prediction interval crosses 1 |
| Reported adherence | RR, low-risk trials only | 28 | RR 1.34 (1.22 to 1.47) | 0.96 to 1.87 | Favourable estimate; interaction P=0.645 |
| Reported adherence | OR, all eligible trials (sensitivity) | 163 | OR 4.78 (4.33 to 5.28) | 2.13 to 10.71 | Prediction interval excludes 1 |
| Reported adherence | RD, all eligible trials (sensitivity) | 163 | RD 0.24 (0.22 to 0.25) | 0.03 to 0.44 | Effect-scale sensitivity |
| Reported adverse drug reactions | RR, all eligible trials (primary) | 94 | RR 0.41 (0.37 to 0.45) | 0.22 to 0.74 | Prediction interval excludes 1 |
| Reported adverse drug reactions | RR, low-risk trials only | 20 | RR 0.38 (0.32 to 0.46) | 0.21 to 0.70 | Favourable estimate; interaction P=0.444 |
| Reported adverse drug reactions | OR, all eligible trials (sensitivity) | 94 | OR 0.29 (0.26 to 0.33) | 0.14 to 0.62 | Prediction interval excludes 1 |
| Unplanned readmission | RR, all eligible trials (primary) | 22 | RR 0.52 (0.46 to 0.60) | 0.42 to 0.66 | Prediction interval excludes 1 |
| Unplanned readmission | RR, low-risk trials only | 4 | RR 0.56 (0.30 to 1.05) | 0.30 to 1.05 | Four results; imprecise; interaction P=0.652 |
| Unplanned readmission | OR, all eligible trials (sensitivity) | 22 | OR 0.36 (0.29 to 0.45) | 0.23 to 0.57 | Prediction interval excludes 1 |

Of the 22 readmission trials, 15 involved heart failure, myocardial infarction, or coronary disease and contributed 72.6% of the weight. Their pooled RR was 0.57 (95% CI 0.51-0.64). Diabetes or hypertension trials were imprecise and heterogeneous, limiting direct application beyond high-risk cardiovascular transitions (Figure 2; Supplementary Table S11E).

Readmission estimates were RR 0.43 (95% CI 0.16-1.13) at 3 months or less, 0.48 (0.38-0.59) at more than 3 to 6 months, and 0.58 (0.48-0.70) beyond 6 months. The observed relative association attenuated with longer reported follow-up (ratio of RRs 1.11 per additional 6 months, 95% CI 1.01-1.22; P=0.033), supporting the use of fixed 30-day and 90-day endpoints in future evaluations. Fixed windows are proposed for comparability rather than because the short-term estimate was larger, since the 3-month stratum rested on four trials with a wide confidence interval (Supplementary Tables S11A-S11B; Supplementary Figure S3).

A ROB-ME-informed assessment found some concerns because 244 of the 266 trials in the complete evidence map did not contribute a readmission result and protocols were generally unavailable. Non-contribution was counted against the full evidence map, and whether readmission had been measured was generally unknown; available information gave no direct indication that outcome contribution depended on result direction or statistical significance (Supplementary Tables S11C-S11D).

Odds-ratio and zero-event sensitivity analyses did not reverse the direction of the three core results. Outcome-specific low-risk estimates remained favourable for reported adherence (28 results; RR 1.34, 95% CI 1.22-1.47) and reported adverse drug reactions (20 results; RR 0.38, 95% CI 0.32-0.46). For readmission, the four-result low-risk point estimate remained below 1 but was imprecise (RR 0.56, 95% CI 0.30-1.05). Reported adherence remained scale-sensitive: RR 1.37, OR 4.78, and risk difference 0.24, which are not interchangeable (Table 3; Supplementary Tables S5, S6A-S6B, S13A; Supplementary Figure S1).

Trustworthiness screening raised no serious integrity signal; unresolved ethics, registration, or report-concordance information produced some-concern classifications in 211 of 238 trials (Supplementary Tables S15A-S15C).

As an additional scope-related robustness check, the sampled non-core validation included 68 contributing reports and 15,160 participants. Effect directions were preserved, but magnitudes differed without a consistent pattern across outcomes; no core-versus-non-core interaction remained significant after multiplicity adjustment, although readmission validation was based on only five trials and had a wide prediction interval. Full effect estimates, interaction tests, and leave-one-out analyses are reported in Supplementary Tables S18-S20.

At the median control risks, pharmacist involvement corresponded to 255 additional patients meeting the reported adherence definition per 1,000, 139 fewer reported adverse drug reactions, and 147 fewer unplanned readmissions. Follow-up was mixed, so these scenarios are not comparable across outcomes (Supplementary Table S7).

Among the secondary outcomes, mortality was lower on average (RR 0.53, 95% CI 0.39-0.72) from nine estimates and 72 deaths, with low certainty. MMAS-8 improved by 1.21 points (95% CI 0.92-1.50) and satisfaction favoured pharmacist involvement, both with high heterogeneity and prediction intervals crossing the null (Supplementary Figure S14).

Pooled averages for disease-control outcomes favoured pharmacist involvement, including systolic blood pressure (MD −8.83, 95% CI −10.08 to −7.58) and glycated haemoglobin (MD −1.09, 95% CI −1.24 to −0.93), but these are descriptive only: I² was 96.7% to 99.5% and every biomarker prediction interval crossed zero. Removing influential estimates left heterogeneity above 95%. Investigator-defined effective rates produced favourable pooled ORs, but varying thresholds limited interpretation and they were retained as exploratory (Figure 4; Table 2; Supplementary Tables S12A-S12B; Supplementary Figures S4, S16-S22).

**Figure 4.**
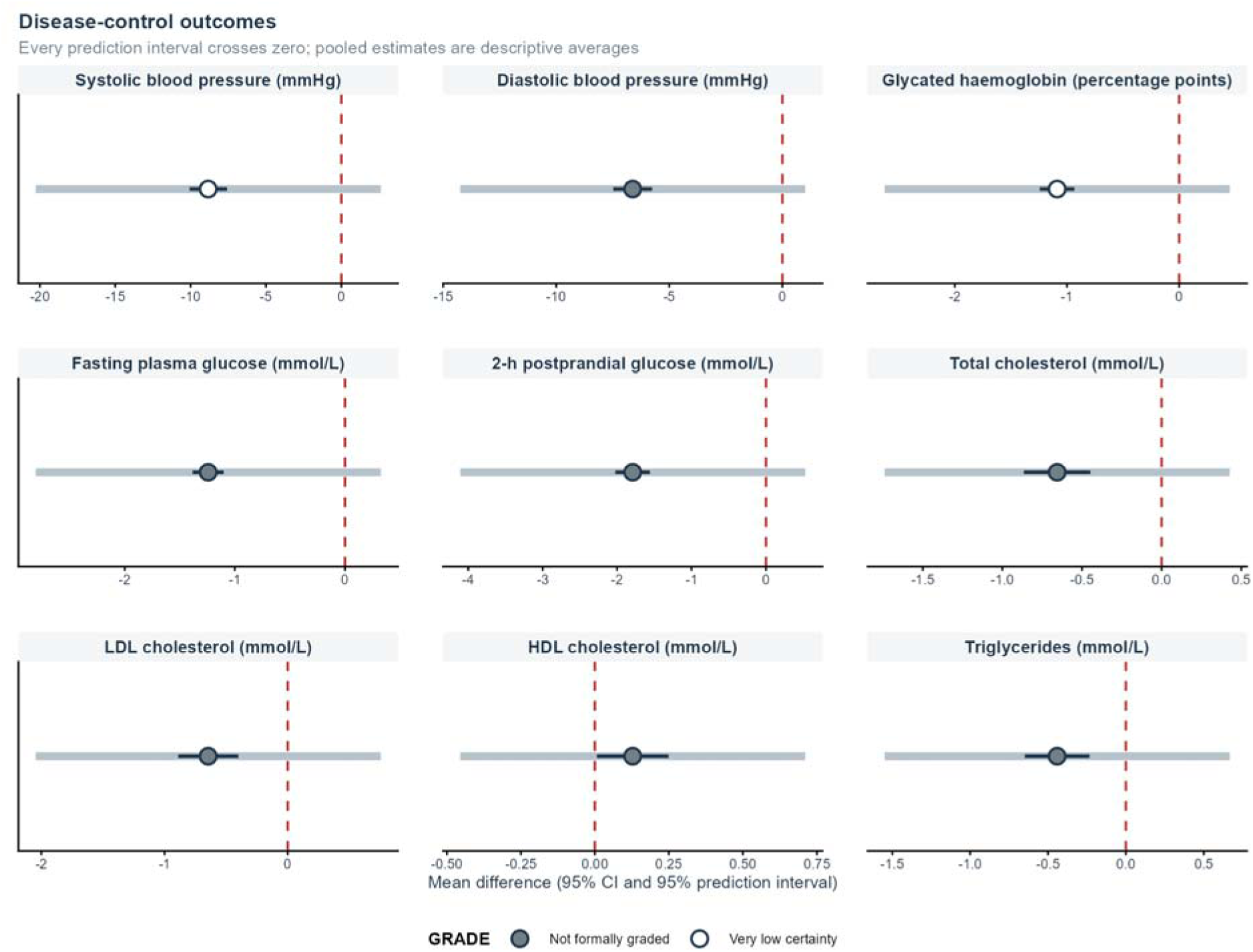
Disease-control outcomes. Dark bars show 95% confidence intervals and pale bars show 95% prediction intervals. Open symbols denote very-low-certainty outcomes and filled symbols outcomes not formally graded. Every prediction interval crosses zero, so pooled estimates are descriptive averages rather than expected effects for an individual setting.

Length of stay was 1.77 days shorter on average (95% CI −2.96 to −0.58), but I² was 92.6% and the prediction interval was −5.71 to 2.17 days. The evidence was rated very low certainty (Supplementary Figure S15).

With healthcare CPI, hospitalisation expenditure was lower by CNY 1258 on average (95% CI −2011 to - 504) and medication expenditure by CNY 1519 (95% CI −2699 to −339). The prediction intervals were - 3293 to 778 and −5325 to 2288, respectively. Overall-CPI sensitivity estimates were CNY −1232 and CNY - 1476. These are selected, unpaired expenditure components. They exclude service-delivery costs and are not estimates of net savings (Figure 5; Table 2; Supplementary Figures S23-S24).

**Figure 5.**
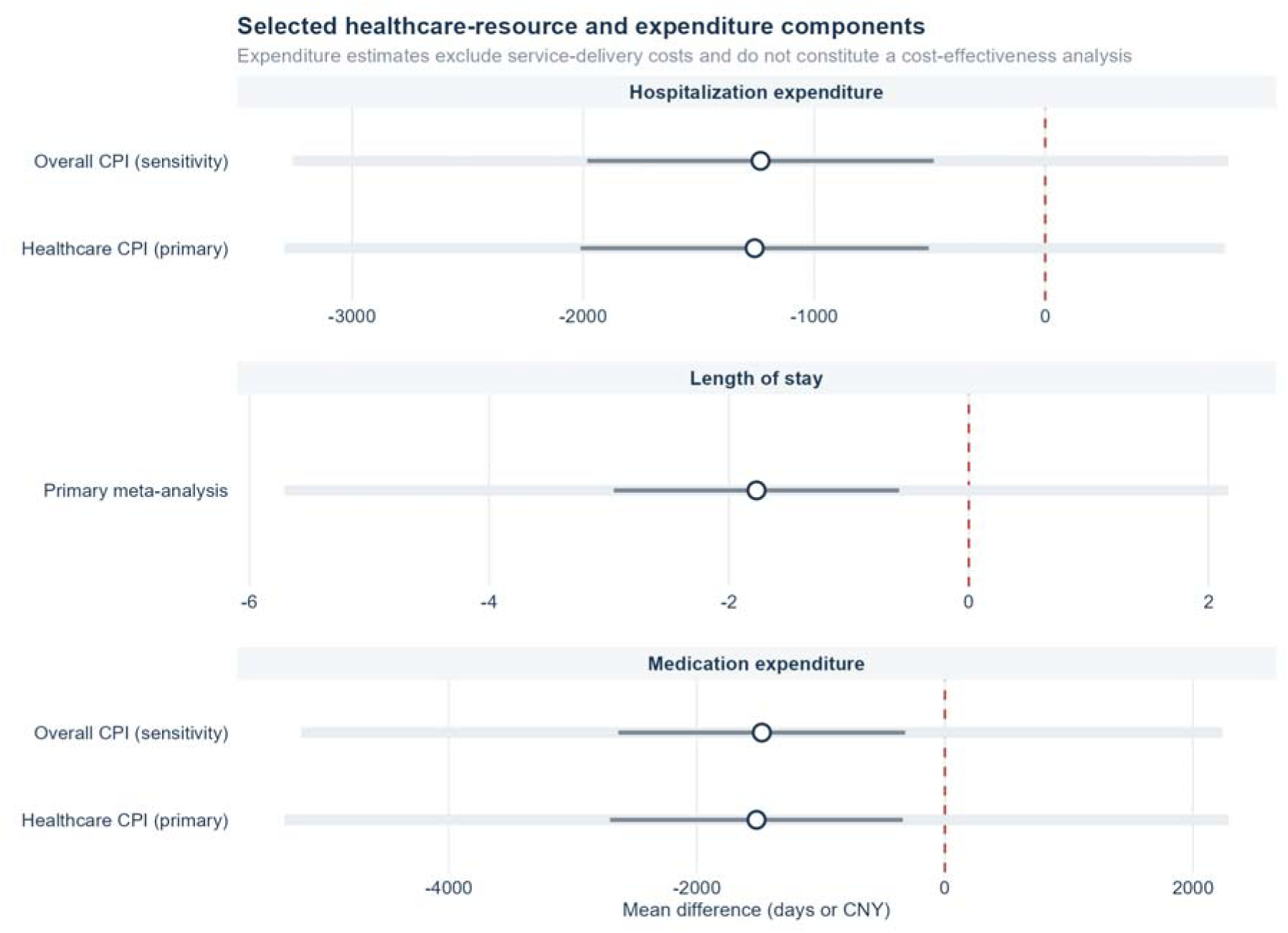
Selected healthcare-resource and expenditure components. Healthcare CPI is the primary price adjustment and overall CPI a sensitivity analysis. The expenditure estimates omit service-delivery costs and do not constitute a cost-effectiveness analysis.

No moderator association survived false-discovery-rate correction except disease category for two investigator-defined outcomes, and no study-level variable explained the extreme biomarker heterogeneity. The prespecified continuous readmission analysis indicated attenuation with longer follow-up (P=0.033) (Supplementary Tables S2A, S2B, and S11A-S11B; Supplementary Figure S2).

Intervention descriptions were incomplete: education was explicitly reported in 219 of 266 trials, adherence support in 136, and post-discharge follow-up in 82. No reporting-status association survived false-discovery-rate correction (Supplementary Tables S1 and S9).

Primary tests suggested small-study effects for glycated haemoglobin (Egger P<0.001), systolic blood pressure (Egger P<0.001), length of stay (Egger P=0.011), and satisfaction (Peters P=0.025) (Supplementary Table S21B; Supplementary Figures S25-S27). For reported adherence, reported adverse drug reactions, and readmission, the primary Peters tests were not significant, whereas supplementary Egger tests were significant. These method-dependent findings were treated as signals of small-study effects rather than proof of publication bias. Across the 19 non-cost outcomes examined, alternative variance estimators, leave-one-out analyses, and exclusion of trials classified as high risk in the descriptive evidence map did not reverse the direction of the pooled estimate (Supplementary Table S21A).

All nine graded outcomes were low or very low certainty (Figure 6; Table 4; Supplementary Table S3). The certainty ratings reflect study limitations, measurement indirectness, imprecision, and reporting concerns. For reported adherence, the broader measurement audit provided contextual evidence of extensive reliance on named self-report and investigator-developed measures, but it was not treated as the measurement distribution of the 163 binary estimates. Readmission was downgraded because most trials and weight came from high-risk cardiovascular populations. Excluding high-risk trials did not reverse effect directions.

**Figure 6.**
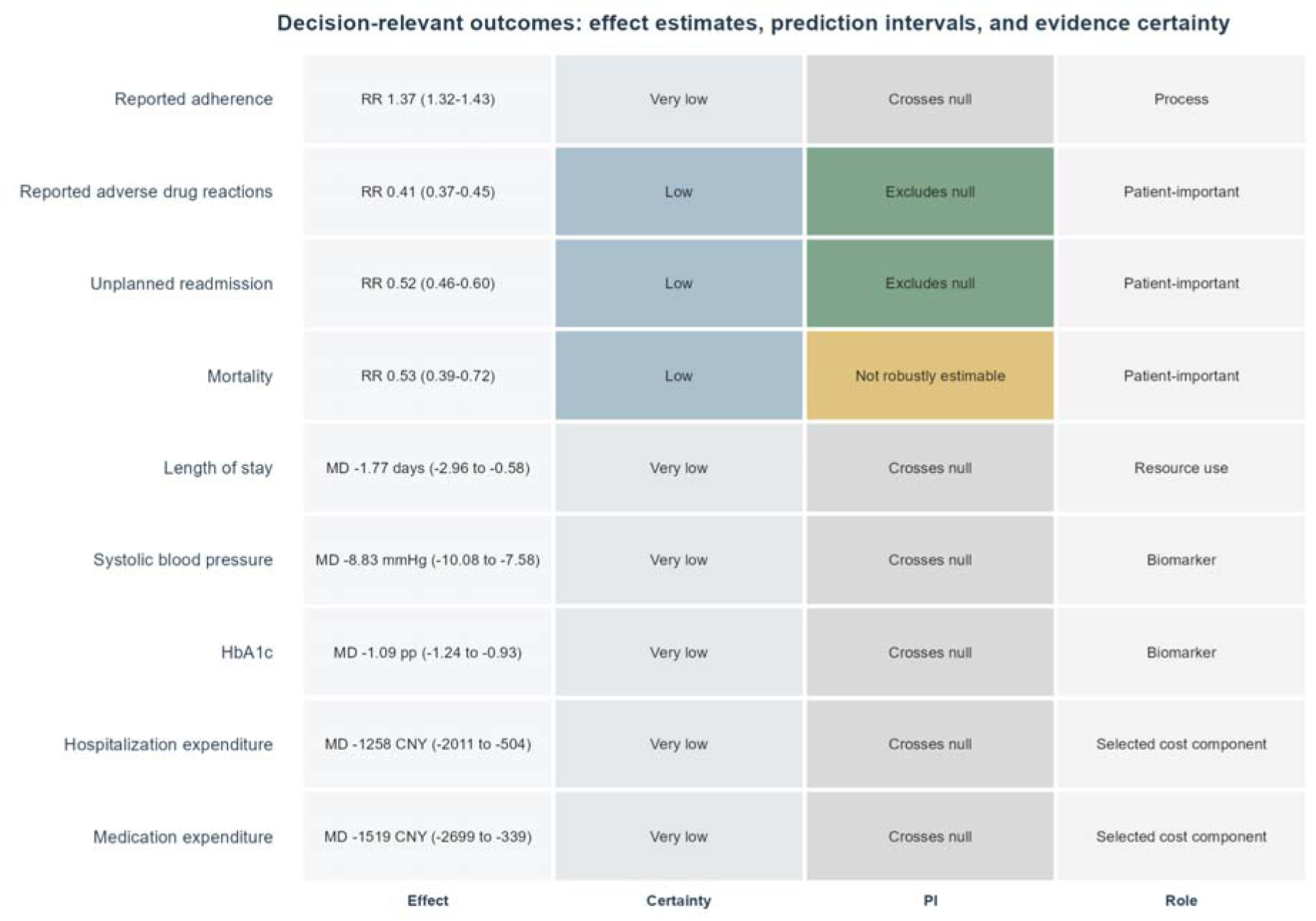
Decision-relevant outcomes: effect estimates, prediction intervals, and evidence certainty. The display integrates the pooled estimate, Grading of Recommendations Assessment, Development and Evaluation (GRADE) certainty, prediction-interval status, and outcome role. Prediction intervals describe expected effects in future comparable studies and do not establish transportability.

**Table 4.** Concise GRADE summary.

| Outcome | Certainty | Main reasons |
| --- | --- | --- |
| Reported adherence | Very low | Risk-of-bias concerns; substantial inconsistency; measurement indirectness. |
| Reported adverse drug reactions | Low | Risk-of-bias concerns; definitions and ascertainment varied. |
| Unplanned readmission | Low | Risk-of-bias concerns; evidence concentrated in high-risk cardiovascular populations. |
| Mortality | Low | Risk-of-bias concerns; nine trials and 72 deaths produced serious imprecision. |
| Length of stay | Very low | Risk-of-bias concerns; very serious inconsistency; suspected small-study effects. |
| Systolic blood pressure | Very low | Risk-of-bias concerns; very serious inconsistency; suspected small-study effects. |
| Glycated haemoglobin | Very low | Risk-of-bias concerns; very serious inconsistency; suspected small-study effects. |
| Hospitalisation expenditure | Very low | Risk-of-bias concerns; inconsistency and indirectness; delivery costs absent. |
| Medication expenditure | Very low | Risk-of-bias concerns; very serious inconsistency and indirectness; delivery costs absent. |

The risk-of-bias downgrade reflects widespread methodological limitations across the evidence base and was retained even when exclusion of high-risk trials did not materially change the full-data estimates. Adherence was downgraded for risk of bias, inconsistency, and indirectness; the broader measurement audit contextualised the indirectness judgement but was not treated as the measurement-method distribution of the 163 pooled binary estimates. Reported adverse drug reactions were downgraded for indirectness because ascertainment varied. Discordant Peters and supplementary Egger tests were recorded without an additional publication-bias downgrade.
Detailed GRADE domain judgements are provided in Supplementary Table S3.

## Discussion

Across 266 Chinese randomised trials, unplanned readmission provided the strongest basis for prospective evaluation. It was internally consistent, with I² of 10.0% and a prediction interval excluding the null, and the endpoint is objectively ascertainable. It applies where the trials sit: 15 of the 22 trials and 72.6% of the weight came from heart failure, myocardial infarction, or coronary disease, and the four low-risk readmission results were too few to confirm the association independently. Reported adverse drug reactions showed the most consistent effect across effect scales, the low-risk subset, and the sampled non-core evidence, although ascertainment was unblinded and definitions varied. Reported adherence showed the largest absolute gain, although most trials relied on self-report or investigator-defined measures and certainty was very low. The evidence therefore narrowed at each step from statistical association to credibility, applicability, and decision usefulness.

The readmission evidence was most directly applicable to high-risk cardiovascular transitions. Two further features narrow it: the diabetes or hypertension estimate was imprecise, and the observed relative association attenuated with longer reported follow-up. Together these support further evaluation in high-risk cardiovascular transitions rather than a uniform claim across all cardiometabolic conditions.

The observed readmission association was substantially larger than the pooled readmission risk ratios reported by international syntheses of medication review and transitional care, which have generally been closer to 0.9.^4,5,28^ Our own earlier review of medication therapy management services, searched to April 2022 in the international literature, reported a readmission odds ratio of 0.78, a materially smaller association than the one observed here on either the odds or the risk scale.^14^ None of its 81 included trials overlaps with the present review, and only six were conducted in China, two of them in cardiovascular disease, so the two estimates rest on separate bodies of evidence. Broader reviews of pharmacist-led chronic disease management and medication therapy management report effects in the same modest range across the outcomes they assess.^3,6,7^ Differences in usual care, baseline risk, disease mix, follow-up, endpoint definitions, and trial methods may contribute to this contrast. Residual bias may also contribute, given widespread RoB 2 concerns and limited blinding of outcome ascertainment. In addition, INSPECT-SR left governance or registration information unresolved for many trials, limiting confidence in the completeness of trial documentation without itself establishing bias. Together with the concentration of evidence in heart failure and post-infarction populations, these differences caution against directly transferring the Chinese pooled estimate to other health systems.

Adherence needs careful interpretation. The pooled RR describes a relative increase in sufficiently comparable reported binary thresholds. The separate 228-trial audit characterised the broader range of adherence measures used across the included trials, including bespoke continuous scales that could not be combined; only nine trials in that broader audit used objective measures. The audit therefore provides context about measurement quality without defining the exact measurement distribution of the 163 pooled estimates.^17^ Disease-control estimates answer a different question. Their pooled means describe the centre of a highly dispersed distribution rather than a site-level expectation: an average systolic reduction near 9 mm Hg coexisted with a prediction interval that included no benefit, and heterogeneity remained above 95% after removing influential estimates. The dispersion more plausibly reflects differences the publications did not measure consistently.

For clinicians, the evidence is most relevant where pharmacist involvement aims to improve medication use, detect and prevent reported adverse drug reactions, or support patients after discharge. These outcomes were generally evaluated within multicomponent programmes that combined review, education, monitoring, prescriber communication, and follow-up; evaluations of transitions-of-care programmes show that outcomes depend on the service bundle, target population, timing, and local workflow.^29,30^ Because a publication that omits an activity does not establish its absence, comparisons between explicit reporting and non-reporting reflect reporting completeness as much as intervention content.^18,31^ Reported adverse drug reactions are measurable indicators of medication-related harm.^32^ Unplanned readmission is likewise a measurable patient-important outcome relevant to care transitions.^5,28–30^ Priority setting also depends on disease burden, feasibility, equity, and opportunity cost.^33^ Because the readmission evidence is concentrated in heart failure and coronary transitions, these populations may be particularly suitable for prospective evaluation.

Transportability is likely to depend on baseline risk and on how the service is delivered locally, including the pharmacist’s clinical authority and intensity of contact, but the publications did not report these consistently enough to test, and the study-level moderators that could be tested explained little. Most trials were conducted in secondary or tertiary hospitals (224 of 266), with relatively little evidence from the primary care settings into which chronic-disease policy is expanding. National standards can define core functions and outcome definitions, while delivery models require local calibration. Future effectiveness trials should therefore collect the implementation and cost data required for payment decisions.^2^ A practical pilot in heart failure or recent myocardial infarction could evaluate a package combining discharge reconciliation, pharmacist review, education, documented handover, and follow-up at 30 and 90 days. Adherence could be measured using dispensing records,^17,34^ patient-important endpoints should include 30-day and 90-day readmission, and delivery inputs and costs should be collected prospectively.

These estimates capture selected hospital and medication expenditure components. A CHEERS-informed audit of the 13 contributing trials found none reporting service-delivery inputs, none reporting a complete incremental cost, and none reporting a health utility; no trial stated an analytic perspective or a source price year (Supplementary Table S17A).^26^ CPI standardisation aligns price years and leaves local prices, benefit packages, accounting definitions, and perspective untouched.^25^ This missing cost structure identifies the main evidence gap for payment design. It is also a policy problem. More than 260 eligible trials were identified in core-indexed journals alone, yet few reported pharmacist time, programme inputs, or incremental costs. Payment reform therefore needs implementation data alongside efficacy estimates.^35^ National DRG/DIP policies define prospective case-based payment, under which an avoided readmission reduces payer expenditure while the hospital delivering the preventive service bears its cost.^36,37^ Supplementary Table S4 sets out a local break-even scenario as a methodological illustration of the service-cost information a payment evaluation would require, and its values are not offered as a tariff or a pricing recommendation.

The review searched six databases and applied the prespecified sensitivity, certainty, and trustworthiness assessments described in the Methods. Together these allowed statistical consistency, credibility, and applicability to be considered separately. Indexing, RoB 2, and INSPECT-SR address different questions, and none changed the principal findings. INSPECT-SR raised no serious integrity signal, and the sampled non-core validation agreed on direction but not magnitude, with no core-versus-non-core interaction surviving multiplicity adjustment. Its stratified design limits inference about the magnitude of publication-channel selection.^38,39^

The limitations span five domains. First, restricting Chinese-language reports by publication venue may have introduced publication-channel-related selection; the sampled non-core analysis supported directional robustness but was neither exhaustive nor population-representative. Second, most trials had methodological and reporting limitations, particularly reliance on self-reported adherence. Third, readmission evidence was concentrated in high-risk cardiovascular transitions, while biomarker effects and follow-up were heterogeneous. Fourth, intervention delivery, pharmacist inputs, and service costs were incompletely reported, and quality of life, although registered, could not be synthesised because the contributing reports used too many different instruments to pool. Fifth, small-study effects were suspected for glycated haemoglobin, systolic blood pressure, and length of stay, and supplementary Egger tests were significant for all three principal outcomes although the prespecified Peters tests were not; if driven by selective publication, these could exaggerate pooled associations. Trials were also clustered within providers, with 266 trials from about 235 institutions and up to five from one centre, so some may share teams and populations.

Among the outcomes assessed, unplanned readmission after admission with heart failure or coronary disease is where this evidence base comes closest to supporting action: the estimate was comparatively consistent and the endpoint is auditable, although it applies to the populations these trials enrolled and remained low certainty. The reduction in reported adverse drug reactions was the most reproducible principal result, although definitions and ascertainment varied. Reported adherence showed the largest apparent gain; the broader measurement audit found only nine objective assessments among 228 adherence-reporting trials. Biomarker estimates were descriptive pooled averages with extreme heterogeneity, and service-delivery and incremental-cost data were insufficient for economic evaluation. The constraint on using this evidence is measurement and not the number of trials: future studies should use fixed 30-day and 90-day clinical endpoints, dispensing-based or other objective adherence measures, rigorous outcome ascertainment, and prospective collection of service inputs and costs.

### Preprint

An earlier version of this work was posted as a preprint on medRxiv on July 25, 2024 (doi:10.1101/2024.07.25.24310734). The present manuscript represents a substantially updated evidence base and methodologically restructured analysis.

## Contributors

Zhijie Deng: Conceptualization, Methodology, Investigation, Data curation, Formal analysis, Validation, Visualization, Project administration, Writing - original draft, Writing - review & editing. Fanglu Chen: Methodology, Software, Formal analysis, Validation, Visualization, Writing - review & editing. Lin Gui: Investigation, Data curation, Validation, Writing - review & editing. Yangjin Huang: Conceptualization, Methodology, Writing - review & editing. Jiahui Chen: Investigation, Data curation, Writing - review & editing. Yansheng Liu: Investigation, Data curation, Writing - review & editing. Yueqiu Gong: Investigation, Data curation, Validation, Writing - review & editing. Lu Zeng: Investigation, Data curation, Writing - review & editing. Anhua Wei: Conceptualization, Supervision, Project administration, Funding acquisition, Writing - review & editing. All authors had full access to all the data in the study and had final responsibility for the decision to submit for publication. Anhua Wei and Fanglu Chen directly accessed and verified the underlying data reported in the manuscript.

## Ethics

This study analysed aggregate results reported in published trial reports and did not involve access to individual participant data, so ethics committee approval and informed consent were not required. Each included trial reported its own ethical approval status, and this information was recorded during trustworthiness screening.

## Data sharing

The data underlying this Article, comprising the data dictionary, the study-level data as extracted from the published trial reports, the cleaned analysis dataset, and the analysis code, are deposited in the Science Data Bank repository at https://doi.org/10.57760/sciencedb.46014. No individual participant data were obtained; all inputs were aggregate results reported in the source publications. The registered protocol is publicly available in PROSPERO (CRD42023414180). Data are available from the date of publication, with no planned end date. Access is restricted: requests should be submitted through the repository and will be granted after approval by the corresponding author, who will review the proposed use before access is provided.

## Data Availability

All data produced in the present study are available upon reasonable request to the authors

https://doi.org/10.57760/sciencedb.46014

## Acknowledgments

We thank Shunshun Peng and Yufeng Ding for their advice on the direction of this work and on the methods used for data extraction.

## Declaration of generative AI and AI-assisted technologies in the writing process

During the preparation of this work the authors used Claude Opus 5 (Anthropic), accessed through the Claude Code desktop application, to edit and restructure manuscript text, to check the internal consistency of reported numbers and cross-references, and to format the reference list, and OpenAI Codex, a GPT-5-based model, to optimise the figure-plotting code. After using these tools, the authors reviewed and edited the content as needed and take full responsibility for the content of the publication. Generative AI was not used to design the review, run the searches, select studies, extract data, or produce or interpret the statistical results; every figure was generated from the locked analytical dataset, and all statistical results, citations, and interpretations were checked against that dataset and the cited sources.

