## Supplementary materials for "Pharmacist-involved services for cardiometabolic conditions in China: a systematic review and meta-analysis"

This file contains supplementary methods, tables, figures, study characteristics, intervention coding, risk-of-bias judgements, detailed Grading of Recommendations Assessment, Development and Evaluation (GRADE) assessments, readmission follow-up and missing-evidence analyses, continuous-outcome influence diagnostics, adherence effect-scale analyses, model and small-study-effect sensitivity analyses, post hoc non-core-journal directional validation, search strategies, and the Preferred Reporting Items for Systematic Reviews and Meta-Analyses (PRISMA) checklist.

Chinese-language study-characteristic fields have been translated into English for presentation. Appendix 1 reproduces the database-specific strategies used in the final search through July 20, 2026. Chinese-language query strings are shown as executed, and no study-design or other database filter was appended.

### Supplementary Methods

Adherence measurement was audited across all 266 included trials. Of these, 228 reported some adherence information. The audit classified measurement as named self-report, investigator-developed, dispensing-based objective, physiological-marker inference, or not reported and was intentionally broader than the quantitative syntheses. Sufficiently comparable binary adherence rates and 8-item Morisky Medication Adherence Scale (MMAS-8) scores were pooled separately; bespoke continuous scores with incompatible ranges, definitions, or directions were retained in the evidence map but not quantitatively combined. Two reviewers independently mapped reporting and design features across all 266 trials using the five domains of the Cochrane risk-of-bias tool for randomised trials, version 2 (RoB 2). This descriptive evidence map was not substituted for outcome-specific RoB 2 in inferential sensitivity analyses. For the three core-outcome low-risk sensitivity analyses, outcome-specific RoB 2 judgements were completed for 52 results (28 for adherence, 20 for reported adverse drug reactions, and four for readmission), all low risk across the five domains and overall. The INSPECT-SR trustworthiness tool used all 21 checks in the official four-domain structure for the 238 trials contributing the three core outcomes only; it was not reapplied to the 68 non-core-journal validation trials. Unavailable registration, ethics, or concordance information was retained as unresolved and led to some concerns rather than no concerns.

#### Software environment

Analyses were conducted in R version 4.5.2. The archived analytical environment recorded metafor 4.8-0, meta 8.2-1, and clubSandwich 0.7.0 as the principal meta-analytic packages. Packages that materially supported data preparation and figure generation are listed below. The figure-plotting code was optimised with the assistance of OpenAI Codex, a GPT-5-based model; all figures were generated from the locked analytical dataset and were checked against the reported estimates by the authors.

| **Package** | **Version** | **Primary role** |
| --- | --- | --- |
| metafor | 4.8-0 | Random-effects meta-analysis and effect-size calculation |
| meta | 8.2-1 | Meta-analysis and sensitivity procedures |
| clubSandwich | 0.7.0 | CR2 robust variance estimation |
| ggplot2 | 4.0.2 | Figure generation |

Supplementary Table S1 | Intervention-activity reporting

| **Activity** | **Explicitly reported** | **Not reported** | **Missing coding** | **Reported (%)** |
| --- | --- | --- | --- | --- |
| Medication reconciliation | 18 | 241 | 7 | 6.9% |
| Medication or treatment review | 70 | 189 | 7 | 27.0% |
| Patient education | 219 | 40 | 7 | 84.6% |
| Adherence support | 136 | 123 | 7 | 52.5% |
| Adverse-event monitoring | 54 | 205 | 7 | 20.8% |
| Prescribing input or authority | 46 | 213 | 7 | 17.8% |
| Post-discharge follow-up | 82 | 177 | 7 | 31.7% |
| Multidisciplinary collaboration | 90 | 169 | 7 | 34.7% |

No individual reporting-status association survived false-discovery-rate (FDR) correction. Full model coefficients are available in the accompanying source-data file. Not reported is an unknown publication state.

Supplementary Table S2A | Summary of moderator analyses. Moderators were tested within families with Benjamini-Hochberg control of the false discovery rate; all are exploratory.

| **Moderator** | **Coverage** | **Adjusted result** | **Interpretation** | **Analysis status** |
| --- | --- | --- | --- | --- |
| Disease category | 21 outcomes | Blood-pressure effective rate q=0.007; diastolic blood pressure q=0.002 | Exploratory; sparse categories | Exploratory |
| Service label | 18 estimable outcomes | No false-discovery-rate-adjusted association | Labels do not define content | Exploratory |
| Mean age | Study-level means | No false-discovery-rate-adjusted association | Ecological analysis | Exploratory |
| Publication year | Eligible outcomes | No q<0.05; 2-h glucose q=0.053 | No evidence of a stable temporal trend | Exploratory |
| Follow-up duration | 682 of 1,107 estimates reported | No false-discovery-rate-adjusted association | 425 estimates missing duration | Exploratory |
| Mainland region | 1,083 of 1,107 estimates classifiable | No false-discovery-rate-adjusted association | Study-level geography | Exploratory |
| Activity reporting status | Eight activities | No individual association survived false-discovery-rate correction | Reporting, not component efficacy | Exploratory |
| Number of reported activities | Complete-case analyses | 2-h glucose q=0.007; HbA1c q=0.046 | Reporting completeness signal | Exploratory |
| Readmission follow-up | 21 of 22 estimates | Per 6 months ratio of RRs 1.11 (1.01 to 1.22), P=0.033; window omnibus P=0.287 | Association attenuated with longer reported follow-up | Prespecified continuous moderator; exploratory windows |

Supplementary Table S2B | Follow-up and regional coverage

| **Item** | **Coverage** | **Result** |
| --- | --- | --- |
| Follow-up reported | 682/1107 | No moderator association survived FDR correction |
| Follow-up missing | 425/1107 | Missing values remained missing |
| Mainland region classifiable | 1083/1107 | No regional association survived FDR correction |
| Hong Kong, Taiwan, or unclassifiable | 24/1107 | Retained in main meta-analysis |

Supplementary Table S3 | Detailed GRADE domains

| **Outcome** | **Risk of bias** | **Inconsistency** | **Indirectness** | **Imprecision** | **Publication bias** | **Upgrade** | **Certainty** |
| --- | --- | --- | --- | --- | --- | --- | --- |
| Medication adherence rate | Serious | Serious | Serious indirectness: a broader evidence-map audit found extensive reliance on named self-report and investigator-developed measures (219 of 228 adherence-reporting trials; nine objective measures). The audit was not limited to the 163 binary estimates and was used as contextual evidence rather than as their contributor-specific measurement distribution. | Not serious | Inconsistent test results |  | Very low |
| Reported adverse drug reactions | Serious | Not serious | Serious | Not serious | Inconsistent test results |  | Low |
| Unplanned readmission | Serious | Not serious | Serious: 15 of 22 trials and 72.6% of weight were heart failure, myocardial infarction, or coronary disease | Not serious | Some concerns; no additional downgrade |  | Low |
| Mortality | Serious | Not serious | Not serious | Serious | Not assessed |  | Low |
| Length of stay | Serious | Very serious | Not serious | Not serious | Suspected |  | Very low |
| Systolic blood pressure | Serious | Very serious | Not serious | Not serious | Suspected |  | Very low |
| Glycated haemoglobin | Serious | Very serious | Not serious | Not serious | Suspected |  | Very low |
| Hospitalisation expenditure | Serious | Serious | Serious | Not serious | Not assessed |  | Very low |
| Medication expenditure | Serious | Very serious | Serious | Not serious | Undetected |  | Very low |

Supplementary Methods | From the payer perspective, the local break-even scenario multiplied the risk-ratio (RR)-based readmission absolute-risk reduction at the median control risk (14.7 percentage points) by published 2025 Changzhi heart-failure diagnosis-related-group (DRG) payments (main-text references 35 and 36). The interval scenario used the 95% confidence limits of the pooled RR. No 30-day or 90-day budget impact was estimated because the meta-analysis lacked fixed-time readmission effects and a defensible observed service-package cost.

Supplementary Table S4 | Local DRG break-even scenario

| **DRG** | **Hospital level** | **Illustrative payment (CNY)** | **Payer break-even service cost per patient, point (95% CI scenario), CNY** | **Gross avoided payer payment per 1,000, point (95% CI scenario), CNY million** |
| --- | --- | --- | --- | --- |
| FR21 | Municipal tertiary | 8,884 | 1,302 (1,092-1,486) | 1.30 (1.09-1.49) |
| FR23 | Municipal tertiary | 7,690 | 1,127 (945-1,286) | 1.13 (0.94-1.29) |
| FR25 | Municipal tertiary | 6,755 | 990 (830-1,130) | 0.99 (0.83-1.13) |
| FR21 | Municipal secondary | 7,045 | 1,033 (866-1,178) | 1.03 (0.87-1.18) |
| FR23 | Municipal secondary | 6,098 | 894 (749-1,020) | 0.89 (0.75-1.02) |
| FR25 | Municipal secondary | 5,356 | 785 (658-896) | 0.79 (0.66-0.90) |
| FR21 | County secondary | 6,693 | 981 (822-1,120) | 0.98 (0.82-1.12) |
| FR23 | County secondary | 5,794 | 849 (712-969) | 0.85 (0.71-0.97) |
| FR25 | County secondary | 5,089 | 746 (625-851) | 0.75 (0.63-0.85) |
| FR21 | Primary | 5,228 | 766 (642-875) | 0.77 (0.64-0.87) |
| FR23 | Primary | 4,526 | 663 (556-757) | 0.66 (0.56-0.76) |
| FR25 | Primary | 3,975 | 583 (488-665) | 0.58 (0.49-0.66) |

The scenario is a methodological illustration, not a national tariff, budget-impact analysis, or economic evaluation. It assumes that an avoided readmission saves the payer the full DRG payment. Hospitals may face the opposite financial incentive because they bear the service cost while losing admission revenue. Heart-failure DRGs were chosen because heart failure, myocardial infarction, and coronary disease supplied 15 of the 22 readmission trials and 72.6% of the random-effects weight (Supplementary Table S11E); the scenario therefore matches the population that dominates the readmission evidence but not the wider cardiometabolic mix of the review. Comparison across hospital levels should use the same DRG group: for FR21 the payer threshold falls from Chinese yuan (CNY) 1,302 at municipal tertiary level to CNY 766 at primary level, a ratio of about 1.7. The four-result outcome-specific low-risk readmission estimate crossed the null, so a positive threshold is not assured. The table gives a break-even threshold, not a judgement of feasibility: no observed pharmacist-service cost anchor was available, and future pilots should measure pharmacist time by grade and fully loaded wage before the threshold can be compared with an actual service cost.

Supplementary Table S5 | Zero-event sensitivity analyses

| **Outcome** | **Analysis** | **k** | **N_Intervention** | **N_Control** | **Effect** | **CI_Lower** | **CI_Upper** | **PI_Lower** | **PI_Upper** | **I2** | **Tau2** |
| --- | --- | --- | --- | --- | --- | --- | --- | --- | --- | --- | --- |
| ADR | Inverse variance, 0.5 only for zero cells | 94 | 6376 | 6277 | 0.405 | 0.367 | 0.448 | 0.222 | 0.739 | 38.333 | 0.089 |
| ADR | Inverse variance, zero-event trials excluded | 91 | 6265 | 6166 | 0.406 | 0.367 | 0.45 | 0.223 | 0.742 | 39.035 | 0.089 |
| ADR | Mantel-Haenszel RR | 94 | 6376 | 6277 | 0.414 | 0.384 | 0.447 | Not reported | Not reported | Not reported | Not reported |
| ADR | Peto odds ratio | 94 | 6376 | 6277 | 0.316 | 0.288 | 0.348 | Not reported | Not reported | Not reported | Not reported |
| Readmission | Inverse variance, 0.5 only for zero cells | 22 | 1967 | 1956 | 0.524 | 0.457 | 0.601 | 0.417 | 0.659 | 9.969 | 0.008 |
| Readmission | Inverse variance, zero-event trials excluded | 21 | 1905 | 1894 | 0.527 | 0.46 | 0.602 | 0.421 | 0.659 | 9.912 | 0.007 |
| Readmission | Mantel-Haenszel RR | 22 | 1967 | 1956 | 0.497 | 0.444 | 0.555 | Not reported | Not reported | Not reported | Not reported |
| Readmission | Peto odds ratio | 22 | 1967 | 1956 | 0.364 | 0.312 | 0.424 | Not reported | Not reported | Not reported | Not reported |
| Mortality | Inverse variance, 0.5 only for zero cells | 9 | 778 | 771 | 0.531 | 0.391 | 0.721 | 0.391 | 0.721 | 0 | 0 |
| Mortality | Inverse variance, zero-event trials excluded | 7 | 558 | 552 | 0.57 | 0.458 | 0.709 | 0.458 | 0.709 | 0 | 0 |
| Mortality | Mantel-Haenszel RR | 9 | 778 | 771 | 0.497 | 0.308 | 0.8 | Not reported | Not reported | Not reported | Not reported |
| Mortality | Peto odds ratio | 9 | 778 | 771 | 0.49 | 0.305 | 0.789 | Not reported | Not reported | Not reported | Not reported |

Supplementary Table S6A | Core outcomes by outcome-specific RoB 2 subset

| **Outcome** | **Subset** | **k** | **N_Intervention** | **N_Control** | **Effect** | **CI_Lower** | **CI_Upper** | **PI_Lower** | **PI_Upper** | **I2** | **Tau2** |
| --- | --- | --- | --- | --- | --- | --- | --- | --- | --- | --- | --- |
| Adherence-Rate | All eligible trials | 163 | 11529 | 11355 | 1.37 | 1.318 | 1.425 | 0.975 | 1.926 | 80.231 | 0.029 |
| Adherence-Rate | Low-risk trials only | 28 | 2589 | 2648 | 1.337 | 1.217 | 1.468 | 0.957 | 1.868 | 81.683 | 0.024 |
| Adherence-Rate | Other trials | 135 | 8940 | 8707 | 1.379 | 1.32 | 1.441 | 0.969 | 1.963 | 80.088 | 0.031 |
| ADR | All eligible trials | 94 | 6376 | 6277 | 0.405 | 0.367 | 0.448 | 0.222 | 0.739 | 38.333 | 0.089 |
| ADR | Low-risk trials only | 20 | 1598 | 1607 | 0.381 | 0.317 | 0.458 | 0.209 | 0.695 | 35.276 | 0.075 |
| ADR | Other trials | 74 | 4778 | 4670 | 0.411 | 0.365 | 0.464 | 0.218 | 0.775 | 39.403 | 0.098 |
| Readmission | All eligible trials | 22 | 1967 | 1956 | 0.524 | 0.457 | 0.601 | 0.417 | 0.659 | 9.969 | 0.008 |
| Readmission | Low-risk trials only | 4 | 522 | 522 | 0.563 | 0.301 | 1.053 | 0.301 | 1.053 | 0.002 | 0 |
| Readmission | Other trials | 18 | 1445 | 1434 | 0.515 | 0.444 | 0.598 | 0.392 | 0.677 | 14.948 | 0.012 |

Supplementary Table S6B | Interaction tests for outcome-specific low-risk versus other results

| **Outcome** | **k** | **Ratio_of_RRs** | **CI_Lower** | **CI_Upper** | **P_interaction** |
| --- | --- | --- | --- | --- | --- |
| Adherence-Rate | 163 | 0.976 | 0.881 | 1.082 | 0.645 |
| ADR | 94 | 0.911 | 0.717 | 1.158 | 0.444 |
| Readmission | 22 | 1.096 | 0.723 | 1.66 | 0.652 |

Supplementary Table S7 | Baseline-risk absolute-effect and number needed to treat for benefit (NNTB) scenarios

| **Outcome** | **Risk_scenario** | **Control_risk** | **Intervention_risk** | **Absolute difference per 1,000** | **NNTB (range)** | **Follow-up reported** | **Follow-up total** | **Median follow-up, months** | **Follow-up range, months** |
| --- | --- | --- | --- | --- | --- | --- | --- | --- | --- |
| Adherence-Rate | Median | 0.689 | 0.944 | 255.223 | 4 | 85 | 163 | 6 | 1-24 |
| Reported ADRs | P25 | 0.145 | 0.059 | 86 | 12 (11-13) | 45 | 94 | 6 | 1-12 |
| Reported ADRs | Median | 0.233 | 0.094 | 139 | 8 (7-8) | 45 | 94 | 6 | 1-12 |
| Reported ADRs | P75 | 0.362 | 0.146 | 216 | 5 (5-5) | 45 | 94 | 6 | 1-12 |
| Unplanned readmission | P25 | 0.216 | 0.113 | 103 | 10 (9-12) | 21 | 22 | 6 | 1-24 |
| Unplanned readmission | Median | 0.308 | 0.161 | 147 | 7 (6-9) | 21 | 22 | 6 | 1-24 |
| Unplanned readmission | P75 | 0.387 | 0.203 | 184 | 6 (5-7) | 21 | 22 | 6 | 1-24 |
| Mortality | P25 | 0.040 | 0.021 | 19 | 53 (41-89) | 9 | 9 | 12 | 3-12 |
| Mortality | Median | 0.060 | 0.032 | 28 | 36 (28-60) | 9 | 9 | 12 | 3-12 |
| Mortality | P75 | 0.071 | 0.038 | 33 | 30 (24-51) | 9 | 9 | 12 | 3-12 |

Supplementary Table S8A | Baseline characteristics and intervention descriptions of the 266 included trials

Study-identity convention: Trial ID is the canonical identifier for core trials. Author-year labels are provided for readability; No. in Table S8A links to the numbered included-study reference list below. The non-core validation sample in Table S19 uses a separate numbering system.

| **Trial ID** | **No.** | **Year** | **Author** | **Type** | **Disease** | **Sample Size,n,I/C** | | **Age** | **Intervention** | **Comparison** | **Duration of Follow-up** | **Province** |
| --- | --- | --- | --- | --- | --- | --- | --- | --- | --- | --- | --- | --- |
| O001 | 1 | 2015 | Huang | PI | Heart Failure | 58 | 50 | IG: 59.2±16 CG: 64.3±12.8 | Clinical pharmacists formulate individualized medication plans and medication education for patients in the disease management group during hospitalization and conduct telephone follow-up after discharge. | Usual care | 1a | Guangdong |
| O002 | 2 | 2021 | Zhou | MTM | Heart Failure | 45 | 45 | IG: 58±12.6 CG: 59±11.6 | The intervention group received medication therapy management services from pharmacists. | The control group only received routine medication education and guidance. | 6 months | Guangdong |
| O003 | 3 | 2022 | Cheng | PI | Heart Failure | 43 | 62 | IG: 72.6±9.47 CG: 73.66±10.15 | Patients in the experimental group received full-course pharmaceutical services on the basis of those in the control group. | Usual care | 1a | Hubei |
| O004 | 4 | 2016 | Chen | PI | Heart Failure | 90 | 90 | IG: 65.31±9.8 CG: 66.17±10.1 | For patients in the pharmacist management group, on the basis of the control group, clinical pharmacists provide personalized pharmaceutical services such as pharmaceutical care, psychological counseling, medication education, and 6-month discharge follow-up. | Usual care | 6 months | Henan |
| O005 | 5 | 2017 | Pan | PI | Heart Failure | 54 | 53 | IG: 59.65±11.4 CG: 58.28±9.53 | The trial group adopted a clinical pathway management method involving clinical pharmacists. | The control group used conventional chronic heart failure clinical pathway management methods. | 3 months | Shandong |
| O006 | 6 | 2019 | Chen | PI | Heart Failure | 50 | 50 | IG: 44.52±5.79 CG: 43.72±6.42 | In the intervention group, on the basis of routine medication guidance, clinical pharmacists from the hospital provide intervention and guidance on patients' medication compliance. | Patients in the control group received routine bedside and post-discharge medication guidance from doctors and nurses. | 3 months | Guangdong |
| O007 | 7 | 2020 | Chen | PI | Heart Failure | 71 | 50 | IG: 74.41±13.85 CG: 73.42±10.68 | Pharmaceutical intervention, including pharmaceutical care, medication evaluation, regular follow-up, etc. | Usual care | 2a | Jiangsu |
| O008 | 8 | 2020 | Rao | PI | Heart Failure | 80 | 80 | IG: 57.62±15.44 CG: 58.48±16.05 | Patients in the intervention group received chronic disease management by clinical pharmacists such as inpatient pharmaceutical care, discharge medication education, and discharge pharmaceutical follow-up on the basis of the control group. | Usual care | 6 months | Hainan |
| O009 | 9 | 2022 | Liu | PC | Heart Failure | 18 | 22 | IG: 63.66±4.22 CG: 62.11±3.67 | Patients in the research group received pharmaceutical services on the basis of routine treatment. | Usual care |  | Heilongjiang |
| O010 | 10 | 2022 | Wang | PC | Heart Failure | 51 | 43 | IG: 61.25±10.35 CG: 63.93±11.23 | Patients in the pharmaceutical service group receive patient-centered precision medication intervention through the WeChat platform and intelligent software. | Patients in the control group were given routine medication guidance. | 90d | Shandong |
| O011 | 11 | 2007 | Tang | PI | Hypertension | 80 | 80 |  | Use health education interventions during consultation. | Only questionnaire survey without any intervention measures. |  | Guangdong |
| O012 | 12 | 2010 | Zhong | PI | Hypertension | 59 | 59 | MeanAge：61 | The intervention group implements individualized medication, medication guidance and scientific and reasonable intervention measures, etc. | Routine treatment without intervention. | 1a | Guangdong |
| O013 | 13 | 2012 | Zhao | PC | Hypertension | 129 | 129 | IG: 62.4±19.1 CG: 65.6±18.8 | Pharmacist interventions involved recommendations to physicians, educational and counseling directly to the patient | No clinical pharmacist involvement and patients received traditional service provided by the hospital clinic | 6 months | Shaanxi |
| O014 | 14 | 2015 | Zhao | PI | Coronary Disease | 45 | 45 |  | Conventional medical treatment plus interventions by clinical pharmacists. | Conventional medical treatment without pharmacist participation | 6 months | Henan |
| O015 | 15 | 2016 | Gan | PI | Myocardial Infarction | 100 | 100 | Mean Age: 62 | The intervention group was provided pharmaceutical services by clinical pharmacists. | The control group took medication according to routine treatment without intervention. | 12 months | Tianjin |
| O016 | 16 | 2019 | Xu | PI | Coronary Disease | 120 | 120 | IG: 63.24±10.19 CG: 64.12±10.19 | Medication assessment and education by the clinical pharmacist at discharge and telephone follow-ups at 1 week and 1 and 3 months after discharge. | Usual care | 12 months | Zhejiang |
| O017 | 17 | 2022 | Liu | MTM | Multi-Disease | 80 | 72 | IG: 71.12±9.76 CG: 67.9±10.9 | The intervention group was implemented at least one complete MTM service in the pharmacy clinic | The control group received general medication counseling | 6 months | Shanghai |
| O018 | 18 | 2013 | Wong | PI | Hypertension | 92 | 139 | IG: 62.3±8.11 CG: 62.5±10.1 | Participants received both usual care followed by community‐based medication counseling service immediately after physician consultation. | Usual care | 6 months | Hong Kong |
| O019 | 19 | 2013 | Qin | PC | Hypertension | 129 | 129 | MeanAge:18-80 | The intervention group was provided with pharmaceutical care by clinical pharmacists. | The control group did not receive any intervention. |  | Beijing |
| O020 | 20 | 2010 | Qin | PI | Diabetes Mellitus | 27 | 27 | Mean Age: 62 years | On the basis of the control group, pharmacists proactively provide pharmaceutical guidance | Oral conventional diabetes drugs and injectable insulin preparations | 3 months | Shanxi |
| O021 | 21 | 2011 | Li | PC | Diabetes Mellitus | 35 | 33 | IG:62.3 ±16.1 CG:57.1± 11.7 | Clinical pharmacists intervene in patients according to the type 2 diabetes inpatient pharmacy pathway. | Conventional drug treatment |  | Shandong |
| O022 | 22 | 2020 | Zheng | PI | Hypercholesterolemia | 100 | 100 | IG: 56.98±4.05 CG: 57.86±4.1 | The observation group was given clinical pharmaceutical intervention. | Patients in the control group did not take the initiative to receive any drug-related guidance during medication. | 1a | Liaoning |
| O023 | 23 | 2015 | Wan | PI | Hypercholesterolemia | 76 | 76 |  | The trial group provides follow-up pharmaceutical services. In addition to in-hospital medication guidance, the pharmaceutical service team establishes online QQ groups and WeChat Moments with patients or their families, opens contact numbers, and ensures timely acceptance of patient consultations, provision of pharmaceutical services, and regularly make appointments with patients by phone or QQ video to understand their medication status, remind patients to pay attention to adverse drug reactions, publish basic knowledge about hyperlipidemia and its drugs through the Internet, provide door-to-door services every 3 months, and distribute written promotional materials to understand patients Self-manage situations and encourage them to correct their bad lifestyle formula, and solve problems in medication and other aspects. | The control group was provided with routine in-hospital medication consultation. | 18 months | Jiangsu |
| O024 | 24 | 2018 | Zhu | PC | Hypercholesterolemia | 82 | 82 | IG: 48-73 CG: 49-75 | The monitoring group was provided with pharmaceutical care by clinical pharmacists for 3 months. | The control group did not receive relevant intervention. | 3 months | Jiangsu |
| O025 | 25 | 2019 | Xu | PI | Coronary Disease | 98 | 95 | IG: 62.69±11 CG: 62.38±11.23 | Direct pharmacist intervention including medication program optimization, formulated pharmaceutical care, medication education, health education, and outpatient follow-up. | Routine treatment and nursing care | 2a | Shanghai |
| O026 | 26 | 2019 | Wu | PC | Myocardial Infarction | 60 | 60 | IG: 63.3±7 CG: 63.1±7 | The experimental group received pharmaceutical care on the basis of the control group, including the formulation of individualized treatment plans, medication education, adverse reaction monitoring, drug interaction monitoring, discharge medication guidance and regular follow-up. | The patients in the control group received routine post-PCI care, including health education and routine post-operative care. The health education was to inform the selected patients of the significance of antiplatelet drug treatment after PCI surgery and possible complications such as bleeding and thrombosis during treatment. | 1a | Guangdong |
| O027 | 27 | 2021 | Wu | PC | Myocardial Infarction | 100 | 99 | IG: 60.21±9.13 CG: 59.27±10.06 | Patients in the experimental group received pharmaceutical services provided by clinical pharmacists. | Usual care | 12 months | Shandong |
| O028 | 28 | 2021 | Su | PI | Myocardial Infarction | 70 | 70 | IG: 63.28±5.24 CG: 64.13±6.67 | The observation group was provided with professional pharmaceutical intervention by clinical pharmacists on the basis of the control group. | The control group was only given routine medical advice | 12 months | Gansu |
| O029 | 29 | 2022 | Jiang | MTM | Coronary Disease | 65 | 60 | IG: 62.3±8.99 CG: 61.8±8.26 | Patients in the intervention group received standardized MTM services on the basis of the control group. | Patients in the control group received routine medical services. | 12 months | Hebei |
| O030 | 30 | 2022 | Li | PI | Multi-Disease | 30 | 30 | IG: 52.87±5.34 CG: 53.12±5.28 | On the basis of the control group, the research group applied clinical pharmacist intervention under the concept of evidence-based pharmacy, which specifically included: conducting pharmaceutical consultation and evaluation of patients, paying close attention to the patient's disease and blood sugar changes, adjusting medication, medication guidance and discharge education. | The control group adopts traditional blood sugar management on the basis of basic intervention, that is, the blood sugar-lowering related treatment plan is determined by an endocrinologist after consultation or determined by the doctor himself, and then the doctor manages it by himself. |  | Guangdong |
| O031 | 31 | 2022 | Jin | PI | Multi-Disease | 30 | 30 | IG: 53.57±4.32 CG: 52.68±4.21 | Provide pharmaceutical services on the basis of routine treatment, including health education, personalized medication guidance and regular follow-up. | Usual care |  | Zhejiang |
| O032 | 32 | 2022 | Gong | MTM | Multi-Disease | 82 | 90 | IG: 66.23±10.59 CG: 68.24±11.72 | Patients in the intervention group received pharmaceutical care based on the MTM concept within 3 months of enrollment. | Patients in the control group received no special intervention during the study period. | 3 months | Shanghai |
| O033 | 33 | 2021 | Jiang | PC | Multi-Disease | 41 | 41 | IG: 32.03±4.77 CG: 31.73±4.92 | The basic intervention of the research group is the same as that of the control group, and they cooperate to carry out pharmaceutical services, including establishing a pharmaceutical service team, formulating and implementing pharmaceutical pathways and procedures, and discharge guidance. | The control group carried out traditional medical service intervention. | 3 months | Liaoning |
| O034 | 34 | 2021 | Fan | PI | Multi-Disease | 41 | 42 | IG: 56.45±9.68 CG: 56.51±9.42 | The observation group adopted pharmaceutical intervention measures based on the IMB model. | The control group used routine follow-up measures. | 180d | Anhui |
| O035 | 35 | 2020 | Yue | MR | Multi-Disease | 50 | 50 | IG: 62.38±2.11 CG: 63.05±2.59 | Medication Reformulation Interventions. | Basic drug intervention is provided to the patient, and the pharmacist provides routine medication guidance to the patient, and instructs the patient to strictly follow the doctor's instructions for medication. |  | Sichuan |
| O036 | 36 | 2020 | Wang | PI | Multi-Disease | 104 | 104 | IG: 62.38±13.04 CG: 61.45±14.06 | Based on the implementation in the control group, the intervention group added clinical pharmacists to participate in blood glucose management, including pharmaceutical consultation and evaluation, pharmaceutical care, adverse reaction monitoring and patient education. | The control group adopts the traditional blood sugar management model. Clinical pharmacists are not involved in the blood sugar treatment. The surgeon determines the blood sugar lowering plan by himself or asks an endocrinologist to consult and give a treatment plan. The surgeon then manages the blood sugar pattern by himself. |  | Jiangsu |
| O037 | 37 | 2020 | Wang | MTM | Multi-Disease | 150 | 150 | IG: 62.4±5.32 CG: 61.09±6.24 | Drug therapy management services, including the establishment of MTM service groups, rational use of cardiovascular drugs, individualized dosing plans, adverse reaction monitoring and life guidance. | General pharmacy services are adopted. When nurses distribute drugs, pharmacists give medication suggestions, including: drug usage and dosage, precautions, monitoring points, etc. There is no follow-up pharmaceutical service. | 6 months | Heilongjiang |
| O038 | 38 | 2020 | He | PI | Multi-Disease | 35 | 35 | IG: 30.49±4.11 CG: 30.18±4.29 | The observation team provides pharmaceutical services, including establishing and analyzing electronic health records, understanding past medical history and medication history, medication education and regular follow-up. | The control group was provided with traditional medical services, told patients about the necessity of lowering blood sugar, monitored blood sugar levels, and reasonably planned their daily diet, etc. |  | Guangdong |
| O039 | 39 | 2020 | Dai | PC | Multi-Disease | 51 | 50 | IG: 71.34±4.95 CG: 72.33±4.21 | Pharmaceutical care, including indicator testing, ultrasound examination, medication monitoring, antibacterial monitoring, health education and medication promotion | Basic medication guidance |  | Xinjiang |
| O040 | 40 | 2019 | Wang | PI | Multi-Disease | 60 | 60 | IG: 56.33±13.34 CG: 56.55±12.53 | The pharmaceutical service team carries out comprehensive medication education and provides different pharmaceutical services at different stages from admission to discharge. | During the hospitalization period, patients in the control group were only provided with medical order review, case review, patient medication status analysis, medication compliance records, etc. No clinical pharmacists provided pharmaceutical services. |  | Shandong |
| O041 | 41 | 2018 | Yang | PI | Multi-Disease | 58 | 58 | IG: 53.6±3.1 CG: 52.7±3.4 | The observation group carried out pharmaceutical intervention led by clinical pharmacists, including establishing a pharmaceutical care system, carrying out pharmaceutical intervention, and regular follow-up. | The control group received routine medication guidance, and took the medication on time and in accordance with the doctor's instructions and prescriptions. | 6 months | Tianjin |
| O042 | 42 | 2017 | Wang | PC | Multi-Disease | 80 | 80 | IG: 60.6±10.2 CG: 56±13.4 | The observation group is provided with full range of chemical and pharmaceutical services by clinical pharmacists. | In the control group, there were no clinical pharmacists providing pharmaceutical services. |  | Shandong |
| O043 | 43 | 2017 | Jiang | PI | Multi-Disease | 61 | 63 | IG: 55.1±16.7 CG: 49.3±17.1 | Establish a working system that combines hierarchical pharmaceutical care and pharmaceutical pathways, and intervene on patients in accordance with hierarchical pharmaceutical care standards and corresponding pharmaceutical pathway tables. | Usual care | 6 months | Zhejiang |
| O044 | 44 | 2017 | Cao | PI | Multi-Disease | 60 | 60 | IG: 68.6±6.5 CG: 67.1±6.2 | On the basis of conventional treatment, the observation group provides pharmaceutical guidance to patients, including establishing health files, strengthening medication guidance, and regular follow-up. | Usual care |  | Shandong |
| O045 | 45 | 2014 | Hao | PC | Multi-Disease | 34 | 33 | IG: 56.97±13.56 CG: 56.73±15.49 | The intervention group implemented medication consultation and health education. | The control group received no intervention. | 6 months | Anhui |
| O046 | 46 | 2012 | Han | PC | Multi-Disease | 51 | 51 |  | Patients in the experimental group received conventional myocardial infarction treatment and pharmaceutical services provided by pharmacists. | Patients in the control group received conventional myocardial infarction treatment. | 1a | Liaoning |
| O047 | 47 | 2022 | Wei | PI | Multi-Disease | 50 | 50 | IG: 61.9±11.4 CG: 61.7±10.8 | The intervention group accepted pharmacists’ participation in treatment management. | Patients in the control group only undergo routine medical order review, analyze the patient's medication status, record the patient's medication compliance and the patient's awareness of their own disease, etc. This does not include the process of pharmacists providing face-to-face medication guidance to patients and discussing with doctors to optimize prescriptions. | 1a | Guangxi |
| O048 | 48 | 2022 | Zhuo | PI | Multi-Disease | 58 | 61 |  | Pharmacist-led app intervention plus usual care | Usual care |  | Guangdong |
| O049 | 49 | 2022 | Wang | PC | Multi-Disease | 40 | 40 |  | Pharmaceutical care included free access to a clinical pharmacist, education material, a WeChat account for live discussion, and a telephone follow-up | Routine care | 3 months | Shandong |
| O050 | 50 | 2022 | Zhou | PI | Multi-Disease | 40 | 40 | IG: 56.65±6.96 CG: 57±7.51 | The observation group provided pharmaceutical service guidance on the basis of conventional drug treatment. | The control group received conventional drug treatment. |  | Guangdong |
| O051 | 51 | 2022 | Pan | PI | Multi-Disease | 43 | 43 | IG: 66.71±5.32 CG: 67.28±4.96 | Clinical pharmacists provide pharmaceutical intervention, including communicating with patients to improve compliance, reviewing prescriptions, medication guidance, and medication education. | The routine diagnosis and treatment process is given, and the dosage, frequency, precautions, etc. of various drugs are clearly marked on the medicine box, and the attending physician is responsible for guiding the patient's medication during diagnosis and treatment. | 24 weeks | Fujian |
| O052 | 52 | 2022 | Liu | PI | Multi-Disease | 30 | 30 | IG: 61.32±0.46 CG: 60.23±0.24 | Pharmacy intervention includes health education, prescription review, medication guidance intervention and life guidance. | The routine medication guidance is as follows: follow the doctor’s instructions to take medication; mark the medication instructions; explain how to take the medication and dosage, etc. |  | Fujian |
| O053 | 53 | 2021 | Shen | PI | Multi-Disease | 34 | 34 | IG: 57.8±5 CG: 58.4±4.8 | The observation group provides pharmaceutical service intervention, including medication education, prescription review, medication guidance and life guidance. | General medication guidance |  | Shaanxi |
| O054 | 54 | 2013 | Lin | PI | Hypertension | 79 | 79 | IG:55.3±2.4 CG:55.6±2.5 | Clinical pharmacists intervene on the basis of routine treatment. | Conventional antihypertensive medication. |  | Guangdong |
| O055 | 55 | 2015 | Li | PC | Hypertension | 50 | 50 | IG:63.3 CG:64.5 | Patients in the intervention group received personalized pharmaceutical services. | Routine treatment without intervention. | 3months | Shanghai |
| O056 | 56 | 2015 | Zhuo | PI | Hypertension | 92 | 89 |  | Adopt the PDCA cycle method, accept pharmacist intervention and record it. | Routine treatment, no intervention | 6months | Fujian |
| O057 | 57 | 2016 | Zhou | PC | Hypertension | 40 | 40 | IG:57.1±2.0 CG:56.9±2.1 | The observation group received pharmaceutical service intervention on the basis of the control group. | Usual care |  | Hubei |
| O058 | 58 | 2016 | Yin | PI | Hypertension | 100 | 100 | MeanAge:67.5 | The intervention group mainly provides medication guidance and health education through telephone follow-up, and discusses the rationality of patients' medication with clinicians. | Routine treatment, no intervention | 5months | Beijing |
| O059 | 59 | 2016 | He | PI | Hypertension | 85 | 87 | IG:61.40±13.34 CG:60.70±10.95 | Pharmacy Services and Psychological Interventions | Routine treatment, no intervention | 6months | Jiangsu |
| O060 | 60 | 2016 | Zhang | PC | Hypertension | 45 | 45 | IG:42.5±1.5 CG:41.5 ±1.3 | Extended pharmaceutical services | General medication guidance |  | Henan |
| O061 | 61 | 2017 | Peng | PI | Hypertension | 73 | 73 | MeanAge:63.39±11.18 | In the observation group, on the basis of the control group, clinical pharmacists will conduct regular telephone follow-up with the patients twice a month. Patients will be provided with hypertension health knowledge education and medication intervention, with a course of treatment lasting 3 months. | Treat with conventional antihypertensive drugs. |  | Jiangxi |
| O062 | 62 | 2017 | Zhou | PI | Hypertension | 62 | 62 | IG:65.7±1.5 CG:65.9±1.4 | On the basis of the control group, pharmacist intervention was given. | Routine discharge guidance intervention. |  | Zhejiang |
| O063 | 63 | 2017 | Ye | PI | Hypertension | 48 | 49 | IG:57.3±2.3 CG:57.3±2.3 | Patients in the study group received clinical pharmacist guidance on the basis of the control group: 1. Health knowledge guidance; 2. Medication plan formulation; 3. Medication precautions guidance. | Patients in the control group were provided with routine antihypertensive treatment and care. |  | Guangdong |
| O064 | 64 | 2017 | Xu | PI | Hypertension | 49 | 49 | IG:65.19±4.21 CG:65.72± 4.46 | Patients in the observation group applied pharmacist intervention measures, which mainly include the following contents: 1. Medication plan formulation; 2. Health education; 3. Medication guidance; 4. Daily guidance. | Routine care and treatments |  | Jiangxi |
| O065 | 65 | 2017 | Hu | PC | Hypertension | 70 | 70 | IG:45.36±5.17 CG:46.41±5.06 | The experimental group provided full-scale chemical and pharmaceutical services while administering medication. | Usual care |  | Henan |
| O066 | 66 | 2018 | Ren | PC | Hypertension | 45 | 45 | IG:64.28±11.33 CG:64.30±11.29 | Adopt routine pharmaceutical intervention at the drug dispensing window, verbally explain the precautions, usage and dosage, and affix medication labels, and provide systematic pharmaceutical service intervention. | Adopt routine pharmaceutical intervention at the dispensing window, verbally explain precautions, usage and dosage, and affix medication labels. | 6months | Jiangsu |
| O067 | 67 | 2018 | Li | PI | Hypertension | 51 | 51 | MeanAge:73.56±3.58 | Patients in the study group received pharmaceutical service intervention on the basis of those in the control group. | Routine medication according to doctor’s instructions. |  | Jiangsu |
| O068 | 68 | 2018 | Lei | PI | Hypertension | 84 | 84 | IG:67.78±4.22 CG:67.82±4.16 | Strengthen pharmaceutical intervention based on the control group | Using routine intervention, nursing staff routinely administer medications, instruct patients to take medications on time, and conduct regular blood pressure measurements. | 1month | Gansu |
| O069 | 69 | 2018 | Zhang | PI | Hypertension | 60 | 60 | IG:59.3±12.0 CG:60.7±11.2 | Clinical pharmacists provide pharmaceutical care to patients using a drug therapy management model. | The hospital pharmacist will establish health records and conduct regular follow-up visits. | 12months | Guangdong |
| O070 | 70 | 2018 | Lyu | PC | Hypertension | 50 | 50 | IG:51.16±6.21 CG:51.27±6.15 | On this basis, the observation group provided full-course chemical and pharmaceutical services and evaluated the medication compliance of the two groups. | The control group was given routine medication guidance. | 3months | Shanghai |
| O071 | 71 | 2018 | Duan | PI | Diabetes Mellitus | 54 | 54 | IG:54.81±2.11 CG:54.89±2.72 | The pharmaceutical intervention group adopts comprehensive pharmaceutical intervention. | The routine intervention group took routine medication measures. |  | Jiangsu |
| O128 | 72 | 2018 | Ni | PC | Hypertension | 45 | 45 | IG:48.71±2.89 CG:48.34±2.43 | The observation group provides pharmaceutical services. | General medication guidance |  | Jiangsu |
| O073 | 73 | 2019 | Wu | PI | Hypertension | 99 | 99 | IG: 64±4.5 CG: 64±4.5 | Patients in the intervention group were given pharmaceutical intervention, including the formulation of rational medication plans, rational medication education, and regular follow-up. | Patients in the routine group were given routine nursing care. | 3 months | Zhejiang |
| O074 | 74 | 2019 | Hu | PC | Hypertension | 50 | 50 | IG:55.8±5.8 CG:56.3±5.8 | The intervention group received community pharmaceutical service intervention. | Usual care |  | Zhejiang |
| O075 | 75 | 2019 | Feng | PI | Hypertension | 120 | 120 | IG:60.25±6.32 CG:60.16±6.64 | Patients in the intervention group received pharmaceutical service intervention from clinical pharmacists, including differences in patients’ blood pressure, development of good living behaviors, and medication compliance. | Intervention with common medications. | 3months | Guangdong |
| O076 | 76 | 2019 | Zhao | PC | Hypertension | 75 | 75 | IG:46.08±2.15 CG:46.47±2.36 | Carry out long-term individualized clinical pharmaceutical care. | Routine medication guidance and out-of-hospital follow-up | 1a | Jiangsu |
| O077 | 77 | 2019 | Zhang | PI | Hypertension | 65 | 65 | IG:72.64±3.25 CG:72.87±3.13 | Implement standardized pharmaceutical intervention. | Usual care |  | Shanghai |
| O078 | 78 | 2019 | Wang | PI | Hypertension | 80 | 80 | IG:58.87±8.76 CG:59.11±8.65 | The observation group took pharmaceutical intervention on the basis of the control group | The control group received conventional treatment |  | Ningxia |
| O079 | 79 | 2019 | Peng | PI | Hypertension | 48 | 59 | MeanAge:69.85±6.59 | On the basis of routine treatment, clinical pharmacists provide full pharmaceutical care. | Provide routine treatment and follow-up. | 6months | Beijing |
| O080 | 80 | 2019 | He | PC | Hypertension | 25 | 25 | IG:68.64±9.62 CG:69.80 ±7.15 | Clinical pharmacists can implement personalized medication based on the monitoring results of smart blood pressure monitors. | Routine hypertension treatment and management by community medical staff. |  | Guangdong |
| O138 | 81 | 2019 | Yao | PI | Diabetes Mellitus | 40 | 40 | IG:54.76±6.21 CG:54.79±6.67 | Adopt glucose control drug treatment combined with pharmaceutical intervention. | Treat with sugar-control drugs. |  | Sichuan |
| O082 | 82 | 2020 | Yang | PC | Hypertension | 65 | 65 | IG:67.13±2.11 CG:67.21±2.08 | Using pharmaceutical service intervention based on the control group. | Usual care |  | Shanghai |
| O083 | 83 | 2020 | Wu | PC | Hypertension | 87 | 93 | MeanAge:78.57±5.27 | Pharmacists provide pharmaceutical services through telephone follow-up or pharmacy outpatient consultation. The follow-up time is once a month and lasts for 12 months. | Only receive routine medication guidance at the medication distribution window, no follow-up visits | 1a | Jiangsu |
| O084 | 84 | 2020 | Sun | PI | Hypertension | 51 | 51 | IG:59.87±9.70 CG:59.06±9.83 | The intervention group implemented the following measures: building an outpatient hypertension pharmaceutical service team, establishing health records for outpatient hypertension patients, implementing health education, conducting follow-up work, strictly reviewing outpatient prescriptions, and fully communicating with doctors. | Usual care | 6months | Jiangsu |
| O085 | 85 | 2020 | Liu | PI | Hypertension | 42 | 42 | IG:62.12±2.58 CG:62.35± 2.62 | take pharmaceutical interventions | Provide routine medication guidance |  | Jiangsu |
| O086 | 86 | 2020 | Hu | PC | Hypertension | 49 | 49 | IG:62.98±7.36 CG:43.02±8.46 | Pharmacists in the observation group proactively intervened, including establishing a pharmaceutical service team, increasing communication with doctors, establishing health files, regularly conducting education lectures on hypertension knowledge, implementing a multi-modal extended service model, and establishing a regular follow-up system. | Patients in the control group obtained medicines from outpatient pharmacies according to doctor's prescriptions and took medicines according to doctor's instructions or drug instructions without active intervention by pharmacists. | 1a | Jiangsu |
| O087 | 87 | 2020 | Chen | PI | Hypertension | 64 | 64 | IG:72.1±4.6 CG:72.0±4.3 | The observation group provided drug service intervention | The control group received routine intervention | 6months | Shanghai |
| O088 | 88 | 2020 | Cai | PC | Hypertension | 40 | 40 | IG:60.23±6.16 CG:59.62±6.03 | Apply the SIMPLE pharmaceutical service model for management on the basis of routine treatment to observe the blood pressure control effect. | Usual care | 12months | Guangdong |
| O089 | 89 | 2021 | Li | PI | Hypertension | 290 | 298 | Mean Age: 65.98±9.48 | Participants in the intervention group were given interventions from pharmacists, including a monthly review of medications, patient education, and medication adjustment advice to medical doctors over 6 months | Both groups received usual care and participated in the community systematic management program of hypertension | 6 months | Guizhou |
| O090 | 90 | 2021 | Cai | PC | Hypertension | 60 | 60 | IG: 56.52±2.61 CG: 57.55±2.81 | On this basis, patients in the experimental group were provided with pharmacist-led pharmaceutical services, and pharmaceutical services were provided by our hospital’s in-service pharmacists. | Patients in the control group took medication under the guidance of the attending physician during their hospitalization. The main guidance included dosage, method of taking, precautions, etc. |  | Jiangsu |
| O091 | 91 | 2021 | Mei | MTM | Hypertension | 45 | 44 | IG:71.89±6.994 CG:72.32±6.958 | Patients in the intervention group received MTM related services. | Patients in the control group received routine medication guidance. | 6months | Sichuan |
| O092 | 92 | 2021 | Liang | PC | Hypertension | 55 | 55 | MeanAge:65.71±1.20 | Pharmaceutical services involving clinical pharmacists | Provide routine care |  | Guangdong |
| O093 | 93 | 2021 | Chen | PC | Hypertension | 63 | 63 | IG:83.6±2.3 CG:83.0±2.4 | The elderly receive full pharmaceutical services on the basis of conventional hypertension treatment. | Routine treatment of hypertension in the elderly |  | Hainan |
| O094 | 94 | 2022 | Yang | PC | Hypertension | 50 | 50 | MeanAge:60-84 | Patients in the observation group received services from a community pharmacy service team composed of clinicians, clinical pharmacists, and community pharmacists. | Routine medication treatment. | 6months | Shanxi |
| O095 | 95 | 2022 | Ma | MTM | Hypertension | 70 | 70 | IG:66.51±6.39 CG:65.57±6.50 | The observation group will adopt chronic disease management for elderly patients with hypertension on the basis of the control group. | Treatment is carried out according to the hypertension control goals and treatment plan requirements of the 2014 version of the "China Primary Management Guidelines for Hypertension". | 3months | Jiangsu |
| O096 | 96 | 2022 | Guo | PI | Hypertension | 48 | 48 |  | On the basis of the control group, the research group adopted home medication guidance intervention with the participation of pharmacists. | Usual care |  | Tianjin |
| O097 | 97 | 2022 | Dong | PI | Hypertension | 60 | 60 | IG:72.43±3.26 CG:72.04±3.15 | Patients in the intervention group adopted the drug intervention service concept. | General medication guidance | 6months | Jiangsu |
| O098 | 98 | 2022 | Chai | PI | Hypertension | 40 | 40 | IG:66.45±8.64 CG:66.78±8.52 | Add pharmaceutical services and psychological intervention to the control group. | Routine hypertension management |  | Gansu |
| O099 | 99 | 2021 | Zhou | PI | Hypertension | 68 | 68 | MeanAge:73.6 | Patients in the observation group received 12 times of enhanced pharmaceutical service intervention on the basis of the control group. | Implement routine hypertension nursing intervention and medication treatment as prescribed by the doctor |  | Henan |
| O100 | 100 | 2013 | Li | PC | Diabetes Mellitus | 82 | 90 | IG:54.3±13.7 CG:52.1±14.8 | Receive full-service chemical and pharmaceutical services provided by clinical pharmacists. | Patients in the control group received traditional medical services. | 1months | Shandong |
| O101 | 101 | 2013 | Zhao | PI | Diabetes Mellitus | 100 | 100 | IG:65.7±3.2 CG:66.6±4.3 | Using pharmaceutical services simultaneously on the basis of the control group | Conventional drug treatment | 3 months | Tianjin |
| O102 | 102 | 2013 | Yang | PI | Diabetes Mellitus | 40 | 40 | IG:67.4± 2.4 CG:67.4±2.4 | Specialized pharmacists will provide medication guidance based on routine guidance. | Provide routine medication guidance and routine hospitalization treatment |  | Guangdong |
| O103 | 103 | 2014 | Yao | PC | Diabetes Mellitus | 75 | 75 | IG:49.5± 8.2 CG:48.2±9.5 | Implement pharmaceutical care based on the control group | group received traditional inpatient medical services from the Department of Diabetes Medicine |  | Guangdong |
| O104 | 104 | 2014 | Wu | PC | Diabetes Mellitus | 40 | 40 | IG:66.74±2.2 CG:64.55±2.5 | Rational drug use guidance intervention mainly includes the following aspects: 1. Pharmacist guidance 2. Control of drug side effects 3. Optimization of drug specifications and dosage forms 4. Telephone guidance | Take medications as prescribed by your doctor and do not take other intervention measures | 4 months | Guangdong |
| O105 | 105 | 2014 | Wen | PC | Diabetes Mellitus | 50 | 50 | IG:55.2±2.5 CG:55.3±2.6 | Application of pharmaceutical guidance on the basis of the control group. | Usual care | 3 months | Chongqing |
| O106 | 106 | 2014 | Tan | PC | Diabetes Mellitus | 33 | 32 | Mean Age：65.3±6.4 | Provide pharmaceutical guidance, that is, pharmacists provide medication guidance to patients through regular telephone guidance, door-to-door visits, and outpatient consultation. | Receive routine treatment | 6 months | Jiangsu |
| O107 | 107 | 2014 | Shen | PC | Diabetes Mellitus | 54 | 54 | IG:60±7.2 CG:61±7.3 | Clinical pharmacists provide scientific and comprehensive pharmaceutical guidance on the basis of conventional anti-diabetic treatment. | Give only conventional hypoglycemic treatment | 3 months | Shanxi |
| O108 | 108 | 2014 | Pei | PC | Diabetes Mellitus | 50 | 50 | IG:68.9±3.8 CG:68.5±3.2 | Adding pharmaceutical health care intervention to routine care | Implement routine care for diabetes treatment |  | Shandong |
| O109 | 109 | 2014 | Pei | PC | Diabetes Mellitus | 50 | 50 | Mean Age：42.6±7.5 | Provide targeted pharmaceutical services: 1. Diabetes health education 2. Provide medication guidance 3. Pay attention to communication with clinicians | Adopt conventional management | 3 months | Shandong |
| O110 | 110 | 2014 | Jin | PC | Diabetes Mellitus | 30 | 30 | IG:65.5±5.5 CG:66.5 ±4.5 | On the basis of routine treatment, pharmacist guidance was provided to him, mainly through telephone contact and regular door-to-door visits. Strict records were kept of the patient's personal situation and the reasons why the doctor's orders were not implemented, and possible changes in the medication process were recorded. Inform patients promptly of adverse reactions during the process of medication administration. | Treatment using conventional methods, i.e. oral diabetes medications and injections of insulin preparations | 3 months | Yunnan |
| O111 | 111 | 2015 | Zhou | PC | Diabetes Mellitus | 85 | 85 | IG:54.7±6.3 CG:53.7±5.9 | On the basis of the control group, clinical pharmacists will provide comprehensive pharmaceutical guidance, including drug selection, regular health education lectures, telephone tracking and follow-up, etc. | Treat only with diabetes medications | 3 months | Zhejiang |
| O112 | 112 | 2015 | Zhao | PI | Diabetes Mellitus | 42 | 42 | IG:52.5±10.7 CG:53.1± 11.5 | Give pharmaceutical intervention | Conventional pharmacological intervention |  | Shandong |
| O113 | 113 | 2015 | Yang | PI | Diabetes Mellitus | 45 | 45 | IG:48.2±2.43 CG:47.3±3.12 | Carry out pharmaceutical service intervention | Get regular service treatment |  | Henan |
| O114 | 114 | 2015 | Luo | PI | Diabetes Mellitus | 60 | 60 | IG:4６.1 ±8.７ CG:4５.６ ±９.2 | Implement comprehensive pharmaceutical services | General medication guidance |  | Guangdong |
| O115 | 115 | 2015 | Liao | PI | Diabetes Mellitus | 214 | 214 | Mean Age:51±21.5 | Professional pharmaceutical guidance from pharmacists | Provide daily chronic disease management records and follow-up | 12 months | Guangdong |
| O116 | 116 | 2015 | Li | PC | Diabetes Mellitus | 50 | 50 | IG:67 9± 3.4 CG:66.5±3 .3 | Provide pharmaceutical services on the basis of control group | Simply carry out routine clinical treatment |  | Sichuan |
| O117 | 117 | 2015 | Feng | PC | Diabetes Mellitus | 60 | 60 | Mean Age:59.2±8.4 | Provide comprehensive pharmaceutical guidance services on the basis of routine treatment | Usual care |  | Zhejiang |
| O118 | 118 | 2015 | Chen | PC | Diabetes Mellitus | 105 | 58 | Group education group: 68.5± 9.3 Home visit group: 65.6±8.3 CG:63.2±7.3 | Adopt door-to-door service or telephone follow-up method | Pharmaceutical intervention through traditional drug dispensing window | 6 months | Guangdong |
| O119 | 119 | 2016 | Yu | PC | Diabetes Mellitus | 78 | 78 | IG:58.91±10.34 CG:60.22±11.61 | On the basis of traditional medical services, we provide comprehensive internal pharmacy services through diabetes-related knowledge education and medication education. | Usual care |  | Inner Mongolia |
| O120 | 120 | 2016 | Wang | PC | Diabetes Mellitus | 44 | 44 | IG:67.4±4.2 CG:67.4±4.1 | The specific service content of using pharmaceutical services to guide is as follows (1) health education (2) explanation of adverse reactions (3) guidance on correct medication use (4) assessment of patient conditions and medication use | General medication guidance |  | Hubei |
| O121 | 121 | 2016 | Sun | PC | Diabetes Mellitus | 129 | 129 | IG:53.0±4.7 CG:54.0±4.9 | On the basis of the regular group, targeted pharmaceutical guidance intervention will be implemented at the same time. | Conventional diabetes treatments | 6 months | Inner Mongolia |
| O122 | 122 | 2016 | Dong | PC | Diabetes Mellitus | 40 | 40 |  | Carry out medication intervention | Routine medication treatment |  | Jilin |
| O123 | 123 | 2016 | Zeng | PI | Diabetes Mellitus | 50 | 50 | IG:45.8 ±9.0 CG:45.7 ±8.8 | Pharmaceutical intervention is organized and completed by clinical pharmacists, including: (1) establishment of personal health files (2) medication guidance (3) pharmaceutical care | Routine medication intervention | | Xinjiang |
| O124 | 124 | 2017 | Wen | PC | Diabetes Mellitus | 40 | 40 | IG:61.33±11.29 CG:60.75±10.49 | Provide pharmaceutical service intervention, mainly including: 1 health education 2 medication intervention 3 strengthening communication with doctors | Provide routine clinical treatment services, that is, only explain the usage and dosage of drugs to patients, distribute health education brochures, etc. |  | Chongqing |
| O125 | 125 | 2017 | Wang | PC | Diabetes Mellitus | 192 | 192 | IG:56±12 CG:56±13 | Add the intervention of clinical pharmacists to the original chronic disease management methods | Continue to manage chronic diseases as usual | 3 months | Guangdong |
| O126 | 126 | 2017 | Pan | PC | Diabetes Mellitus | 71 | 71 | IG: 57.06±9.11 CG:55.77±9.86 | On the basis of routine medical services, we provide medication education, telephone follow-up and other extended pharmaceutical services inside and outside the hospital. | Routine pharmacy services |  | Jiangsu |
| O127 | 127 | 2017 | Han | PC | Diabetes Mellitus | 46 | 46 | IG:56.8±8.3 CG:57.6±10.2 | On this basis, pharmaceutical services such as medication education and telephone follow-up are provided. | Routine treatment services | 3 months | Shanghai |
| O129 | 128 | 2018 | Tan | PI | Diabetes Mellitus | 40 | 40 | IG:55.34±4.28 CG:53.64±1.34 | Give pharmaceutical intervention | Routine medication intervention | 3 months | Sichuan |
| O130 | 129 | 2018 | Lyu | PC | Diabetes Mellitus | 50 | 50 | IG:49.50±10.73 CG:49.62±10.82 | Implement comprehensive pharmaceutical intervention: 1. Health record establishment 2. Medication intervention 3. Pharmaceutical monitoring 4. Post-discharge intervention | Implement routine medication measures |  | Jiangsu |
| O131 | 130 | 2018 | Li | PC | Diabetes Mellitus | 50 | 50 | IG:76±6.87 CG:51.82±6.9 | Use pharmacy services | Implement routine medication measures |  | Shandong |
| O132 | 131 | 2018 | Huang | PC | Diabetes Mellitus | 50 | 50 | IG:61.75± 5.16 CG:60.52±4.36 | Application of pharmaceutical services 1. Medication education 2. Telephone follow-up | Conventional drug treatment |  | Fujian |
| O133 | 132 | 2018 | Hou | PC | Diabetes Mellitus | 51 | 51 | IG:45.4±3.2 CG:45.8±3.5 | During outpatient drug use, clinical pharmacists formulate individualized dosage plans based on the patient's specific conditions. | Clinical pharmacists did not provide pharmaceutical services during outpatient drug use | 6 months | Shandong |
| O134 | 133 | 2018 | Guo | PC | Diabetes Mellitus | 85 | 85 | IG:56.33±2.19 CG:56.40±2.11 | Pharmacy services will be provided on the basis of the control group, as follows: 1. Prescription review 2 Rational drug use intervention 3 Diabetes health education 4 Follow-up and tracking | Provide routine diabetes care |  | Liaoning |
| O135 | 134 | 2018 | Fang | PC | Diabetes Mellitus | 39 | 39 | IG:67.29±3.27 CG:67.35±3.14 | The observation group was given pharmaceutical guidance and education on the basis of the control group. 1 Establish medication files 2 Pharmacy education; 3 Telephone education; 4 Home visits twice a month; 5 Active encouragement | Provide routine guidance: every time the patient takes medicine, inform him or her of the method of use. If the patient asks for advice, answer the questions patiently. | 3 months | Henan |
| O136 | 135 | 2018 | Du | PC | Diabetes Mellitus | 30 | 30 | Mean Age：49±6.1 | Implement pharmaceutical monitoring service methods formulated by pharmacists on the basis of the control group | Routine medical care services |  | Jiangsu |
| O137 | 136 | 2018 | Chen | PI | Diabetes Mellitus | 50 | 50 | IG:67.53±1.79 CG:69.23±1.44 | The implementation of pharmaceutical intervention and comprehensive pharmaceutical guidance specifically includes the following measures: 1. Provide multi-level clinical medication guidance; 2. Timely assessment of medication compliance; 3. Implement telephone guidance and patient visits. | Routine diabetes treatment |  | Jiangsu |
| O139 | 137 | 2019 | Wang | PC | Diabetes Mellitus | 50 | 50 | IG:55.89±2.72 CG:54.81±2.76 | Clinical pharmacists participate to provide pharmaceutical services: 1. Provide diabetes health education to patients. 2 Provide guidance to patients on rational medication use. 3 Clinical pharmacists formulate medication plans 4 Supervise patients’ medication use | General medication guidance, clinical pharmacists are not involved. |  | Jiangsu |
| O140 | 138 | 2019 | Mi | PC | Diabetes Mellitus | 65 | 60 | IG:51.07 ± 8.75 CG:51.24 ± 8.55 | On the basis of the control group, the following specifications were implemented: chemical and pharmaceutical service intervention measures, establishment of health records, and promotion of basic diabetes knowledge: correct medication guidance, pharmaceutical services and supervised medication follow-up. | General medication guidance | 6 months | Xinjiang |
| O141 | 139 | 2019 | Liu | MTM | Diabetes Mellitus | 40 | 36 | IG:56.21±4.65 CG:57.30±5.38 | Accept the "medicine-drug-nursing" comprehensive service model | Accepting traditional medical services is the "medical-nursing" service model. | 3 months | Guangdong |
| O142 | 140 | 2019 | Liang | PC | Diabetes Mellitus | 70 | 70 | IG:45.9±5.3 CG:46.4±4.3 | Provide pharmaceutical guidance based on evidence-based support. The specific measures are as follows: 1. Establish an evidence-based group. 2 Ask evidence-based questions.3 Evidence-based support4 Evidence-based application | Provide routine medication guidance |  | Chongqing |
| O143 | 141 | 2019 | Li | PC | Diabetes Mellitus | 60 | 60 | IG:55.29±11.25 CG:54.48±10.43 | Receive multi-drug combination treatment, and the observation group will receive drug intervention on this basis. | Provide routine medication guidance |  | Shandong |
| O144 | 142 | 2019 | Du | PC | Diabetes Mellitus | 79 | 63 | Mean Age:65.5±15.5 | The work of the intervention team is as follows: 1. Specialist clinical pharmacists work in the department to evaluate patients' pharmacy in a targeted and individualized manner, and work with doctors to formulate drug intervention plans. 2. Strengthen medication education. 3. Regularly organize medication education. 4. Carry out monitoring pharmacy, strictly observe the patient's condition and vital signs after medication, and pay attention to adverse drug reactions. 5. Regular telephone follow-up and medication guidance. | Conventional treatment |  | Gansu |
| O145 | 143 | 2019 | Chen | PC | Diabetes Mellitus | 45 | 45 | IG:48.24±12.81 CG:48.90±12.21 | Pharmaceutical service strategies | Conventional treatment |  | Shandong |
| O146 | 144 | 2019 | Cai | PC | Diabetes Mellitus | 50 | 50 | IG:54.88 ± 4.89 CG:54.14±6.55 | Apply the methods and principles of clinical pathways and establish PCP for patients with type 2 diabetes according to the characteristics of diabetes treatment | Provide routine management | 6months | Hubei |
| O147 | 145 | 2018 | Zhi | PC | Diabetes Mellitus | 55 | 55 | IG:58.4±2.3 CG:58.1±2.6 | Adopt full-course chemical pharmaceutical intervention during the treatment process | Routine intervention methods, including traditional medical services and pharmaceutical services in hospitals |  | Shandong |
| O148 | 146 | 2020 | Zhao | PC | Diabetes Mellitus | 42 | 42 | IG:56.5±4.1 CG:56.9±4.2 | Pharmaceutical service intervention methods for treatment 1 Medication intervention 2 Regular service intervention 3 Health education | Treat with medicines prescribed by traditional doctors |  | Shandong |
| O149 | 147 | 2020 | Xin | PI | Diabetes Mellitus | 50 | 50 | IG:57.1±6.0 CG:56.8±5.7 | On the basis of the control group, pharmacist intervention nursing measures were given. The detailed measures are as follows: 1. Psychological care. 2 Dietary care. 3 Medication care 4 Health education 5 Daily care 6 Follow-up | Routine diagnosis and treatment services |  | Shandong |
| O150 | 148 | 2020 | Xin | PC | Diabetes Mellitus | 10 | 31 | IG:49.12 ± 15.52 CG:44.10 ± 14.26 | Pharmacists participate in chronic disease management and provide pharmaceutical intervention to patients | General medication guidance | 6 months | Beijing |
| O151 | 149 | 2020 | Shang | PC | Diabetes Mellitus | 100 | 100 | IG:70.12±6.94 CG:70.06±6.84 | On this basis, clinical pharmacists intervene: 1. Provide medication guidance; 2. Diabetes health education. 3 Pay attention to communication with clinicians. 4. Monitor the changes and development of the disease course. | General medication guidance | 12 months | Shanghai |
| O152 | 150 | 2020 | Ma | PI | Diabetes Mellitus | 50 | 50 | IG:49.7±4.2 CG:46.8±3.5 | The specific content of pharmaceutical intervention includes: 1. Setting up a special organization composed of clinical pharmacists; 2. Establishing personal health files for patients; 3. Providing medication guidance to patients; 4. Patiently communicating with patients; 5. Carry out pharmaceutical care of patients. | Routine medication intervention |  | Qinghai |
| O153 | 151 | 2020 | Li | PC | Diabetes Mellitus | 66 | 66 | IG:57.15±6.92 CG:56.79±7.81 | On the basis of the control group, a pharmaceutical care model with pharmaceutical service characteristics was provided. | Routine monitoring, including basic monitoring during hospitalization, such as blood sugar control, medication use guidance, etc. | 6months | Hebei |
| O154 | 152 | 2020 | Li | PC | Diabetes Mellitus | 70 | 70 | IG:56.21±8.06 CG:57.65±8.94 | Implement and strengthen pharmaceutical services: (1) Strengthen the popularization of drug knowledge (2) Formulate and design personalized medication plans (3) Pay close attention to special patients (4) Strictly follow the pharmacoeconomic perspective in the medication process | Patients in the control group were managed with conventional methods, such as correct medication use, effective control of patients’ blood sugar levels, life management, etc. |  | Shandong |
| O155 | 153 | 2020 | Li | PC | Diabetes Mellitus | 37 | 37 | IG:61.4±8.2 CG:62.1±7.9 | Adopt Pharmacy Services Guidance | General medication guidance |  | Hubei |
| O156 | 154 | 2020 | Huang | PC | Diabetes Mellitus | 40 | 40 | Mean Age:64.52±4.36 | In addition, personalized pharmaceutical services are provided: 1. Medication education 2. Follow-up management | General medication guidance |  | Zhejiang |
| O157 | 155 | 2019 | Zhu | PI | Diabetes Mellitus | 50 | 50 | IG:52.0 ± 4.2 CG:52.5 ± 4.8 | Drug intervention is completed by clinicians and pharmacists: 1. Set up a drug intervention team and determine the theme 2. Establish personal files 3. Medication guidance 4. Pharmaceutical monitoring | Routine medication intervention, including medication dosage, drug name, medication time, medication method, medication frequency, etc., mainly oral health education |  | Sichuan |
| O158 | 156 | 2020 | Chen | PC | Diabetes Mellitus | 100 | 100 | IG:46.52±7.28 CG:46.44±7.22 | On the basis of the control group, pharmaceutical services are provided | Conventional treatment |  | Jiangsu |
| O159 | 157 | 2021 | Zhou | PI | Diabetes Mellitus | 50 | 50 | IG:8.52±6.4 CG:8.46±6. | Analyze and guide patients’ medication use, and record patients’ medication use | General medication guidance | 2months | Yunnan |
| O160 | 158 | 2021 | Zhao | PI | Diabetes Mellitus | 91 | 91 | IG:62.79±15.38 CG:63.28±14.76 | The intervention group implemented the following measures: established a clinical pharmacy service team, established health records for outpatient diabetes patients, increased communication with doctors, regularly conducted health lectures and education on diabetes medication knowledge, strengthened drug prescription review, implemented multi-modal antidiabetic drug guidance, and conducted regular follow-up visits. | Only receive routine medication guidance, including introduction to medications and explanations of treatment methods |  | Jiangsu |
| O161 | 159 | 2021 | Yu | PC | Diabetes Mellitus | 50 | 50 | IG:55.6±4.3 CG:54.6±4.2 | On the basis of the regular group, clinicians will be involved in the management of standardized diabetes treatment. | Provide patients with routine medical service management, and provide patients with blood sugar monitoring, health education and routine medication plans in accordance with the diagnosis and treatment plan of the attending physician. |  | Xinjiang |
| O162 | 160 | 2021 | Wang | PI | Diabetes Mellitus | 50 | 50 | IG:62.50±2.50 CG:62.75±2.75 | Pharmacist intervention was used on the basis of the control group | Receive medication guidance and medical services from clinical medical staff |  | Jiangsu |
| O163 | 161 | 2021 | Tian | PI | Diabetes Mellitus | 82 | 82 | IG:54.17±4.64 CG:55.28±3.19 | Clinical pharmacists participate in chronic disease management | Apply routine chronic disease management |  | Ningxia |
| O164 | 162 | 2021 | Song | PI | Diabetes Mellitus | 80 | 80 | IG:50.14±8.64 CG:49.19±12.19 | Clinical pharmacists provide pharmaceutical services to patients in the intervention group | Receive MMC standardized diagnosis and treatment | 3months | Jiangsu |
| O165 | 163 | 2021 | Niu | PI | Diabetes Mellitus | 30 | 30 | IG:69.25±3.74 CG:69.22±3.33 | Pharmaceutical intervention based on the control group | Multidrug combination therapy |  | Jiangsu |
| O166 | 164 | 2021 | Ma | PC | Diabetes Mellitus | 60 | 60 | IG:65.18±7.62 CG:67.21±8.34 | Professional pharmaceutical services | Routine medical advice and care |  | Jiangsu |
| O167 | 165 | 2021 | Liang | PI | Diabetes Mellitus | 50 | 50 | IG:62.4±2.1 CG:63.2±2.7 | Pharmaceutical service intervention in the observation group | For routine medication, pharmacists guide patients on medication time and precautions. |  | Guangdong |
| O168 | 166 | 2021 | Li | PC | Diabetes Mellitus | 66 | 66 | IG:55.00±4.59 CG:55.22± 4.37 | Adopt chronic disease management services led by clinical pharmacists. Specific contents include: 1. Pharmaceutical monitoring during hospitalization; 2. Medication education upon discharge; 3. Follow-up after discharge. | Take traditional medical services | 6months | Hebei |
| O169 | 167 | 2021 | Hong | PC | Diabetes Mellitus | 50 | 50 | IG:69.12±5.4 CG:54±5.08 | Patients in the observation group added active intervention by clinical pharmacists on the basis of the original chronic disease management model. | Our hospital's traditional chronic disease management model is adopted, with doctors and nurses providing medication guidance, and clinical pharmacists not actively providing pharmaceutical services. | 6months | Fujian |
| O170 | 168 | 2021 | Geng | PI | Diabetes Mellitus | 61 | 61 | IG:58.98±5.25 CG:58.32±5.74 | Give pharmaceutical intervention | Routine medication intervention |  | Heilongjiang |
| O171 | 169 | 2021 | Duan | PI | Diabetes Mellitus | 50 | 50 | IG:60.12±5.36 CG:60.25±5.21 | Pharmacist intervention methods | Routine intervention measures are taken, and the pharmacist distributes medicines to patients, explains the dosage and usage of the medicines, and answers questions raised by patients, etc. |  | Jiangsu |
| O172 | 170 | 2021 | Dai | PI | Diabetes Mellitus | 68 | 68 | IG:58.12±7.39 CG:58.57±8.05 | A T2DM medication guidance clinic has been opened. Our hospital's endocrinology professional clinical pharmacists with the title of pharmacist or above provide medication guidance for each patient diagnosed with T2DM. They explain the uses of insulin, the use of insulin injection pens, storage methods, and medications according to the doctor's prescription, adverse reactions, usage precautions, etc. | Traditional health education is adopted. After the doctor writes a prescription, he explains the use of insulin to the patient. The nurse simulates the insulin injection process to the patient, distributes an instruction manual for insulin use, and instructs him to follow up if he feels unwell. He also conducts a follow-up phone call once a month to understand the blood sugar control situation, simple health promotion | 3months | Anhui |
| O173 | 171 | 2021 | Chi | PI | Diabetes Mellitus | 44 | 44 | IG:66.45±5.76 CG:66.27±5.91 | Receive clinical pharmacist intervention | For routine medical advice, medical staff will introduce to patients the time to take the medication, frequency of use, and relevant precautions during medication based on the prescription. |  | Fujian |
| O174 | 172 | 2022 | Zhao | PC | Diabetes Mellitus | 55 | 55 | IG:55.70±7.98 CG:55.65±8.03 | On this basis, accept pharmaceutical services from clinical pharmacists, including: 1. Pharmaceutical knowledge education 2. Drug interaction management 3. Adverse drug reaction prevention and control 4. Establishing a follow-up mechanism for pharmaceutical services. | General medication guidance | 3months | Shandong |
| O175 | 173 | 2022 | Zhang | PC | Diabetes Mellitus | 60 | 60 | IG:10.85±0.31 CG:11.00±0.28 | Implement clinical pharmacy guidance. | Routine medication guidance, pharmacists need to inform children and parents to follow the doctor's instructions to take medication. |  | Guangdong |
| O176 | 174 | 2022 | Yin | PC | Diabetes Mellitus | 50 | 50 | IG:75.56±2.33 CG:69.78±3.54 | In addition to drug treatment, professional clinical pharmacists provide guidance. | Medications and general nursing care. | 12months | Jiangsu |
| O177 | 175 | 2022 | Wu | PC | Diabetes Mellitus | 50 | 50 | IG:52.6±4.4 CG:48.8±5.6 | The experimental group fully complied with the treatment model of the control group and cooperated with community pharmaceutical service intervention methods, including 1. establishing patient personal information files. 2. regularly conducting community lectures 3. organizing community health activities 4. pharmacist planning medication guidance 5. regular follow-up. | Conventional treatment modalities for type 2 diabetes. | 5months | Beijing |
| O178 | 176 | 2022 | Wang | PC | Diabetes Mellitus | 30 | 30 | IG:68.49±5.72 CG:68.51±5.37 | On the basis of the control group, family pharmacists' clinical pharmaceutical service intervention was added: (1) medical order review. (2) pharmaceutical monitoring (3) medication guidance. (4) Family follow-up. | Usual care | 6months | Sichuan |
| O179 | 177 | 2022 | Sun | PC | Diabetes Mellitus | 50 | 50 | IG:58.8±6.0 CG:58.4±6.4 | Providing community pharmacy services interventions | Provide medication guidance at routine drug distribution windows | 6 months | Beijing |
| O180 | 178 | 2014 | Lin | PI | Multi-Disease | 150 | 150 | Mean Age: 62.1±11.6 | Pharmacist intervention, including formulation of individualized medication, detection of adverse reactions and regular follow-up. | The control group was not given any intervention measures. |  | Guangdong |
| O181 | 179 | 2022 | Rao | PC | Diabetes Mellitus | 48 | 49 | IG:44.07±13.12 CG:45.56±15.60 | In addition to routine medical treatment, professional pharmaceutical guidance and follow-up are provided. | Follow normal medical procedures. | 3months | Jiangsu |
| O182 | 180 | 2022 | Li | PC | Diabetes Mellitus | 35 | 35 | IG:53.28±4.22 CG:54.29±4.24 | Pharmaceutical Services Management | Routine medication management |  | Shandong |
| O183 | 181 | 2022 | Hu | PC | Diabetes Mellitus | 150 | 150 | IG:51.65±1.62 CG:51.32±1.65 | The observation group received pharmaceutical services | Receive routine medication guidance |  | Jiangsu |
| O184 | 182 | 2022 | Guo | PC | Diabetes Mellitus | 46 | 46 | IG:63.56±7.52 CG:63.51±7.46 | Pharmaceutical services were provided based on individualized management of patients in the control group. | Intervention using simple personalized management |  | Hebei |
| O185 | 183 | 2022 | Du | PI | Diabetes Mellitus | 63 | 63 | IG:59.60±9.17 CG:59.14±8.33 | The research team provided pharmaceutical intervention in the hospital pharmacy on the basis of the reference group. | Conventional pharmacological intervention |  | Guangdong |
| U001 | 184 | 2026 | Gao | PC | Coronary Disease | 30 | 30 | IG: 64.67±7.31 CG: 65.1±8.62 | The intervention group received the CPC programme | Standard care without additional pharmaceutical interventions | 1 month | Beijing |
| U002 | 185 | 2023 | Wang | PI | Coronary Disease | 82 | 80 | IG: 64.1±10 CG: 63.2±12.1 | On the basis of the control group, patients in the intervention group received a home pharmacist service model. The home pharmacists were served by four pharmacists who had obtained clinical pharmacist certificates in our hospital. They discussed with the cardiology clinicians and formulated unified follow-up standards. | Patients in the control group received routine discharge health education, including guidance on lifestyle changes, regular review, rehabilitation exercises, medication precautions, etc., and regular telephone or outpatient follow-up. |  | Chongqing |
| U005 | 186 | 2023 | Xiao | PI | Diabetes Mellitus | 136 | 131 | IG: 54.1±9.8 CG: 53.2±14 | Patients in the physician-pharmacist collaborative clinics received pharmaceutical care at each followup visit, including diabetes education, medication guidance, lifestyle intervention, treatment of adverse drug reactions and identification of complications. | There is no intervention provided by pharmacists in usual clinics. | 12 months | Hunan |
| U013 | 187 | 2023 | Miu | PI | Diabetes Mellitus | 100 | 100 | IG: 71.2±4.3 CG: 70.5±3.8 | The intervention group added online and offline pharmacist services to conventional pharmacist services. | The control group received routine pharmacist services, that is, after a doctor issued a prescription, they went to the pharmacy to pick up the medicine. The pharmacist distributed and provided guidance on the usage, dosage and precautions of oral medicines. |  | Fujian |
| U017 | 188 | 2023 | Wang | PI | Diabetes Mellitus | 300 | 300 | IG: 64.39±2.47 CG: 64.21±2.52 | Pharmacist-led remote medication guidance | Traditional medication guidance requires hospital pharmacists to provide corresponding drug treatment and simple medication guidance based on the patient's clinical symptoms, including instructing to take medications in a timely and quantitative manner, and assisting in indicating medication time, dosage, and specific drug types, etc. | 6 months | Hebei |
| U018 | 189 | 2023 | Lou | PI | Diabetes Mellitus | 60 | 60 | IG: 70.32±2.5 CG: 69.27±4.02 | Carry out pharmacist intervention based on IMB model | Routine follow-up management | 6 months | Zhejiang |
| U010 | 190 | 2024 | Li | MTM | Diabetes Mellitus | 40 | 40 | IG: 60.72±12.54 CG: 59.52±12.22 | Receive pharmaceutical care based on the traditional model | Receive traditional "medical-nursing" medical services | 3 months | Liaoning |
| U012 | 191 | 2024 | Zhang | PI | Diabetes Mellitus | 98 | 96 | IG: 77±8.5 CG: 77.35±8.69 | In the intervention group, the pharmacists of the South Hospital (who must first undergo training and pass the assessment) carry out standardized (online and offline) pharmacist services based on the contract signed by the family pharmacist. | The control group only provided traditional window pharmacy services | 3 months | Chongqing |
| U016 | 192 | 2024 | Ning | PI | Diabetes Mellitus | 200 | 200 | IG: 53.64±11.8 CG: 54.3±11.27 | On the basis of the control group, the intervention group adopted the MDT model with clinical pharmacists as the core. | The control group received routine diabetes chronic disease education. During hospitalization, the attending physician and responsible nurse provided the patients with dietary and exercise guidance, medication education, and treatment of adverse reactions. | 12 months | Guangxi |
| U008 | 193 | 2025 | Zhang | PI | Diabetes Mellitus | 100 | 100 | IG: 54.36±8.42 CG: 55.28±8.72 | Perioperative blood glucose management intervention based on ERAS theory with participation of pharmacists from primary hospitals on a routine basis | Routine perioperative blood glucose management intervention |  | Guangdong |
| U009 | 194 | 2025 | Ma | PC | Diabetes Mellitus | 66 | 66 | IG: 60.62±8.21 CG: 59.81±9.35 | Pharmaceutical care of patients based on PCNE classification system | Routine medical monitoring | 3 months | Qinghai |
| U015 | 195 | 2025 | Zhang | PI | Diabetes Mellitus | 103 | 103 | IG: 63.8±10.1 CG: 64.6±9.3 | On the basis of conventional pharmacist services, home pharmacist services based on family hospital beds are added. | Patients receive routine pharmacist services and receive basic medication consultation and guidance at traditional medication distribution windows. | 9 months | Guangdong |
| U006 | 196 | 2026 | Xiao | PI | Diabetes Mellitus | 291 | 283 | IG: 54.0 (49.0-60.0) CG: 55.0 (46.0-67.0) | Centers in the intervention group provided routine physician-led therapy combined with pharmacist-delivered pharmaceutical care through outpatient visits at baseline and at the third, sixth, ninth, and 12th months | Patients in the control centers received usual treatment and management by physicians, with no pharmacist available to provide additional patient education. | 12 months | Hunan |
| U007 | 197 | 2026 | Zhang | PI | Diabetes Mellitus | 60 | 60 | IG: 61.52±7.36 CG: 60.89±7.51 | On the basis of the conventional intervention group, pharmacist services based on the KAP model are provided | Standardized anti-diabetic treatment and health education | 3 months | Guangxi |
| U014 | 198 | 2026 | Dong | PI | Diabetes Mellitus | 40 | 40 | IG: 67.82±2.18 CG: 67.12±2.38 | The intervention group implemented medication education led by pharmacists | The control group received routine medication guidance. | 3 months | Zhejiang |
| U019 | 199 | 2024 | Lai | PI | Dyslipidemia | 50 | 50 | IG: 53.02±6.59 CG: 53.15±6.72 | Lipid-lowering treatments and pharmacist interventions | Conventional lipid-lowering treatment | 12 months | Xinjiang |
| U020 | 200 | 2024 | Chen | PI | Myocardial Infarction | 56 | 56 | IG: 60.17±5.03 CG: 61.36±5.84 | Accept ALHA pharmacist intervention combined with drug reformulation based on the iceberg theory | The control group received routine clinical pharmacist intervention | 6 months | Jiangsu |
| U021 | 201 | 2025 | Chen | PI | Myocardial Infarction | 36 | 36 | IG: 74.3±5.8 CG: 74.1±6.4 | Provide refined clinical pharmacist-led pharmacist intervention based on the control group | Conventional pharmacist intervention model | 3 months | Anhui |
| U023 | 202 | 2023 | Wei | PC | Heart Failure | 140 | 143 | IG: 56.47±13.66 CG: 57.24±12.71 | Graded pharmaceutical care provided by pharmacists | Treated individually by a doctor | 12 months | Jiangsu |
| U025 | 203 | 2023 | Li | PI | Heart Failure | 46 | 46 | IG: 61.07±11.49 CG: 62.35±10.11 | Disease management is led by clinical pharmacists and is jointly carried out by specialist physicians, clinical pharmacists and nurses. | Routine discharge education | 6 months | Fujian |
| U022 | 204 | 2025 | Xiong | PI | Heart Failure | 30 | 30 | IG: 72.2±8.41 CG: 67.5±8.32 | Patients in the intervention group were jointly discussed and selected by clinical pharmacists and clinicians to select a clinical drug path for CHF treatment that is suitable for our hospital. The patients in the intervention group received full medication monitoring during their hospitalization, starting from the confirmation that the patient was the subject of monitoring until the patient was discharged with the treatment target. | Patients in the control group were managed according to the routine treatment plan of the cardiology department. |  | Heilongjiang |
| U024 | 205 | 2026 | Hou | PI | Heart Failure | 113 | 113 | IG: 66.42±12.51 CG: 67.02±13.32 | CPP interventions provided by clinical pharmacists include medication monitoring during hospitalization, development of individualized discharge medication plans, and pharmaceutical follow-up after discharge, etc. | conventional treatment | 6 months | Inner Mongolia |
| U030 | 206 | 2023 | Wang | PI | Hypertension | 40 | 40 | IG: 72.19±6.82 CG: 71.42±6.51 | drug service intervention | General medication guidance |  | Hebei |
| U027 | 207 | 2024 | Li | PI | Hypertension | 89 | 84 | IG: 50.92±8.16 CG: 52.74±6.83 | The intervention group implemented the "Internet +" pharmacist service model based on the control group. | The control group received routine pharmacist services | 6 months | Shanghai |
| U031 | 208 | 2024 | Wu | MTM | Hypertension | 148 | 159 | IG: 64.88±7.66 CG: 66.11±7.61 | Standard Medication Therapy Management Services | Provides traditional pharmacist services and does not provide active intervention | 6 months | Beijing |
| U026 | 209 | 2025 | Zhang | PI | Hypertension | 41 | 41 | IG: 61.47±2.45 CG: 61.81±2.57 | Receive 3 months of "Internet +" pharmacist services on the basis of the control group | Subject to routine pharmacist service intervention, the pharmacist will provide discharge medication guidance before discharge. | 3 months | Shanxi |
| U028 | 210 | 2025 | Sun | MTM | Hypertension | 199 | 199 | IG: 70.86±5.55 CG: 69.53±7.02 | Smart MTM services provided by home pharmacists | Intervention measures mainly use routine follow-up and health education | 6 months | Jiangsu |
| U029 | 211 | 2026 | Hu | PI | Hypertension | 57 | 57 | IG: 56.77±4.32 CG: 56.32±4.51 | Receive enhanced pharmacist intervention on the basis of the conventional group | General medication guidance | 3 months | Hubei |
| U033 | 212 | 2023 | Han | PC | Multi-Disease | 97 | 98 | IG: 60.65±14.11 CG: 60.09±10.6 | Based on the traditional treatment of "doctor-nurse" team, pharmacists are added to participate in the diagnosis and treatment of patients. | Traditional treatment using a "doctor-nurse" team | 3 months | Hebei |
| U035 | 213 | 2023 | Li | MTM | Multi-Disease | 51 | 50 | IG: 64.06±9.43 CG: 63.42±9.06 | a standard MTM service was given to the intervention group | General services for noninfectious chronic diseases | 12 months | Jiangsu |
| U032 | 214 | 2024 | Xu | PI | Multi-Disease | 80 | 80 | IG: 56.89±2.64 CG: 57.14±3.01 | pharmacist intervention on the basis of conventional treatment | Routine treatment includes drug treatment, dietary guidance, regular follow-up, etc. | 3 months | Beijing |
| U034 | 215 | 2024 | Mai | MTM | Multi-Disease | 41 | 41 | IG: 72.83±5.12 CG: 72.54±4.68 | MTM model pharmacy outpatient service | Routine drug consultation and medication guidance | 3 months | Guangdong |
| U036 | 216 | 2024 | Tang | PI | Multi-Disease | 48 | 48 | IG: 57.7±11.71 CG: 55.42±13.66 | pharmacist interventions based on the IMB model | Traditional pharmacist intervention methods | 12 months | Jiangxi |
| U037 | 217 | 2025 | Yang | MR | Multi-Disease | 50 | 50 | IG: 57.8±10.56 CG: 58.88±9.35 | In addition to conventional treatment, receive drug reformulation services provided by clinical pharmacists | Routine treatment (no pharmacist services) | 6 months | Anhui |
| S001 | 218 | 2024 | Jiang | MTM | Coronary Disease | 61 | 59 | IG: 67.26±9.89 CG: 66.73±8.92 | Add standardized MTM services based on the control group | Routine diagnosis and treatment services | 6 months | Hebei |
| S002 | 219 | 2019 | Xiao | PI | Coronary Disease | 52 | 52 | IG: 80.1±5.58 CG: 79.44±6.08 | On the basis of the treatment in the control group, pharmacist intervention was also given, including telephone follow-up after discharge. | Normal clinical cardiac rehabilitation treatment plan, including comprehensive treatment such as exercise rehabilitation training, conventional drug treatment, health education, dietary intervention and psychological counseling | 3 months | Zhejiang |
| S003 | 220 | 2021 | Yuan | PI | Coronary Disease | 35 | 36 | IG: 68.2±8.49 CG: 70.3±13.5 | On the basis of the control group, the intervention group was provided with pharmacist intervention by clinical pharmacists at discharge and after discharge. | The control group was provided with routine pharmacist services during hospitalization. | 6 months | Jiangsu |
| S027 | 221 | 2008 | Li | PI | Diabetes Mellitus | 30 | 30 |  | Regularly provide pharmacist services through phone calls, visits, lectures, etc. | Routine diagnosis and treatment | 6 months | Jiangxi |
| S010 | 222 | 2012 | Luo | PI | Diabetes Mellitus | 30 | 30 | IG: 59.1±9.8 CG: 59.3±10.4 | Clinical pharmacists provide individualized guidance to patients in the intervention group | Routine diagnosis and treatment | 3 months | Jiangxi |
| S026 | 223 | 2012 | Xue | PI | Diabetes Mellitus | 80 | 80 |  | Carry out safe medication and health education interventions during the consultation process | No intervention measures were taken, only questionnaire survey | 3 months | Hubei |
| S008 | 224 | 2013 | Ji | PI | Diabetes Mellitus | 32 | 31 | IG: 43.66±13.03 CG: 41.81±17.38 | In addition to receiving routine medical services, receive full pharmacist services from pharmacists | Routine medical services | 6 months | Jiangsu |
| S028 | 225 | 2013 | Chen | PI | Diabetes Mellitus | 98 | 97 |  | The clinical pharmacist will establish a special electronic medication record for him and receive full-process, focused individual monitoring and intervention treatment from the intervention team (including clinical pharmacists, endocrinologists, and nurses). | conventional treatment |  | Sichuan |
| S012 | 226 | 2015 | Li | PI | Diabetes Mellitus | 18 | 17 | IG: 50.5±13.6 CG: 47.2±12.1 | Receive routine treatment and regular follow-up and intervention from pharmacists | receive conventional treatment | 3 months | Beijing |
| S009 | 227 | 2016 | Gong | PI | Diabetes Mellitus | 62 | 62 | IG: 57.3±8.7 CG: 55.7±10.7 | On the basis of the treatment in the control group, the group received professional guidance from clinical pharmacists in the Department of Endocrinology. At least one family member of the patient was given the same education, and the family members were required to remind or supervise the patient's medication and lifestyle management. | The control group received general medication education, diet control, exercise guidance, etc. from doctors or nurses. | 3 months | Zhejiang |
| S015 | 228 | 2016 | Yang | PI | Diabetes Mellitus | 60 | 60 |  | Full-service pharmacist service approach | Guidance on routine medication methods | 6 months | Shandong |
| S022 | 229 | 2016 | Shen | PC | Diabetes Mellitus | 73 | 75 |  | In addition to receiving medical services, pharmacists will provide continuous pharmacist services according to the pharmacist intervention path. | Only receive routine medical services, no pharmacist service intervention | 12 months | Shanghai |
| S004 | 230 | 2017 | Zheng | PC | Diabetes Mellitus | 150 | 150 | IG: 65.7±3.2 CG: 66.2±3.1 | Provide pharmacist services to patients using case management model based on OS | conventional treatment | 6 months | Guangdong |
| S013 | 231 | 2017 | Su | PI | Diabetes Mellitus | 47 | 45 | IG: 49.2±4.43 CG: 48.2±3.72 | The intervention group also received clinical pharmacist services provided by clinical pharmacists. clinical pharmacist services focused on review of medical orders, medication education and medication monitoring of patients. | routine intervention | 6 months | Guangdong |
| S023 | 232 | 2018 | Ran | PI | Diabetes Mellitus | 100 | 100 | IG: 48.72±10.57 CG: 48.46±10.83 | Combined use of pharmacist intervention on the basis of the control group | clinical basic plan |  | Hubei |
| S014 | 233 | 2019 | Li | PI | Diabetes Mellitus | 27 | 29 | IG: 52±12.4 CG: 56.3±8.4 | Routine treatment and chronic disease management by pharmacists. The chronic disease management method is telephone follow-up, and patients are encouraged to use WeChat to contact pharmacists. | conventional treatment | 6 months | Beijing |
| S016 | 234 | 2020 | Ji | PI | Diabetes Mellitus | 90 | 90 | IG: 52.67±12.39 CG: 53.2±17 | The experimental group received pharmacist services from licensed pharmacists | The control group did not receive services from licensed pharmacists | 6 months | Jiangsu |
| S021 | 235 | 2019 | Wang | PI | Diabetes Mellitus | 40 | 39 | IG: 61.53±13.26 CG: 64±12.6 | Receive standardized pharmacist services related to anti-diabetic drugs | General lifestyle education |  | Shanghai |
| S019 | 236 | 2020 | Nie | MR | Diabetes Mellitus | 181 | 176 | IG: 72.2±6.2 CG: 72.7±5.9 | In addition to conventional treatment, patients also receive MR services provided by clinical pharmacists | conventional treatment | 6 months | Shaanxi |
| S007 | 237 | 2021 | Liu | MTM | Diabetes Mellitus | 64 | 64 | IG: 60.5±10.5 CG: 62.2±9.8 | Regional collaborative diabetes medication management service model | Conventional Diabetes Management Model | 6 months | Shanghai |
| S025 | 238 | 2021 | Xuan | PI | Diabetes Mellitus | 100 | 99 | IG: 58.86±10.59 CG: 59.2±10.34 | In addition to conventional treatment, patients also received diabetes medication education and information-based follow-up from clinical pharmacists. | Routine treatment by medical staff | 6 months | Jiangsu |
| S005 | 239 | 2022 | Chu | PC | Diabetes Mellitus | 35 | 35 | IG: 52.1±5.2 CG: 53.3±4.9 | Pharmacist-physician joint service model intervention, clinical pharmacists use the PCNE classification method to comprehensively evaluate the patient's medication use and the types and causes of DRPs, and work with general practitioners to formulate targeted intervention measures and carry out individualized pharmacist services. | Traditional community medical and nursing joint service model intervention | 12 months | Anhui |
| S011 | 240 | 2022 | Ji | PI | Diabetes Mellitus | 107 | 118 | IG: 53.61±6.62 CG: 53.78±4.55 | Managed by the "Comprehensive Diabetes Clinic" team | Receive routine care in the endocrinology clinic | 6 months | Jiangsu |
| S006 | 241 | 2023 | Ke | PC | Diabetes Mellitus | 83 | 83 | IG: 57.86±14.5 CG: 61.57±11.82 | Pharmacist-involved pharmaceutical care | General Medical Diagnosis and Treatment |  | Hubei |
| S018 | 242 | 2025 | Zhan | PI | Diabetes Mellitus | 80 | 80 | IG: 52.53±3.38 CG: 52.32±3.34 | Multidisciplinary chronic disease management by clinical pharmacists combined with medical care | General management | 6 months | Guangxi |
| S020 | 243 | 2025 | Ge | MR | Diabetes Mellitus | 155 | 141 | IG: 71.9±5 CG: 72±5.1 | In the control group, clinical pharmacists carried out medication reformulation and applied the teach-back method in medication education. | Conventional medical model treatment | 6 months | Jiangsu |
| S024 | 244 | 2025 | Ding | PI | Diabetes Mellitus | 240 | 100 |  | The subjects in the intervention group received a one-year pharmaceutical health education intervention on the basis of routine pharmacist services. | Routine pharmacist services include pharmacist prescription review, routine medication instructions and drug use reminders when picking up medicine at the window, and medication consultation and guidance at the medication consultation window. | 12 months | Shanghai |
| S029 | 245 | 2022 | Bai | PI | Dyslipidemia | 276 | 243 | IG: 64.36±9.67 CG: 65.59±11.31 | Implement complete chronic disease management, that is, follow the established pharmacological chronic disease management worksheet for dyslipidemia to carry out patient data collection, medication guidance, medication education (including basic knowledge about disease science, monitoring of drug efficacy indicators, drug interactions, self-management of adverse drug reactions), and lifestyle management. | Implement routine medication guidance, that is, clinical pharmacists provide necessary medication guidance and patient medication consultation, and the clinical data are obtained from the hospital's electronic medical record system |  | Guizhou |
| S030 | 246 | 2014 | Yu | PC | Dyslipidemia | 55 | 47 | IG: 67.32±16.78 CG: 65.94±15.54 | Interventions are carried out according to established pharmaceutical care plans | No intervention | 12 months | Jiangsu |
| S031 | 247 | 2022 | Zhang | MTM | Heart Failure | 41 | 36 | IG: 61.36±14.83 CG: 64.21±11.3 | Referring to the American MTM service model, with specialist clinical pharmacists as the mainstay and multidisciplinary pharmacists participating together, a professional pharmaceutical treatment service model is developed for CHF patients. | Only general medication consultations are accepted | 6 months | Jiangsu |
| S035 | 248 | 2013 | Guo | PI | Hypertension | 71 | 71 | IG: 57.25±3.28 CG: 56.77±3.57 | Clinical pharmacist intervention in medication administration | Adopt conventional antihypertensive drug treatment without clinical pharmacist intervention |  | Fujian |
| S037 | 249 | 2013 | Li | PI | Hypertension | 60 | 60 |  | A team of clinical pharmacists regularly participate in return visits to provide patients with medication guidance, life and health education, and adjustment of treatment drug plans. | Go directly to the outpatient clinic regularly for routine treatment by a doctor | 6 months | Sichuan |
| S032 | 250 | 2015 | Li | PC | Hypertension | 60 | 60 | IG: 46.34±4.34 CG: 45.87±4.57 | Combined with pharmacist services on the basis of conventional treatment, that is, clinical pharmacists participate in services for patients in drug selection, medication adherence, medication timing, combined drug treatment, prevention of adverse reactions, and lifestyle intervention, and provide corresponding treatment to those with combined diseases. | conventional treatment | 3 months | Henan |
| S042 | 251 | 2015 | Zhang | PI | Hypertension | 44 | 44 | IG: 68.5±2.7 CG: 69.8±2.5 | Strengthen the following pharmacist service intervention measures based on the control group | Routine treatment and health education |  | Shaanxi |
| S033 | 252 | 2017 | Liang | PI | Hypertension | 26 | 23 | IG: 52.8±10.5 CG: 49.7±10.4 | The pharmacist intervention group's intervention measures for patients include education for discharged patients and WeChat follow-up for discharged patients. | No pharmacist intervention | 3 months | Beijing |
| S036 | 253 | 2018 | Jiao | PI | Hypertension | 30 | 30 | IG: 63.21±6.19 CG: 62.96±6.42 | clinical pharmacist intervention | No clinical pharmacist intervention will be implemented | 12 months | Shanghai |
| S039 | 254 | 2019 | Bai | PI | Hypertension | 41 | 39 | IG: 67±5 CG: 68±5 | Provide 8 times of health education, inpatient education or telephone follow-up focusing on lifestyle and medication adherence | Only 2 health education sessions will be given | 3 months | Beijing |
| S034 | 255 | 2021 | Zhou | PI | Hypertension | 68 | 68 |  | Patients in the intervention group received enhanced pharmacist service intervention measures on the basis of the control group. | Routine hypertension nursing intervention and medication treatment as prescribed by the doctor | 6 months | Henan |
| S041 | 256 | 2021 | Shi | PI | Hypertension | 25 | 25 | IG: 62.14±12.6 CG: 63.01±14.4 | On the basis of family doctor services, pharmacists participate in and implement a 12-month personalized pharmacist service intervention. | No pharmacists participate in family doctor services | 12 months | Shanghai |
| S040 | 257 | 2023 | Liu | PI | Hypertension | 200 | 200 | IG: 67.1±8.8 CG: 65.5±9.2 | Receive a 12-month intensive pharmacist intervention involving multiple indicators, including drugs and lifestyle, provided by clinical pharmacists. | Pharmacists will only record various indicators and evaluate medication adherence during enrollment and follow-up (the frequency is the same as that of the intervention group), and will not receive any education or intervention. | 12 months | Henan |
| S043 | 258 | 2024 | Lu | PC | Hypertension | 167 | 170 | IG: 68.6±8.5 CG: 68.2±9.3 | Increase remote pharmacy services, specifically for pharmacists to issue medication instructions to patients for medication guidance, which is convenient for patients to review outside the hospital; during the follow-up period, clinical pharmacists conduct regular follow-up visits with patients via WeChat and phone calls (once a week) | Routine medical services | 6 months | Shanghai |
| S038 | 259 | 2025 | Li | PI | Hypertension | 40 | 40 | IG: 52±8 CG: 51±9 | On the basis of routine management, clinical pharmacists will conduct standardized management in accordance with the "Multidisciplinary Expert Consensus on Clinical Screening for Secondary Hypertension in China (2023)" | A routine hypertension management process is implemented, and cardiologists routinely perform diagnosis, treatment plan formulation and follow-up when patients first visit. Patients are instructed to measure and record their blood pressure on time every day, follow up with outpatients every four weeks, adjust the antihypertensive plan based on blood pressure conditions and office blood pressure, and provide patients with standardized education on basic hypertension knowledge (salt-restricted diet, regular medication, etc.) without active intervention by pharmacists. | 3 months | Gansu |
| S049 | 260 | 2017 | Guo | PI | Multi-Disease | 102 | 102 | IG: 33.6±8.1 CG: 32.2±7.8 | Traditional medical treatment + medication monitoring | Traditional medical treatment (conventional treatment + diet control + exercise therapy) | 6 months | Guangdong |
| S044 | 261 | 2020 | Yan | PI | Multi-Disease | 191 | 187 |  | The team records the patient's general condition on site, discusses prescription optimization with the doctor and decides whether to implement it, and provides medication guidance to the patient. | Routine diagnosis and treatment services | 12 months | Jiangsu |
| S046 | 262 | 2020 | Wang | PI | Multi-Disease | 42 | 42 | IG: 36.76±1.322 CG: 36.69±1.22 | Pharmacist + nutritionist joint clinic to provide personalized guidance on pharmaceutical combined nutrition | No individualized guidance on medicine and nutrition |  | Shandong |
| S051 | 263 | 2022 | Xu | PI | Multi-Disease | 60 | 60 | IG: 60.92±7.03 CG: 60.85±6.98 | pharmacist service intervention under the integrated model | Basic pharmacist service intervention |  | Shaanxi |
| S050 | 264 | 2023 | Zhang | MTM | Multi-Disease | 40 | 40 |  | Based on the control group, the MTM model was adopted | Traditional medical service model |  | Hebei |
| S047 | 265 | 2024 | Zhang | PC | Multi-Disease | 60 | 60 | IG: 57.83±10.44 CG: 58.97±9.65 | Pharmacy monitoring through PCNE system | Take medication according to doctor’s instructions without pharmaceutical supervision |  | Shanghai |
| S045 | 266 | 2025 | Bai | PI | Multi-Disease | 40 | 40 | IG: 74.72±12.77 CG: 75.31±11.28 | In addition to checking patient information and drug information according to routine nursing measures, and providing drug guidance to patients, we also analyze the drugs and factors involved in unreasonable medical orders, implement medical order intervention, and ensure the rational application of medical orders. | Routine nursing measures |  | Beijing |

Supplementary Table S8B | Care setting and hospital grade of the 266 included trials. Hospital grade reflects the official grade of the institution in the trial’s publication year. Trial-level institution assignments are provided in the shared dataset.

| **Care setting** | **Trials** | **% of 266** |
| --- | --- | --- |
| Tertiary hospital | 149 | 56.0% |
| Secondary hospital | 75 | 28.2% |
| Primary-level hospital | 7 | 2.6% |
| Primary care or community health institution | 35 | 13.2% |

Supplementary Table S9 | Trial-level intervention-activity reporting

| **Trial ID** | **Author-year** | **Medication reconciliation** | **Medication review** | **Education** | **Adherence support** | **ADR/ADE monitoring** | **Prescribing input** | **Post-discharge follow-up** | **Multidisciplinary** | **Number explicitly reported** |
| --- | --- | --- | --- | --- | --- | --- | --- | --- | --- | --- |
| O001 | Huang 2015 | Not reported | Not reported | Explicitly reported | Not reported | Not reported | Explicitly reported | Explicitly reported | Explicitly reported | 4 |
| O002 | Zhou 2021 | Not reported | Explicitly reported | Explicitly reported | Not reported | Not reported | Not reported | Not reported | Not reported | 2 |
| O003 | Cheng 2022 | Not reported | Not reported | Explicitly reported | Explicitly reported | Not reported | Not reported | Explicitly reported | Explicitly reported | 4 |
| O004 | Chen 2016 | Not reported | Not reported | Not reported | Not reported | Not reported | Not reported | Explicitly reported | Not reported | 1 |
| O005 | Pan 2017 | Not reported | Explicitly reported | Explicitly reported | Explicitly reported | Not reported | Not reported | Not reported | Not reported | 3 |
| O006 | Chen 2019 | Not reported | Not reported | Explicitly reported | Explicitly reported | Not reported | Not reported | Explicitly reported | Not reported | 3 |
| O007 | Chen 2020 | Not reported | Not reported | Not reported | Explicitly reported | Not reported | Not reported | Explicitly reported | Explicitly reported | 3 |
| O008 | Rao 2020 | Not reported | Explicitly reported | Explicitly reported | Not reported | Not reported | Not reported | Explicitly reported | Explicitly reported | 4 |
| O009 | Liu 2022 | Not reported | Not reported | Explicitly reported | Not reported | Not reported | Not reported | Not reported | Not reported | 1 |
| O010 | Wang 2022 | Not reported | Not reported | Explicitly reported | Not reported | Not reported | Not reported | Not reported | Explicitly reported | 2 |
| O011 | Tang 2007 | Not reported | Not reported | Explicitly reported | Not reported | Not reported | Not reported | Not reported | Not reported | 1 |
| O012 | Zhong 2010 | Not reported | Not reported | Explicitly reported | Explicitly reported | Not reported | Not reported | Not reported | Not reported | 2 |
| O013 | Zhao 2012 | Not reported | Not reported | Explicitly reported | Explicitly reported | Not reported | Explicitly reported | Not reported | Explicitly reported | 4 |
| O015 | Gan 2016 | Not reported | Not reported | Explicitly reported | Explicitly reported | Not reported | Not reported | Not reported | Explicitly reported | 3 |
| O016 | Xu 2019 | Not reported | Explicitly reported | Not reported | Not reported | Not reported | Explicitly reported | Not reported | Explicitly reported | 3 |
| O017 | Liu 2022 | Not reported | Explicitly reported | Explicitly reported | Not reported | Not reported | Not reported | Not reported | Explicitly reported | 3 |
| O018 | Wong 2013 | Not reported | Not reported | Explicitly reported | Explicitly reported | Not reported | Not reported | Not reported | Not reported | 2 |
| O019 | Qin 2013 | Not reported | Not reported | Explicitly reported | Not reported | Not reported | Explicitly reported | Not reported | Explicitly reported | 3 |
| O020 | Qin 2010 | Not reported | Not reported | Explicitly reported | Explicitly reported | Not reported | Not reported | Not reported | Explicitly reported | 3 |
| O021 | Li 2011 | Not reported | Explicitly reported | Explicitly reported | Not reported | Explicitly reported | Not reported | Explicitly reported | Not reported | 4 |
| O022 | Zheng 2020 | Not reported | Not reported | Explicitly reported | Not reported | Not reported | Explicitly reported | Not reported | Explicitly reported | 3 |
| O023 | Wan 2015 | Not reported | Not reported | Explicitly reported | Not reported | Not reported | Not reported | Not reported | Not reported | 1 |
| O024 | Zhu 2018 | Not reported | Not reported | Explicitly reported | Explicitly reported | Not reported | Not reported | Not reported | Not reported | 2 |
| O025 | Xu 2019 | Not reported | Not reported | Explicitly reported | Not reported | Not reported | Not reported | Not reported | Not reported | 1 |
| O026 | Wu 2019 | Not reported | Not reported | Explicitly reported | Not reported | Not reported | Not reported | Explicitly reported | Not reported | 2 |
| O027 | Wu 2021 | Not reported | Explicitly reported | Explicitly reported | Not reported | Not reported | Not reported | Explicitly reported | Not reported | 3 |
| O028 | Su 2021 | Not reported | Not reported | Explicitly reported | Explicitly reported | Not reported | Explicitly reported | Not reported | Not reported | 3 |
| O029 | Jiang 2022 | Not reported | Explicitly reported | Not reported | Explicitly reported | Not reported | Explicitly reported | Not reported | Explicitly reported | 4 |
| O030 | Li 2022 | Not reported | Not reported | Explicitly reported | Not reported | Not reported | Explicitly reported | Not reported | Not reported | 2 |
| O031 | Jin 2022 | Not reported | Not reported | Explicitly reported | Not reported | Not reported | Explicitly reported | Explicitly reported | Not reported | 3 |
| O032 | Gong 2022 | Not reported | Explicitly reported | Not reported | Explicitly reported | Not reported | Not reported | Not reported | Not reported | 2 |
| O033 | Jiang 2021 | Not reported | Not reported | Explicitly reported | Not reported | Not reported | Not reported | Explicitly reported | Explicitly reported | 3 |
| O034 | Fan 2021 | Not reported | Not reported | Explicitly reported | Explicitly reported | Not reported | Explicitly reported | Not reported | Explicitly reported | 4 |
| O035 | Yue 2020 | Explicitly reported | Not reported | Explicitly reported | Not reported | Not reported | Not reported | Not reported | Not reported | 2 |
| O036 | Wang 2020 | Not reported | Not reported | Explicitly reported | Not reported | Not reported | Explicitly reported | Not reported | Explicitly reported | 3 |
| O037 | Wang 2020 | Explicitly reported | Explicitly reported | Explicitly reported | Explicitly reported | Explicitly reported | Explicitly reported | Not reported | Explicitly reported | 7 |
| O038 | He 2020 | Not reported | Not reported | Explicitly reported | Not reported | Not reported | Not reported | Not reported | Not reported | 1 |
| O039 | Dai 2020 | Not reported | Not reported | Explicitly reported | Not reported | Not reported | Not reported | Not reported | Explicitly reported | 2 |
| O040 | Wang 2019 | Not reported | Not reported | Explicitly reported | Not reported | Not reported | Not reported | Not reported | Not reported | 1 |
| O041 | Yang 2018 | Not reported | Explicitly reported | Explicitly reported | Explicitly reported | Not reported | Not reported | Not reported | Not reported | 3 |
| O042 | Wang 2017 | Not reported | Not reported | Explicitly reported | Not reported | Not reported | Not reported | Not reported | Not reported | 1 |
| O043 | Jiang 2017 | Not reported | Explicitly reported | Explicitly reported | Not reported | Not reported | Not reported | Explicitly reported | Not reported | 3 |
| O044 | Cao 2017 | Not reported | Not reported | Explicitly reported | Explicitly reported | Not reported | Not reported | Not reported | Not reported | 2 |
| O045 | Hao 2014 | Not reported | Not reported | Not reported | Explicitly reported | Not reported | Not reported | Not reported | Not reported | 1 |
| O046 | Han 2012 | Not reported | Not reported | Explicitly reported | Explicitly reported | Not reported | Not reported | Not reported | Not reported | 2 |
| O047 | Wei 2022 | Not reported | Explicitly reported | Explicitly reported | Explicitly reported | Explicitly reported | Not reported | Not reported | Explicitly reported | 5 |
| O048 | Zhuo 2022 | Not reported | Not reported | Explicitly reported | Not reported | Not reported | Explicitly reported | Not reported | Explicitly reported | 3 |
| O049 | Wang 2022 | Not reported | Not reported | Explicitly reported | Explicitly reported | Not reported | Not reported | Explicitly reported | Not reported | 3 |
| O050 | Zhou 2022 | Not reported | Not reported | Explicitly reported | Explicitly reported | Not reported | Not reported | Not reported | Not reported | 2 |
| O051 | Pan 2022 | Not reported | Not reported | Not reported | Explicitly reported | Not reported | Not reported | Not reported | Not reported | 1 |
| O053 | Shen 2021 | Not reported | Not reported | Explicitly reported | Not reported | Not reported | Explicitly reported | Explicitly reported | Explicitly reported | 4 |
| O054 | Lin 2013 | Not reported | Not reported | Not reported | Not reported | Not reported | Not reported | Not reported | Not reported | 0 |
| O055 | Li 2015 | Not reported | Not reported | Explicitly reported | Explicitly reported | Explicitly reported | Not reported | Explicitly reported | Explicitly reported | 5 |
| O056 | Zhuo 2015 | Not reported | Not reported | Explicitly reported | Not reported | Not reported | Explicitly reported | Not reported | Explicitly reported | 3 |
| O057 | Zhou 2016 | Not reported | Not reported | Explicitly reported | Explicitly reported | Not reported | Not reported | Not reported | Not reported | 2 |
| O058 | Yin 2016 | Not reported | Not reported | Explicitly reported | Not reported | Not reported | Not reported | Not reported | Explicitly reported | 2 |
| O059 | He 2016 | Not reported | Explicitly reported | Explicitly reported | Explicitly reported | Explicitly reported | Not reported | Not reported | Explicitly reported | 5 |
| O060 | Zhang 2016 | Not reported | Not reported | Explicitly reported | Explicitly reported | Not reported | Not reported | Not reported | Not reported | 2 |
| O061 | Peng 2017 | Not reported | Not reported | Explicitly reported | Not reported | Not reported | Not reported | Not reported | Explicitly reported | 2 |
| O062 | Zhou 2017 | Not reported | Not reported | Explicitly reported | Explicitly reported | Not reported | Not reported | Not reported | Not reported | 2 |
| O063 | Ye 2017 | Not reported | Not reported | Explicitly reported | Explicitly reported | Not reported | Not reported | Not reported | Explicitly reported | 3 |
| O064 | Xu 2017 | Not reported | Not reported | Explicitly reported | Explicitly reported | Not reported | Explicitly reported | Not reported | Not reported | 3 |
| O065 | Hu 2017 | Not reported | Not reported | Explicitly reported | Explicitly reported | Not reported | Not reported | Not reported | Explicitly reported | 3 |
| O066 | Ren 2018 | Not reported | Explicitly reported | Explicitly reported | Explicitly reported | Not reported | Not reported | Not reported | Not reported | 3 |
| O067 | Li 2018 | Not reported | Explicitly reported | Explicitly reported | Explicitly reported | Not reported | Explicitly reported | Not reported | Explicitly reported | 5 |
| O068 | Lei 2018 | Not reported | Not reported | Explicitly reported | Not reported | Not reported | Explicitly reported | Not reported | Explicitly reported | 3 |
| O069 | Zhang 2018 | Not reported | Explicitly reported | Explicitly reported | Not reported | Not reported | Not reported | Not reported | Not reported | 2 |
| O070 | Lyu 2018 | Not reported | Not reported | Explicitly reported | Not reported | Not reported | Not reported | Not reported | Explicitly reported | 2 |
| O071 | Duan 2018 | Not reported | Not reported | Explicitly reported | Explicitly reported | Not reported | Not reported | Not reported | Not reported | 2 |
| O073 | Wu 2019 | Not reported | Not reported | Explicitly reported | Explicitly reported | Not reported | Not reported | Not reported | Not reported | 2 |
| O074 | Hu 2019 | Not reported | Not reported | Not reported | Not reported | Not reported | Not reported | Not reported | Not reported | 0 |
| O075 | Feng 2019 | Not reported | Not reported | Explicitly reported | Explicitly reported | Not reported | Not reported | Not reported | Explicitly reported | 3 |
| O076 | Zhao 2019 | Not reported | Not reported | Explicitly reported | Explicitly reported | Not reported | Not reported | Explicitly reported | Explicitly reported | 4 |
| O077 | Zhang 2019 | Not reported | Not reported | Explicitly reported | Explicitly reported | Not reported | Not reported | Not reported | Not reported | 2 |
| O078 | Wang 2019 | Not reported | Not reported | Explicitly reported | Explicitly reported | Explicitly reported | Not reported | Not reported | Explicitly reported | 4 |
| O079 | Peng 2019 | Explicitly reported | Not reported | Explicitly reported | Explicitly reported | Not reported | Not reported | Explicitly reported | Not reported | 4 |
| O080 | He 2019 | Not reported | Not reported | Explicitly reported | Not reported | Not reported | Explicitly reported | Not reported | Not reported | 2 |
| O082 | Yang 2020 | Not reported | Not reported | Explicitly reported | Not reported | Not reported | Not reported | Not reported | Not reported | 1 |
| O083 | Wu 2020 | Not reported | Not reported | Not reported | Not reported | Not reported | Not reported | Not reported | Not reported | 0 |
| O084 | Sun 2020 | Not reported | Explicitly reported | Explicitly reported | Explicitly reported | Not reported | Not reported | Not reported | Explicitly reported | 4 |
| O085 | Liu 2020 | Not reported | Not reported | Explicitly reported | Explicitly reported | Not reported | Not reported | Not reported | Explicitly reported | 3 |
| O086 | Hu 2020 | Not reported | Explicitly reported | Explicitly reported | Explicitly reported | Not reported | Explicitly reported | Not reported | Explicitly reported | 5 |
| O087 | Chen 2020 | Not reported | Not reported | Not reported | Explicitly reported | Not reported | Not reported | Not reported | Not reported | 1 |
| O088 | Cai 2020 | Not reported | Not reported | Not reported | Not reported | Not reported | Not reported | Not reported | Not reported | 0 |
| O089 | Li 2021 | Not reported | Explicitly reported | Explicitly reported | Not reported | Not reported | Not reported | Not reported | Not reported | 2 |
| O090 | Cai 2021 | Not reported | Not reported | Explicitly reported | Not reported | Not reported | Not reported | Not reported | Explicitly reported | 2 |
| O091 | Mei 2021 | Not reported | Explicitly reported | Explicitly reported | Not reported | Not reported | Not reported | Not reported | Not reported | 2 |
| O092 | Liang 2021 | Not reported | Not reported | Not reported | Not reported | Not reported | Not reported | Not reported | Not reported | 0 |
| O093 | Chen 2021 | Not reported | Not reported | Not reported | Explicitly reported | Not reported | Not reported | Not reported | Not reported | 1 |
| O094 | Yang 2022 | Not reported | Explicitly reported | Explicitly reported | Explicitly reported | Not reported | Not reported | Not reported | Explicitly reported | 4 |
| O095 | Ma 2022 | Not reported | Not reported | Explicitly reported | Not reported | Not reported | Not reported | Explicitly reported | Explicitly reported | 3 |
| O096 | Guo 2022 | Not reported | Not reported | Explicitly reported | Not reported | Not reported | Not reported | Not reported | Not reported | 1 |
| O097 | Dong 2022 | Not reported | Not reported | Explicitly reported | Explicitly reported | Not reported | Not reported | Not reported | Not reported | 2 |
| O098 | Chai 2022 | Not reported | Not reported | Explicitly reported | Explicitly reported | Not reported | Not reported | Not reported | Explicitly reported | 3 |
| O099 | Zhou 2021 | Not reported | Not reported | Explicitly reported | Not reported | Not reported | Not reported | Not reported | Not reported | 1 |
| O100 | Li 2013 | Not reported | Not reported | Explicitly reported | Not reported | Not reported | Not reported | Not reported | Not reported | 1 |
| O101 | Zhao 2013 | Not reported | Not reported | Explicitly reported | Not reported | Not reported | Explicitly reported | Not reported | Not reported | 2 |
| O102 | Yang 2013 | Not reported | Not reported | Explicitly reported | Explicitly reported | Explicitly reported | Not reported | Not reported | Not reported | 3 |
| O103 | Yao 2014 | Not reported | Not reported | Explicitly reported | Explicitly reported | Not reported | Not reported | Not reported | Not reported | 2 |
| O104 | Wu 2014 | Not reported | Not reported | Explicitly reported | Explicitly reported | Not reported | Not reported | Not reported | Not reported | 2 |
| O105 | Wen 2014 | Not reported | Not reported | Not reported | Not reported | Not reported | Not reported | Not reported | Not reported | 0 |
| O106 | Tan 2014 | Not reported | Not reported | Not reported | Not reported | Not reported | Not reported | Not reported | Explicitly reported | 1 |
| O107 | Shen 2014 | Not reported | Not reported | Explicitly reported | Explicitly reported | Not reported | Not reported | Explicitly reported | Explicitly reported | 4 |
| O108 | Pei 2014 | Not reported | Not reported | Not reported | Explicitly reported | Not reported | Not reported | Explicitly reported | Not reported | 2 |
| O109 | Pei 2014 | Not reported | Not reported | Explicitly reported | Explicitly reported | Not reported | Not reported | Not reported | Explicitly reported | 3 |
| O110 | Jin 2014 | Not reported | Not reported | Not reported | Not reported | Not reported | Not reported | Not reported | Not reported | 0 |
| O111 | Zhou 2015 | Not reported | Not reported | Not reported | Not reported | Not reported | Not reported | Not reported | Not reported | 0 |
| O112 | Zhao 2015 | Not reported | Not reported | Not reported | Not reported | Not reported | Not reported | Not reported | Not reported | 0 |
| O113 | Yang 2015 | Not reported | Not reported | Not reported | Explicitly reported | Not reported | Not reported | Not reported | Not reported | 1 |
| O114 | Luo 2015 | Not reported | Not reported | Explicitly reported | Not reported | Not reported | Not reported | Not reported | Not reported | 1 |
| O115 | Liao 2015 | Not reported | Not reported | Explicitly reported | Explicitly reported | Not reported | Not reported | Not reported | Not reported | 2 |
| O116 | Li 2015 | Not reported | Not reported | Explicitly reported | Explicitly reported | Not reported | Not reported | Not reported | Not reported | 2 |
| O117 | Feng 2015 | Not reported | Not reported | Not reported | Explicitly reported | Not reported | Not reported | Not reported | Not reported | 1 |
| O118 | Chen 2015 | Not reported | Not reported | Explicitly reported | Not reported | Not reported | Not reported | Not reported | Not reported | 1 |
| O119 | Yu 2016 | Not reported | Not reported | Explicitly reported | Explicitly reported | Not reported | Not reported | Not reported | Not reported | 2 |
| O120 | Wang 2016 | Not reported | Not reported | Explicitly reported | Not reported | Not reported | Not reported | Not reported | Not reported | 1 |
| O121 | Sun 2016 | Not reported | Not reported | Not reported | Not reported | Not reported | Not reported | Not reported | Not reported | 0 |
| O122 | Dong 2016 | Not reported | Not reported | Explicitly reported | Not reported | Not reported | Not reported | Not reported | Not reported | 1 |
| O123 | Zeng 2016 | Not reported | Not reported | Explicitly reported | Not reported | Not reported | Not reported | Not reported | Not reported | 1 |
| O124 | Wen 2017 | Not reported | Not reported | Explicitly reported | Explicitly reported | Not reported | Not reported | Not reported | Explicitly reported | 3 |
| O125 | Wang 2017 | Not reported | Not reported | Explicitly reported | Explicitly reported | Not reported | Not reported | Not reported | Explicitly reported | 3 |
| O126 | Pan 2017 | Not reported | Not reported | Explicitly reported | Explicitly reported | Explicitly reported | Not reported | Not reported | Not reported | 3 |
| O127 | Han 2017 | Not reported | Not reported | Explicitly reported | Not reported | Not reported | Not reported | Not reported | Not reported | 1 |
| O128 | Ni 2018 | Not reported | Not reported | Explicitly reported | Explicitly reported | Not reported | Not reported | Not reported | Not reported | 2 |
| O129 | Tan 2018 | Not reported | Not reported | Explicitly reported | Explicitly reported | Not reported | Not reported | Not reported | Not reported | 2 |
| O130 | Lyu 2018 | Not reported | Not reported | Explicitly reported | Not reported | Not reported | Not reported | Not reported | Not reported | 1 |
| O131 | Li 2018 | Not reported | Not reported | Not reported | Not reported | Not reported | Not reported | Not reported | Not reported | 0 |
| O132 | Huang 2018 | Not reported | Not reported | Explicitly reported | Not reported | Not reported | Not reported | Not reported | Not reported | 1 |
| O133 | Hou 2018 | Not reported | Not reported | Explicitly reported | Not reported | Not reported | Not reported | Not reported | Not reported | 1 |
| O134 | Guo 2018 | Not reported | Explicitly reported | Explicitly reported | Explicitly reported | Not reported | Not reported | Explicitly reported | Explicitly reported | 5 |
| O135 | Fang 2018 | Not reported | Not reported | Explicitly reported | Explicitly reported | Not reported | Not reported | Not reported | Not reported | 2 |
| O136 | Du 2018 | Not reported | Not reported | Not reported | Not reported | Not reported | Not reported | Not reported | Not reported | 0 |
| O137 | Chen 2018 | Not reported | Not reported | Explicitly reported | Not reported | Not reported | Not reported | Not reported | Not reported | 1 |
| O138 | Yao 2019 | Not reported | Not reported | Not reported | Not reported | Not reported | Not reported | Not reported | Not reported | 0 |
| O139 | Wang 2019 | Not reported | Not reported | Explicitly reported | Explicitly reported | Not reported | Not reported | Explicitly reported | Not reported | 3 |
| O140 | Mi 2019 | Not reported | Not reported | Explicitly reported | Not reported | Not reported | Not reported | Not reported | Not reported | 1 |
| O141 | Liu 2019 | Explicitly reported | Explicitly reported | Explicitly reported | Not reported | Not reported | Not reported | Explicitly reported | Not reported | 4 |
| O142 | Liang 2019 | Not reported | Not reported | Explicitly reported | Explicitly reported | Not reported | Explicitly reported | Not reported | Not reported | 3 |
| O143 | Li 2019 | Not reported | Not reported | Not reported | Not reported | Not reported | Not reported | Not reported | Not reported | 0 |
| O144 | Du 2019 | Not reported | Not reported | Explicitly reported | Not reported | Not reported | Not reported | Not reported | Not reported | 1 |
| O145 | Chen 2019 | Not reported | Not reported | Explicitly reported | Not reported | Explicitly reported | Not reported | Explicitly reported | Explicitly reported | 4 |
| O146 | Cai 2019 | Explicitly reported | Not reported | Explicitly reported | Not reported | Not reported | Not reported | Not reported | Not reported | 2 |
| O147 | Zhi 2018 | Not reported | Explicitly reported | Explicitly reported | Explicitly reported | Not reported | Not reported | Not reported | Not reported | 3 |
| O148 | Zhao 2020 | Not reported | Not reported | Explicitly reported | Not reported | Not reported | Not reported | Not reported | Not reported | 1 |
| O149 | Xin 2020 | Not reported | Not reported | Explicitly reported | Not reported | Not reported | Not reported | Explicitly reported | Explicitly reported | 3 |
| O150 | Xin 2020 | Not reported | Not reported | Not reported | Not reported | Not reported | Not reported | Not reported | Not reported | 0 |
| O151 | Shang 2020 | Not reported | Not reported | Not reported | Not reported | Not reported | Not reported | Not reported | Not reported | 0 |
| O152 | Ma 2020 | Not reported | Not reported | Explicitly reported | Not reported | Not reported | Not reported | Not reported | Not reported | 1 |
| O153 | Li 2020 | Not reported | Not reported | Explicitly reported | Not reported | Not reported | Not reported | Not reported | Not reported | 1 |
| O154 | Li 2020 | Not reported | Not reported | Not reported | Not reported | Not reported | Not reported | Not reported | Not reported | 0 |
| O155 | Li 2020 | Not reported | Not reported | Explicitly reported | Not reported | Not reported | Not reported | Not reported | Not reported | 1 |
| O156 | Huang 2020 | Not reported | Not reported | Explicitly reported | Explicitly reported | Not reported | Not reported | Explicitly reported | Not reported | 3 |
| O157 | Zhu 2019 | Not reported | Not reported | Explicitly reported | Not reported | Not reported | Not reported | Not reported | Not reported | 1 |
| O158 | Chen 2020 | Not reported | Not reported | Not reported | Not reported | Not reported | Not reported | Not reported | Not reported | 0 |
| O159 | Zhou 2021 | Not reported | Not reported | Explicitly reported | Explicitly reported | Not reported | Not reported | Not reported | Not reported | 2 |
| O160 | Zhao 2021 | Not reported | Explicitly reported | Explicitly reported | Explicitly reported | Not reported | Not reported | Not reported | Explicitly reported | 4 |
| O161 | Yu 2021 | Not reported | Not reported | Not reported | Not reported | Not reported | Not reported | Not reported | Not reported | 0 |
| O162 | Wang 2021 | Not reported | Not reported | Explicitly reported | Not reported | Not reported | Not reported | Explicitly reported | Not reported | 2 |
| O163 | Tian 2021 | Not reported | Not reported | Explicitly reported | Not reported | Not reported | Not reported | Not reported | Explicitly reported | 2 |
| O164 | Song 2021 | Not reported | Explicitly reported | Explicitly reported | Explicitly reported | Not reported | Not reported | Not reported | Explicitly reported | 4 |
| O165 | Niu 2021 | Not reported | Explicitly reported | Explicitly reported | Explicitly reported | Not reported | Not reported | Not reported | Not reported | 3 |
| O166 | Ma 2021 | Not reported | Not reported | Not reported | Explicitly reported | Not reported | Not reported | Not reported | Not reported | 1 |
| O167 | Liang 2021 | Not reported | Not reported | Not reported | Not reported | Not reported | Not reported | Not reported | Not reported | 0 |
| O168 | Li 2021 | Not reported | Not reported | Explicitly reported | Not reported | Not reported | Not reported | Explicitly reported | Not reported | 2 |
| O169 | Hong 2021 | Explicitly reported | Not reported | Explicitly reported | Not reported | Explicitly reported | Not reported | Not reported | Not reported | 3 |
| O170 | Geng 2021 | Not reported | Not reported | Explicitly reported | Not reported | Not reported | Not reported | Explicitly reported | Not reported | 2 |
| O171 | Duan 2021 | Not reported | Not reported | Explicitly reported | Explicitly reported | Not reported | Not reported | Not reported | Not reported | 2 |
| O172 | Dai 2021 | Not reported | Not reported | Explicitly reported | Not reported | Not reported | Explicitly reported | Not reported | Not reported | 2 |
| O173 | Chi 2021 | Not reported | Not reported | Explicitly reported | Not reported | Not reported | Not reported | Not reported | Not reported | 1 |
| O174 | Zhao 2022 | Not reported | Not reported | Explicitly reported | Explicitly reported | Not reported | Not reported | Not reported | Not reported | 2 |
| O175 | Zhang 2022 | Not reported | Not reported | Explicitly reported | Not reported | Not reported | Explicitly reported | Not reported | Not reported | 2 |
| O176 | Yin 2022 | Not reported | Not reported | Explicitly reported | Not reported | Not reported | Not reported | Explicitly reported | Explicitly reported | 3 |
| O177 | Wu 2022 | Not reported | Not reported | Explicitly reported | Not reported | Not reported | Not reported | Not reported | Not reported | 1 |
| O178 | Wang 2022 | Not reported | Not reported | Explicitly reported | Explicitly reported | Not reported | Explicitly reported | Not reported | Not reported | 3 |
| O179 | Sun 2022 | Not reported | Not reported | Explicitly reported | Explicitly reported | Not reported | Not reported | Not reported | Not reported | 2 |
| O180 | Lin 2014 | Not reported | Not reported | Explicitly reported | Explicitly reported | Not reported | Explicitly reported | Not reported | Explicitly reported | 4 |
| O181 | Rao 2022 | Not reported | Not reported | Not reported | Not reported | Not reported | Not reported | Not reported | Not reported | 0 |
| O182 | Li 2022 | Not reported | Not reported | Not reported | Not reported | Not reported | Not reported | Not reported | Not reported | 0 |
| O183 | Hu 2022 | Not reported | Not reported | Explicitly reported | Explicitly reported | Explicitly reported | Not reported | Not reported | Not reported | 3 |
| O184 | Guo 2022 | Not reported | Not reported | Explicitly reported | Explicitly reported | Not reported | Not reported | Not reported | Not reported | 2 |
| O185 | Du 2022 | Not reported | Not reported | Explicitly reported | Explicitly reported | Not reported | Not reported | Not reported | Not reported | 2 |
| U001 | Gao 2026 | Not reported | Explicitly reported | Not reported | Not reported | Not reported | Explicitly reported | Explicitly reported | Not reported | 3 |
| U002 | Wang 2023 | Not reported | Not reported | Explicitly reported | Explicitly reported | Not reported | Not reported | Not reported | Explicitly reported | 3 |
| U005 | Xiao 2023 | Not reported | Not reported | Not reported | Not reported | Not reported | Not reported | Not reported | Explicitly reported | 1 |
| U006 | Xiao 2026 | Not reported | Not reported | Explicitly reported | Not reported | Not reported | Explicitly reported | Not reported | Explicitly reported | 3 |
| U007 | Zhang 2026 | Not reported | Not reported | Explicitly reported | Not reported | Not reported | Not reported | Not reported | Explicitly reported | 2 |
| U008 | Zhang 2025 | Not reported | Not reported | Explicitly reported | Not reported | Not reported | Not reported | Not reported | Explicitly reported | 2 |
| U009 | Ma 2025 | Not reported | Explicitly reported | Explicitly reported | Not reported | Not reported | Not reported | Not reported | Not reported | 2 |
| U010 | Li 2024 | Explicitly reported | Explicitly reported | Explicitly reported | Not reported | Not reported | Not reported | Explicitly reported | Not reported | 4 |
| U012 | Zhang 2024 | Not reported | Not reported | Explicitly reported | Not reported | Not reported | Not reported | Not reported | Not reported | 1 |
| U013 | Miu 2023 | Not reported | Not reported | Not reported | Explicitly reported | Not reported | Not reported | Not reported | Not reported | 1 |
| U014 | Dong 2026 | Not reported | Not reported | Explicitly reported | Not reported | Not reported | Not reported | Not reported | Not reported | 1 |
| U015 | Zhang 2025 | Not reported | Explicitly reported | Explicitly reported | Explicitly reported | Not reported | Not reported | Explicitly reported | Explicitly reported | 5 |
| U016 | Ning 2024 | Not reported | Explicitly reported | Explicitly reported | Explicitly reported | Not reported | Not reported | Explicitly reported | Explicitly reported | 5 |
| U017 | Wang 2023 | Not reported | Explicitly reported | Explicitly reported | Not reported | Not reported | Not reported | Not reported | Not reported | 2 |
| U018 | Lou 2023 | Not reported | Explicitly reported | Explicitly reported | Not reported | Not reported | Not reported | Not reported | Explicitly reported | 3 |
| U019 | Lai 2024 | Not reported | Not reported | Explicitly reported | Explicitly reported | Not reported | Not reported | Not reported | Explicitly reported | 3 |
| U020 | Chen 2024 | Explicitly reported | Not reported | Explicitly reported | Explicitly reported | Not reported | Not reported | Not reported | Explicitly reported | 4 |
| U021 | Chen 2025 | Explicitly reported | Not reported | Explicitly reported | Explicitly reported | Not reported | Not reported | Explicitly reported | Explicitly reported | 5 |
| U022 | Xiong 2025 | Not reported | Not reported | Explicitly reported | Not reported | Not reported | Not reported | Not reported | Explicitly reported | 2 |
| U023 | Wei 2023 | Not reported | Explicitly reported | Explicitly reported | Explicitly reported | Explicitly reported | Not reported | Not reported | Not reported | 4 |
| U024 | Hou 2026 | Explicitly reported | Explicitly reported | Explicitly reported | Not reported | Explicitly reported | Not reported | Explicitly reported | Explicitly reported | 6 |
| U025 | Li 2023 | Not reported | Not reported | Explicitly reported | Not reported | Not reported | Not reported | Not reported | Explicitly reported | 2 |
| U026 | Zhang 2025 | Not reported | Not reported | Explicitly reported | Explicitly reported | Not reported | Explicitly reported | Not reported | Not reported | 3 |
| U027 | Li 2024 | Not reported | Not reported | Explicitly reported | Not reported | Not reported | Not reported | Not reported | Not reported | 1 |
| U028 | Sun 2025 | Not reported | Not reported | Explicitly reported | Explicitly reported | Not reported | Not reported | Not reported | Not reported | 2 |
| U029 | Hu 2026 | Not reported | Explicitly reported | Explicitly reported | Not reported | Not reported | Not reported | Not reported | Explicitly reported | 3 |
| U030 | Wang 2023 | Not reported | Not reported | Explicitly reported | Not reported | Not reported | Not reported | Not reported | Not reported | 1 |
| U031 | Wu 2024 | Not reported | Explicitly reported | Not reported | Not reported | Not reported | Not reported | Not reported | Not reported | 1 |
| U032 | Xu 2024 | Not reported | Not reported | Explicitly reported | Not reported | Explicitly reported | Explicitly reported | Not reported | Not reported | 3 |
| U033 | Han 2023 | Not reported | Not reported | Explicitly reported | Explicitly reported | Not reported | Explicitly reported | Not reported | Not reported | 3 |
| U034 | Mai 2024 | Not reported | Not reported | Explicitly reported | Not reported | Not reported | Not reported | Not reported | Not reported | 1 |
| U035 | Li 2023 | Not reported | Explicitly reported | Explicitly reported | Not reported | Not reported | Not reported | Explicitly reported | Not reported | 3 |
| U036 | Tang 2024 | Not reported | Not reported | Explicitly reported | Not reported | Not reported | Not reported | Not reported | Explicitly reported | 2 |
| U037 | Yang 2025 | Explicitly reported | Not reported | Explicitly reported | Not reported | Not reported | Not reported | Explicitly reported | Explicitly reported | 4 |
| O014 | Zhao 2015 | Explicitly reported | Explicitly reported | Explicitly reported | Explicitly reported | Explicitly reported | Explicitly reported | Explicitly reported | Not reported | 7 |
| O052 | Liu 2022 | Not reported | Explicitly reported | Explicitly reported | Not reported | Explicitly reported | Not reported | Explicitly reported | Not reported | 4 |
| S001 | Jiang 2024 | Explicitly reported | Explicitly reported | Explicitly reported | Explicitly reported | Explicitly reported | Explicitly reported | Explicitly reported | Explicitly reported | 8 |
| S002 | Xiao 2019 | Not reported | Not reported | Explicitly reported | Explicitly reported | Explicitly reported | Not reported | Explicitly reported | Explicitly reported | 5 |
| S003 | Yuan 2021 | Not reported | Not reported | Explicitly reported | Explicitly reported | Explicitly reported | Not reported | Explicitly reported | Not reported | 4 |
| S004 | Zheng 2017 | Not reported | Explicitly reported | Explicitly reported | Explicitly reported | Explicitly reported | Not reported | Explicitly reported | Not reported | 5 |
| S005 | Chu 2022 | Not reported | Explicitly reported | Explicitly reported | Explicitly reported | Explicitly reported | Not reported | Explicitly reported | Explicitly reported | 6 |
| S006 | Ke 2023 | Not reported | Explicitly reported | Explicitly reported | Explicitly reported | Explicitly reported | Not reported | Explicitly reported | Not reported | 5 |
| S007 | Liu 2021 | Explicitly reported | Explicitly reported | Explicitly reported | Explicitly reported | Explicitly reported | Not reported | Explicitly reported | Explicitly reported | 7 |
| S008 | Ji 2013 | Not reported | Not reported | Explicitly reported | Explicitly reported | Explicitly reported | Not reported | Explicitly reported | Not reported | 4 |
| S009 | Gong 2016 | Not reported | Not reported | Explicitly reported | Explicitly reported | Explicitly reported | Not reported | Explicitly reported | Not reported | 4 |
| S010 | Luo 2012 | Not reported | Not reported | Explicitly reported | Explicitly reported | Not reported | Not reported | Explicitly reported | Not reported | 3 |
| S011 | Ji 2022 | Not reported | Not reported | Explicitly reported | Not reported | Not reported | Not reported | Explicitly reported | Explicitly reported | 3 |
| S012 | Li 2015 | Not reported | Explicitly reported | Explicitly reported | Explicitly reported | Explicitly reported | Explicitly reported | Explicitly reported | Explicitly reported | 7 |
| S013 | Su 2017 | Not reported | Explicitly reported | Explicitly reported | Explicitly reported | Explicitly reported | Explicitly reported | Explicitly reported | Explicitly reported | 7 |
| S014 | Li 2019 | Not reported | Explicitly reported | Explicitly reported | Explicitly reported | Explicitly reported | Explicitly reported | Explicitly reported | Not reported | 6 |
| S015 | Yang 2016 | Not reported | Explicitly reported | Explicitly reported | Explicitly reported | Explicitly reported | Not reported | Explicitly reported | Explicitly reported | 6 |
| S016 | Ji 2020 | Not reported | Explicitly reported | Explicitly reported | Explicitly reported | Not reported | Not reported | Explicitly reported | Not reported | 4 |
| S018 | Zhan 2025 | Not reported | Explicitly reported | Explicitly reported | Explicitly reported | Explicitly reported | Explicitly reported | Explicitly reported | Explicitly reported | 7 |
| S019 | Nie 2020 | Explicitly reported | Explicitly reported | Explicitly reported | Explicitly reported | Explicitly reported | Explicitly reported | Explicitly reported | Explicitly reported | 8 |
| S020 | Ge 2025 | Explicitly reported | Explicitly reported | Explicitly reported | Explicitly reported | Explicitly reported | Explicitly reported | Explicitly reported | Explicitly reported | 8 |
| S021 | Wang 2019 | Not reported | Explicitly reported | Explicitly reported | Explicitly reported | Explicitly reported | Not reported | Explicitly reported | Not reported | 5 |
| S022 | Shen 2016 | Not reported | Explicitly reported | Explicitly reported | Explicitly reported | Explicitly reported | Not reported | Explicitly reported | Explicitly reported | 6 |
| S023 | Ran 2018 | Not reported | Explicitly reported | Explicitly reported | Explicitly reported | Explicitly reported | Not reported | Explicitly reported | Not reported | 5 |
| S024 | Ding 2025 | Not reported | Not reported | Explicitly reported | Explicitly reported | Explicitly reported | Not reported | Explicitly reported | Not reported | 4 |
| S025 | Xuan 2021 | Not reported | Explicitly reported | Explicitly reported | Explicitly reported | Explicitly reported | Not reported | Explicitly reported | Not reported | 5 |
| S026 | Xue 2012 | Not reported | Not reported | Explicitly reported | Explicitly reported | Not reported | Not reported | Explicitly reported | Not reported | 3 |
| S027 | Li 2008 | Not reported | Explicitly reported | Explicitly reported | Explicitly reported | Explicitly reported | Not reported | Explicitly reported | Not reported | 5 |
| S028 | Chen 2013 | Not reported | Explicitly reported | Explicitly reported | Explicitly reported | Explicitly reported | Not reported | Explicitly reported | Explicitly reported | 6 |
| S029 | Bai 2022 | Not reported | Explicitly reported | Explicitly reported | Explicitly reported | Explicitly reported | Not reported | Explicitly reported | Not reported | 5 |
| S030 | Yu 2014 | Not reported | Not reported | Explicitly reported | Explicitly reported | Explicitly reported | Not reported | Explicitly reported | Not reported | 4 |
| S031 | Zhang 2022 | Explicitly reported | Explicitly reported | Explicitly reported | Explicitly reported | Explicitly reported | Explicitly reported | Explicitly reported | Explicitly reported | 8 |
| S032 | Li 2015 | Not reported | Explicitly reported | Explicitly reported | Explicitly reported | Explicitly reported | Explicitly reported | Explicitly reported | Explicitly reported | 7 |
| S033 | Liang 2017 | Not reported | Not reported | Explicitly reported | Explicitly reported | Explicitly reported | Not reported | Explicitly reported | Not reported | 4 |
| S034 | Zhou 2021 | Not reported | Explicitly reported | Explicitly reported | Explicitly reported | Explicitly reported | Explicitly reported | Explicitly reported | Explicitly reported | 7 |
| S035 | Guo 2013 | Not reported | Explicitly reported | Explicitly reported | Explicitly reported | Explicitly reported | Not reported | Explicitly reported | Not reported | 5 |
| S036 | Jiao 2018 | Not reported | Not reported | Explicitly reported | Explicitly reported | Explicitly reported | Explicitly reported | Explicitly reported | Not reported | 5 |
| S037 | Li 2013 | Not reported | Explicitly reported | Explicitly reported | Explicitly reported | Explicitly reported | Explicitly reported | Explicitly reported | Explicitly reported | 7 |
| S038 | Li 2025 | Not reported | Explicitly reported | Explicitly reported | Explicitly reported | Explicitly reported | Explicitly reported | Explicitly reported | Explicitly reported | 7 |
| S039 | Bai 2019 | Not reported | Not reported | Explicitly reported | Explicitly reported | Explicitly reported | Not reported | Explicitly reported | Not reported | 4 |
| S040 | Liu 2023 | Not reported | Explicitly reported | Explicitly reported | Explicitly reported | Explicitly reported | Not reported | Explicitly reported | Not reported | 5 |
| S041 | Shi 2021 | Not reported | Not reported | Explicitly reported | Explicitly reported | Explicitly reported | Not reported | Explicitly reported | Not reported | 4 |
| S042 | Zhang 2015 | Not reported | Not reported | Explicitly reported | Explicitly reported | Explicitly reported | Not reported | Explicitly reported | Not reported | 4 |
| S043 | Lu 2024 | Explicitly reported | Explicitly reported | Explicitly reported | Explicitly reported | Explicitly reported | Explicitly reported | Explicitly reported | Explicitly reported | 8 |
| S044 | Yan 2020 | Not reported | Explicitly reported | Explicitly reported | Explicitly reported | Explicitly reported | Explicitly reported | Not reported | Not reported | 5 |
| S045 | Bai 2025 | Not reported | Explicitly reported | Explicitly reported | Explicitly reported | Explicitly reported | Not reported | Explicitly reported | Explicitly reported | 6 |
| S046 | Wang 2020 | Explicitly reported | Explicitly reported | Explicitly reported | Explicitly reported | Explicitly reported | Explicitly reported | Explicitly reported | Explicitly reported | 8 |
| S047 | Zhang 2024 | Explicitly reported | Explicitly reported | Explicitly reported | Explicitly reported | Explicitly reported | Explicitly reported | Explicitly reported | Explicitly reported | 8 |
| S049 | Guo 2017 | Explicitly reported | Explicitly reported | Explicitly reported | Explicitly reported | Explicitly reported | Explicitly reported | Explicitly reported | Explicitly reported | 8 |
| S050 | Zhang 2023 | Not reported | Explicitly reported | Explicitly reported | Explicitly reported | Explicitly reported | Not reported | Explicitly reported | Not reported | 5 |
| S051 | Xu 2022 | Not reported | Explicitly reported | Explicitly reported | Not reported | Explicitly reported | Not reported | Explicitly reported | Not reported | 4 |

Supplementary Table S10A | Descriptive trial-level evidence map using RoB 2 domains

| **Trial ID** | **Author-year** | **Randomization process** | **Deviations from intended interventions** | **Missing outcome data** | **Measurement of the outcome** | **Selection of the reported result** | **Overall Bias** |
| --- | --- | --- | --- | --- | --- | --- | --- |
| O001 | Huang 2015 | Some concerns | Low | Low | Low | Low | Some concerns |
| O002 | Zhou 2021 | Some concerns | Low | Low | Low | Low | Some concerns |
| O003 | Cheng 2022 | Some concerns | Low | Low | Low | Low | Some concerns |
| O004 | Chen 2016 | Some concerns | Low | Low | Low | Low | Some concerns |
| O005 | Pan 2017 | Some concerns | Low | Low | Low | Low | Some concerns |
| O006 | Chen 2019 | Some concerns | Low | Low | Low | Low | Some concerns |
| O007 | Chen 2020 | Some concerns | Low | Low | Low | Low | Some concerns |
| O008 | Rao 2020 | Some concerns | Low | Low | Low | Low | Some concerns |
| O009 | Liu 2022 | Some concerns | Low | Low | Low | Low | Some concerns |
| O010 | Wang 2022 | Some concerns | Low | Low | Low | Low | Some concerns |
| O011 | Tang 2007 | Low | Low | Low | Low | Low | Low |
| O012 | Zhong 2010 | Low | Low | Low | Low | Low | Low |
| O013 | Zhao 2012 | Low | Low | Low | Low | Low | Low |
| O014 | Zhao 2015 | Low | Low | Low | Low | Low | Low |
| O015 | Gan 2016 | Some concerns | Low | Low | Low | Low | Some concerns |
| O016 | Xu 2019 | Some concerns | Low | Low | Low | Low | Some concerns |
| O017 | Liu 2022 | High | Low | Low | Low | Low | High |
| O018 | Wong 2013 | Some concerns | Low | Low | Low | Low | Some concerns |
| O019 | Qin 2013 | Some concerns | Low | Low | Low | Low | Some concerns |
| O020 | Qin 2010 | Some concerns | Low | Low | Low | Low | Some concerns |
| O021 | Li 2011 | Some concerns | Low | Low | Low | Low | Some concerns |
| O022 | Zheng 2020 | Some concerns | Low | Low | Some concerns | Low | Some concerns |
| O023 | Wan 2015 | Some concerns | Low | Low | High | Low | High |
| O024 | Zhu 2018 | Some concerns | Low | Low | Low | Low | Some concerns |
| O025 | Xu 2019 | Some concerns | Low | Low | Low | Low | Some concerns |
| O026 | Wu 2019 | Some concerns | Low | Low | Some concerns | Low | Some concerns |
| O027 | Wu 2021 | Some concerns | Low | Low | Low | Low | Some concerns |
| O028 | Su 2021 | Some concerns | Low | Low | High | Low | High |
| O029 | Jiang 2022 | Some concerns | Low | Low | Low | Low | Some concerns |
| O030 | Li 2022 | Some concerns | Low | Low | Low | Low | Some concerns |
| O031 | Jin 2022 | Some concerns | Low | Low | Low | Low | Some concerns |
| O032 | Gong 2022 | Some concerns | Low | Low | Low | Low | Some concerns |
| O033 | Jiang 2021 | Some concerns | Low | Low | Low | Low | Some concerns |
| O034 | Fan 2021 | Some concerns | Low | Low | Some concerns | Low | Some concerns |
| O035 | Yue 2020 | Some concerns | Low | Low | Low | Low | Some concerns |
| O036 | Wang 2020 | Some concerns | Low | Low | Some concerns | Low | Some concerns |
| O037 | Wang 2020 | Some concerns | Low | Low | High | Low | High |
| O038 | He 2020 | Some concerns | Low | Low | Low | Low | Some concerns |
| O039 | Dai 2020 | Some concerns | Low | Low | Some concerns | Low | Some concerns |
| O040 | Wang 2019 | Some concerns | Low | Low | Low | Low | Some concerns |
| O041 | Yang 2018 | Some concerns | Low | Low | Low | Low | Some concerns |
| O042 | Wang 2017 | Some concerns | Low | Low | Low | Low | Some concerns |
| O043 | Jiang 2017 | Some concerns | Low | Low | Some concerns | Low | Some concerns |
| O044 | Cao 2017 | Some concerns | Low | Low | Some concerns | Low | Some concerns |
| O045 | Hao 2014 | Some concerns | Low | Low | Low | Low | Some concerns |
| O046 | Han 2012 | Some concerns | Low | Low | Low | Low | Some concerns |
| O047 | Wei 2022 | Some concerns | Low | Low | Low | Low | Some concerns |
| O048 | Zhuo 2022 | Some concerns | Low | Low | Some concerns | Low | Some concerns |
| O049 | Wang 2022 | Some concerns | Low | Low | Low | Low | Some concerns |
| O050 | Zhou 2022 | Some concerns | Low | Low | High | Low | High |
| O051 | Pan 2022 | Low | Low | Low | Low | Low | Low |
| O052 | Liu 2022 | Some concerns | Low | Low | Some concerns | Low | Some concerns |
| O053 | Shen 2021 | Some concerns | Low | Low | Low | Low | Some concerns |
| O054 | Lin 2013 | Some concerns | Low | Low | Low | Low | Some concerns |
| O055 | Li 2015 | Some concerns | Low | Low | Low | Low | Some concerns |
| O056 | Zhuo 2015 | Some concerns | Low | Low | Low | Low | Some concerns |
| O057 | Zhou 2016 | Some concerns | Low | Low | Some concerns | Low | Some concerns |
| O058 | Yin 2016 | Some concerns | Low | Low | High | Low | High |
| O059 | He 2016 | Some concerns | Low | Low | High | Low | High |
| O060 | Zhang 2016 | Some concerns | Low | Low | Low | Low | Some concerns |
| O061 | Peng 2017 | Some concerns | Low | Low | High | Low | High |
| O062 | Zhou 2017 | Some concerns | Low | Low | Low | Low | Some concerns |
| O063 | Ye 2017 | Some concerns | Low | Low | Low | Low | Some concerns |
| O064 | Xu 2017 | Some concerns | Low | Low | Low | Low | Some concerns |
| O065 | Hu 2017 | Some concerns | Low | Low | High | Low | High |
| O066 | Ren 2018 | Some concerns | Low | Low | Some concerns | Low | Some concerns |
| O067 | Li 2018 | Some concerns | Low | Low | Low | Low | Some concerns |
| O068 | Lei 2018 | Some concerns | Low | Low | Some concerns | Low | Some concerns |
| O069 | Zhang 2018 | Some concerns | Low | Low | Low | Low | Some concerns |
| O070 | Lyu 2018 | Some concerns | Low | Low | Low | Low | Some concerns |
| O071 | Duan 2018 | Some concerns | Low | Low | Low | Low | Some concerns |
| O128 | Ni 2018 | Some concerns | Low | Low | Some concerns | Low | Some concerns |
| O073 | Wu 2019 | Some concerns | Low | Low | Low | Low | Some concerns |
| O074 | Hu 2019 | Some concerns | Low | Low | Some concerns | Low | Some concerns |
| O075 | Feng 2019 | Some concerns | Low | Low | Some concerns | Low | Some concerns |
| O076 | Zhao 2019 | Some concerns | Low | Low | Low | Low | Some concerns |
| O077 | Zhang 2019 | Some concerns | Low | Low | Low | Low | Some concerns |
| O078 | Wang 2019 | Some concerns | Low | Low | Low | Low | Some concerns |
| O079 | Peng 2019 | Some concerns | Low | Low | Low | Low | Some concerns |
| O080 | He 2019 | Some concerns | Low | Low | Low | Low | Some concerns |
| O138 | Yao 2019 | Low | Low | Low | Low | Low | Low |
| O082 | Yang 2020 | Some concerns | Low | Low | Low | Low | Some concerns |
| O083 | Wu 2020 | High | Low | Low | Low | Low | High |
| O084 | Sun 2020 | Some concerns | Low | Low | Some concerns | Low | Some concerns |
| O085 | Liu 2020 | Some concerns | Low | Low | Low | Low | Some concerns |
| O086 | Hu 2020 | Some concerns | Low | Low | Some concerns | Low | Some concerns |
| O087 | Chen 2020 | Some concerns | Low | Low | Low | Low | Some concerns |
| O088 | Cai 2020 | Some concerns | Low | Low | Some concerns | Low | Some concerns |
| O089 | Li 2021 | Some concerns | Low | Low | Some concerns | Low | Some concerns |
| O090 | Cai 2021 | Some concerns | Low | Low | Low | Low | Some concerns |
| O091 | Mei 2021 | Some concerns | Low | Low | Low | Low | Some concerns |
| O092 | Liang 2021 | Some concerns | Low | Low | Low | Low | Some concerns |
| O093 | Chen 2021 | Some concerns | Low | Low | Low | Low | Some concerns |
| O094 | Yang 2022 | Some concerns | Low | Low | Low | Low | Some concerns |
| O095 | Ma 2022 | Some concerns | Low | Low | Low | Low | Some concerns |
| O096 | Guo 2022 | Some concerns | Low | Low | Low | Low | Some concerns |
| O097 | Dong 2022 | Some concerns | Low | Low | Low | Low | Some concerns |
| O098 | Chai 2022 | Some concerns | Low | Low | Some concerns | Low | Some concerns |
| O099 | Zhou 2021 | Some concerns | Low | Low | Low | Low | Some concerns |
| O100 | Li 2013 | Some concerns | Low | Low | Low | Low | Some concerns |
| O101 | Zhao 2013 | Some concerns | Low | Low | Some concerns | Low | Some concerns |
| O102 | Yang 2013 | Some concerns | Low | Low | Low | Low | Some concerns |
| O103 | Yao 2014 | Some concerns | Low | Low | Low | Low | Some concerns |
| O104 | Wu 2014 | Some concerns | Low | Low | Low | Low | Some concerns |
| O105 | Wen 2014 | Some concerns | Low | Low | Low | Low | Some concerns |
| O106 | Tan 2014 | Low | Low | Low | Low | Low | Low |
| O107 | Shen 2014 | Some concerns | Low | Low | Low | Low | Some concerns |
| O108 | Pei 2014 | Some concerns | Low | Low | Low | Low | Some concerns |
| O109 | Pei 2014 | Some concerns | Low | Low | Low | Low | Some concerns |
| O110 | Jin 2014 | Some concerns | Low | Low | Low | Low | Some concerns |
| O111 | Zhou 2015 | Some concerns | Low | Low | Low | Low | Some concerns |
| O112 | Zhao 2015 | Some concerns | Low | Some concerns | Some concerns | Low | Some concerns |
| O113 | Yang 2015 | Some concerns | Low | Low | Low | Low | Some concerns |
| O114 | Luo 2015 | High | Low | Low | Some concerns | Low | High |
| O115 | Liao 2015 | Some concerns | Low | Low | Low | Low | Some concerns |
| O116 | Li 2015 | Some concerns | Low | Low | Low | Low | Some concerns |
| O117 | Feng 2015 | Some concerns | Low | Low | Low | Low | Some concerns |
| O118 | Chen 2015 | Some concerns | Low | Low | Low | Low | Some concerns |
| O119 | Yu 2016 | Some concerns | Low | Low | Low | Low | Some concerns |
| O120 | Wang 2016 | Some concerns | Some concerns | Low | Some concerns | Low | Some concerns |
| O121 | Sun 2016 | Some concerns | Low | Low | Some concerns | Low | Some concerns |
| O122 | Dong 2016 | Some concerns | Low | Low | Low | Low | Some concerns |
| O123 | Zeng 2016 | Some concerns | Low | Low | Low | Low | Some concerns |
| O124 | Wen 2017 | Some concerns | Low | Low | Low | Low | Some concerns |
| O125 | Wang 2017 | Some concerns | Low | Low | Low | Low | Some concerns |
| O126 | Pan 2017 | Some concerns | Low | Low | Low | Low | Some concerns |
| O127 | Han 2017 | Some concerns | Low | Low | Low | Low | Some concerns |
| O129 | Tan 2018 | Some concerns | Low | Low | Low | Low | Some concerns |
| O130 | Lyu 2018 | Some concerns | Low | Low | Low | Low | Some concerns |
| O131 | Li 2018 | Some concerns | Low | Low | Low | Low | Some concerns |
| O132 | Huang 2018 | Low | Low | Low | Low | Low | Low |
| O133 | Hou 2018 | Some concerns | Low | Low | Low | Low | Some concerns |
| O134 | Guo 2018 | Some concerns | Low | Low | Low | Low | Some concerns |
| O135 | Fang 2018 | Some concerns | Low | Low | Low | Low | Some concerns |
| O136 | Du 2018 | Some concerns | Low | Low | Low | Low | Some concerns |
| O137 | Chen 2018 | Some concerns | Low | Low | Some concerns | Low | Some concerns |
| O139 | Wang 2019 | Low | Low | Low | Low | Low | Low |
| O140 | Mi 2019 | Low | Low | Low | Low | Low | Low |
| O141 | Liu 2019 | Some concerns | Low | Low | Low | Low | Some concerns |
| O142 | Liang 2019 | Some concerns | Some concerns | Low | Low | Low | Some concerns |
| O143 | Li 2019 | Some concerns | Low | Some concerns | Some concerns | Low | Some concerns |
| O144 | Du 2019 | Some concerns | Low | Low | Low | Low | Some concerns |
| O145 | Chen 2019 | Some concerns | Low | Low | Low | Low | Some concerns |
| O146 | Cai 2019 | Low | Low | Low | Low | Low | Low |
| O147 | Zhi 2018 | Some concerns | Low | Some concerns | Some concerns | Low | Some concerns |
| O148 | Zhao 2020 | Low | Low | Low | Low | Low | Low |
| O149 | Xin 2020 | Some concerns | Low | Low | Some concerns | Low | Some concerns |
| O150 | Xin 2020 | Some concerns | Low | Low | Low | Low | Some concerns |
| O151 | Shang 2020 | Some concerns | Low | Low | Some concerns | Low | Some concerns |
| O152 | Ma 2020 | High | Low | Low | Low | Low | High |
| O153 | Li 2020 | Some concerns | Some concerns | Low | Low | Low | Some concerns |
| O154 | Li 2020 | Some concerns | Low | Low | Low | Low | Some concerns |
| O155 | Li 2020 | Some concerns | Low | Low | Low | Low | Some concerns |
| O156 | Huang 2020 | Some concerns | Low | Low | Low | Low | Some concerns |
| O157 | Zhu 2019 | Some concerns | Low | Low | Low | Low | Some concerns |
| O158 | Chen 2020 | Some concerns | Low | Low | Low | Low | Some concerns |
| O159 | Zhou 2021 | Some concerns | Low | Low | Low | Low | Some concerns |
| O160 | Zhao 2021 | Low | Low | Low | Low | Low | Low |
| O161 | Yu 2021 | Some concerns | Low | Low | Low | Low | Some concerns |
| O162 | Wang 2021 | Some concerns | Low | Low | Low | Low | Some concerns |
| O163 | Tian 2021 | Some concerns | Low | Low | Low | Low | Some concerns |
| O164 | Song 2021 | Some concerns | Low | Low | Low | Low | Some concerns |
| O165 | Niu 2021 | Some concerns | Some concerns | Low | Low | Low | Some concerns |
| O166 | Ma 2021 | Some concerns | Low | Low | Low | Low | Some concerns |
| O167 | Liang 2021 | Some concerns | Low | Some concerns | Low | Low | Some concerns |
| O168 | Li 2021 | Some concerns | Some concerns | Low | Some concerns | Low | Some concerns |
| O169 | Hong 2021 | Some concerns | Low | Low | Low | Low | Some concerns |
| O170 | Geng 2021 | Some concerns | Low | Low | Low | Low | Some concerns |
| O171 | Duan 2021 | Some concerns | Low | Low | Low | Low | Some concerns |
| O172 | Dai 2021 | Some concerns | Low | Low | Low | Low | Some concerns |
| O173 | Chi 2021 | Some concerns | Low | Low | Low | Low | Some concerns |
| O174 | Zhao 2022 | Some concerns | Low | Low | Low | Low | Some concerns |
| O175 | Zhang 2022 | Some concerns | Low | Low | Low | Low | Some concerns |
| O176 | Yin 2022 | Some concerns | Low | Low | Low | Low | Some concerns |
| O177 | Wu 2022 | Some concerns | Some concerns | Low | Low | Low | Some concerns |
| O178 | Wang 2022 | High | Low | Low | Low | Low | High |
| O179 | Sun 2022 | Some concerns | Low | Low | Low | Low | Some concerns |
| O180 | Lin 2014 | Some concerns | Low | Low | Low | Low | Some concerns |
| O181 | Rao 2022 | Some concerns | Some concerns | Low | Low | Low | Some concerns |
| O182 | Li 2022 | Some concerns | High | Low | Low | Low | High |
| O183 | Hu 2022 | Some concerns | Some concerns | Low | Low | Low | Some concerns |
| O184 | Guo 2022 | Some concerns | Low | Low | Low | Low | Some concerns |
| O185 | Du 2022 | Some concerns | Low | Low | Some concerns | Low | Some concerns |
| U001 | Gao 2026 | Low | Low | Low | Low | Low | Low |
| U002 | Wang 2023 | Some concerns | Low | Low | Low | Low | Some concerns |
| U005 | Xiao 2023 | Some concerns | Low | Low | Low | Low | Some concerns |
| U013 | Miu 2023 | Low | Low | Low | Low | Low | Low |
| U017 | Wang 2023 | Some concerns | Low | Low | Low | Low | Some concerns |
| U018 | Lou 2023 | Low | Low | Low | Low | Low | Low |
| U010 | Li 2024 | Low | Low | Low | Low | Low | Low |
| U012 | Zhang 2024 | Low | Low | Low | Low | Low | Low |
| U016 | Ning 2024 | Low | Low | Low | Low | Low | Low |
| U008 | Zhang 2025 | Some concerns | Low | Low | Low | Low | Some concerns |
| U009 | Ma 2025 | Some concerns | Low | Low | Low | Low | Some concerns |
| U015 | Zhang 2025 | Low | Low | Low | Low | Low | Low |
| U006 | Xiao 2026 | Low | Low | Low | Low | Low | Low |
| U007 | Zhang 2026 | Low | Low | Low | Low | Low | Low |
| U014 | Dong 2026 | Low | Low | Low | Low | Low | Low |
| U019 | Lai 2024 | Low | Low | Low | Low | Low | Low |
| U020 | Chen 2024 | Some concerns | Low | Low | Low | Low | Some concerns |
| U021 | Chen 2025 | Low | Low | Low | Low | Low | Low |
| U023 | Wei 2023 | Some concerns | Low | Low | Low | Low | Some concerns |
| U025 | Li 2023 | Low | Low | Low | Low | Low | Low |
| U022 | Xiong 2025 | Low | Low | Low | Low | Low | Low |
| U024 | Hou 2026 | Low | Low | Low | Low | Low | Low |
| U030 | Wang 2023 | Low | Low | Low | Low | Low | Low |
| U027 | Li 2024 | Low | Low | Low | Low | Low | Low |
| U031 | Wu 2024 | Low | Low | Low | Low | Low | Low |
| U026 | Zhang 2025 | Low | Low | Low | Low | Low | Low |
| U028 | Sun 2025 | Low | Low | Low | Low | Low | Low |
| U029 | Hu 2026 | Low | Low | Low | Low | Low | Low |
| U033 | Han 2023 | Some concerns | Low | Low | Low | Low | Some concerns |
| U035 | Li 2023 | Low | Low | Low | Low | Low | Low |
| U032 | Xu 2024 | Low | Low | Low | Low | Low | Low |
| U034 | Mai 2024 | High | Low | Low | Low | Low | High |
| U036 | Tang 2024 | Low | Low | Low | Low | Low | Low |
| U037 | Yang 2025 | Low | Low | Low | Low | Low | Low |
| S001 | Jiang 2024 | Low | Low | Low | Low | Low | Low |
| S002 | Xiao 2019 | Low | Low | Low | Low | Low | Low |
| S003 | Yuan 2021 | Some concerns | Low | Low | Low | Low | Some concerns |
| S027 | Li 2008 | Low | Low | Low | Low | Low | Low |
| S010 | Luo 2012 | Low | Low | Low | Low | Low | Low |
| S026 | Xue 2012 | Low | Low | Low | Low | Low | Low |
| S008 | Ji 2013 | Low | Low | Low | Low | Low | Low |
| S028 | Chen 2013 | Low | Low | Low | Low | Low | Low |
| S012 | Li 2015 | Low | Low | Low | Low | Low | Low |
| S009 | Gong 2016 | Some concerns | Low | Low | Low | Low | Some concerns |
| S015 | Yang 2016 | Low | Low | Low | Low | Low | Low |
| S022 | Shen 2016 | Some concerns | Low | Low | Low | Low | Some concerns |
| S004 | Zheng 2017 | Low | Low | Low | Low | Low | Low |
| S013 | Su 2017 | Some concerns | Low | Low | Low | Low | Some concerns |
| S023 | Ran 2018 | High | Low | Low | Low | Low | High |
| S014 | Li 2019 | Some concerns | Low | Low | Low | Low | Some concerns |
| S016 | Ji 2020 | Some concerns | Low | Low | Low | Low | Some concerns |
| S021 | Wang 2019 | Some concerns | High | Low | Low | Low | High |
| S019 | Nie 2020 | Low | Low | Low | Low | Low | Low |
| S007 | Liu 2021 | Low | Low | Low | Low | Low | Low |
| S025 | Xuan 2021 | Low | Low | Low | Low | Low | Low |
| S005 | Chu 2022 | High | Low | Low | Low | Low | High |
| S011 | Ji 2022 | Some concerns | Low | Low | Low | Low | Some concerns |
| S006 | Ke 2023 | High | Low | Low | Low | Low | High |
| S018 | Zhan 2025 | Some concerns | Low | Low | Low | Low | Some concerns |
| S020 | Ge 2025 | Some concerns | Low | Low | Low | Low | Some concerns |
| S024 | Ding 2025 | Some concerns | Low | Low | Low | Low | Some concerns |
| S029 | Bai 2022 | Some concerns | Low | Low | Low | Low | Some concerns |
| S030 | Yu 2014 | Low | Low | Low | Low | Low | Low |
| S031 | Zhang 2022 | Low | Low | Low | Low | Low | Low |
| S035 | Guo 2013 | Some concerns | Low | Low | Low | Low | Some concerns |
| S037 | Li 2013 | Some concerns | Low | Low | Low | Low | Some concerns |
| S032 | Li 2015 | Low | Low | Low | Low | Low | Low |
| S042 | Zhang 2015 | Some concerns | Low | Low | Low | Low | Some concerns |
| S033 | Liang 2017 | Some concerns | Low | Low | Low | Low | Some concerns |
| S036 | Jiao 2018 | Some concerns | Low | Low | Low | Low | Some concerns |
| S039 | Bai 2019 | Low | Low | Low | Low | Low | Low |
| S034 | Zhou 2021 | Low | Low | Low | Low | Low | Low |
| S041 | Shi 2021 | Low | Low | Low | Low | Low | Low |
| S040 | Liu 2023 | Low | Low | Low | High | Some concerns | High |
| S043 | Lu 2024 | Some concerns | Low | Low | Low | Low | Some concerns |
| S038 | Li 2025 | Low | Low | Low | Low | Low | Low |
| S049 | Guo 2017 | Low | Low | Low | Low | Low | Some concerns |
| S044 | Yan 2020 | Some concerns | Low | Low | Low | Low | Some concerns |
| S046 | Wang 2020 | Some concerns | Low | Low | Low | Low | Some concerns |
| S051 | Xu 2022 | Some concerns | Low | Low | Low | Low | Some concerns |
| S050 | Zhang 2023 | Low | Low | Low | Low | Low | Low |
| S047 | Zhang 2024 | Some concerns | Low | Low | Low | Low | Some concerns |
| S045 | Bai 2025 | Low | Low | Low | Low | Low | Low |

Supplementary Table S10B | Outcome-specific RoB 2 judgements for low-risk sensitivity analyses

| **Trial ID** | **Author-year** | **Outcome** | **Randomization process** | **Deviations from intended interventions** | **Missing outcome data** | **Measurement of the outcome** | **Selection of the reported result** | **Overall Bias** |
| --- | --- | --- | --- | --- | --- | --- | --- | --- |
| O013 | Zhao 2012 | Adherence | Low | Low | Low | Low | Low | Low |
| O089 | Li 2021 | Adherence | Low | Low | Low | Low | Low | Low |
| O018 | Wong 2013 | Adherence | Low | Low | Low | Low | Low | Low |
| O016 | Xu 2019 | Readmission | Low | Low | Low | Low | Low | Low |
| O046 | Han 2012 | Adherence | Low | Low | Low | Low | Low | Low |
| O079 | Peng 2019 | Adherence | Low | Low | Low | Low | Low | Low |
| O184 | Guo 2022 | Adherence | Low | Low | Low | Low | Low | Low |
| O156 | Huang 2020 | Adherence | Low | Low | Low | Low | Low | Low |
| O142 | Liang 2019 | ADR | Low | Low | Low | Low | Low | Low |
| O143 | Li 2019 | ADR | Low | Low | Low | Low | Low | Low |
| O143 | Li 2019 | Adherence | Low | Low | Low | Low | Low | Low |
| O130 | Lyu 2018 | ADR | Low | Low | Low | Low | Low | Low |
| O130 | Lyu 2018 | Adherence | Low | Low | Low | Low | Low | Low |
| O132 | Huang 2018 | Adherence | Low | Low | Low | Low | Low | Low |
| O123 | Zeng 2016 | ADR | Low | Low | Low | Low | Low | Low |
| O123 | Zeng 2016 | Adherence | Low | Low | Low | Low | Low | Low |
| U001 | Gao 2026 | Adherence | Low | Low | Low | Low | Low | Low |
| U008 | Zhang 2025 | Readmission | Low | Low | Low | Low | Low | Low |
| U010 | Li 2024 | ADR | Low | Low | Low | Low | Low | Low |
| U014 | Dong 2026 | Adherence | Low | Low | Low | Low | Low | Low |
| U015 | Zhang 2025 | ADR | Low | Low | Low | Low | Low | Low |
| U017 | Wang 2023 | ADR | Low | Low | Low | Low | Low | Low |
| U022 | Xiong 2025 | Readmission | Low | Low | Low | Low | Low | Low |
| U024 | Hou 2026 | ADR | Low | Low | Low | Low | Low | Low |
| U019 | Lai 2024 | ADR | Low | Low | Low | Low | Low | Low |
| U026 | Zhang 2025 | ADR | Low | Low | Low | Low | Low | Low |
| U026 | Zhang 2025 | Adherence | Low | Low | Low | Low | Low | Low |
| U028 | Sun 2025 | ADR | Low | Low | Low | Low | Low | Low |
| U028 | Sun 2025 | Adherence | Low | Low | Low | Low | Low | Low |
| U029 | Hu 2026 | Adherence | Low | Low | Low | Low | Low | Low |
| U030 | Wang 2023 | Readmission | Low | Low | Low | Low | Low | Low |
| U030 | Wang 2023 | ADR | Low | Low | Low | Low | Low | Low |
| U030 | Wang 2023 | Adherence | Low | Low | Low | Low | Low | Low |
| U031 | Wu 2024 | ADR | Low | Low | Low | Low | Low | Low |
| U031 | Wu 2024 | Adherence | Low | Low | Low | Low | Low | Low |
| U033 | Han 2023 | Adherence | Low | Low | Low | Low | Low | Low |
| U020 | Chen 2024 | ADR | Low | Low | Low | Low | Low | Low |
| U021 | Chen 2025 | ADR | Low | Low | Low | Low | Low | Low |
| S001 | Jiang 2024 | ADR | Low | Low | Low | Low | Low | Low |
| S004 | Zheng 2017 | Adherence | Low | Low | Low | Low | Low | Low |
| S005 | Chu 2022 | Adherence | Low | Low | Low | Low | Low | Low |
| S011 | Ji 2022 | ADR | Low | Low | Low | Low | Low | Low |
| S018 | Zhan 2025 | Adherence | Low | Low | Low | Low | Low | Low |
| S019 | Nie 2020 | Adherence | Low | Low | Low | Low | Low | Low |
| S020 | Ge 2025 | Adherence | Low | Low | Low | Low | Low | Low |
| S034 | Zhou 2021 | Adherence | Low | Low | Low | Low | Low | Low |
| S038 | Li 2025 | ADR | Low | Low | Low | Low | Low | Low |
| S040 | Liu 2023 | Adherence | Low | Low | Low | Low | Low | Low |
| S049 | Guo 2017 | ADR | Low | Low | Low | Low | Low | Low |
| S049 | Guo 2017 | Adherence | Low | Low | Low | Low | Low | Low |
| S051 | Xu 2022 | ADR | Low | Low | Low | Low | Low | Low |
| S051 | Xu 2022 | Adherence | Low | Low | Low | Low | Low | Low |

These 52 result-level assessments covered the studies retained in the outcome-specific low-risk sensitivity analyses: 28 for adherence, 20 for reported adverse drug reactions, and four for readmission. All 52 results were judged low risk across all five RoB 2 domains and overall. The preceding 266-trial table is a descriptive evidence map of reporting and design features and was not used in place of these outcome-specific judgements.

### Additional outcome-specific methods

Chinese-language journal reports were retained when the journal appeared in at least one of the Peking University core list (2023 edition), CSSCI source-journal list (2023-2024), or Chinese Science and Technology Core list (2024 edition). English-language reports were not filtered by these lists. The three-list union was used so that a journal needed to appear in only one list.

Unplanned readmission was analysed by follow-up window and per 6-month increase in follow-up. The risk of bias due to missing evidence (ROB-ME) framework informed the assessment of missing evidence. For continuous outcomes with more than one eligible assessment, the final reported follow-up value was selected; the locked analytical datasets contained no missing outcome values requiring imputation. The continuous-outcome audit recomputed mean differences and variances and used studentised residuals, Cook's distance, and Baujat diagnostics. Reported adherence was examined on RR, OR, and risk-difference scales.

Supplementary Table S11A | Unplanned readmission by follow-up window

| **Window** | **k** | **Trials** | **RR** | **CI lower** | **CI upper** | **PI lower** | **PI upper** | **I² (%)** |
| --- | --- | --- | --- | --- | --- | --- | --- | --- |
| Overall | 22 | 22 | 0.524 | 0.457 | 0.601 | 0.417 | 0.659 | 9.969 |
| <=3 months | 4 | 4 | 0.425 | 0.16 | 1.132 | 0.16 | 1.132 | 0.001 |
| >3-6 months | 7 | 7 | 0.477 | 0.384 | 0.592 | 0.35 | 0.651 | 11.236 |
| >6 months | 10 | 10 | 0.577 | 0.475 | 0.701 | 0.475 | 0.701 | 0.003 |

Supplementary Table S11B | Readmission follow-up moderator tests

| **Analysis** | **k** | **Estimate** | **P value** | **BH q value** |
| --- | --- | --- | --- | --- |
| Per 6-month increase | 21 | Ratio of RRs 1.11 (1.01 to 1.22) | 0.033 | 0.067 |
| Three follow-up windows, omnibus | 21 | Qm=1.34, df=2 | 0.287 | 0.287 |

Supplementary Table S11C | ROB-ME-informed assessment for readmission

| **Domain** | **Judgement** | **Rationale** |
| --- | --- | --- |
| Eligible studies with missing readmission results | 244 of 266 trials did not contribute a readmission result; outcome eligibility or intent was usually not documented | Non-reporting alone was not treated as proof that a result was measured and suppressed. |
| Availability of protocols or analysis plans | Generally unavailable in the extracted evidence base | The absence of prespecified outcome documents limits result-level verification. |
| Evidence that missingness depended on result direction or significance | No direct evidence; selective outcome non-reporting cannot be excluded | Observed reporting was concentrated in selected clinical populations and follow-up windows. |
| Small-study-effect evidence | Peters P=0.109 | An asymmetry test cannot detect selective outcome non-reporting within published trials. |
| Overall ROB-ME-informed judgement | Some concerns | Uncertainty remains, but the available evidence does not establish a high risk of bias due to missing evidence. |

Supplementary Table S11D | Profile of readmission reporting

| **Disease category** | **Total included trials** | **Trials reporting readmission** | **Trials not reporting readmission** | **Reporting proportion (%)** |
| --- | --- | --- | --- | --- |
| Overall | 266 | 22 | 244 | 8.3 |
| Coronary disease | 9 | 3 | 6 | 33.3 |
| Diabetes mellitus | 124 | 4 | 120 | 3.2 |
| Dyslipidaemia | 6 | 0 | 6 | 0.0 |
| Heart failure | 15 | 9 | 6 | 60.0 |
| Hypertension | 67 | 2 | 65 | 3.0 |
| Mixed cardiometabolic conditions | 39 | 1 | 38 | 2.6 |
| Myocardial infarction | 6 | 3 | 3 | 50.0 |

Supplementary Table S11E | Unplanned readmission by disease spectrum

| **Disease spectrum** | **Trials** | **RR** | **CI lower** | **CI upper** | **PI lower** | **PI upper** | **I² (%)** | **Random-effects weight (%)** |
| --- | --- | --- | --- | --- | --- | --- | --- | --- |
| Diabetes or hypertension | 6 | 0.318 | 0.096 | 1.054 | 0.03 | 3.409 | 78.88 | 18.115 |
| Heart failure, myocardial infarction, or coronary disease | 15 | 0.57 | 0.511 | 0.635 | 0.511 | 0.635 | 0 | 72.604 |
| Mixed cardiometabolic conditions | 1 | 0.37 | 0.26 | 0.527 | Not estimable | Not estimable | 0 | 9.28 |

Note: A prediction interval was not estimated for the one-trial mixed-condition subgroup. In the 15-trial heart-failure, myocardial-infarction, or coronary-disease subgroup, τ² was estimated at the boundary value of zero, so the conventional prediction interval collapsed to the confidence interval; this should not be read as proof of homogeneous effects.

The omnibus interaction P value was 0.105. The mixed-condition stratum contained one trial.

Supplementary Table S12A | Continuous-outcome data and influence audit

| **Outcome** | **k, all** | **Source-data flags** | **Influential trials removed** | **MD, all** | **95% CI, all** | **I², all (%)** | **MD, influence sensitivity** | **95% CI, influence sensitivity** | **I², sensitivity (%)** |
| --- | --- | --- | --- | --- | --- | --- | --- | --- | --- |
| SBP | 91 | 0 | 3 | -8.83 | -10.08 to -7.58 | 97.66 | -8.65 | -9.71 to -7.59 | 96.594 |
| DBP | 87 | 0 | 1 | -6.616 | -7.47 to -5.76 | 97.242 | -6.756 | -7.58 to -5.94 | 96.979 |
| FPG | 137 | 0 | 9 | -1.241 | -1.38 to -1.10 | 97.741 | -1.115 | -1.23 to -1.00 | 96.09 |
| 2hPG | 105 | 0 | 3 | -1.79 | -2.02 to -1.56 | 97.631 | -1.688 | -1.90 to -1.48 | 97.052 |
| HbA1c | 110 | 0 | 3 | -1.088 | -1.24 to -0.93 | 97.916 | -1.019 | -1.16 to -0.88 | 97.292 |
| TC | 28 | 0 | 1 | -0.656 | -0.86 to -0.45 | 96.716 | -0.606 | -0.80 to -0.41 | 95.928 |
| TG | 29 | 0 | 2 | -0.441 | -0.65 to -0.23 | 99.266 | -0.314 | -0.43 to -0.19 | 97.56 |
| LDL-C | 35 | 0 | 2 | -0.645 | -0.89 to -0.40 | 99.503 | -0.508 | -0.68 to -0.34 | 98.787 |
| HDL-C | 27 | 0 | 2 | 0.128 | 0.01 to 0.25 | 98.886 | 0.067 | -0.02 to 0.16 | 97.52 |

Supplementary Table S12B | Statistically influential continuous-outcome estimates

Trial IDs correspond to the canonical trial identifiers used throughout the Supplementary Material.

| **Outcome** | **Trial ID** | **Author** | **Year** | **MD** | **Variance** | **Studentised residual** | **Cook's distance** | **Baujat Q contribution** | **Baujat effect influence** |
| --- | --- | --- | --- | --- | --- | --- | --- | --- | --- |
| SBP | O085 | Liu | 2020 | -23.14 | 3.725 | -2.478 | 0.063 | 5.609 | 0.065 |
| SBP | O063 | Ye | 2017 | -30 | 1.521 | -3.988 | 0.153 | 13.065 | 0.176 |
| SBP | U036 | Tang | 2024 | 10.07 | 2.738 | 3.418 | 0.127 | 10.056 | 0.141 |
| DBP | U036 | Tang | 2024 | 5.78 | 2.69 | 3.152 | 0.106 | 8.917 | 0.116 |
| FPG | O159 | Zhou | 2021 | -3.6 | 0.021 | -3.048 | 0.077 | 8.645 | 0.081 |
| FPG | O150 | Xin | 2020 | -3.29 | 0.077 | -2.504 | 0.047 | 6 | 0.049 |
| FPG | O132 | Huang | 2018 | -3.4 | 0.067 | -2.668 | 0.055 | 6.764 | 0.057 |
| FPG | O137 | Chen | 2018 | -2.93 | 0.078 | -2.048 | 0.032 | 4.072 | 0.032 |
| FPG | O111 | Zhou | 2015 | -3.5 | 0.065 | -2.803 | 0.061 | 7.427 | 0.063 |
| FPG | O117 | Feng | 2015 | -3.1 | 0.062 | -2.289 | 0.04 | 5.048 | 0.041 |
| FPG | O109 | Pei | 2014 | -3.2 | 0.068 | -2.407 | 0.044 | 5.558 | 0.046 |
| FPG | U032 | Xu | 2024 | -3.59 | 0.086 | -2.871 | 0.061 | 7.786 | 0.064 |
| FPG | S046 | Wang | 2020 | 0.29 | 0.004 | 1.964 | 0.029 | 3.744 | 0.03 |
| 2hPG | O149 | Xin | 2020 | -4.3 | 0.203 | -2.092 | 0.041 | 4.05 | 0.042 |
| 2hPG | O137 | Chen | 2018 | -4.62 | 0.07 | -2.493 | 0.064 | 5.633 | 0.067 |
| 2hPG | O111 | Zhou | 2015 | -6 | 0.068 | -3.86 | 0.15 | 12.479 | 0.168 |
| HbA1c | O158 | Chen | 2020 | -2.9 | 0.105 | -2.189 | 0.044 | 4.677 | 0.045 |
| HbA1c | O111 | Zhou | 2015 | -4.4 | 0.047 | -4.473 | 0.211 | 17.025 | 0.247 |
| HbA1c | S008 | Ji | 2013 | -2.92 | 0.144 | -2.15 | 0.04 | 4.527 | 0.041 |
| TC | S030 | Yu | 2014 | -1.95 | 0.02 | -2.781 | 0.245 | 5.801 | 0.294 |
| TG | O024 | Zhu | 2018 | -2.52 | 0.011 | -5.887 | 0.715 | 14.73 | 1.514 |
| TG | O121 | Sun | 2016 | -1.5 | 0.034 | -2.021 | 0.128 | 3.548 | 0.138 |
| LDL-C | O024 | Zhu | 2018 | -2.96 | 0.01 | -4.19 | 0.458 | 11.395 | 0.68 |
| LDL-C | S032 | Li | 2015 | -2.55 | 0.088 | -2.764 | 0.181 | 6.615 | 0.213 |
| HDL-C | U036 | Tang | 2024 | 0.68 | 0.005 | 1.96 | 0.164 | 3.704 | 0.18 |
| HDL-C | S032 | Li | 2015 | 1.07 | 0.021 | 3.334 | 0.337 | 9.016 | 0.486 |

Supplementary Table S13A | Adherence effect-scale sensitivity

| **Scale** | **k** | **Trials** | **Effect** | **CI lower** | **CI upper** | **PI lower** | **PI upper** | **I² (%)** |
| --- | --- | --- | --- | --- | --- | --- | --- | --- |
| RR | 163 | 161 | 1.37 | 1.318 | 1.425 | 0.975 | 1.926 | 80.221 |
| OR | 163 | 161 | 4.781 | 4.333 | 5.275 | 2.134 | 10.714 | 41.616 |
| RD | 163 | 161 | 0.235 | 0.216 | 0.254 | 0.031 | 0.439 | 73.534 |

Supplementary Table S13B | Audit of identifiable adherence measurement methods across all included trials

| **Measurement category** | **Trials** | **% of 266 trials** | **Interpretive category** |
| --- | --- | --- | --- |
| Morisky Medication Adherence Scale | 110 | 41.4% | Named self-report instrument |
| Medication Adherence Rating Scale, 5-item (MARS-5) | 3 | 1.1% | Named self-report instrument |
| Treatment Adherence Scale for Hypertensive Patients | 2 | 0.8% | Named self-report instrument |
| Investigator-developed scale or definition | 104 | 39.1% | Unvalidated self-report |
| Dispensing-based objective measure (proportion of days covered or medication possession ratio) | 8 | 3.0% | Objective, indirect |
| Adherence inferred from a physiological marker | 1 | 0.4% | Objective, indirect |
| Adherence not reported | 38 | 14.3% | Not applicable |

Among the 228 trials that reported adherence, 115 (50.4%) used a named self-report instrument, 104 (45.6%) used an investigator-developed questionnaire or definition, and nine (3.9%) used an objective measure: eight dispensing-based and one inferred from a physiological marker. The Treatment Adherence Scale for Hypertensive Patients is an instrument developed to assess treatment adherence in Chinese patients with hypertension. Measurement type was ascertained from the original report rather than from the extraction database; two reviewers read each report independently and a third adjudicated disagreements. This audit spans all included trials and is not limited to those contributing the adherence syntheses. Only sufficiently comparable binary adherence rates and MMAS-8 scores were pooled; bespoke continuous scores using incompatible scoring ranges or definitions remained in the evidence map. Adherence rate and MMAS-8 score were analysed as separate outcomes, so the two trials reporting both contributed one estimate to each.

Supplementary Table S14A | Chinese core-index sources

| **Index** | **List** | **Edition** | **Verification source** |
| --- | --- | --- | --- |
| Peking University core | Guide to the Core Journals of China | 2023 edition | https://lib.bjut.edu.cn/info/1475/2489.htm |
| CSSCI | Chinese Social Sciences Citation Index source journals | 2023-2024 | https://cssrac.nju.edu.cn/gywm/zxjj/index.html |
| Chinese Science and Technology Core | Chinese Science and Technology Core Journals | 2024 edition | https://lib.bjut.edu.cn/info/1475/2609.htm |

Supplementary Table S14B | Publication language of included trials

| **Publication_language** | **Trials** | **Percent** |
| --- | --- | --- |
| Chinese | 254 | 95.489 |
| English | 12 | 4.511 |

Supplementary Table S15A | INSPECT-SR summary for the three core outcomes

Author-defined operational rule for summarising unresolved INSPECT-SR checks: all 21 checks were reviewed. An unresolved item was retained as unclear and led to some concerns in its domain. Any domain with some concerns led to an overall judgement of some concerns. Serious concerns required strong evidence of an integrity problem. The 211 some-concern judgements arose in conduct, governance, and transparency, principally because ethics information could not be verified in 157 trials and registration or report-to-registration concordance could not be verified in 211.

| **Outcome set** | **Trials screened** | **No concerns** | **Some concerns** | **Serious concerns** |
| --- | --- | --- | --- | --- |
| All three core outcomes | 238 | 27 | 211 | 0 |
| Reported adherence | 187 | 18 | 169 | 0 |
| Reported adverse drug reactions | 93 | 9 | 84 | 0 |
| Unplanned readmission | 22 | 7 | 15 | 0 |

INSPECT-SR is a report-trustworthiness screen and does not replace RoB 2 or GRADE.

Supplementary Table S15B | Trial-level INSPECT-SR judgements

| **Trial ID** | **Author-year** | **Core outcomes** | **Post-publication notices** | **Conduct, governance, and transparency** | **Text and figures** | **Results** | **Overall** |
| --- | --- | --- | --- | --- | --- | --- | --- |
| O001 | Huang 2015 | unplanned readmission during follow-up | No concerns | Some concerns | No concerns | No concerns | Some concerns |
| O002 | Zhou 2021 | MMAS-8 adherence score; reported adverse drug reactions | No concerns | Some concerns | No concerns | No concerns | Some concerns |
| O003 | Cheng 2022 | reported adverse drug reactions; unplanned readmission during follow-up | No concerns | No concerns | No concerns | No concerns | No concerns |
| O004 | Chen 2016 | unplanned readmission during follow-up | No concerns | Some concerns | No concerns | No concerns | Some concerns |
| O005 | Pan 2017 | reported adherence rate; unplanned readmission during follow-up | No concerns | Some concerns | No concerns | No concerns | Some concerns |
| O006 | Chen 2019 | reported adherence rate; unplanned readmission during follow-up | No concerns | Some concerns | No concerns | No concerns | Some concerns |
| O007 | Chen 2020 | unplanned readmission during follow-up | No concerns | Some concerns | No concerns | No concerns | Some concerns |
| O008 | Rao 2020 | unplanned readmission during follow-up | No concerns | No concerns | No concerns | No concerns | No concerns |
| O009 | Liu 2022 | reported adverse drug reactions | No concerns | Some concerns | No concerns | No concerns | Some concerns |
| O010 | Wang 2022 | reported adherence rate | No concerns | Some concerns | No concerns | No concerns | Some concerns |
| O011 | Tang 2007 | reported adherence rate | No concerns | Some concerns | No concerns | No concerns | Some concerns |
| O012 | Zhong 2010 | reported adherence rate | No concerns | Some concerns | No concerns | No concerns | Some concerns |
| O013 | Zhao 2012 | reported adherence rate | No concerns | Some concerns | No concerns | No concerns | Some concerns |
| O014 | Zhao 2015 | reported adverse drug reactions | No concerns | Some concerns | No concerns | No concerns | Some concerns |
| O015 | Gan 2016 | unplanned readmission during follow-up | No concerns | Some concerns | No concerns | No concerns | Some concerns |
| O016 | Xu 2019 | unplanned readmission during follow-up | No concerns | No concerns | No concerns | No concerns | No concerns |
| O017 | Liu 2022 | MMAS-8 adherence score | No concerns | Some concerns | No concerns | No concerns | Some concerns |
| O018 | Wong 2013 | MMAS-8 adherence score; reported adherence rate | No concerns | Some concerns | No concerns | No concerns | Some concerns |
| O019 | Qin 2013 | reported adherence rate | No concerns | Some concerns | No concerns | No concerns | Some concerns |
| O020 | Qin 2010 | reported adherence rate | No concerns | Some concerns | No concerns | No concerns | Some concerns |
| O021 | Li 2011 | reported adverse drug reactions | No concerns | Some concerns | No concerns | No concerns | Some concerns |
| O022 | Zheng 2020 | reported adherence rate | No concerns | Some concerns | No concerns | No concerns | Some concerns |
| O023 | Wan 2015 | reported adherence rate | No concerns | Some concerns | No concerns | No concerns | Some concerns |
| O024 | Zhu 2018 | reported adherence rate; reported adverse drug reactions | No concerns | Some concerns | No concerns | No concerns | Some concerns |
| O025 | Xu 2019 | reported adherence rate; unplanned readmission during follow-up | No concerns | Some concerns | No concerns | No concerns | Some concerns |
| O026 | Wu 2019 | reported adherence rate | No concerns | Some concerns | No concerns | No concerns | Some concerns |
| O027 | Wu 2021 | reported adherence rate; unplanned readmission during follow-up | No concerns | No concerns | No concerns | No concerns | No concerns |
| O028 | Su 2021 | MMAS-8 adherence score; unplanned readmission during follow-up | No concerns | Some concerns | No concerns | No concerns | Some concerns |
| O029 | Jiang 2022 | MMAS-8 adherence score; reported adverse drug reactions | No concerns | Some concerns | No concerns | No concerns | Some concerns |
| O031 | Jin 2022 | reported adherence rate; reported adverse drug reactions | No concerns | Some concerns | No concerns | No concerns | Some concerns |
| O032 | Gong 2022 | MMAS-8 adherence score | No concerns | No concerns | No concerns | No concerns | No concerns |
| O034 | Fan 2021 | MMAS-8 adherence score | No concerns | No concerns | No concerns | No concerns | No concerns |
| O035 | Yue 2020 | reported adherence rate; reported adverse drug reactions | No concerns | Some concerns | No concerns | No concerns | Some concerns |
| O037 | Wang 2020 | reported adherence rate; unplanned readmission during follow-up | No concerns | Some concerns | No concerns | No concerns | Some concerns |
| O038 | He 2020 | reported adherence rate | No concerns | Some concerns | No concerns | No concerns | Some concerns |
| O039 | Dai 2020 | reported adverse drug reactions | No concerns | Some concerns | No concerns | No concerns | Some concerns |
| O040 | Wang 2019 | reported adherence rate | No concerns | Some concerns | No concerns | No concerns | Some concerns |
| O041 | Yang 2018 | reported adherence rate | No concerns | Some concerns | No concerns | No concerns | Some concerns |
| O042 | Wang 2017 | reported adherence rate; reported adverse drug reactions | No concerns | Some concerns | No concerns | No concerns | Some concerns |
| O043 | Jiang 2017 | reported adherence rate; reported adverse drug reactions | No concerns | Some concerns | No concerns | No concerns | Some concerns |
| O044 | Cao 2017 | MMAS-8 adherence score; reported adverse drug reactions | No concerns | Some concerns | No concerns | No concerns | Some concerns |
| O045 | Hao 2014 | reported adherence rate | No concerns | Some concerns | No concerns | No concerns | Some concerns |
| O046 | Han 2012 | reported adherence rate | No concerns | Some concerns | No concerns | No concerns | Some concerns |
| O047 | Wei 2022 | reported adherence rate | No concerns | Some concerns | No concerns | No concerns | Some concerns |
| O048 | Zhuo 2022 | reported adherence rate | No concerns | Some concerns | No concerns | No concerns | Some concerns |
| O049 | Wang 2022 | reported adherence rate | No concerns | No concerns | No concerns | No concerns | No concerns |
| O050 | Zhou 2022 | reported adherence rate | No concerns | Some concerns | No concerns | No concerns | Some concerns |
| O051 | Pan 2022 | MMAS-8 adherence score; reported adverse drug reactions | No concerns | Some concerns | No concerns | No concerns | Some concerns |
| O052 | Liu 2022 | reported adherence rate; reported adverse drug reactions | No concerns | Some concerns | No concerns | No concerns | Some concerns |
| O053 | Shen 2021 | reported adherence rate; reported adverse drug reactions | No concerns | Some concerns | No concerns | No concerns | Some concerns |
| O054 | Lin 2013 | reported adherence rate | No concerns | Some concerns | No concerns | No concerns | Some concerns |
| O055 | Li 2015 | reported adherence rate | No concerns | Some concerns | No concerns | No concerns | Some concerns |
| O056 | Zhuo 2015 | reported adherence rate | No concerns | Some concerns | No concerns | No concerns | Some concerns |
| O057 | Zhou 2016 | reported adherence rate | No concerns | Some concerns | No concerns | No concerns | Some concerns |
| O058 | Yin 2016 | reported adherence rate | No concerns | Some concerns | No concerns | No concerns | Some concerns |
| O059 | He 2016 | reported adherence rate | No concerns | Some concerns | No concerns | No concerns | Some concerns |
| O060 | Zhang 2016 | reported adherence rate | No concerns | Some concerns | No concerns | No concerns | Some concerns |
| O061 | Peng 2017 | reported adherence rate | No concerns | Some concerns | No concerns | No concerns | Some concerns |
| O062 | Zhou 2017 | reported adherence rate | No concerns | Some concerns | No concerns | No concerns | Some concerns |
| O063 | Ye 2017 | reported adherence rate | No concerns | Some concerns | No concerns | No concerns | Some concerns |
| O064 | Xu 2017 | reported adherence rate | No concerns | Some concerns | No concerns | No concerns | Some concerns |
| O065 | Hu 2017 | reported adherence rate | No concerns | Some concerns | No concerns | No concerns | Some concerns |
| O066 | Ren 2018 | reported adherence rate | No concerns | Some concerns | No concerns | No concerns | Some concerns |
| O067 | Li 2018 | reported adherence rate | No concerns | Some concerns | No concerns | No concerns | Some concerns |
| O068 | Lei 2018 | reported adherence rate | No concerns | Some concerns | No concerns | No concerns | Some concerns |
| O069 | Zhang 2018 | reported adherence rate | No concerns | Some concerns | No concerns | No concerns | Some concerns |
| O070 | Lyu 2018 | reported adherence rate | No concerns | Some concerns | No concerns | No concerns | Some concerns |
| O071 | Duan 2018 | reported adverse drug reactions | No concerns | Some concerns | No concerns | No concerns | Some concerns |
| O073 | Wu 2019 | reported adherence rate | No concerns | Some concerns | No concerns | No concerns | Some concerns |
| O074 | Hu 2019 | reported adherence rate | No concerns | Some concerns | No concerns | No concerns | Some concerns |
| O075 | Feng 2019 | reported adherence rate | No concerns | Some concerns | No concerns | No concerns | Some concerns |
| O076 | Zhao 2019 | reported adverse drug reactions; unplanned readmission during follow-up | No concerns | Some concerns | No concerns | No concerns | Some concerns |
| O077 | Zhang 2019 | reported adherence rate | No concerns | Some concerns | No concerns | No concerns | Some concerns |
| O078 | Wang 2019 | MMAS-8 adherence score | No concerns | Some concerns | No concerns | No concerns | Some concerns |
| O079 | Peng 2019 | reported adherence rate | No concerns | No concerns | No concerns | No concerns | No concerns |
| O080 | He 2019 | reported adherence rate | No concerns | Some concerns | No concerns | No concerns | Some concerns |
| O082 | Yang 2020 | reported adherence rate | No concerns | Some concerns | No concerns | No concerns | Some concerns |
| O083 | Wu 2020 | reported adherence rate | No concerns | Some concerns | No concerns | No concerns | Some concerns |
| O084 | Sun 2020 | reported adherence rate | No concerns | Some concerns | No concerns | No concerns | Some concerns |
| O085 | Liu 2020 | reported adherence rate | No concerns | Some concerns | No concerns | No concerns | Some concerns |
| O086 | Hu 2020 | reported adherence rate | No concerns | Some concerns | No concerns | No concerns | Some concerns |
| O087 | Chen 2020 | reported adherence rate | No concerns | Some concerns | No concerns | No concerns | Some concerns |
| O088 | Cai 2020 | reported adherence rate | No concerns | Some concerns | No concerns | No concerns | Some concerns |
| O089 | Li 2021 | reported adherence rate | No concerns | No concerns | No concerns | No concerns | No concerns |
| O090 | Cai 2021 | reported adherence rate | No concerns | Some concerns | No concerns | No concerns | Some concerns |
| O091 | Mei 2021 | reported adherence rate | No concerns | Some concerns | No concerns | No concerns | Some concerns |
| O092 | Liang 2021 | reported adherence rate; reported adverse drug reactions | No concerns | Some concerns | No concerns | No concerns | Some concerns |
| O093 | Chen 2021 | reported adverse drug reactions | No concerns | Some concerns | No concerns | No concerns | Some concerns |
| O094 | Yang 2022 | reported adherence rate; reported adverse drug reactions | No concerns | Some concerns | No concerns | No concerns | Some concerns |
| O095 | Ma 2022 | reported adverse drug reactions | No concerns | Some concerns | No concerns | No concerns | Some concerns |
| O096 | Guo 2022 | reported adherence rate; reported adverse drug reactions | No concerns | Some concerns | No concerns | No concerns | Some concerns |
| O097 | Dong 2022 | reported adherence rate | No concerns | Some concerns | No concerns | No concerns | Some concerns |
| O098 | Chai 2022 | reported adherence rate | No concerns | Some concerns | No concerns | No concerns | Some concerns |
| O099 | Zhou 2021 | reported adherence rate | No concerns | Some concerns | No concerns | No concerns | Some concerns |
| O100 | Li 2013 | reported adverse drug reactions; unplanned readmission during follow-up | No concerns | Some concerns | No concerns | No concerns | Some concerns |
| O101 | Zhao 2013 | reported adverse drug reactions | No concerns | Some concerns | No concerns | No concerns | Some concerns |
| O102 | Yang 2013 | reported adverse drug reactions | No concerns | Some concerns | No concerns | No concerns | Some concerns |
| O103 | Yao 2014 | reported adherence rate; reported adverse drug reactions | No concerns | Some concerns | No concerns | No concerns | Some concerns |
| O104 | Wu 2014 | reported adherence rate | No concerns | Some concerns | No concerns | No concerns | Some concerns |
| O105 | Wen 2014 | reported adverse drug reactions | No concerns | Some concerns | No concerns | No concerns | Some concerns |
| O106 | Tan 2014 | reported adherence rate | No concerns | Some concerns | No concerns | No concerns | Some concerns |
| O107 | Shen 2014 | reported adherence rate; reported adverse drug reactions | No concerns | Some concerns | No concerns | No concerns | Some concerns |
| O108 | Pei 2014 | reported adherence rate | No concerns | Some concerns | No concerns | No concerns | Some concerns |
| O109 | Pei 2014 | reported adherence rate | No concerns | Some concerns | No concerns | No concerns | Some concerns |
| O110 | Jin 2014 | reported adherence rate | No concerns | Some concerns | No concerns | No concerns | Some concerns |
| O111 | Zhou 2015 | reported adherence rate | No concerns | Some concerns | No concerns | No concerns | Some concerns |
| O112 | Zhao 2015 | reported adverse drug reactions | No concerns | Some concerns | No concerns | No concerns | Some concerns |
| O113 | Yang 2015 | reported adherence rate | No concerns | Some concerns | No concerns | No concerns | Some concerns |
| O114 | Luo 2015 | reported adherence rate; reported adverse drug reactions | No concerns | Some concerns | No concerns | No concerns | Some concerns |
| O115 | Liao 2015 | reported adherence rate | No concerns | Some concerns | No concerns | No concerns | Some concerns |
| O116 | Li 2015 | reported adherence rate | No concerns | Some concerns | No concerns | No concerns | Some concerns |
| O117 | Feng 2015 | reported adherence rate | No concerns | Some concerns | No concerns | No concerns | Some concerns |
| O118 | Chen 2015 | reported adherence rate; reported adverse drug reactions | No concerns | Some concerns | No concerns | No concerns | Some concerns |
| O119 | Yu 2016 | reported adverse drug reactions | No concerns | Some concerns | No concerns | No concerns | Some concerns |
| O120 | Wang 2016 | reported adherence rate | No concerns | Some concerns | No concerns | No concerns | Some concerns |
| O121 | Sun 2016 | reported adherence rate; reported adverse drug reactions | No concerns | Some concerns | No concerns | No concerns | Some concerns |
| O122 | Dong 2016 | reported adherence rate | No concerns | Some concerns | No concerns | No concerns | Some concerns |
| O123 | Zeng 2016 | reported adherence rate; reported adverse drug reactions | No concerns | Some concerns | No concerns | No concerns | Some concerns |
| O124 | Wen 2017 | reported adherence rate | No concerns | Some concerns | No concerns | No concerns | Some concerns |
| O125 | Wang 2017 | reported adherence rate | No concerns | Some concerns | No concerns | No concerns | Some concerns |
| O126 | Pan 2017 | reported adherence rate; reported adverse drug reactions | No concerns | Some concerns | No concerns | No concerns | Some concerns |
| O127 | Han 2017 | reported adherence rate | No concerns | Some concerns | No concerns | No concerns | Some concerns |
| O128 | Ni 2018 | reported adverse drug reactions | No concerns | Some concerns | No concerns | No concerns | Some concerns |
| O129 | Tan 2018 | reported adherence rate; reported adverse drug reactions | No concerns | Some concerns | No concerns | No concerns | Some concerns |
| O130 | Lyu 2018 | reported adherence rate; reported adverse drug reactions | No concerns | Some concerns | No concerns | No concerns | Some concerns |
| O131 | Li 2018 | reported adherence rate | No concerns | Some concerns | No concerns | No concerns | Some concerns |
| O132 | Huang 2018 | reported adherence rate | No concerns | Some concerns | No concerns | No concerns | Some concerns |
| O133 | Hou 2018 | reported adherence rate | No concerns | Some concerns | No concerns | No concerns | Some concerns |
| O134 | Guo 2018 | reported adverse drug reactions | No concerns | Some concerns | No concerns | No concerns | Some concerns |
| O135 | Fang 2018 | reported adherence rate | No concerns | Some concerns | No concerns | No concerns | Some concerns |
| O136 | Du 2018 | reported adherence rate; reported adverse drug reactions | No concerns | Some concerns | No concerns | No concerns | Some concerns |
| O137 | Chen 2018 | reported adherence rate | No concerns | Some concerns | No concerns | No concerns | Some concerns |
| O138 | Yao 2019 | reported adverse drug reactions | No concerns | Some concerns | No concerns | No concerns | Some concerns |
| O139 | Wang 2019 | reported adverse drug reactions | No concerns | Some concerns | No concerns | No concerns | Some concerns |
| O140 | Mi 2019 | reported adherence rate | No concerns | Some concerns | No concerns | No concerns | Some concerns |
| O141 | Liu 2019 | reported adherence rate | No concerns | Some concerns | No concerns | No concerns | Some concerns |
| O142 | Liang 2019 | reported adverse drug reactions | No concerns | Some concerns | No concerns | No concerns | Some concerns |
| O143 | Li 2019 | reported adherence rate; reported adverse drug reactions | No concerns | Some concerns | No concerns | No concerns | Some concerns |
| O144 | Du 2019 | reported adverse drug reactions | No concerns | Some concerns | No concerns | No concerns | Some concerns |
| O145 | Chen 2019 | reported adverse drug reactions | No concerns | Some concerns | No concerns | No concerns | Some concerns |
| O146 | Cai 2019 | reported adherence rate; reported adverse drug reactions | No concerns | Some concerns | No concerns | No concerns | Some concerns |
| O147 | Zhi 2018 | reported adherence rate; reported adverse drug reactions | No concerns | Some concerns | No concerns | No concerns | Some concerns |
| O148 | Zhao 2020 | reported adherence rate | No concerns | Some concerns | No concerns | No concerns | Some concerns |
| O149 | Xin 2020 | reported adherence rate | No concerns | Some concerns | No concerns | No concerns | Some concerns |
| O150 | Xin 2020 | MMAS-8 adherence score; reported adverse drug reactions | No concerns | Some concerns | No concerns | No concerns | Some concerns |
| O151 | Shang 2020 | reported adherence rate; reported adverse drug reactions | No concerns | Some concerns | No concerns | No concerns | Some concerns |
| O152 | Ma 2020 | reported adherence rate; reported adverse drug reactions | No concerns | Some concerns | No concerns | No concerns | Some concerns |
| O153 | Li 2020 | reported adherence rate | No concerns | Some concerns | No concerns | No concerns | Some concerns |
| O154 | Li 2020 | reported adherence rate | No concerns | Some concerns | No concerns | No concerns | Some concerns |
| O155 | Li 2020 | reported adherence rate | No concerns | Some concerns | No concerns | No concerns | Some concerns |
| O156 | Huang 2020 | reported adherence rate | No concerns | Some concerns | No concerns | No concerns | Some concerns |
| O157 | Zhu 2019 | reported adherence rate; reported adverse drug reactions | No concerns | Some concerns | No concerns | No concerns | Some concerns |
| O158 | Chen 2020 | reported adverse drug reactions | No concerns | Some concerns | No concerns | No concerns | Some concerns |
| O159 | Zhou 2021 | reported adherence rate; reported adverse drug reactions | No concerns | Some concerns | No concerns | No concerns | Some concerns |
| O160 | Zhao 2021 | reported adherence rate; reported adverse drug reactions | No concerns | Some concerns | No concerns | No concerns | Some concerns |
| O161 | Yu 2021 | reported adherence rate; reported adverse drug reactions | No concerns | Some concerns | No concerns | No concerns | Some concerns |
| O162 | Wang 2021 | reported adherence rate | No concerns | Some concerns | No concerns | No concerns | Some concerns |
| O163 | Tian 2021 | reported adherence rate | No concerns | Some concerns | No concerns | No concerns | Some concerns |
| O164 | Song 2021 | reported adherence rate | No concerns | Some concerns | No concerns | No concerns | Some concerns |
| O165 | Niu 2021 | reported adherence rate | No concerns | Some concerns | No concerns | No concerns | Some concerns |
| O166 | Ma 2021 | reported adherence rate; reported adverse drug reactions | No concerns | Some concerns | No concerns | No concerns | Some concerns |
| O167 | Liang 2021 | reported adherence rate | No concerns | Some concerns | No concerns | No concerns | Some concerns |
| O168 | Li 2021 | MMAS-8 adherence score | No concerns | Some concerns | No concerns | No concerns | Some concerns |
| O169 | Hong 2021 | reported adverse drug reactions | No concerns | Some concerns | No concerns | No concerns | Some concerns |
| O170 | Geng 2021 | reported adherence rate | No concerns | Some concerns | No concerns | No concerns | Some concerns |
| O171 | Duan 2021 | reported adherence rate | No concerns | Some concerns | No concerns | No concerns | Some concerns |
| O172 | Dai 2021 | MMAS-8 adherence score | No concerns | Some concerns | No concerns | No concerns | Some concerns |
| O173 | Chi 2021 | reported adverse drug reactions | No concerns | Some concerns | No concerns | No concerns | Some concerns |
| O174 | Zhao 2022 | reported adherence rate | No concerns | Some concerns | No concerns | No concerns | Some concerns |
| O175 | Zhang 2022 | reported adherence rate | No concerns | Some concerns | No concerns | No concerns | Some concerns |
| O176 | Yin 2022 | reported adherence rate | No concerns | Some concerns | No concerns | No concerns | Some concerns |
| O177 | Wu 2022 | reported adherence rate; reported adverse drug reactions | No concerns | Some concerns | No concerns | No concerns | Some concerns |
| O178 | Wang 2022 | reported adherence rate | No concerns | Some concerns | No concerns | No concerns | Some concerns |
| O179 | Sun 2022 | reported adverse drug reactions | No concerns | Some concerns | No concerns | No concerns | Some concerns |
| O180 | Lin 2014 | reported adherence rate | No concerns | Some concerns | No concerns | No concerns | Some concerns |
| O181 | Rao 2022 | MMAS-8 adherence score | No concerns | Some concerns | No concerns | No concerns | Some concerns |
| O182 | Li 2022 | reported adherence rate | No concerns | Some concerns | No concerns | No concerns | Some concerns |
| O183 | Hu 2022 | reported adherence rate | No concerns | Some concerns | No concerns | No concerns | Some concerns |
| O184 | Guo 2022 | reported adherence rate | No concerns | Some concerns | No concerns | No concerns | Some concerns |
| O185 | Du 2022 | reported adherence rate; reported adverse drug reactions | No concerns | Some concerns | No concerns | No concerns | Some concerns |
| S001 | Jiang 2024 | MMAS-8 adherence score; reported adverse drug reactions | No concerns | No concerns | No concerns | No concerns | No concerns |
| S003 | Yuan 2021 | unplanned readmission during follow-up | No concerns | No concerns | No concerns | No concerns | No concerns |
| S004 | Zheng 2017 | reported adherence rate | No concerns | No concerns | No concerns | No concerns | No concerns |
| S005 | Chu 2022 | reported adherence rate | No concerns | Some concerns | No concerns | No concerns | Some concerns |
| S007 | Liu 2021 | MMAS-8 adherence score | No concerns | No concerns | No concerns | No concerns | No concerns |
| S009 | Gong 2016 | reported adverse drug reactions; unplanned readmission during follow-up | No concerns | Some concerns | No concerns | No concerns | Some concerns |
| S011 | Ji 2022 | reported adverse drug reactions | No concerns | Some concerns | No concerns | No concerns | Some concerns |
| S013 | Su 2017 | MMAS-8 adherence score; reported adverse drug reactions | No concerns | Some concerns | No concerns | No concerns | Some concerns |
| S015 | Yang 2016 | reported adherence rate | No concerns | Some concerns | No concerns | No concerns | Some concerns |
| S016 | Ji 2020 | MMAS-8 adherence score | No concerns | Some concerns | No concerns | No concerns | Some concerns |
| S018 | Zhan 2025 | reported adherence rate | No concerns | Some concerns | No concerns | No concerns | Some concerns |
| S019 | Nie 2020 | MMAS-8 adherence score; reported adherence rate | No concerns | Some concerns | No concerns | No concerns | Some concerns |
| S020 | Ge 2025 | reported adherence rate | No concerns | No concerns | No concerns | No concerns | No concerns |
| S021 | Wang 2019 | reported adverse drug reactions | No concerns | Some concerns | No concerns | No concerns | Some concerns |
| S022 | Shen 2016 | reported adverse drug reactions | No concerns | Some concerns | No concerns | No concerns | Some concerns |
| S023 | Ran 2018 | reported adherence rate; reported adverse drug reactions | No concerns | Some concerns | No concerns | No concerns | Some concerns |
| S024 | Ding 2025 | reported adherence rate | No concerns | No concerns | No concerns | No concerns | No concerns |
| S026 | Xue 2012 | reported adherence rate | No concerns | Some concerns | No concerns | No concerns | Some concerns |
| S027 | Li 2008 | reported adherence rate | No concerns | Some concerns | No concerns | No concerns | Some concerns |
| S029 | Bai 2022 | reported adherence rate; reported adverse drug reactions | No concerns | Some concerns | No concerns | No concerns | Some concerns |
| S031 | Zhang 2022 | MMAS-8 adherence score | No concerns | Some concerns | No concerns | No concerns | Some concerns |
| S034 | Zhou 2021 | reported adherence rate | No concerns | Some concerns | No concerns | No concerns | Some concerns |
| S035 | Guo 2013 | reported adherence rate | No concerns | Some concerns | No concerns | No concerns | Some concerns |
| S037 | Li 2013 | reported adherence rate; reported adverse drug reactions | No concerns | Some concerns | No concerns | No concerns | Some concerns |
| S038 | Li 2025 | reported adverse drug reactions | No concerns | No concerns | No concerns | No concerns | No concerns |
| S040 | Liu 2023 | reported adherence rate | No concerns | No concerns | No concerns | No concerns | No concerns |
| S042 | Zhang 2015 | reported adherence rate | No concerns | Some concerns | No concerns | No concerns | Some concerns |
| S047 | Zhang 2024 | MMAS-8 adherence score | No concerns | Some concerns | No concerns | No concerns | Some concerns |
| S049 | Guo 2017 | reported adherence rate; reported adverse drug reactions | No concerns | Some concerns | No concerns | No concerns | Some concerns |
| S050 | Zhang 2023 | reported adherence rate; reported adverse drug reactions | No concerns | Some concerns | No concerns | No concerns | Some concerns |
| S051 | Xu 2022 | reported adherence rate; reported adverse drug reactions | No concerns | Some concerns | No concerns | No concerns | Some concerns |
| U001 | Gao 2026 | reported adherence rate | No concerns | No concerns | No concerns | No concerns | No concerns |
| U002 | Wang 2023 | MMAS-8 adherence score; reported adverse drug reactions | No concerns | No concerns | No concerns | No concerns | No concerns |
| U005 | Xiao 2023 | unplanned readmission during follow-up | No concerns | No concerns | No concerns | No concerns | No concerns |
| U007 | Zhang 2026 | reported adverse drug reactions | No concerns | Some concerns | No concerns | No concerns | Some concerns |
| U008 | Zhang 2025 | reported adverse drug reactions | No concerns | No concerns | No concerns | No concerns | No concerns |
| U010 | Li 2024 | reported adherence rate | No concerns | No concerns | No concerns | No concerns | No concerns |
| U013 | Miu 2023 | reported adverse drug reactions | No concerns | Some concerns | No concerns | No concerns | Some concerns |
| U014 | Dong 2026 | reported adverse drug reactions | No concerns | Some concerns | No concerns | No concerns | Some concerns |
| U015 | Zhang 2025 | reported adverse drug reactions | No concerns | No concerns | No concerns | No concerns | No concerns |
| U017 | Wang 2023 | reported adverse drug reactions; unplanned readmission during follow-up | No concerns | Some concerns | No concerns | No concerns | Some concerns |
| U018 | Lou 2023 | MMAS-8 adherence score | No concerns | Some concerns | No concerns | No concerns | Some concerns |
| U019 | Lai 2024 | reported adverse drug reactions | No concerns | Some concerns | No concerns | No concerns | Some concerns |
| U020 | Chen 2024 | reported adverse drug reactions | No concerns | Some concerns | No concerns | No concerns | Some concerns |
| U021 | Chen 2025 | MMAS-8 adherence score; reported adverse drug reactions | No concerns | No concerns | No concerns | No concerns | No concerns |
| U022 | Xiong 2025 | reported adverse drug reactions | No concerns | Some concerns | No concerns | No concerns | Some concerns |
| U023 | Wei 2023 | MMAS-8 adherence score; reported adverse drug reactions | No concerns | Some concerns | No concerns | No concerns | Some concerns |
| U024 | Hou 2026 | unplanned readmission during follow-up | No concerns | Some concerns | No concerns | No concerns | Some concerns |
| U025 | Li 2023 | unplanned readmission during follow-up | No concerns | No concerns | No concerns | No concerns | No concerns |
| U026 | Zhang 2025 | reported adherence rate; reported adverse drug reactions | No concerns | Some concerns | No concerns | No concerns | Some concerns |
| U027 | Li 2024 | MMAS-8 adherence score | No concerns | Some concerns | No concerns | No concerns | Some concerns |
| U028 | Sun 2025 | reported adherence rate; reported adverse drug reactions | No concerns | No concerns | No concerns | No concerns | No concerns |
| U029 | Hu 2026 | reported adherence rate | No concerns | Some concerns | No concerns | No concerns | Some concerns |
| U030 | Wang 2023 | reported adherence rate; reported adverse drug reactions; unplanned readmission during follow-up | No concerns | Some concerns | No concerns | No concerns | Some concerns |
| U031 | Wu 2024 | reported adherence rate; reported adverse drug reactions | No concerns | No concerns | No concerns | No concerns | No concerns |
| U033 | Han 2023 | reported adherence rate | No concerns | Some concerns | No concerns | No concerns | Some concerns |
| U036 | Tang 2024 | MMAS-8 adherence score | No concerns | Some concerns | No concerns | No concerns | Some concerns |
| U037 | Yang 2025 | reported adverse drug reactions | No concerns | Some concerns | No concerns | No concerns | Some concerns |

Supplementary Table S15C | Item-level resolution of the 21 INSPECT-SR checks

| **Check** | **Domain** | **Question** | **No** | **Unclear** |
| --- | --- | --- | --- | --- |
| 1.1 | Post-publication notices | Associated retraction? | 238 | 0 |
| 1.2 | Post-publication notices | Expression of concern or other relevant notice? | 238 | 0 |
| 1.3 | Post-publication notices | Related work by the research team raises concern? | 238 | 0 |
| 2.1 | Conduct, governance, and transparency | Concerns relating to ethical approval? | 81 | 157 |
| 2.2 | Conduct, governance, and transparency | Concerns relating to timing or absence of registration? | 27 | 211 |
| 2.3 | Conduct, governance, and transparency | Important inconsistency between publication and registration? | 27 | 211 |
| 2.4 | Conduct, governance, and transparency | Implausible participant recruitment? | 238 | 0 |
| 2.5 | Conduct, governance, and transparency | Methods implausible given reported resources? | 238 | 0 |
| 3.1 | Text and figures | Duplicated or study-incompatible text or tables? | 238 | 0 |
| 3.2 | Text and figures | Manipulation or duplication of figures? | 238 | 0 |
| 4.1 | Results | Unexplained discrepancy with eligibility criteria? | 238 | 0 |
| 4.2 | Results | Group sizes implausible given allocation method? | 238 | 0 |
| 4.3 | Results | Implausible baseline data? | 238 | 0 |
| 4.4 | Results | Discrepancy among figures, tables, and text? | 238 | 0 |
| 4.5 | Results | Implausible loss to follow-up? | 238 | 0 |
| 4.6 | Results | Unexplained inconsistency in participant numbers? | 238 | 0 |
| 4.7 | Results | Implausible outcome data or treatment effects? | 238 | 0 |
| 4.8 | Results | Impossible means or variances for integer data? | 238 | 0 |
| 4.9 | Results | Errors in statistical results? | 238 | 0 |
| 4.10 | Results | Other contradictions implied by the data? | 238 | 0 |
| 4.11 | Results | Inconsistency across publications from the same study? | 238 | 0 |

No item was marked yes. An unclear response means that the available report did not permit verification; it is not an allegation of misconduct. Items 2.2 and 2.3 were necessarily both unresolved when no registration record could be located.

Supplementary Table S16 | Protocol amendments, register versions, and analytical status. Register version numbers and their publication dates are listed in the PROSPERO version history for CRD42023414180.

| **Stage and register version** | **Amendment** | **Rationale and inferential status** |
| --- | --- | --- |
| Initial registration (April 14, 2023; version 1.0) | Narrower set of clinical, medication-use, and resource outcomes; quality of life registered; trials at high risk of bias to be excluded | Prespecified registration |
| During conduct (registered July 18, 2026; version 2.0) | Full database search strategies recorded in the register | The initial record described the strategies only in summary form |
| During conduct, before final data lock (registered August 10, 2026; version 4.0) | Introduced a Chinese-language journal-scope criterion based on the union of three core-index lists | Defined the primary Chinese-language evidence scope; not part of the initial registration and not used as a proxy for trial quality |
| During conduct, before final data lock (registered August 9, 2026; version 3.0) | Expanded structured framework covering 21 outcomes | Map multidimensional evidence; analyses treated as outcome-specific evidence mapping |
| During conduct, before final data lock (registered August 9-10, 2026; versions 3.0 and 5.0) | Added prediction intervals, GRADE, ROB-ME, follow-up and disease-spectrum analyses, and INSPECT-SR | Clarify cross-study consistency, missing evidence, indirectness, and trial trustworthiness |
| During conduct, before final data lock (registered August 10, 2026; version 5.0) | Trials at high risk of bias were retained rather than excluded | Departure from the registered exclusion rule; risk of bias was addressed through outcome-specific low-risk sensitivity analyses, which preserve the full evidence base |
| During conduct (registered August 10, 2026; version 4.0) | Quality of life was not synthesised | Registered in version 1.0; the contributing reports used too many different instruments to pool |
| After final data lock | Post hoc sampled non-core-journal directional validation and secondary leave-one-out checks | Exploratory robustness checks; not part of the prespecified primary synthesis and not an equivalence test |
| Administrative (registered August 11, 2026; version 6.0) | Outcomes itemised individually in the register and the review team completed | No change to eligibility, analysis, or results |

Supplementary Table S17A | Qualitative completeness audit of selected cost-component reports

| **CHEERS-informed item** | **Observed completeness** | **Implication** |
| --- | --- | --- |
| Explicit decision perspective | Not clearly reported | No payer, provider, patient, or societal perspective could be assigned consistently |
| Service-delivery inputs and costs | 0 of 13 trials | Pharmacist time, staffing, overhead, and implementation costs were absent |
| Explicit source price year | Not clearly reported | Publication year was used as the price-year assumption |
| Complete incremental cost | 0 of 13 trials | Hospitalization and medication expenditures were unpaired components |
| Health utility or QALY | 0 of 13 trials | No cost-utility synthesis was possible |
| Joint uncertainty in costs and outcomes | Not reported | Net-benefit and cost-effectiveness uncertainty could not be estimated |

The 13 trials are the unique reports contributing either expenditure component; hospitalisation expenditure was analysed in nine estimates and medication expenditure in 12, with overlap between the two sets. Following the Consolidated Health Economic Evaluation Reporting Standards (CHEERS) 2022, completeness was recorded item by item without a numerical score, because the checklist is not designed to be scored.

Supplementary Table S17B | CPI indices and official source pages used for expenditure standardisation

| **Year** | **Healthcare CPI change (%)** | **Overall CPI change (%)** | **Period** | **Official source** |
| --- | --- | --- | --- | --- |
| 2016 | 3.8 | 2.0 | annual | NBS National Data annual CPI-by-category series |
| 2017 | 6.0 | 1.6 | annual | NBS National Data annual CPI-by-category series |
| 2018 | 4.3 | 2.1 | annual | NBS National Data annual CPI-by-category series |
| 2019 | 2.4 | 2.9 | annual | NBS National Data annual CPI-by-category series |
| 2020 | 1.8 | 2.5 | annual | NBS National Data annual CPI-by-category series |
| 2021 | 0.4 | 0.9 | annual | NBS National Data annual CPI-by-category series |
| 2022 | 0.6 | 2.0 | annual | NBS National Data annual CPI-by-category series |
| 2023 | 1.1 | 0.2 | annual | NBS National Data annual CPI-by-category series |
| 2024 | 1.3 | 0.2 | annual | NBS National Data annual CPI-by-category series |
| 2025 | 0.8 | 0.0 | annual | NBS National Data annual CPI-by-category series |
| 2026 | 2.0 | 1.0 | January-June average | NBS June 2026 CPI release (January-June column) |

Annual 2016-2025 values were obtained from the National Bureau of Statistics National Data annual CPI-by-category series (https://data.stats.gov.cn/english/easyquery.htm?cn=E0104). The 2026 link used the January-June cumulative changes reported in the official June 2026 release (https://www.stats.gov.cn/sj/zxfbhjd/202607/t20260709_1964084.html): healthcare 2.0% and overall CPI 1.0%. Publication year was treated as price year. Source pages were accessed on August 11, 2026.

### Post hoc non-core-journal directional validation

Of 115 full-text reports reviewed, 111 met the population, intervention, comparator, and randomised-study-design criteria. Sixty-eight reported at least one validation outcome and contributed to the validation analyses, whereas 43 were eligible but did not report a validation outcome. Four reports were excluded because both groups received pharmaceutical services. The validation sample originated from 1,800 sampled records, drawn as 100 records from each of six disease strata in each of the three Chinese databases; merging and deduplication yielded 217 unique records before title-and-abstract screening. The validation used the primary effect definitions and random-effects restricted maximum likelihood (REML) model with Hartung-Knapp confidence intervals. MMAS-8 was analysed separately as a continuous exploratory outcome, and mortality was described without pooled validation because only two non-core trials reported it. Follow-up was reported in 33 contributing trials (48.5%) and was not reported in 35 (51.5%). This sampled post-data-lock analysis assessed directional robustness and did not replace the prespecified GRADE assessment or comprehensively review non-core-journal reports. INSPECT-SR was not reapplied.

Supplementary Table S18 | Core versus non-core-journal validation of principal outcomes

| **Outcome** | **Core k (N intervention/control)** | **Core effect (95% CI)** | **Non-core k (N intervention/control)** | **Non-core effect (95% CI)** | **Non-core PI** | **Non-core I²** | **Non-core τ²** | **Combined effect (95% CI)** | **P interaction** | **BH-adjusted q** | **Change vs core** | **Interpretation** |
| --- | --- | --- | --- | --- | --- | --- | --- | --- | --- | --- | --- | --- |
| Reported adherence rate | 163 11529/11355 | RR 1.370 (1.318 to 1.425) | 47 6174/6227 | RR 1.223 (1.186 to 1.261) | 1.089 to 1.374 | 44.5% | 0.0031 | RR 1.327 (1.288 to 1.366) | 0.034 | 0.137 | -3.20% | Direction preserved; magnitude differed |
| Reported ADR | 94 6376/6277 | RR 0.405 (0.367 to 0.448) | 33 4866/4920 | RR 0.349 (0.294 to 0.415) | 0.155 to 0.789 | 57.1% | 0.1526 | RR 0.392 (0.359 to 0.427) | 0.351 | 0.702 | -3.34% | Directional robustness supported |
| Unplanned readmission | 22 1967/1956 | RR 0.524 (0.457 to 0.601) | 5 200/197 | RR 0.407 (0.186 to 0.888) | 0.092 to 1.790 | 40.8% | 0.2058 | RR 0.526 (0.462 to 0.600) | 0.790 | 0.790 | +0.35% | Direction preserved; precision limited |
| MMAS-8 score | 28 1909/1956 | MD 1.211 (0.920 to 1.501) | 7 449/448 | MD 1.030 (0.502 to 1.558) | -0.359 to 2.419 | 96.8% | 0.2757 | MD 1.175 (0.930 to 1.420) | 0.579 | 0.772 | -2.90% | Exploratory; precision limited |

RR, risk ratio; MD, mean difference; PI, 95% prediction interval. Core and combined estimates were reproduced from the locked primary analyses. The interaction P value is from a one-variable core-versus-non-core meta-regression; the four validation outcomes were treated as one exploratory family and q values use the Benjamini-Hochberg procedure. Change versus core is calculated from unrounded pooled estimates. Because the combined random-effects model re-estimates between-study variance and study weights, the combined estimate need not equal arithmetic interpolation between the core and non-core point estimates. Because the non-core sample was stratified for validation rather than sampled proportionally from the excluded pool, combined estimates are sensitivity estimates and should not be interpreted as population-representative estimates of the complete literature.

Supplementary Table S19 | Characteristics of trials included in the non-core-journal validation sample

| **Non-core study label** | **Disease** | **N (I/C)** | **Follow-up** | **Outcome(s)** | **Intervention category** | **Comparator** | **RoB 2 overall** |
| --- | --- | --- | --- | --- | --- | --- | --- |
| Zeng 2012 (Validation No. 2) | Coronary Disease | 631/631 | 6 months | adherence rate | Pharmaceutical care | Usual care or routine medication guidance (as reported) | Some Concerns |
| Liu 2013 (Validation No. 5) | Myocardial infarction | 31/31 | Not reported | readmission | Pharmaceutical care | Usual care or routine medication guidance (as reported) | Some Concerns |
| Chen 2016 (Validation No. 8) | Heart Failure | 70/70 | Not reported | adherence rate | Pharmacist intervention | Usual care or routine medication guidance (as reported) | Some Concerns |
| Li 2016 (Validation No. 10) | Coronary Disease | 55/55 | Not reported | ADR | Pharmacist intervention | Usual care or routine medication guidance (as reported) | Some Concerns |
| Sun 2016 (Validation No. 11) | Dyslipidemia | 50/50 | 6 months | adherence rate | Pharmacist intervention | Usual care or routine medication guidance (as reported) | Some Concerns |
| Cao 2017 (Validation No. 12) | Coronary Disease | 50/50 | 3 months | adherence rate, ADR | Pharmacist intervention | Usual care or routine medication guidance (as reported) | Some Concerns |
| Chen 2017 (Validation No. 13) | Diabetes Mellitus | 39/39 | Not reported | adherence rate | Pharmacist intervention | Usual care or routine medication guidance (as reported) | Some Concerns |
| Han 2017 (Validation No. 14) | Diabetes Mellitus | 50/50 | Not reported | adherence rate | Pharmacist intervention | Usual care or routine medication guidance (as reported) | Some Concerns |
| Liao 2017 (Validation No. 15) | Diabetes Mellitus | 96/96 | 3 months | adherence rate | Pharmacist intervention | Usual care or routine medication guidance (as reported) | High |
| Liu 2017 (Validation No. 16) | Diabetes Mellitus | 35/35 | Not reported | adherence rate, ADR | Pharmacist intervention | Usual care or routine medication guidance (as reported) | Low |
| Chen 2017 (Validation No. 17) | Heart Failure | 42/42 | 6 months | readmission, mortality | Pharmacist intervention | Usual care or routine medication guidance (as reported) | Some Concerns |
| Li 2018 (Validation No. 20) | Heart Failure | 38/38 | 12 months | readmission, mortality | Pharmacist intervention | Usual care or routine medication guidance (as reported) | Some Concerns |
| Wu 2018 (Validation No. 23) | Mixed cardiometabolic | 30/30 | 12 months | adherence rate, ADR | Pharmacist intervention | Usual care or routine medication guidance (as reported) | High |
| Cai 2019 (Validation No. 28) | Diabetes Mellitus | 34/34 | Not reported | adherence rate, ADR | Pharmacist intervention | Usual care or routine medication guidance (as reported) | Some Concerns |
| Chen 2019 (Validation No. 30) | Mixed cardiometabolic | 45/45 | Not reported | adherence rate | Pharmacist intervention | Usual care or routine medication guidance (as reported) | Some Concerns |
| Deng 2019 (Validation No. 32) | Mixed cardiometabolic | 100/100 | Not reported | ADR | Pharmacist intervention | Usual care or routine medication guidance (as reported) | Some Concerns |
| Dou 2019 (Validation No. 33) | Diabetes Mellitus | 50/50 | 12 months | adherence rate | Pharmacist intervention | Usual care or routine medication guidance (as reported) | Some Concerns |
| Huang 2019 (Validation No. 34) | Diabetes Mellitus | 37/37 | 3 months | adherence rate | Medication therapy management | Usual care or routine medication guidance (as reported) | Low |
| Liao 2019 (Validation No. 36) | Mixed cardiometabolic | 86/87 | 6 months | adherence rate | Pharmacist intervention | Usual care or routine medication guidance (as reported) | Some Concerns |
| Mei 2019 (Validation No. 37) | Mixed cardiometabolic | 25/25 | Not reported | ADR | Pharmacist intervention | Usual care or routine medication guidance (as reported) | Some Concerns |
| Rong 2019 (Validation No. 39) | Mixed cardiometabolic | 50/50 | 6 months | adherence rate | Pharmacist intervention | Usual care or routine medication guidance (as reported) | Some Concerns |
| Wang 2019 (Validation No. 42) | Hypertension | 90/90 | Not reported | ADR | Pharmacist intervention | Usual care or routine medication guidance (as reported) | High |
| Zhang 2019 (Validation No. 47) | Diabetes Mellitus | 120/120 | Not reported | adherence rate | Pharmacist intervention | Usual care or routine medication guidance (as reported) | Some Concerns |
| Zhu 2019 (Validation No. 49) | Diabetes Mellitus | 50/50 | 12 months | adherence rate | Pharmacist intervention | Usual care or routine medication guidance (as reported) | Some Concerns |
| Jiang 2020 (Validation No. 52) | Diabetes Mellitus | 35/35 | Not reported | adherence rate | Pharmacist intervention | Usual care or routine medication guidance (as reported) | Low |
| Jiang 2020 (Validation No. 53) | Myocardial infarction | 128/128 | 1 month | ADR, MMAS-8 | Pharmacist intervention | Usual care or routine medication guidance (as reported) | Low |
| Shao 2020 (Validation No. 55) | Diabetes Mellitus | 50/50 | Not reported | adherence rate | Pharmacist intervention | Usual care or routine medication guidance (as reported) | Low |
| Yang 2020 (Validation No. 59) | Mixed cardiometabolic | 20/20 | Not reported | adherence rate | Pharmacist intervention | Usual care or routine medication guidance (as reported) | High |
| Zhang 2020 (Validation No. 60) | Hypertension | 41/41 | Not reported | adherence rate | Pharmacist intervention | Usual care or routine medication guidance (as reported) | Some Concerns |
| Zeng 2021 (Validation No. 64) | Diabetes Mellitus | 58/58 | 3 months | adherence rate | Pharmacist intervention | Usual care or routine medication guidance (as reported) | Some Concerns |
| Chu 2021 (Validation No. 67) | Diabetes Mellitus | 41/41 | 3 months | adherence rate | Pharmacist intervention | Usual care or routine medication guidance (as reported) | Low |
| Han 2021 (Validation No. 71) | Diabetes Mellitus | 80/80 | Not reported | adherence rate | Pharmaceutical care | Usual care or routine medication guidance (as reported) | Some Concerns |
| Hua 2021 (Validation No. 73) | Hypertension | 50/50 | Not reported | ADR | Pharmacist intervention | Usual care or routine medication guidance (as reported) | Low |
| Kong 2021 (Validation No. 74) | Diabetes Mellitus | 50/50 | Not reported | adherence rate, ADR | Pharmacist intervention | Usual care or routine medication guidance (as reported) | Some Concerns |
| Lai 2021 (Validation No. 75) | Mixed cardiometabolic | 50/50 | 1 month | adherence rate, ADR, MMAS-8 | Pharmaceutical care | Usual care or routine medication guidance (as reported) | Low |
| Li 2021 (Validation No. 76) | Hypertension | 50/50 | 6 months | adherence rate | Pharmacist intervention | Usual care or routine medication guidance (as reported) | Low |
| Li 2021 (Validation No. 77) | Hypertension | 52/52 | Not reported | adherence rate, ADR | Pharmacist intervention | Usual care or routine medication guidance (as reported) | Low |
| Li 2021 (Validation No. 78) | Diabetes Mellitus | 43/43 | Not reported | ADR | Pharmacist intervention | Usual care or routine medication guidance (as reported) | Some Concerns |
| Liu 2021 (Validation No. 80) | Diabetes Mellitus | 40/40 | Not reported | adherence rate | Pharmacist intervention | Usual care or routine medication guidance (as reported) | Some Concerns |
| Liu 2021 (Validation No. 81) | Hypertension | 80/80 | 6 months | adherence rate, ADR | Pharmaceutical care | Usual care or routine medication guidance (as reported) | High |
| Liu 2021 (Validation No. 83) | Mixed cardiometabolic | 32/32 | Not reported | ADR | Pharmacist intervention | Usual care or routine medication guidance (as reported) | Some Concerns |
| Luo 2021 (Validation No. 84) | Diabetes Mellitus | 51/51 | Not reported | adherence rate, ADR | Pharmacist intervention | Usual care or routine medication guidance (as reported) | Some Concerns |
| Sun 2021 (Validation No. 89) | Hypertension | 50/50 | 3 months | adherence rate | Pharmacist intervention | Usual care or routine medication guidance (as reported) | Some Concerns |
| Wang 2021 (Validation No. 90) | Mixed cardiometabolic | 60/60 | Not reported | adherence rate, ADR | Pharmacist intervention | Usual care or routine medication guidance (as reported) | Low |
| Yin 2021 (Validation No. 94) | Diabetes Mellitus | 88/88 | Not reported | ADR | Pharmacist intervention | Usual care or routine medication guidance (as reported) | Low |
| Guo 2022 (Validation No. 101) | Hypertension | 56/58 | 6 months | ADR, MMAS-8 | Medication therapy management | Usual care or routine medication guidance (as reported) | Low |
| Hu 2022 (Validation No. 103) | Dyslipidemia | 56/56 | 1 month | ADR | Pharmacist intervention | Usual care or routine medication guidance (as reported) | Low |
| Shao 2022 (Validation No. 106) | Coronary Disease | 63/63 | 1 month | ADR | Pharmaceutical care | Usual care or routine medication guidance (as reported) | Some Concerns |
| Ni 2023 (Validation No. 108) | Diabetes Mellitus | 35/35 | Not reported | adherence rate, ADR | Pharmaceutical care | Usual care or routine medication guidance (as reported) | Some Concerns |
| Xiao 2023 (Validation No. 109) | Diabetes Mellitus | 72/72 | Not reported | adherence rate, ADR | Pharmacist intervention | Usual care or routine medication guidance (as reported) | Low |
| Yin 2023 (Validation No. 110) | Hypertension | 3024/3076 | 6 months | adherence rate, ADR | Medication therapy management | Usual care or routine medication guidance (as reported) | Low |
| Zhai 2024 (Validation No. 118) | Hypertension | 156/156 | Not reported | adherence rate | Pharmacist intervention | Usual care or routine medication guidance (as reported) | Low |
| Sun 2024 (Validation No. 119) | Heart Failure | 43/40 | 6 months | readmission, MMAS-8 | Pharmacist intervention | Usual care or routine medication guidance (as reported) | Some Concerns |
| Tao 2024 (Validation No. 120) | Myocardial infarction | 46/46 | Not reported | adherence rate, ADR, readmission | Pharmacist intervention | Usual care or routine medication guidance (as reported) | Low |
| Zhang 2024 (Validation No. 124) | Hypertension | 30/30 | 1 month | adherence rate | Pharmacist intervention | Usual care or routine medication guidance (as reported) | Low |
| Zhu 2024 (Validation No. 126) | Hypertension | 50/50 | Not reported | adherence rate, ADR | Pharmacist intervention | Usual care or routine medication guidance (as reported) | Low |
| Cao 2025 (Validation No. 127) | Mixed cardiometabolic | 42/42 | 6 months | MMAS-8 | Medication therapy management | Usual care or routine medication guidance (as reported) | Some Concerns |
| Liu 2025 (Validation No. 131) | Diabetes Mellitus | 47/47 | Not reported | adherence rate | Pharmacist intervention | Usual care or routine medication guidance (as reported) | Some Concerns |
| Ou 2025 (Validation No. 132) | Mixed cardiometabolic | 150/150 | 6 months | adherence rate | Pharmacist intervention | Usual care or routine medication guidance (as reported) | Some Concerns |
| Shu 2025 (Validation No. 133) | Hypertension | 80/80 | 6 months | adherence rate | Pharmacist intervention | Usual care or routine medication guidance (as reported) | Low |
| Sun 2025 (Validation No. 134) | Mixed cardiometabolic | 40/40 | Not reported | adherence rate | Pharmacist intervention | Usual care or routine medication guidance (as reported) | Some Concerns |
| Yin 2025 (Validation No. 135) | Dyslipidemia | 50/50 | Not reported | ADR | Pharmacist intervention | Usual care or routine medication guidance (as reported) | Low |
| Yuan 2025 (Validation No. 137) | Coronary Disease | 30/30 | 3 months | ADR, MMAS-8 | Medication therapy management | Usual care or routine medication guidance (as reported) | Low |
| Fu 2026 (Validation No. 139) | Hypertension | 100/100 | 6 months | MMAS-8 | Pharmacist intervention | Usual care or routine medication guidance (as reported) | Some Concerns |
| Hua 2026 (Validation No. 140) | Hypertension | 41/41 | Not reported | adherence rate, ADR | Pharmacist intervention | Usual care or routine medication guidance (as reported) | Some Concerns |
| Wu 2026 (Validation No. 142) | Hypertension | 200/200 | 12 months | ADR | Pharmacist intervention | Usual care or routine medication guidance (as reported) | Low |
| Xue 2026 (Validation No. 143) | Diabetes Mellitus | 50/50 | 3 months | adherence rate, ADR | Medication therapy management | Usual care or routine medication guidance (as reported) | Low |
| Zhao 2026 (Validation No. 144) | Dyslipidemia | 40/40 | 6 months | adherence rate, ADR | Pharmacist intervention | Usual care or routine medication guidance (as reported) | Low |

The table lists the 68 unique non-core-journal trials that reported at least one validation outcome and contributed to the validation analyses. Forty-three additional reports met the underlying study-level criteria but did not report a validation outcome and therefore contributed no validation effect estimate; four reports were excluded because both groups received pharmaceutical services. N is the randomised sample size in the intervention/control groups. Follow-up was reported in 33 contributing trials (48.5%) and not reported in 35 (51.5%). Comparator descriptions are harmonised to a compact label in this table; the source reports and extraction workbook retain the original wording. INSPECT-SR was applied only to the 238 core-outcome trials in the primary evidence base, not to these 68 validation trials.

Supplementary Table S20 | Leave-one-out sensitivities for non-core validation

| **Sensitivity** | **k** | **N (I/C)** | **RR (95% CI)** | **95% PI** | **I²** | **τ²** | **Interaction P (raw)** | **BH q (primary family)** |
| --- | --- | --- | --- | --- | --- | --- | --- | --- |
| Yin 2023 excluded: adherence | 46 | 3150/3151 | RR 1.228 (1.192 to 1.266) | 1.107 to 1.363 | 32.5% | 0.0025 | 0.049 | 0.197 |
| Yin 2023 excluded: ADR | 32 | 1842/1844 | RR 0.335 (0.289 to 0.387) | 0.194 to 0.578 | 21.7% | 0.0666 | 0.044 | Not in primary family |

Excluding Yin (2023), the non-core adherence estimate remained weaker than the core estimate, and the ADR estimate remained directionally protective (RR 0.335, 95% CI 0.289-0.387). Raw interaction P values are shown for these secondary leave-one-out checks. The ADR check was not added to the primary four-outcome BH family; therefore no BH q value is reported for that row. Neither leave-one-out result establishes that the remaining contrast is journal-related.

Supplementary Table S21A | Model, leave-one-out, and high-risk-excluded sensitivity analyses

| **Outcome** | **Measure** | **k** | **Primary REML-HK (95% CI)** | **DL/PM/HE point-estimate range** | **Leave-one-out point-estimate range** | **High-risk-excluded REML-HK (95% CI)** |
| --- | --- | --- | --- | --- | --- | --- |
| 2-h postprandial glucose | MD | 105 | -1.79 (-2.02 to -1.56) | -1.79 to -1.79 | -1.83 to -1.74 | k=102; -1.76 (-1.99 to -1.53) |
| MMAS-8 | MD | 28 | 1.21 (0.92 to 1.50) | 1.21 to 1.21 | 1.14 to 1.25 | k=27; 1.23 (0.94 to 1.53) |
| Reported adherence rate | RR | 163 | 1.37 (1.32 to 1.43) | 1.35 to 1.43 | 1.36 to 1.37 | k=150; 1.37 (1.31 to 1.43) |
| Reported adverse drug reactions | RR | 94 | 0.41 (0.37 to 0.45) | 0.41 to 0.46 | 0.40 to 0.41 | k=91; 0.41 (0.37 to 0.45) |
| Glycaemic effective rate | OR | 31 | 4.54 (3.43 to 6.00) | 4.41 to 4.57 | 4.07 to 4.74 | k=31; 4.54 (3.43 to 6.00) |
| Blood-pressure effective rate | OR | 56 | 3.46 (2.89 to 4.14) | 3.46 to 3.46 | 3.35 to 3.57 | k=51; 3.50 (2.89 to 4.24) |
| Diastolic blood pressure | MD | 87 | -6.62 (-7.47 to -5.76) | -6.65 to -6.62 | -6.76 to -6.53 | k=83; -6.77 (-7.65 to -5.89) |
| Overall effective rate | OR | 9 | 2.63 (1.83 to 3.77) | 2.63 to 2.99 | 2.51 to 2.76 | k=8; 2.76 (1.77 to 4.30) |
| Fasting plasma glucose | MD | 137 | -1.24 (-1.38 to -1.10) | -1.24 to -1.24 | -1.26 to -1.22 | k=131; -1.22 (-1.36 to -1.08) |
| Glycated haemoglobin | MD | 110 | -1.09 (-1.24 to -0.93) | -1.09 to -1.09 | -1.11 to -1.05 | k=104; -1.06 (-1.21 to -0.90) |
| HDL cholesterol | MD | 27 | 0.13 (0.01 to 0.25) | 0.10 to 0.13 | 0.09 to 0.15 | k=27; 0.13 (0.01 to 0.25) |
| LDL cholesterol | MD | 35 | -0.64 (-0.89 to -0.40) | -0.65 to -0.61 | -0.66 to -0.56 | k=35; -0.64 (-0.89 to -0.40) |
| Length of stay | MD | 13 | -1.77 (-2.96 to -0.58) | -1.81 to -1.75 | -1.98 to -1.35 | k=13; -1.77 (-2.96 to -0.58) |
| Mortality | RR | 9 | 0.53 (0.39 to 0.72) | 0.53 to 0.53 | 0.50 to 0.57 | k=9; 0.53 (0.39 to 0.72) |
| Unplanned readmission | RR | 22 | 0.52 (0.46 to 0.60) | 0.48 to 0.52 | 0.50 to 0.55 | k=22; 0.52 (0.46 to 0.60) |
| Satisfaction | OR | 12 | 5.39 (2.91 to 9.99) | 5.34 to 5.40 | 4.38 to 6.23 | k=12; 5.39 (2.91 to 9.99) |
| Systolic blood pressure | MD | 91 | -8.83 (-10.08 to -7.58) | -8.83 to -8.81 | -9.05 to -8.58 | k=87; -8.76 (-10.06 to -7.46) |
| Total cholesterol | MD | 28 | -0.66 (-0.86 to -0.45) | -0.66 to -0.65 | -0.68 to -0.61 | k=28; -0.66 (-0.86 to -0.45) |
| Triglycerides | MD | 29 | -0.44 (-0.65 to -0.23) | -0.44 to -0.43 | -0.47 to -0.36 | k=29; -0.44 (-0.65 to -0.23) |

REML-HK denotes restricted maximum likelihood with Hartung-Knapp inference. Alternative models used DerSimonian-Laird (DL), Paule-Mandel (PM), and Hunter-Schmidt (HE) estimators with Hartung-Knapp inference. High-risk exclusion used the descriptive trial-level evidence-map classification and is distinct from the outcome-specific low-risk analyses in Supplementary Table S6A. None of the 19 non-cost outcomes changed direction. Expenditure outcomes are reported separately with healthcare-CPI and overall-CPI specifications in the main results and Figure 5.

Supplementary Table S21B | Outcome-specific small-study-effect tests

| **Outcome** | **Measure** | **k** | **Prespecified primary test** | **Primary P** | **Supplementary test** | **Supplementary P** | **Note** |
| --- | --- | --- | --- | --- | --- | --- | --- |
| 2-h postprandial glucose | MD | 105 | Egger regression | 0.879 | Not prespecified | Not estimated | Primary test only |
| MMAS-8 | MD | 28 | Egger regression | 0.355 | Not prespecified | Not estimated | Primary test only |
| Reported adherence rate | RR | 163 | Peters regression | 0.203 | Egger regression | <0.001 | Egger is supplementary |
| Reported adverse drug reactions | RR | 94 | Peters regression | 0.137 | Egger regression | <0.001 | Egger is supplementary |
| Glycaemic effective rate | OR | 31 | Peters regression | 0.590 | Egger regression | 0.078 | Egger is supplementary |
| Blood-pressure effective rate | OR | 56 | Peters regression | 0.496 | Egger regression | 0.387 | Egger is supplementary |
| Diastolic blood pressure | MD | 87 | Egger regression | 0.629 | Not prespecified | Not estimated | Primary test only |
| Overall effective rate | OR | 9 | Not performed | Not estimated | Not performed | Not estimated | Not performed: fewer than 10 estimates |
| Fasting plasma glucose | MD | 137 | Egger regression | 0.970 | Not prespecified | Not estimated | Primary test only |
| Glycated haemoglobin | MD | 110 | Egger regression | <0.001 | Not prespecified | Not estimated | Primary test only |
| HDL cholesterol | MD | 27 | Egger regression | 0.505 | Not prespecified | Not estimated | Primary test only |
| LDL cholesterol | MD | 35 | Egger regression | 0.275 | Not prespecified | Not estimated | Primary test only |
| Length of stay | MD | 13 | Egger regression | 0.011 | Not prespecified | Not estimated | Primary test only |
| Mortality | RR | 9 | Not performed | Not estimated | Not performed | Not estimated | Not performed: fewer than 10 estimates |
| Unplanned readmission | RR | 22 | Peters regression | 0.613 | Egger regression | 0.021 | Egger is supplementary |
| Satisfaction | OR | 12 | Peters regression | 0.025 | Egger regression | 0.004 | Egger is supplementary |
| Systolic blood pressure | MD | 91 | Egger regression | <0.001 | Not prespecified | Not estimated | Primary test only |
| Total cholesterol | MD | 28 | Egger regression | 0.347 | Not prespecified | Not estimated | Primary test only |
| Triglycerides | MD | 29 | Egger regression | 0.062 | Not prespecified | Not estimated | Primary test only |

Peters regression was the prespecified primary assessment for binary outcomes; Egger regression is shown as a supplementary, method-dependent sensitivity. Egger regression was the primary assessment for continuous outcomes. Tests were not performed when fewer than ten estimates were available. Funnel-plot asymmetry and regression-test results are signals of small-study effects and are not direct proof of publication bias.

### Supplementary figures

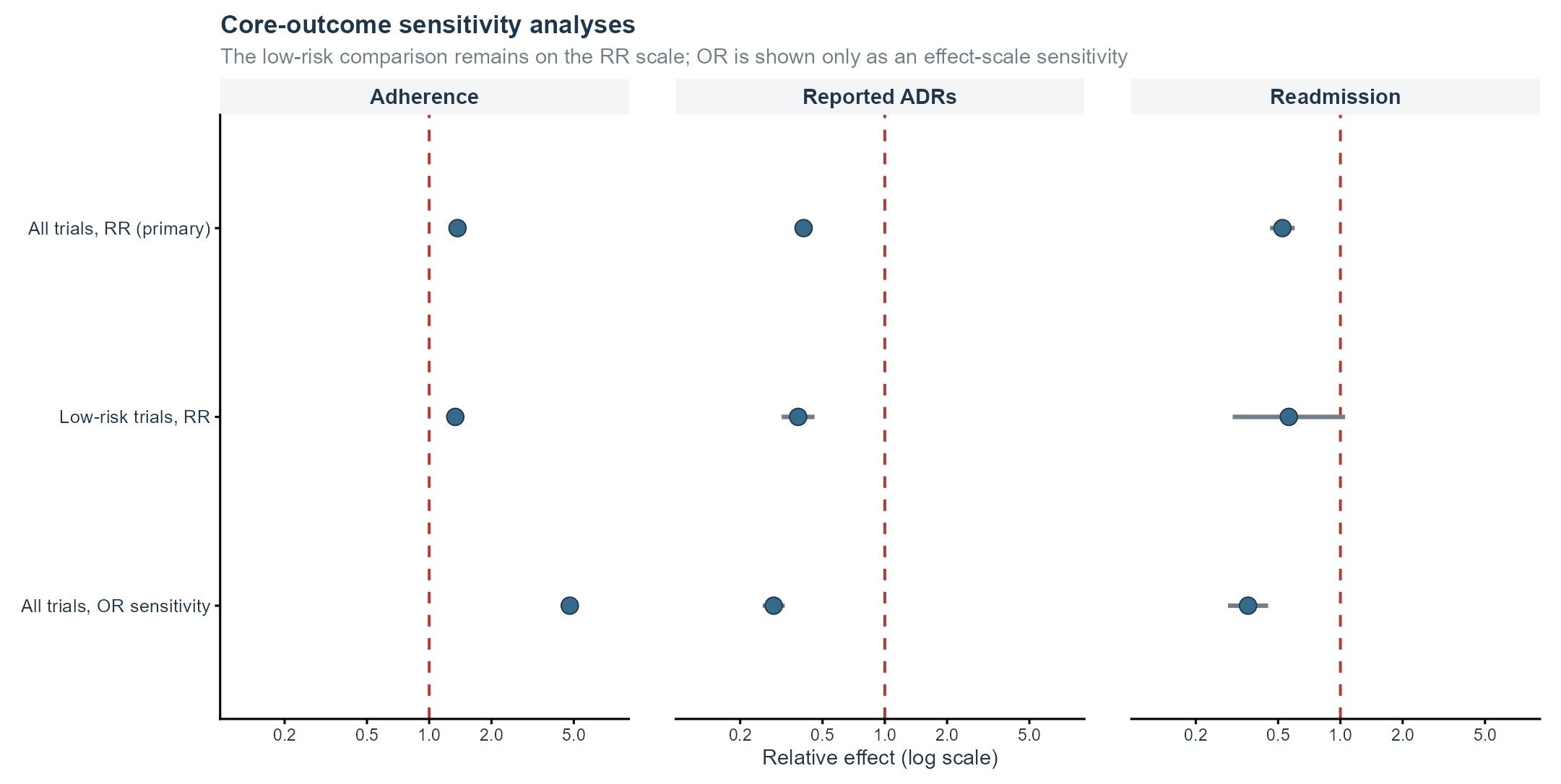

Supplementary Figure S1 | Core-outcome sensitivity analyses. Each panel shows the primary all-trial RR, the outcome-specific low-risk RR, and the all-trial OR effect-scale sensitivity. Keeping the low-risk comparison on the RR scale separates risk-of-bias restriction from effect-scale sensitivity.

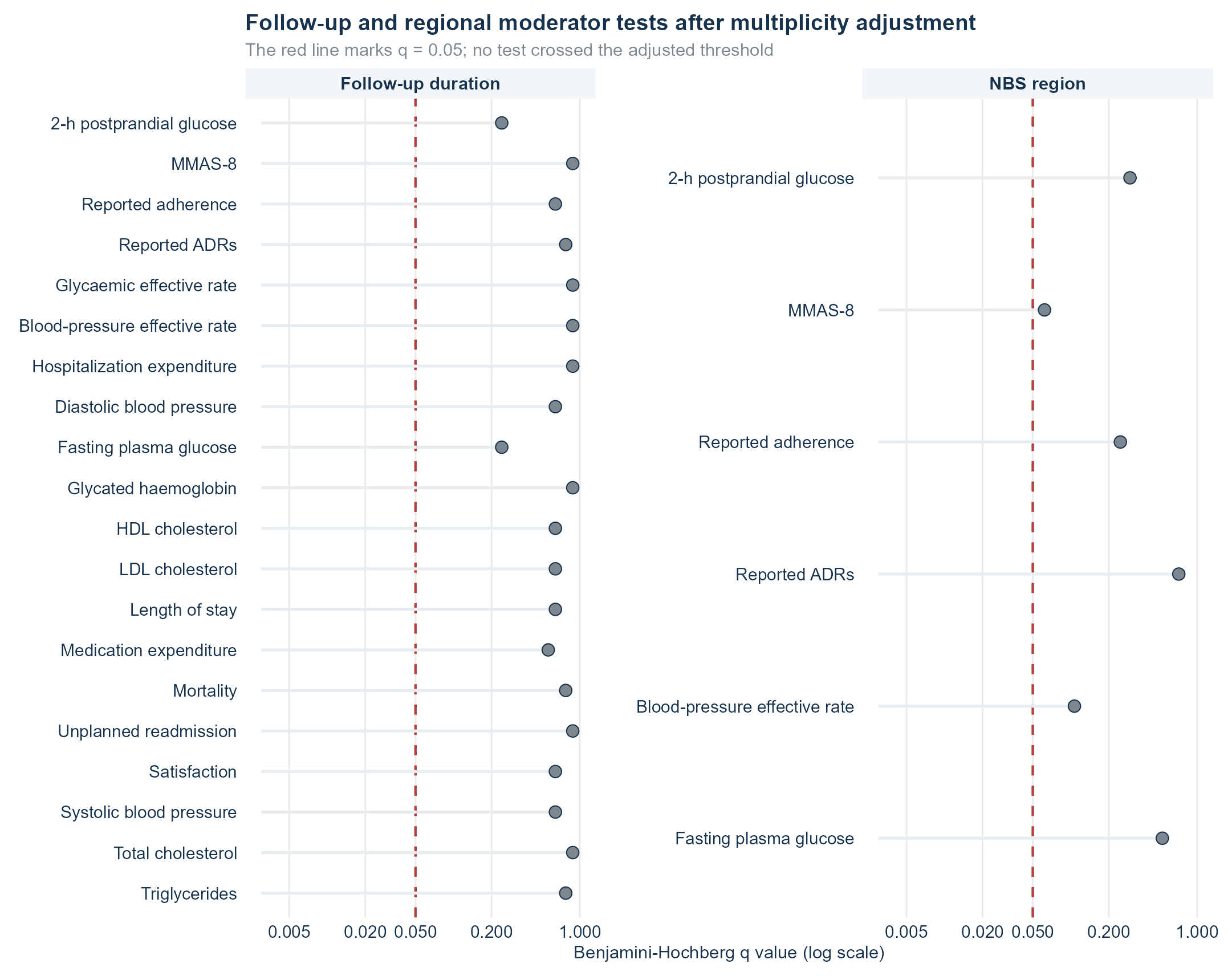

Supplementary Figure S2 | Follow-up and regional moderator tests after false-discovery-rate correction. The prespecified continuous readmission follow-up moderator is reported separately using its unadjusted P value; the FDR display is provided for the broader exploratory moderator family.

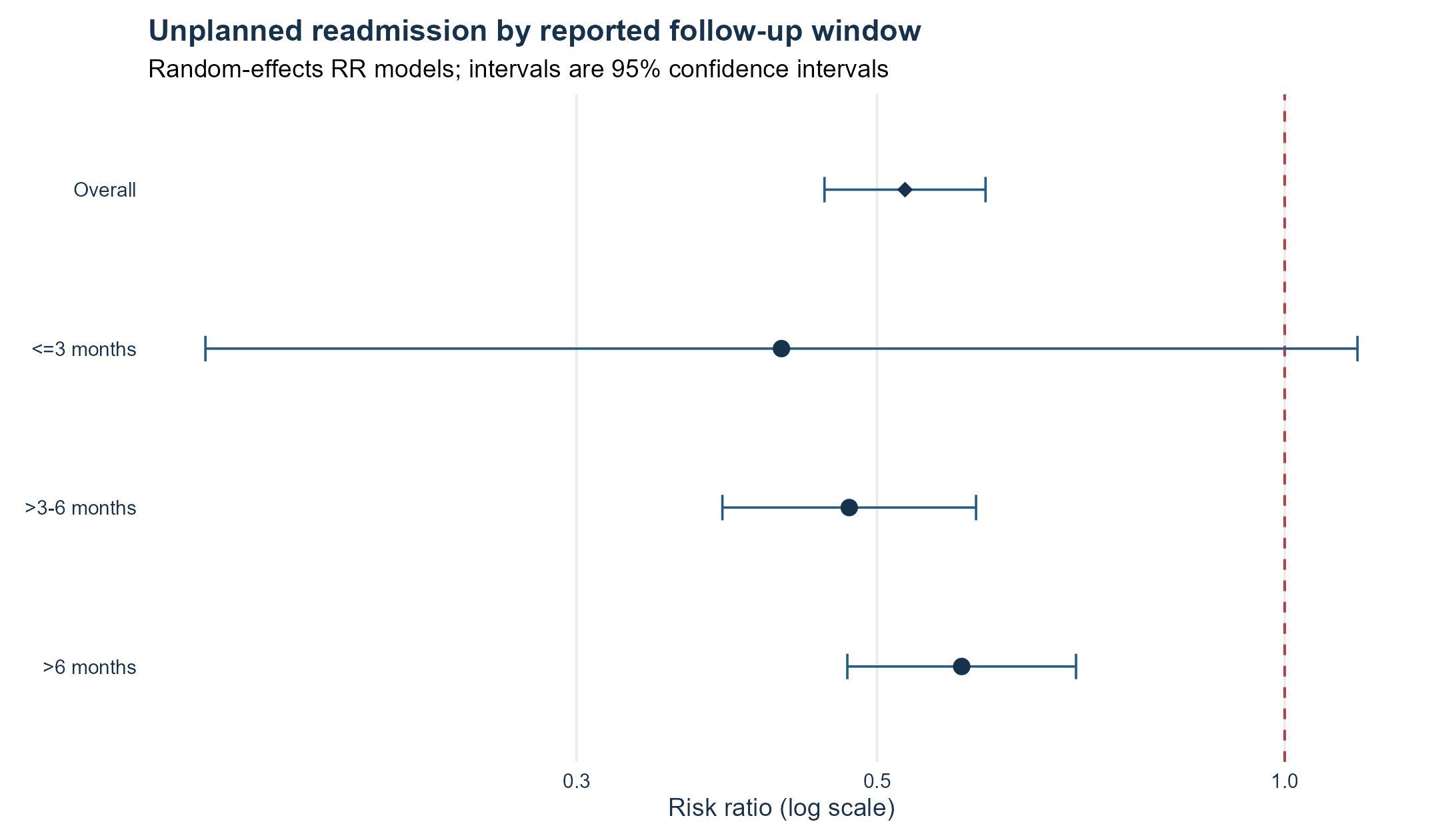

Supplementary Figure S3 | Unplanned readmission by reported follow-up window. Points are random-effects RRs and bars are 95% confidence intervals. One estimate lacked follow-up duration and is included only in the overall model.

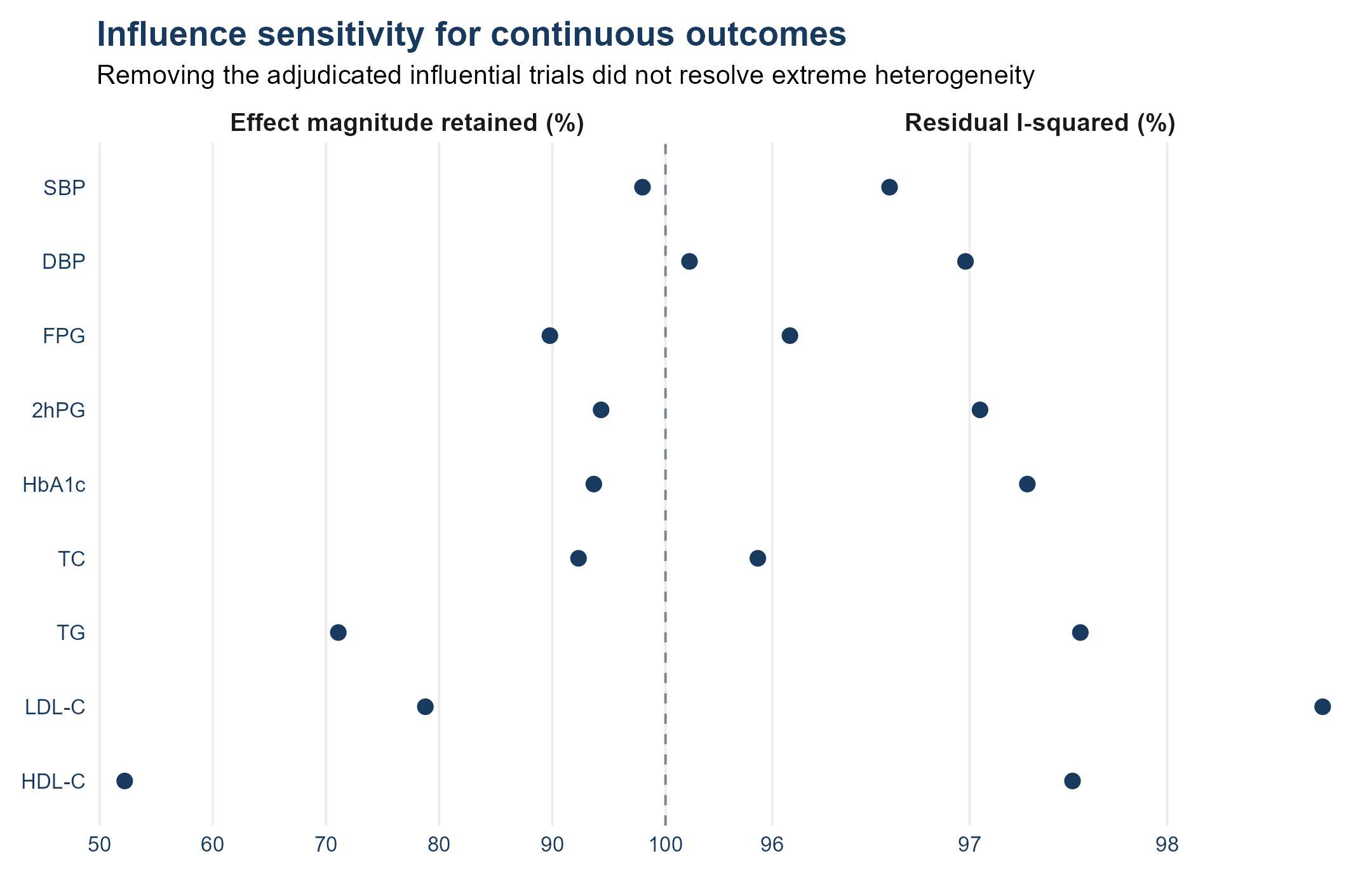

Supplementary Figure S4 | Influence sensitivity for continuous outcomes. The left panel shows the proportion of the primary pooled effect magnitude retained after removal of adjudicated influential trials. The right panel shows residual I². Influence flags were not treated as data errors.

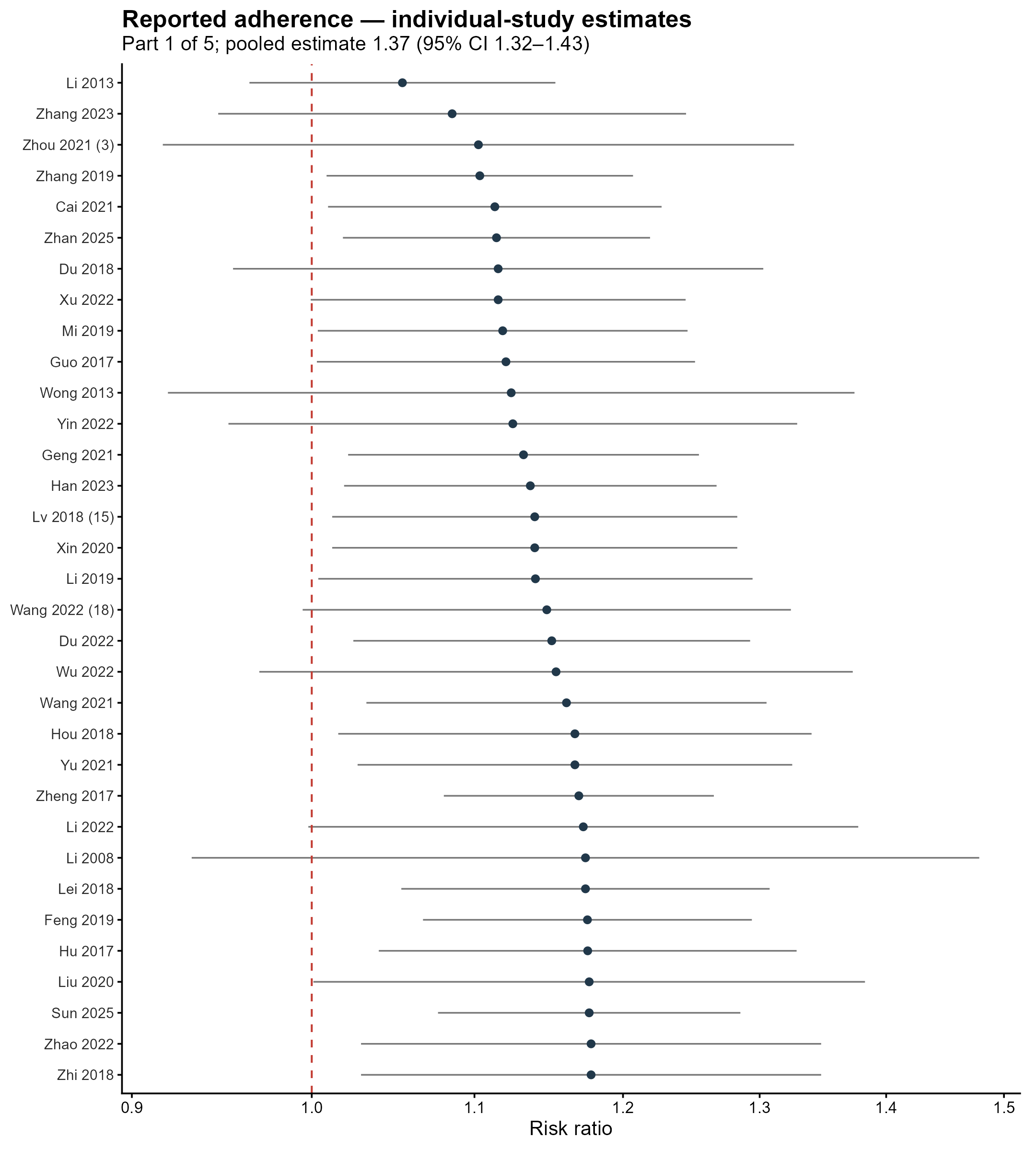

Supplementary Figure S5 | Adherence-Rate individual-study forest plot, part 1 of 5. The dashed red line is the null; pooled estimates are stated in the subtitle.

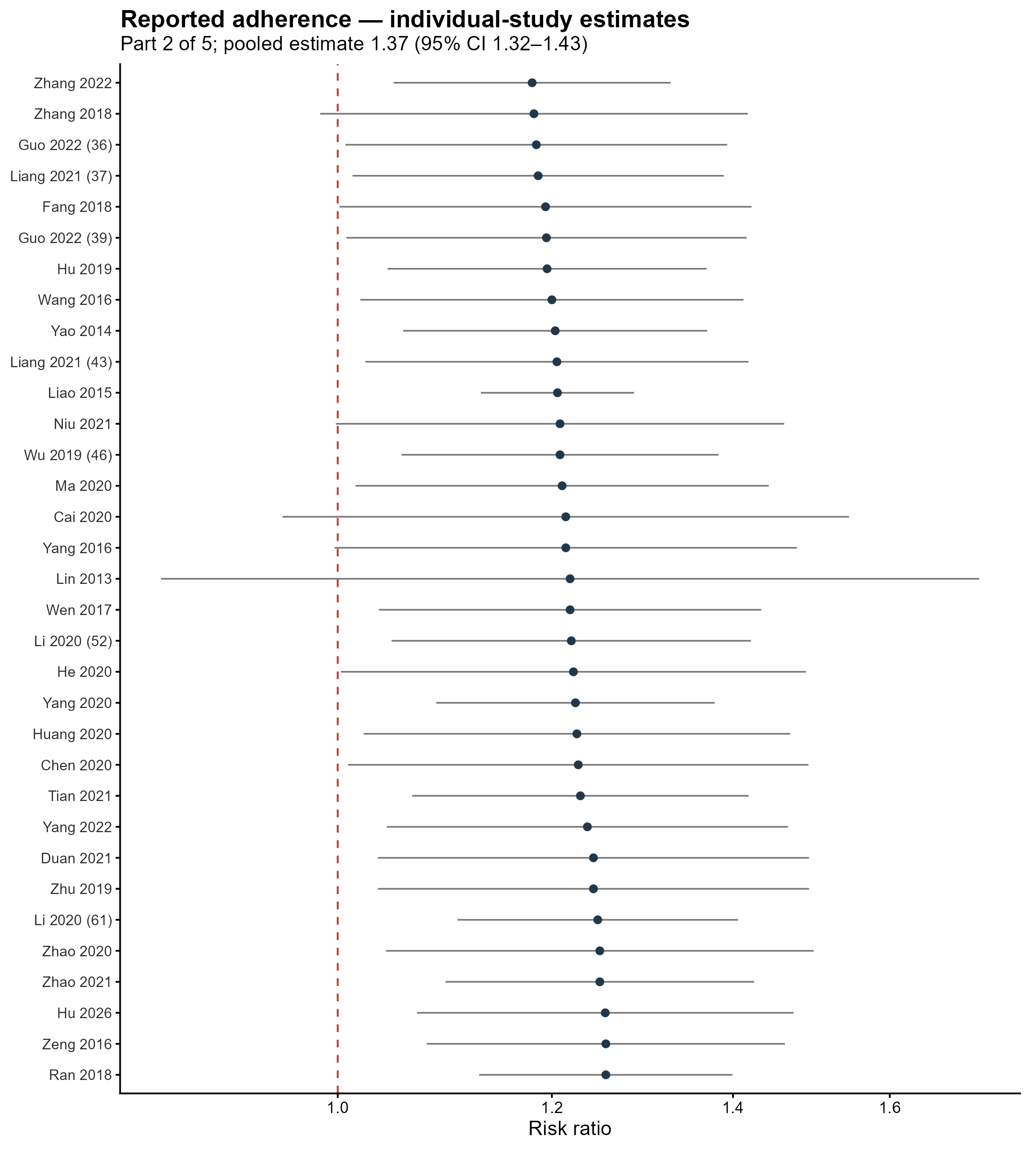

Supplementary Figure S6 | Adherence-Rate individual-study forest plot, part 2 of 5. The dashed red line is the null; pooled estimates are stated in the subtitle.

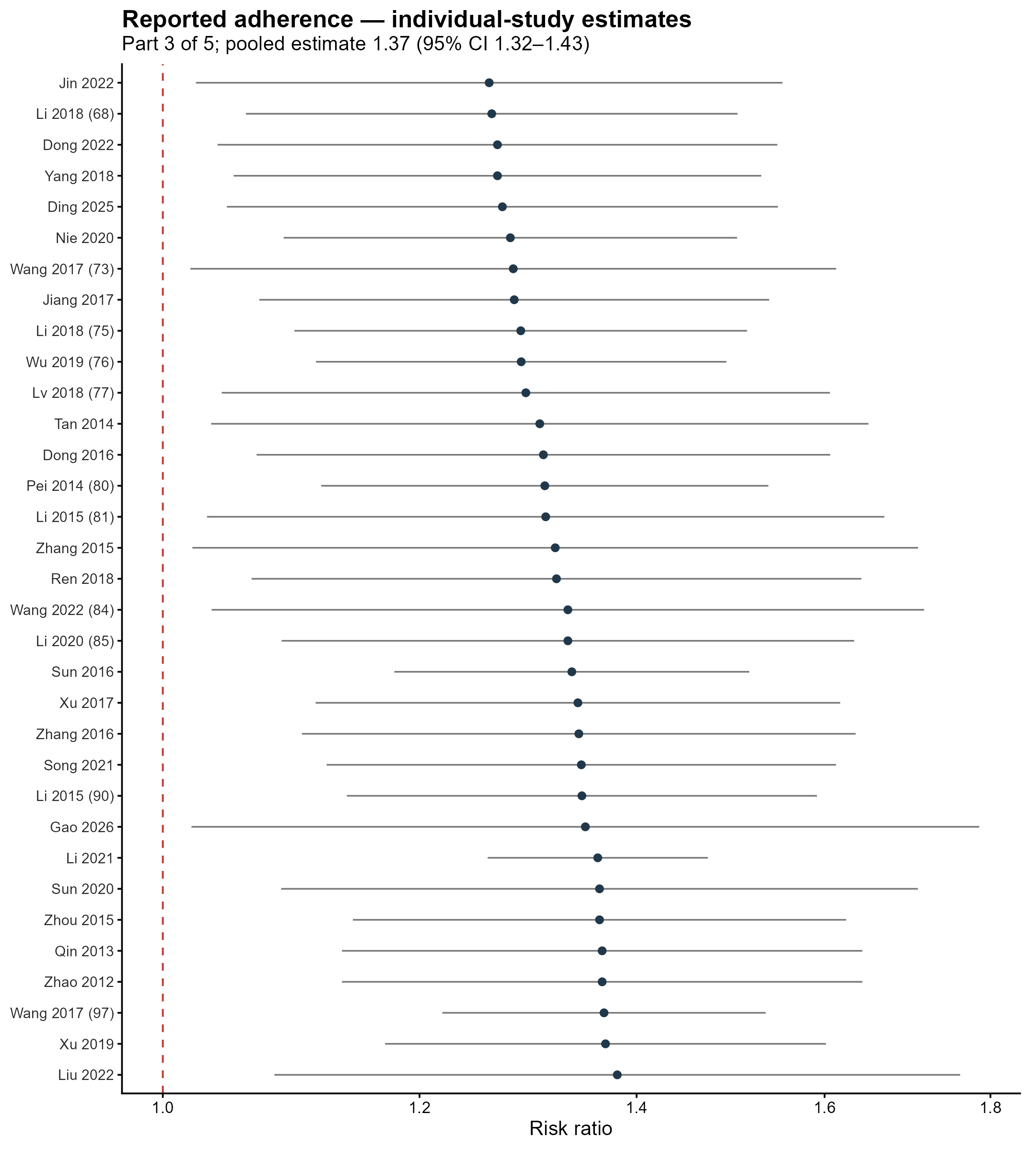

Supplementary Figure S7 | Adherence-Rate individual-study forest plot, part 3 of 5. The dashed red line is the null; pooled estimates are stated in the subtitle.

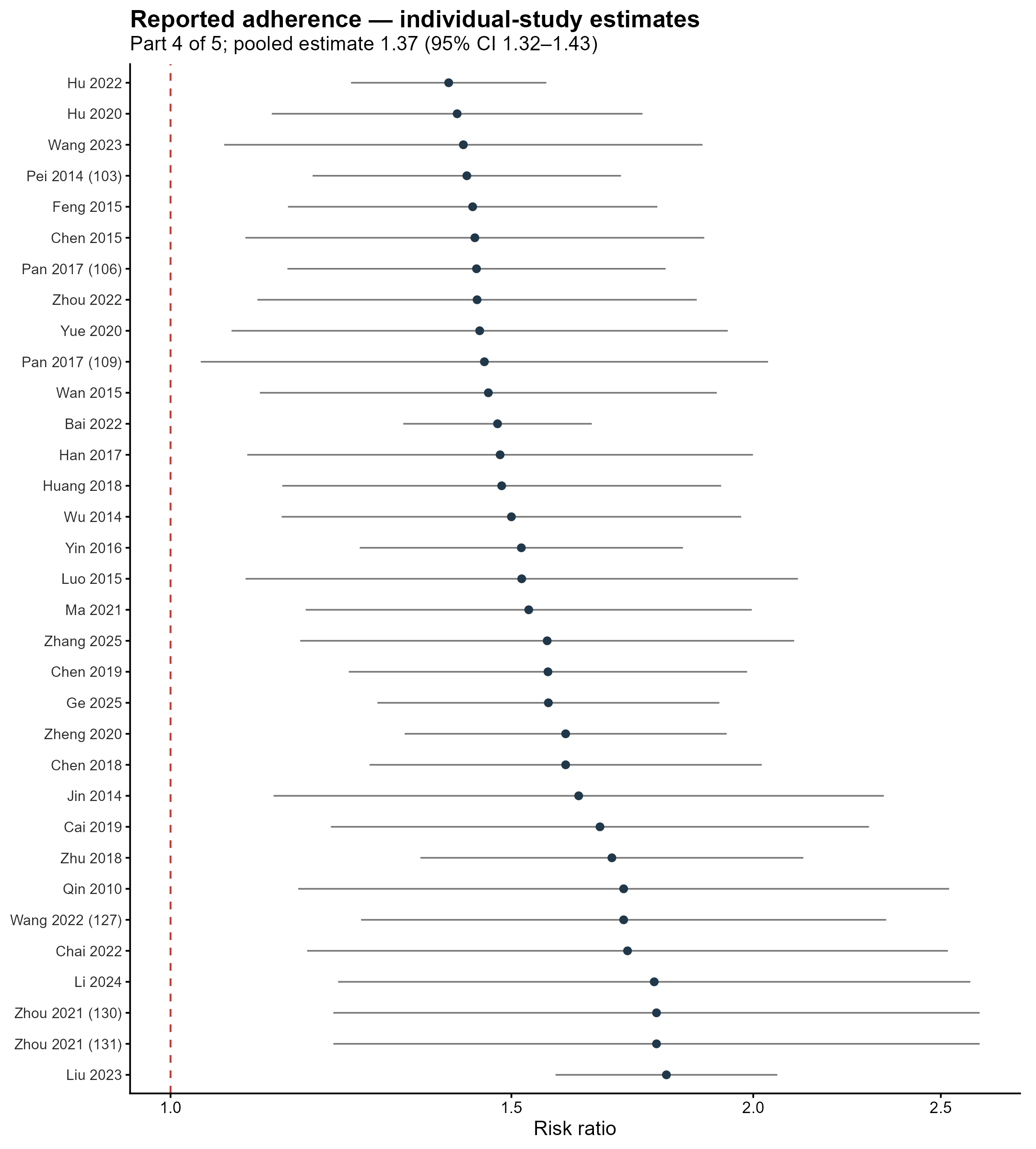

Supplementary Figure S8 | Adherence-Rate individual-study forest plot, part 4 of 5. The dashed red line is the null; pooled estimates are stated in the subtitle.

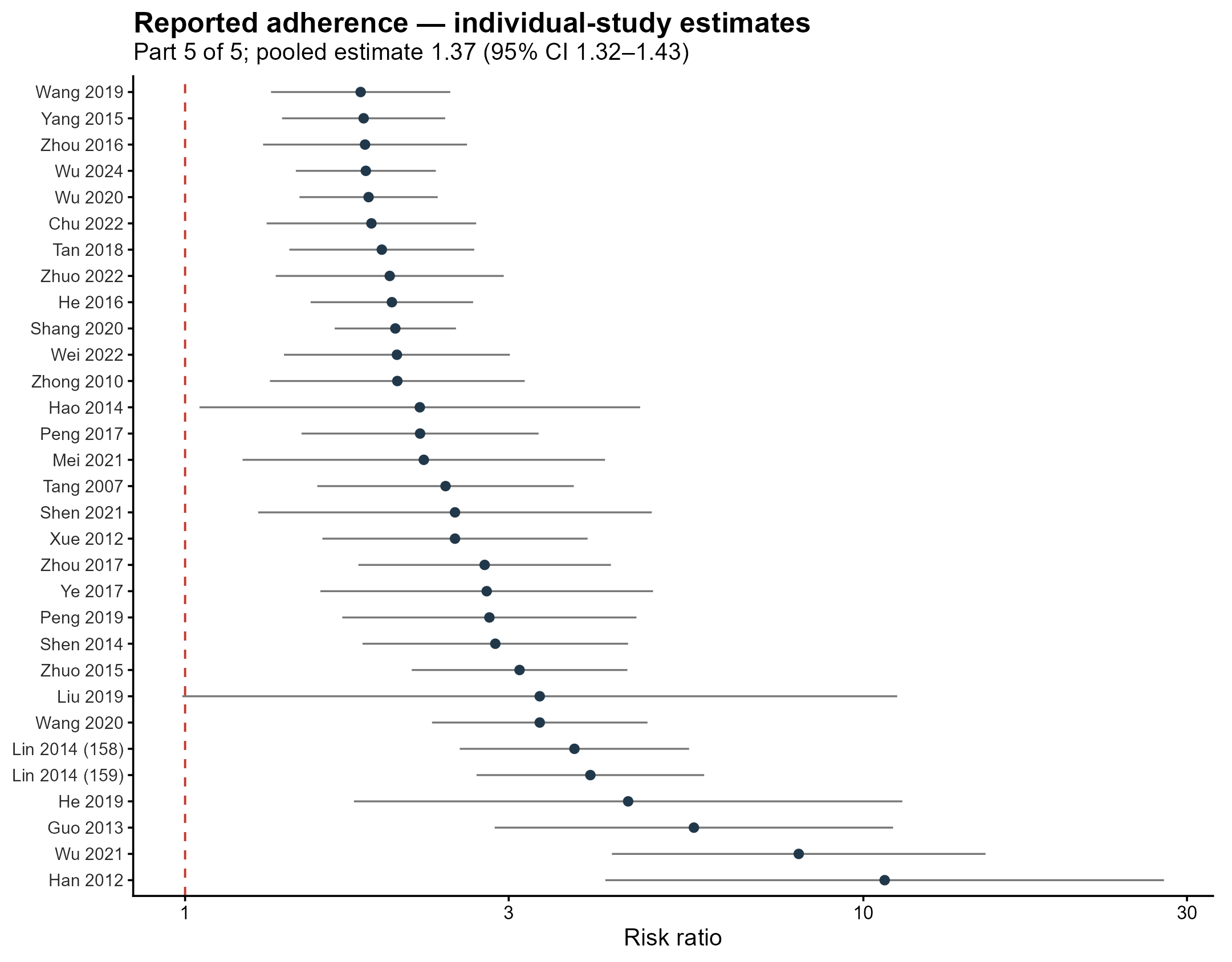

Supplementary Figure S9 | Adherence-Rate individual-study forest plot, part 5 of 5. The dashed red line is the null; pooled estimates are stated in the subtitle.

Supplementary Figure S10 | ADR individual-study forest plot, part 1 of 3. The dashed red line is the null; pooled estimates are stated in the subtitle.

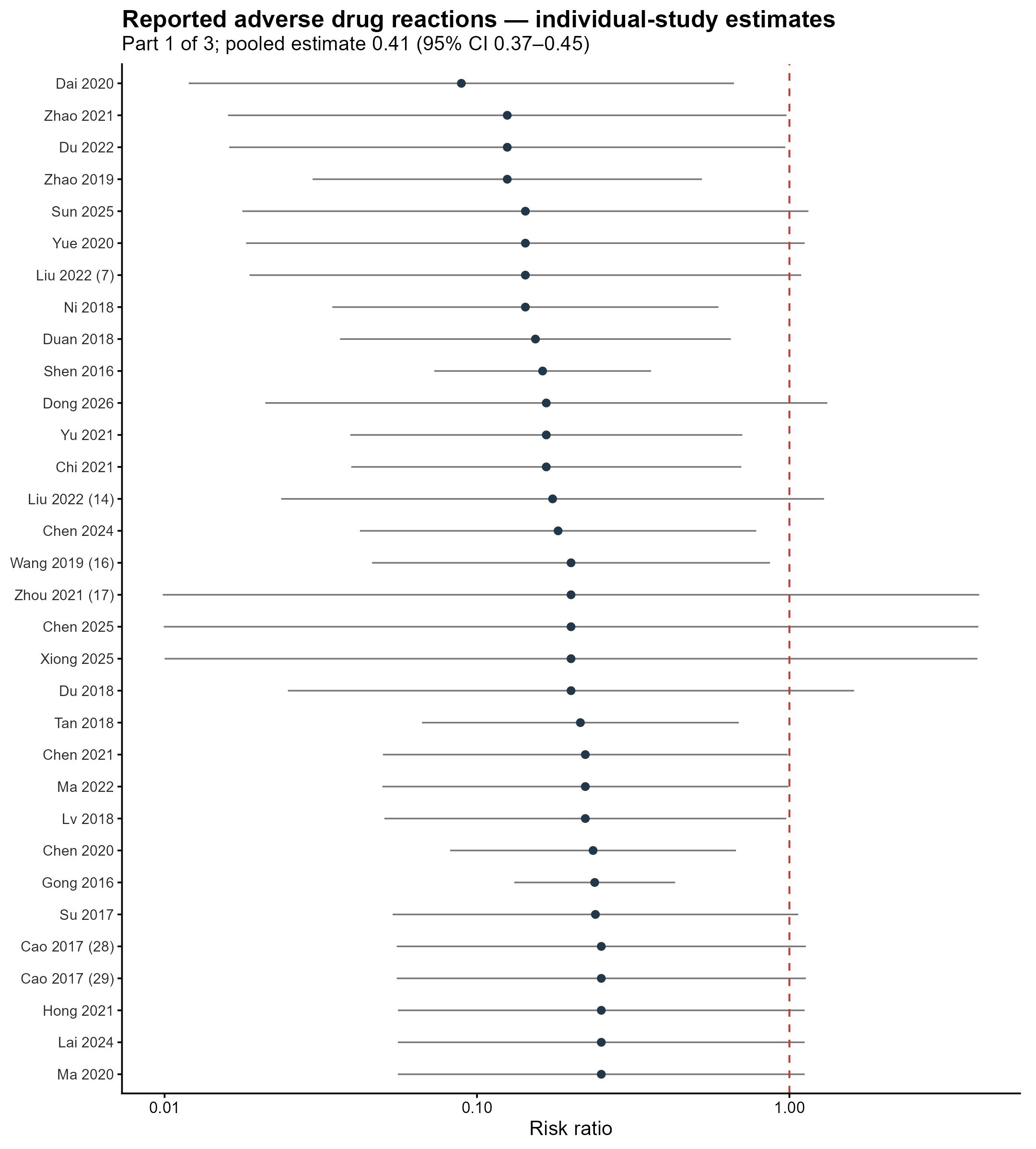

Supplementary Figure S11 | ADR individual-study forest plot, part 2 of 3. The dashed red line is the null; pooled estimates are stated in the subtitle.

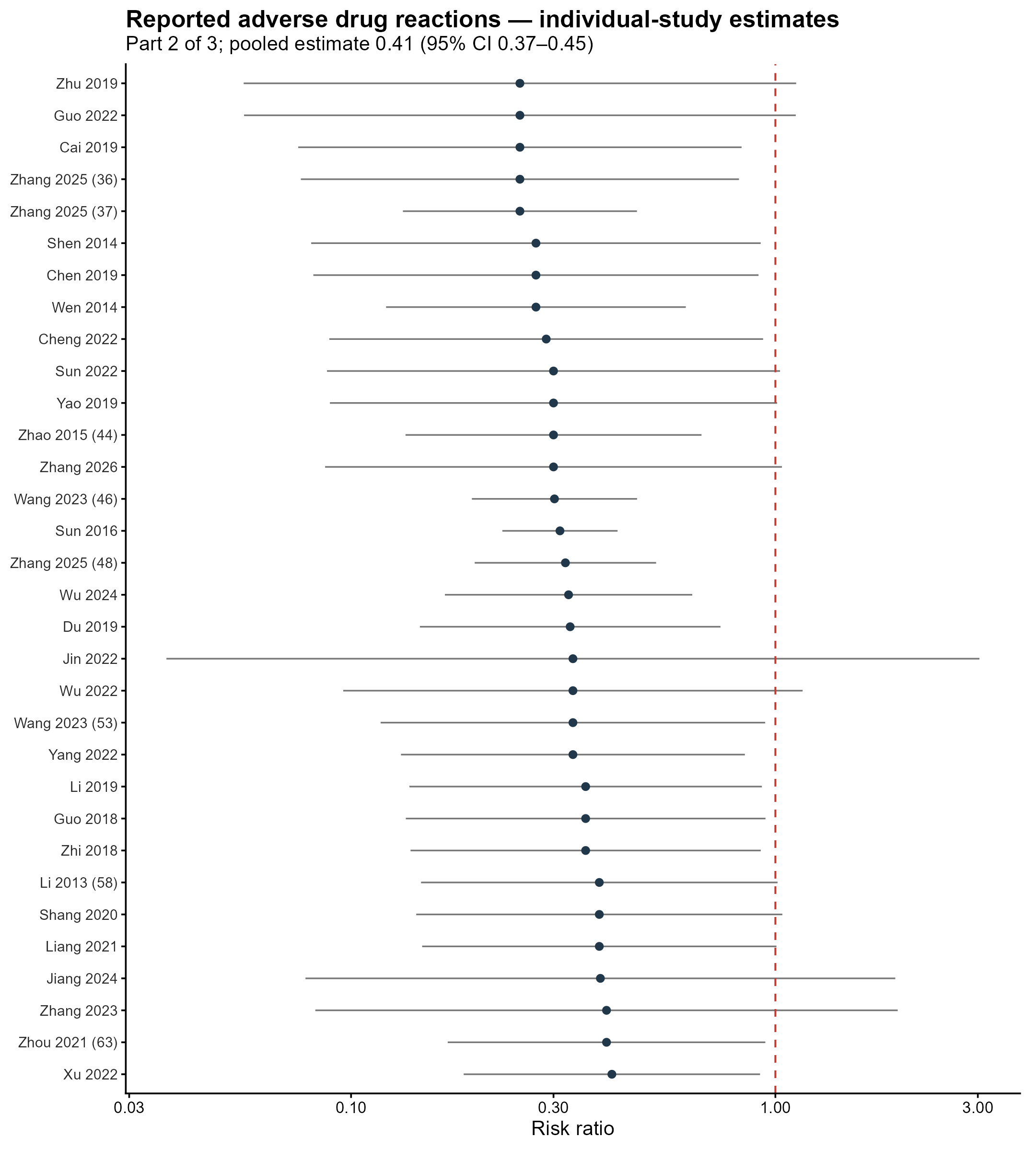

Supplementary Figure S12 | ADR individual-study forest plot, part 3 of 3. The dashed red line is the null; pooled estimates are stated in the subtitle.

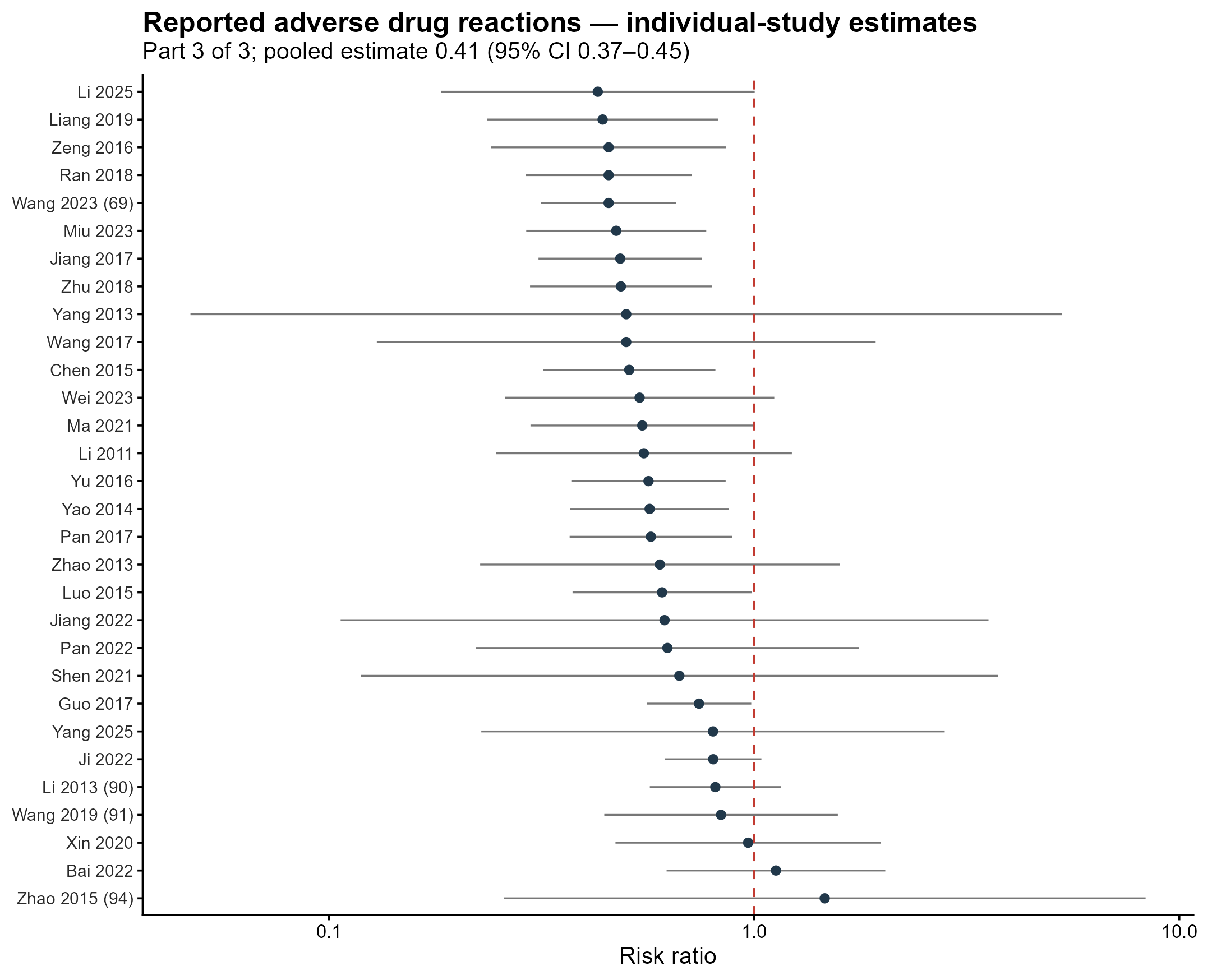

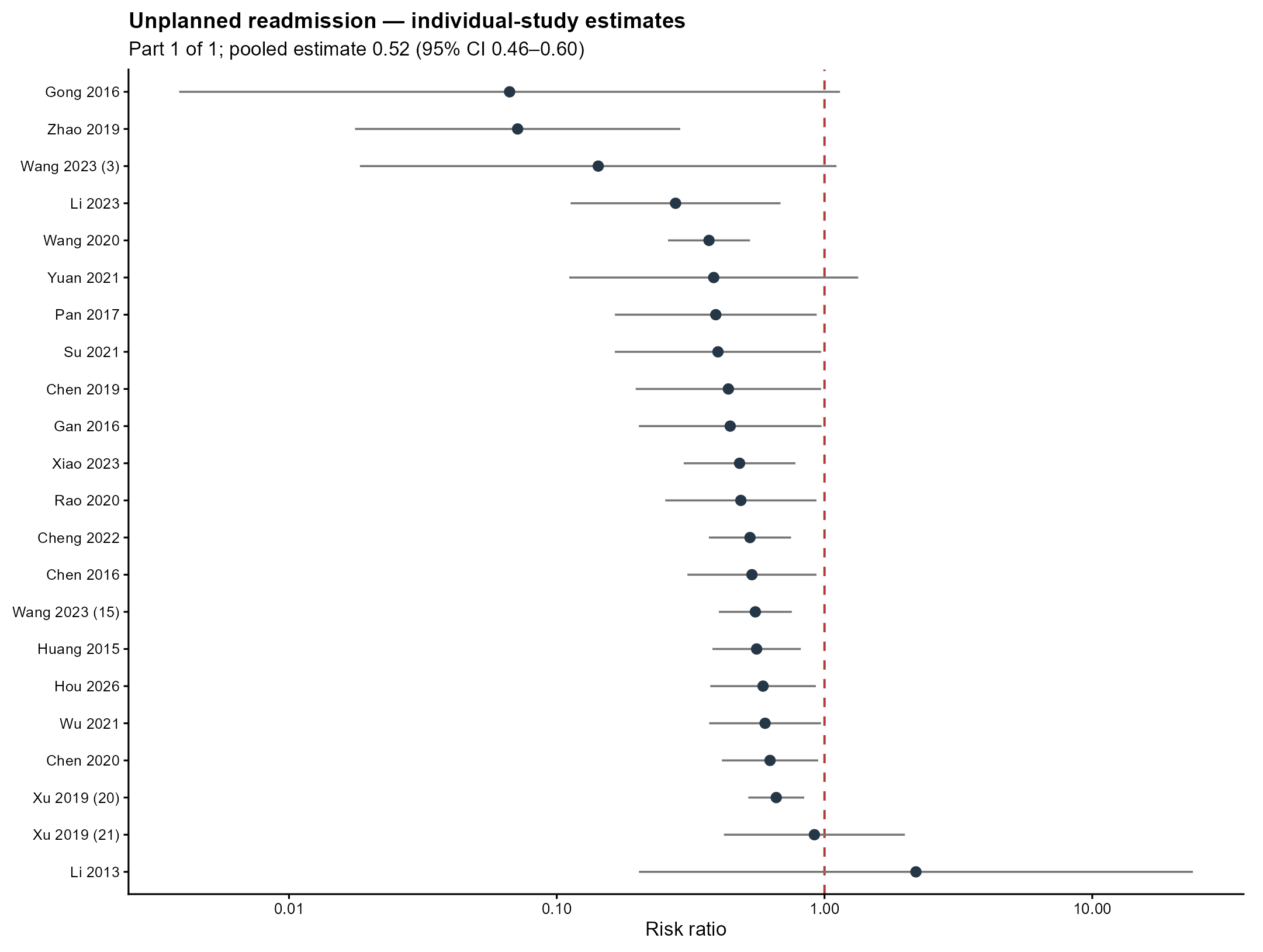

Supplementary Figure S13 | Readmission individual-study forest plot, part 1 of 1. The dashed red line is the null; pooled estimates are stated in the subtitle.

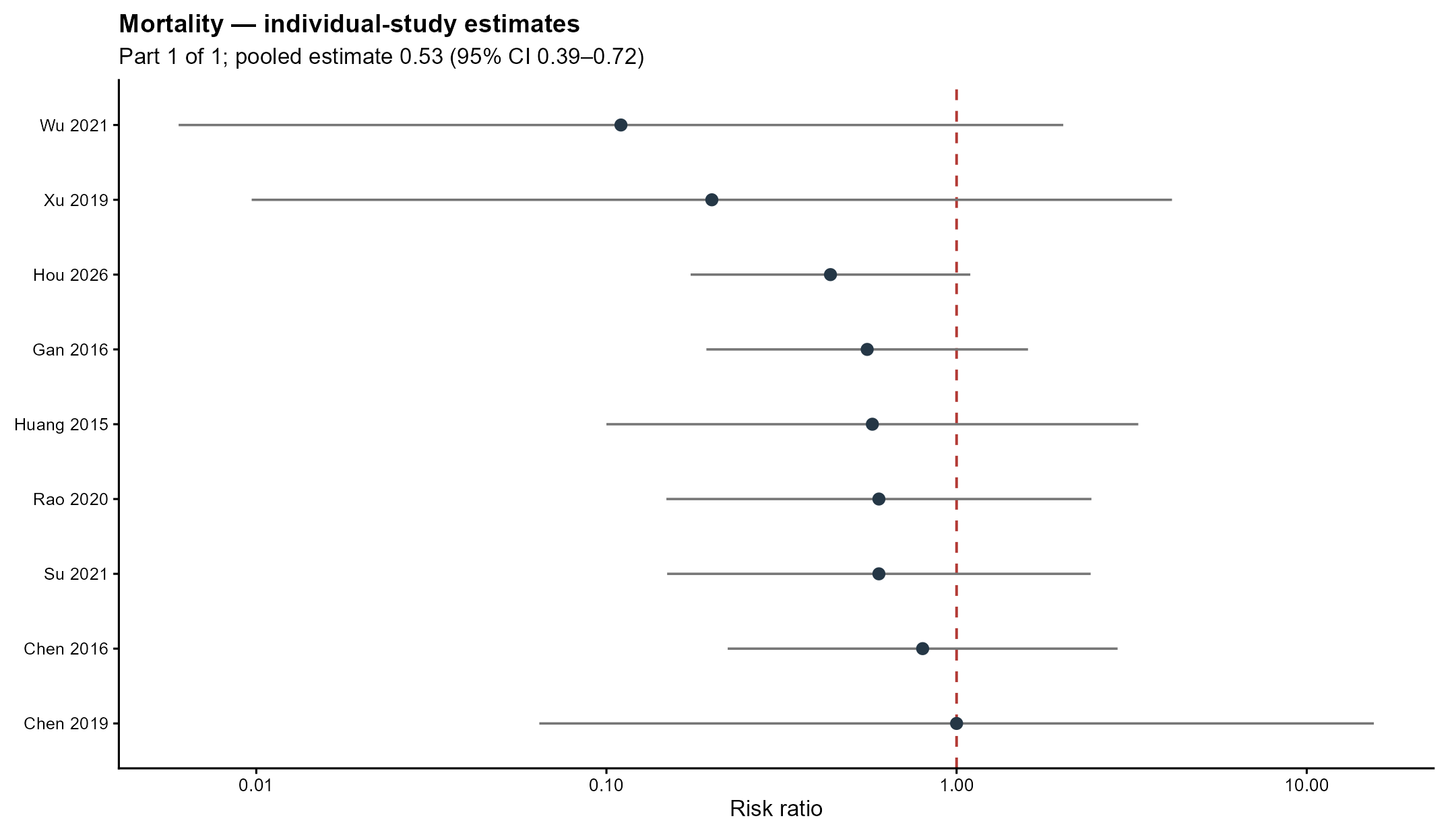

Supplementary Figure S14 | Mortality individual-study forest plot, part 1 of 1. The dashed red line is the null; pooled estimates are stated in the subtitle.

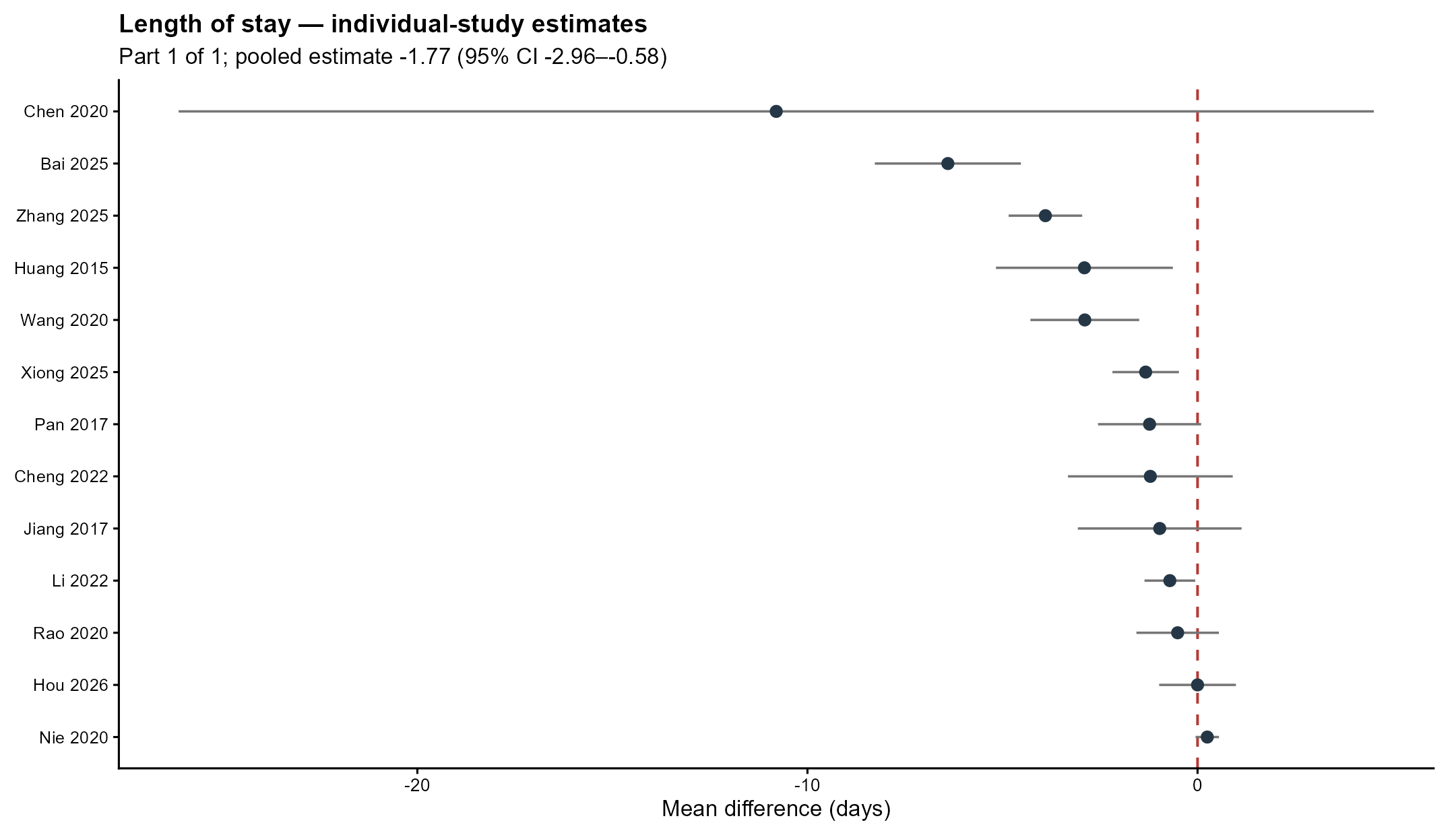

Supplementary Figure S15 | Length_of_Stay individual-study forest plot, part 1 of 1. The dashed red line is the null; pooled estimates are stated in the subtitle.

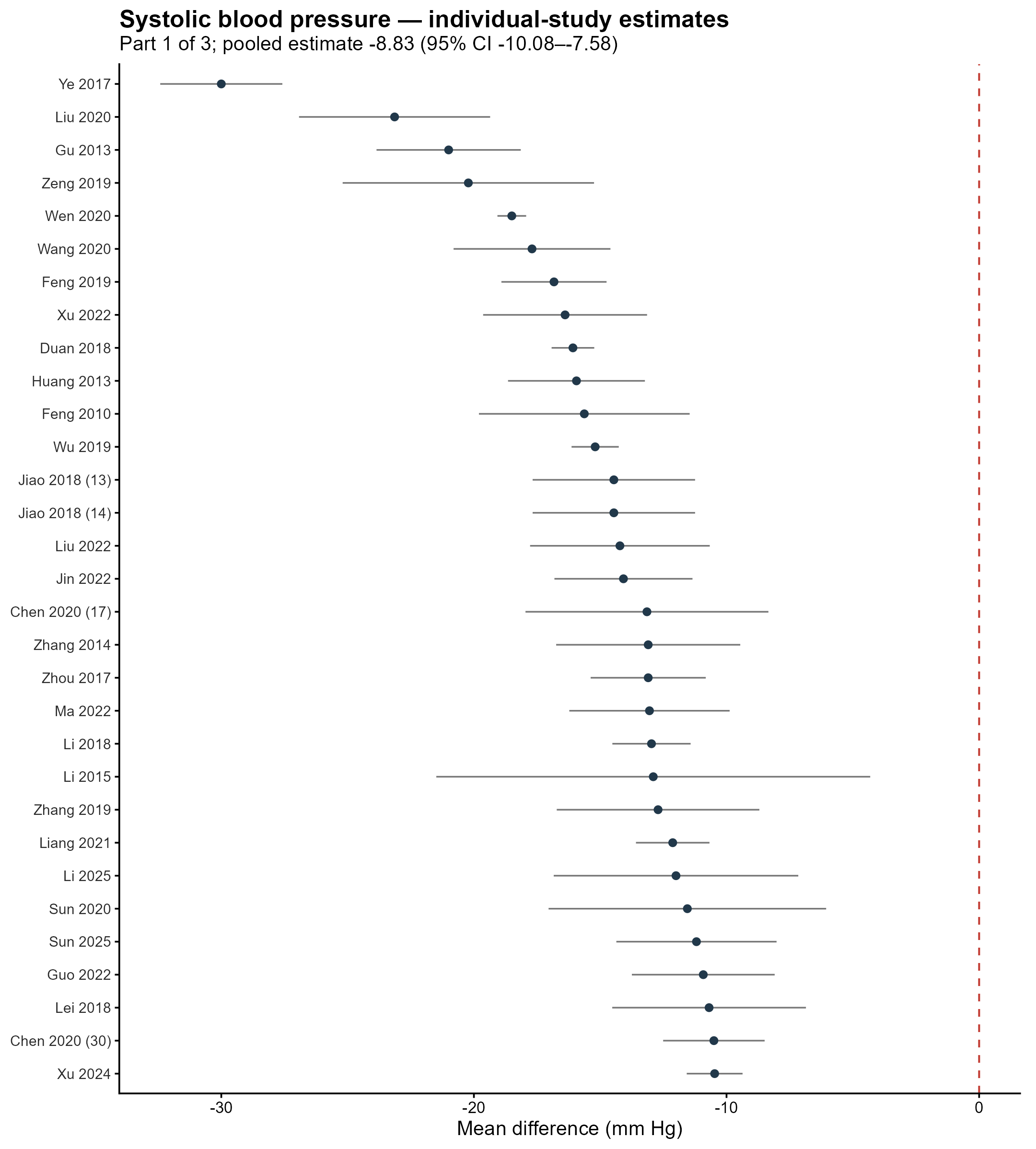

Supplementary Figure S16 | SBP individual-study forest plot, part 1 of 3. The dashed red line is the null; pooled estimates are stated in the subtitle.

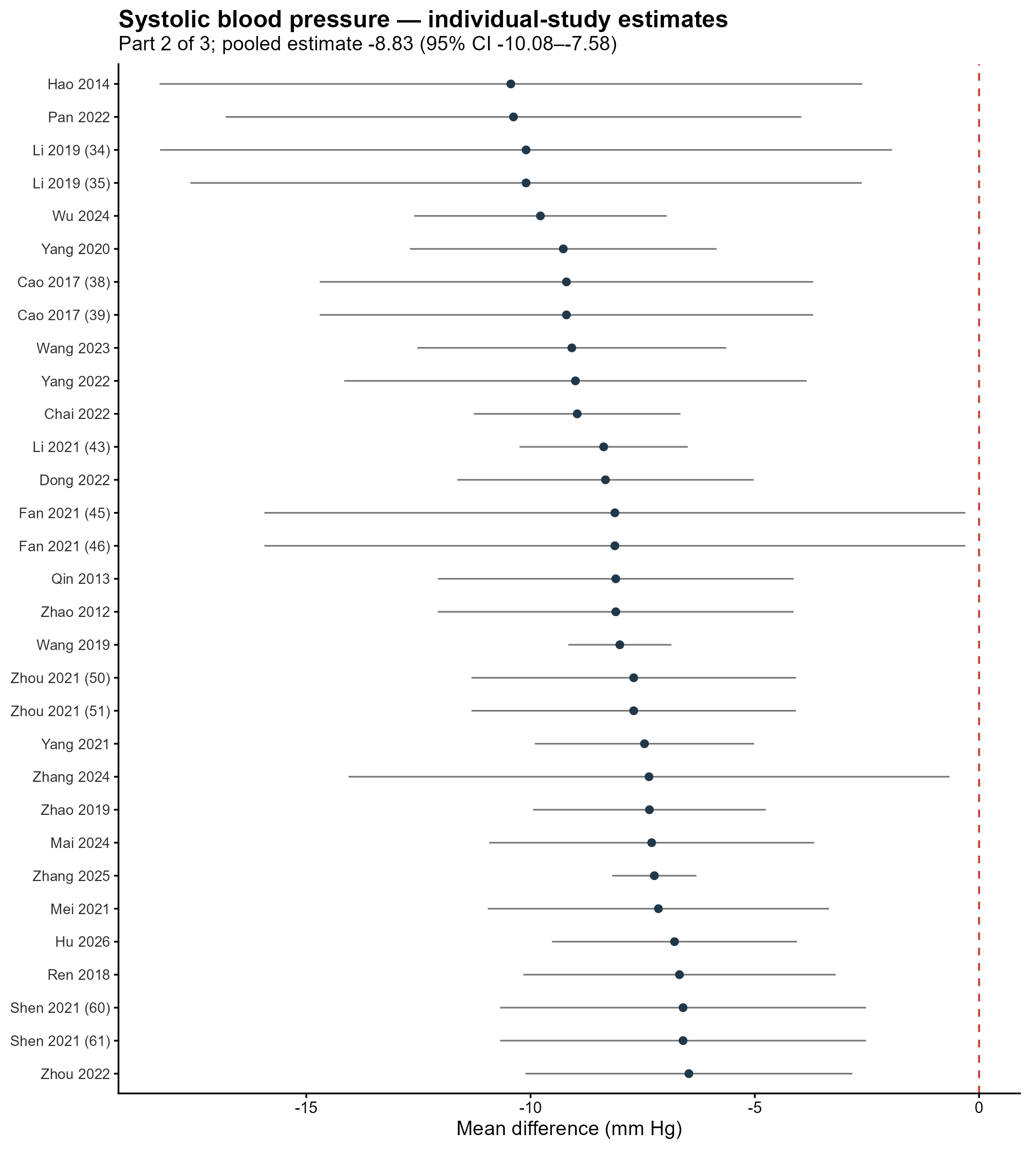

Supplementary Figure S17 | SBP individual-study forest plot, part 2 of 3. The dashed red line is the null; pooled estimates are stated in the subtitle.

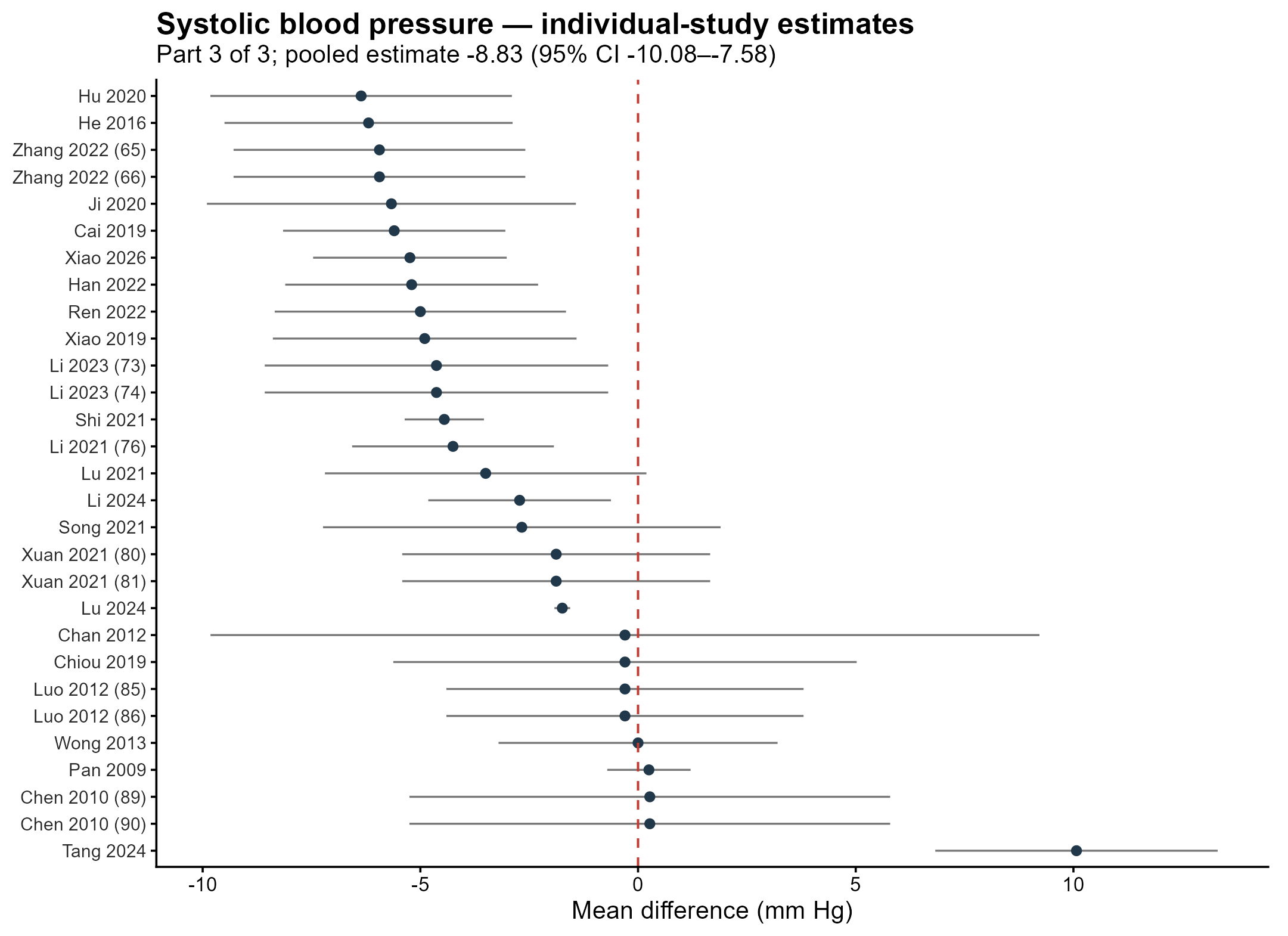

Supplementary Figure S18 | SBP individual-study forest plot, part 3 of 3. The dashed red line is the null; pooled estimates are stated in the subtitle.

Supplementary Figure S19 | HbA1c individual-study forest plot, part 1 of 4. The dashed red line is the null; pooled estimates are stated in the subtitle.

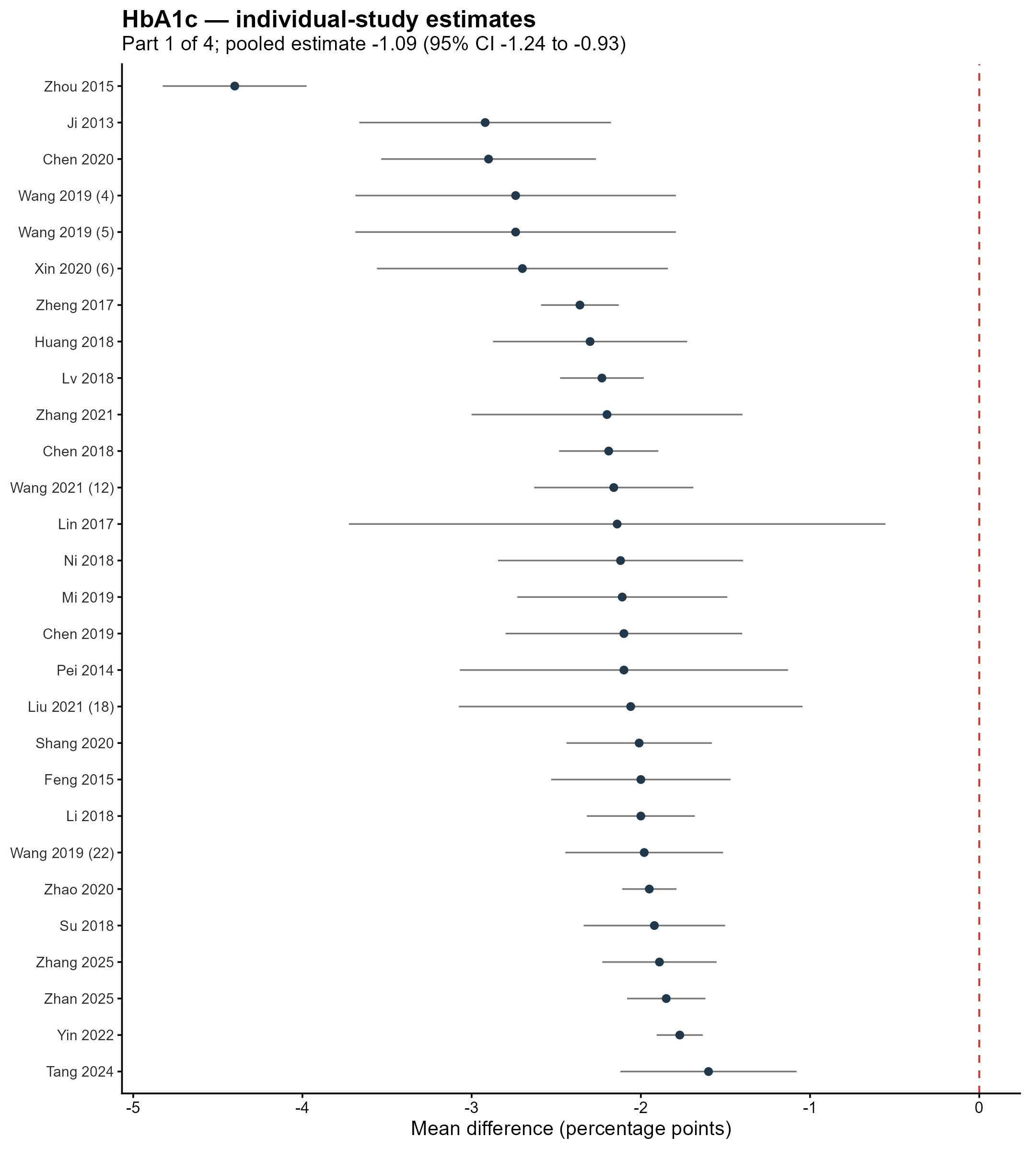

Supplementary Figure S20 | HbA1c individual-study forest plot, part 2 of 4. The dashed red line is the null; pooled estimates are stated in the subtitle.

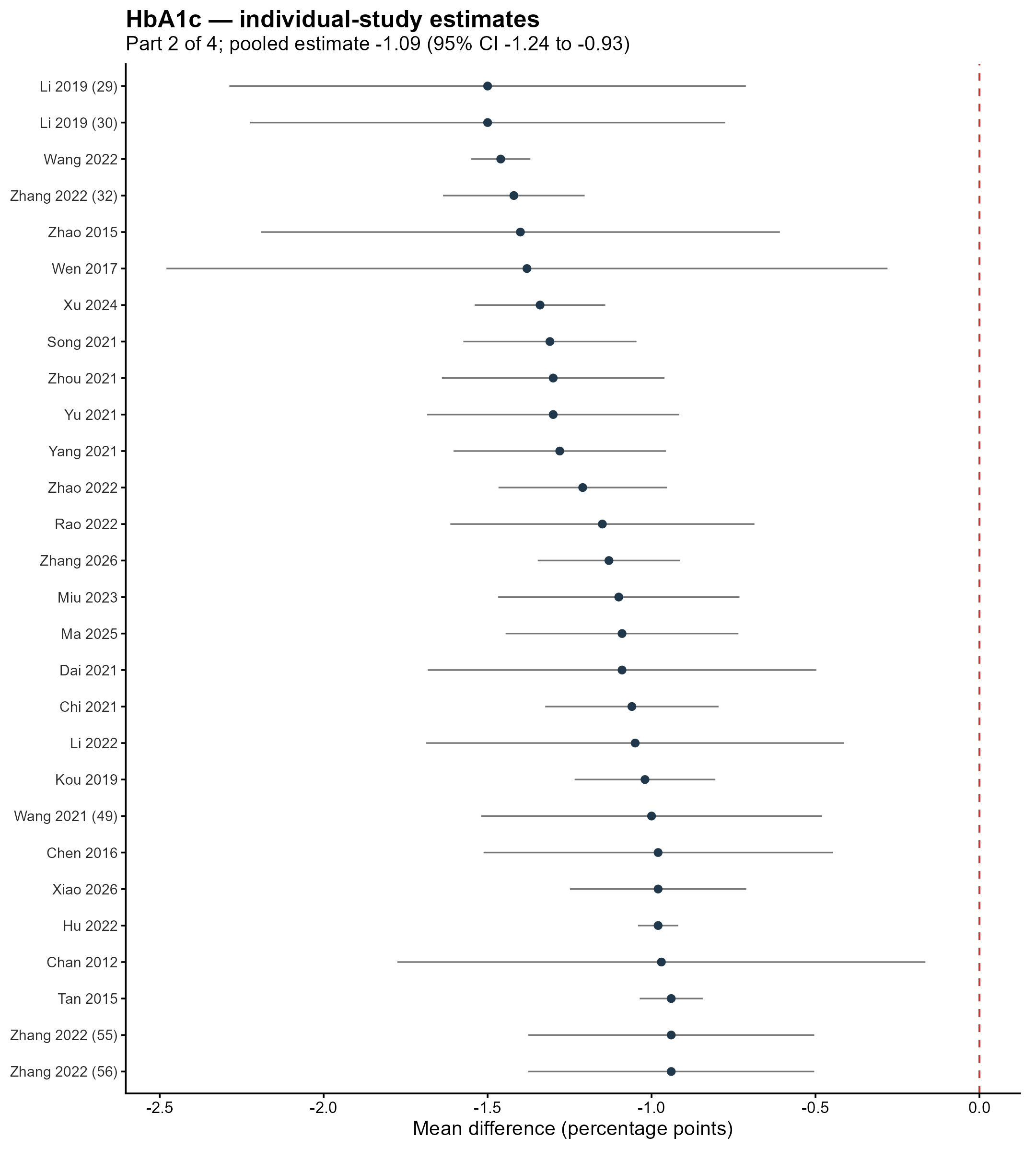

Supplementary Figure S21 | HbA1c individual-study forest plot, part 3 of 4. The dashed red line is the null; pooled estimates are stated in the subtitle.

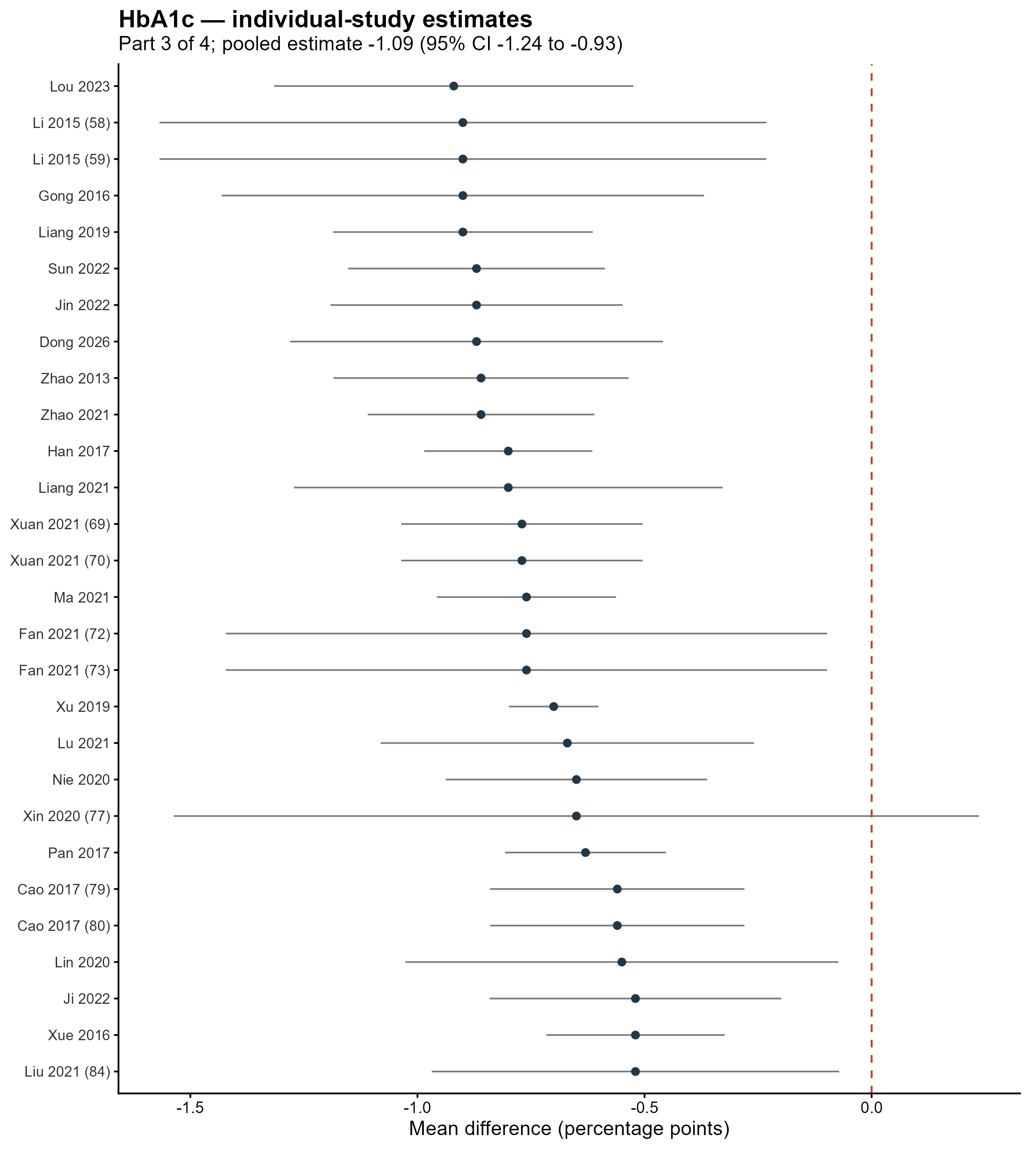

Supplementary Figure S22 | HbA1c individual-study forest plot, part 4 of 4. The dashed red line is the null; pooled estimates are stated in the subtitle.

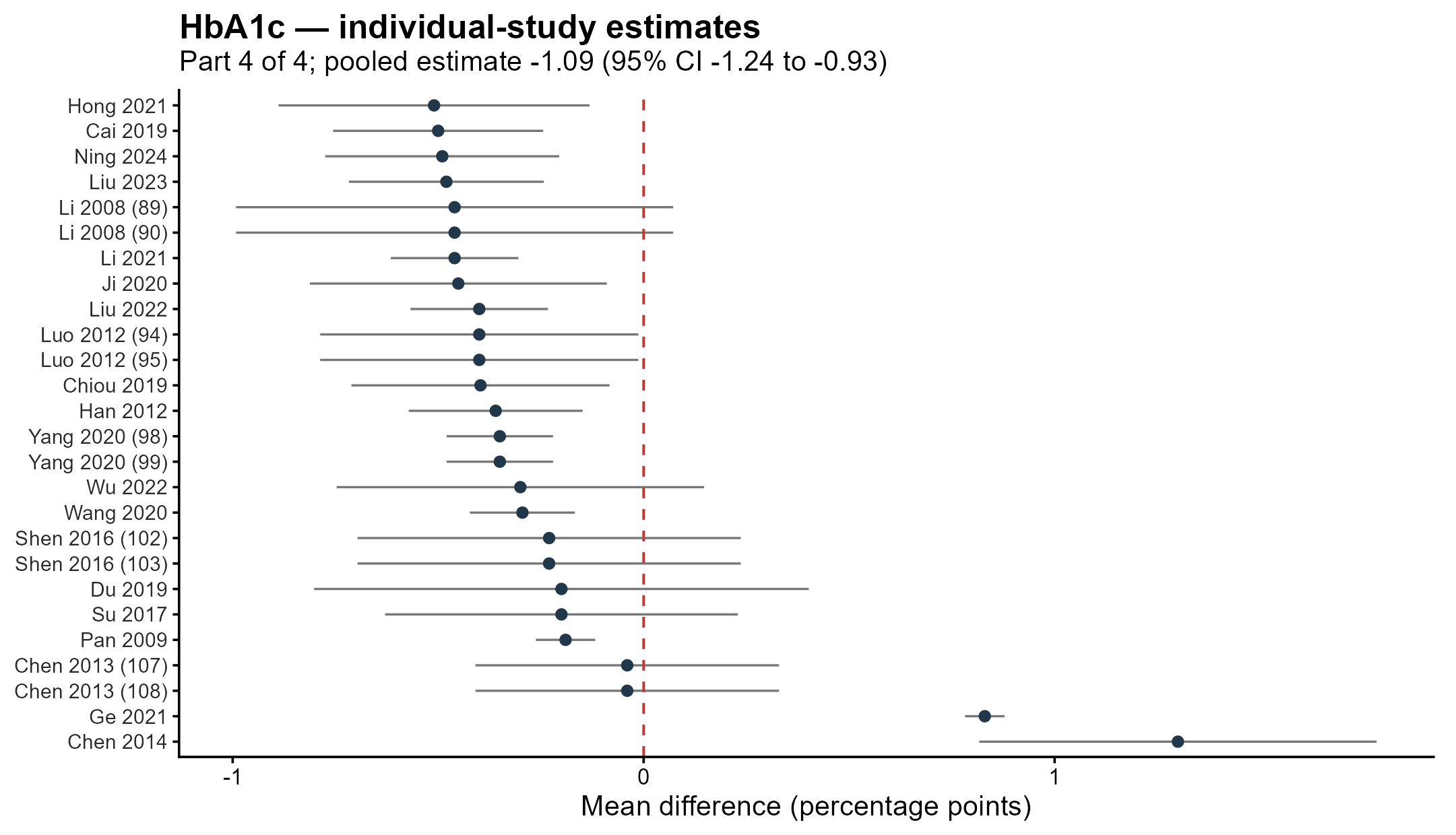

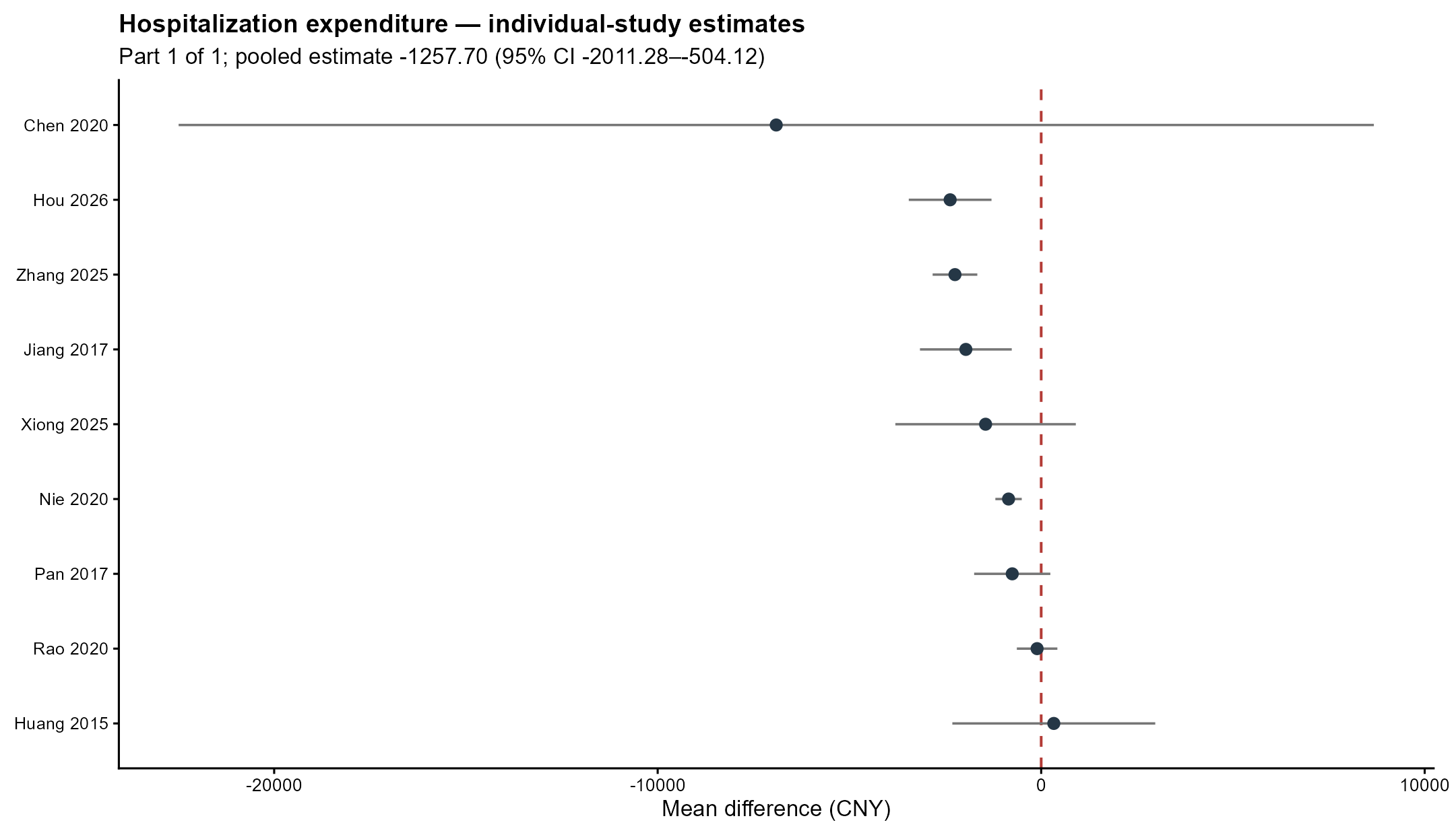

Supplementary Figure S23 | Cost_of_Hospitalization individual-study forest plot, part 1 of 1. The dashed red line is the null; pooled estimates are stated in the subtitle.

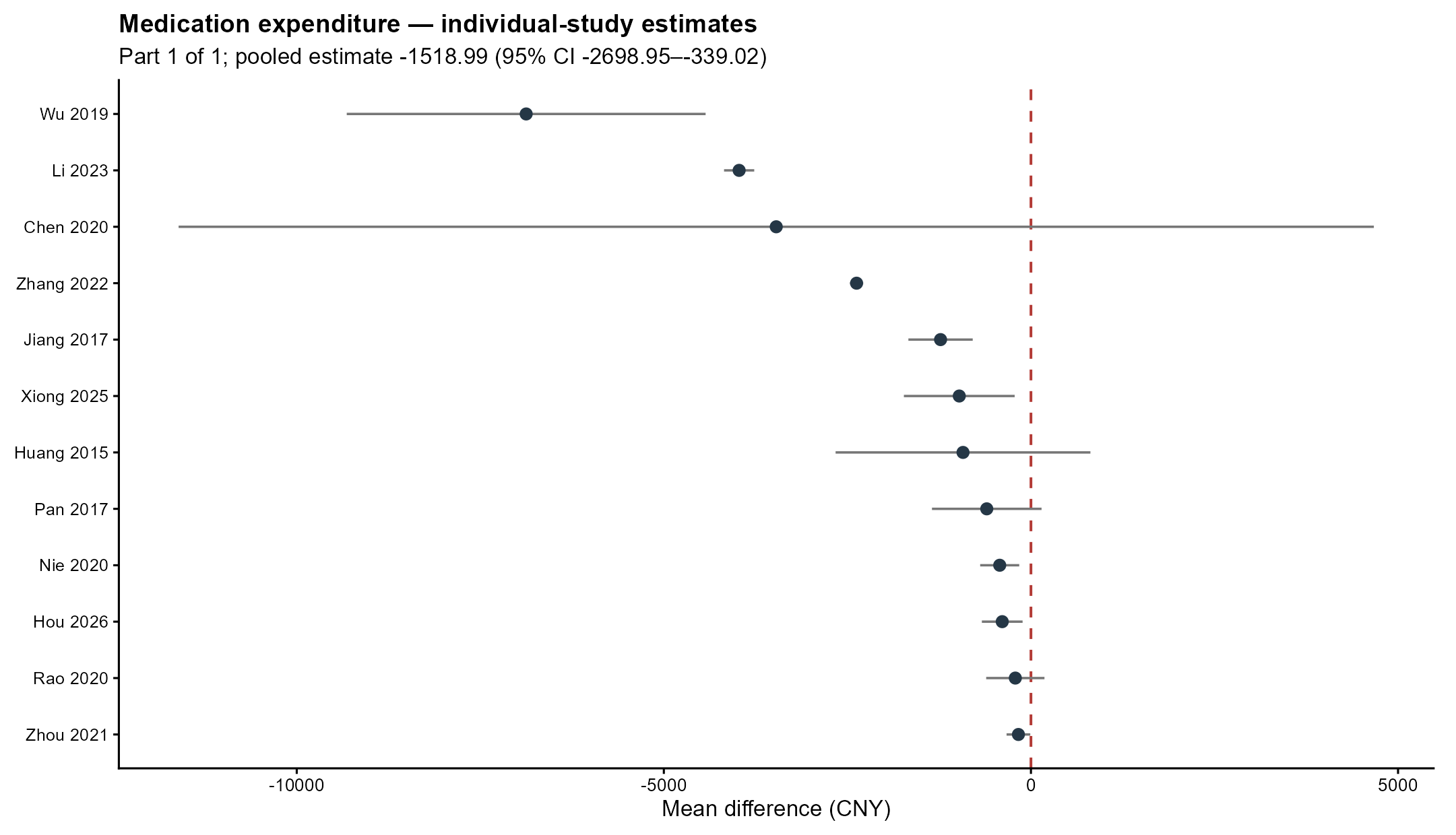

Supplementary Figure S24 | Medication_Cost individual-study forest plot, part 1 of 1. The dashed red line is the null; pooled estimates are stated in the subtitle.

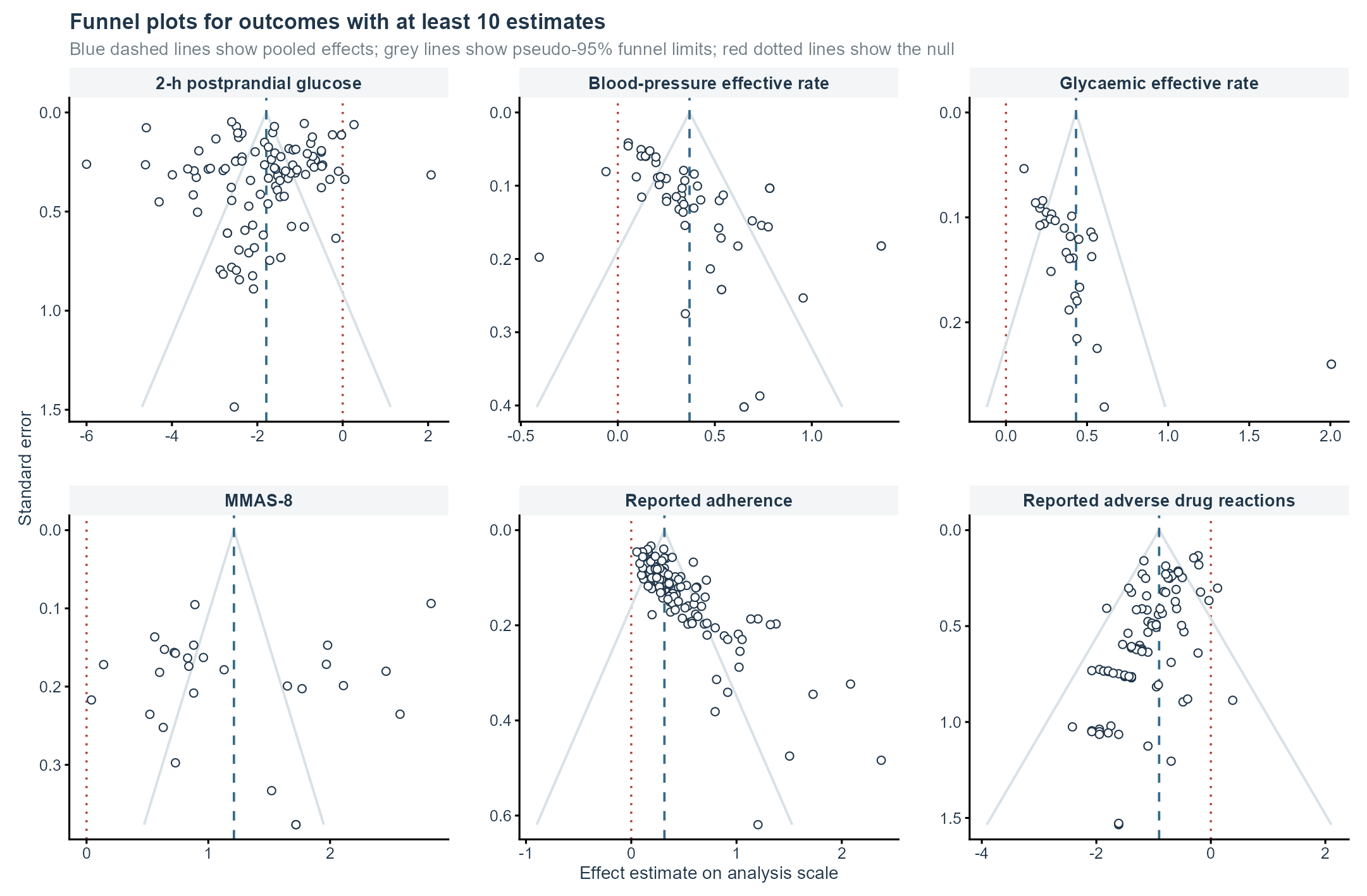

Supplementary Figure S25 | Funnel plots for outcomes with at least 10 estimates, part 1. Panels use outcome-specific axes. Blue dashed lines show pooled effects, grey lines pseudo-95% funnel limits, and red dotted lines the null. These diagnostic displays are not proof of publication bias.

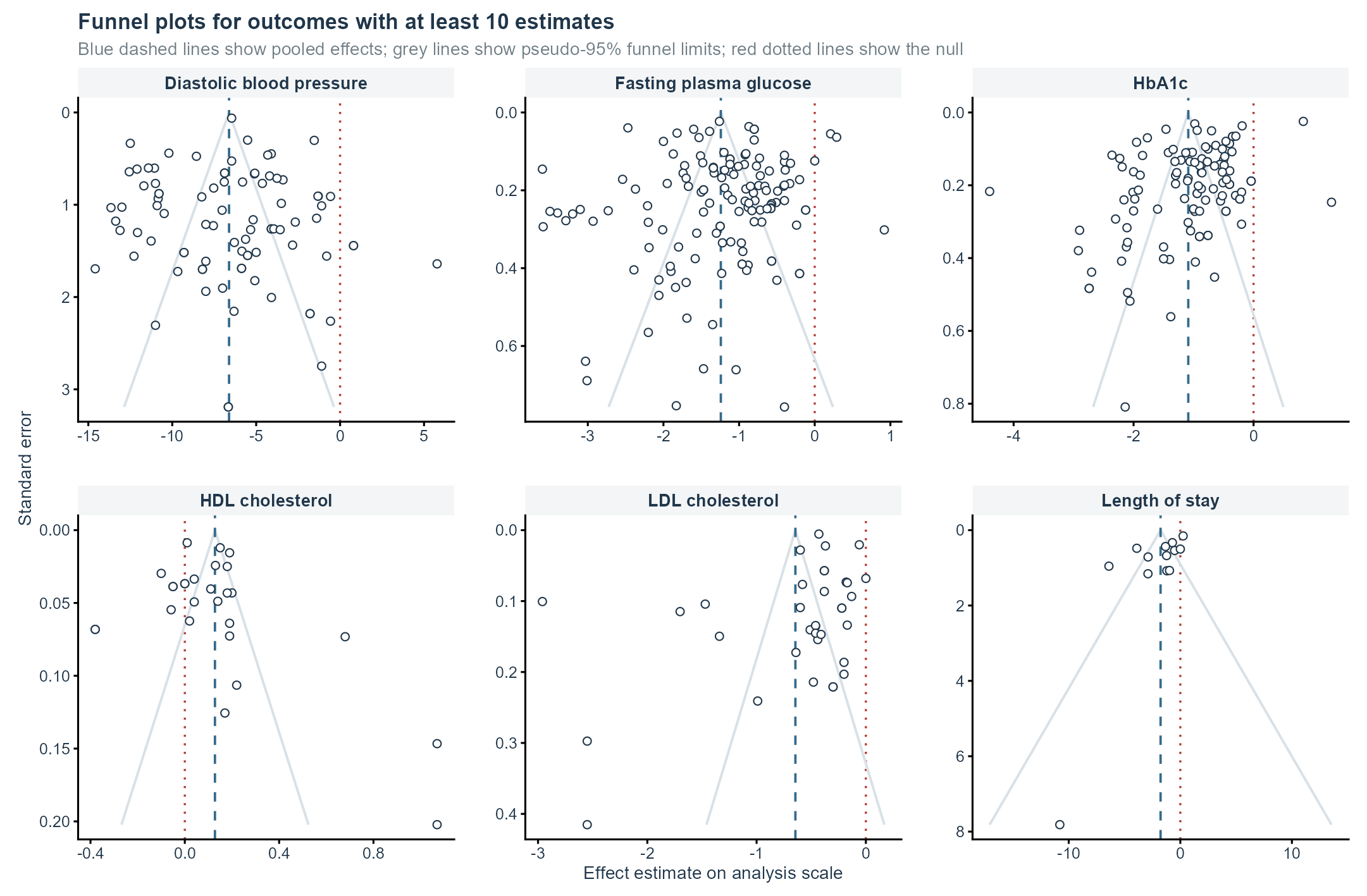

Supplementary Figure S26 | Funnel plots for outcomes with at least 10 estimates, part 2. Panels use outcome-specific axes. Blue dashed lines show pooled effects, grey lines pseudo-95% funnel limits, and red dotted lines the null. These diagnostic displays are not proof of publication bias.

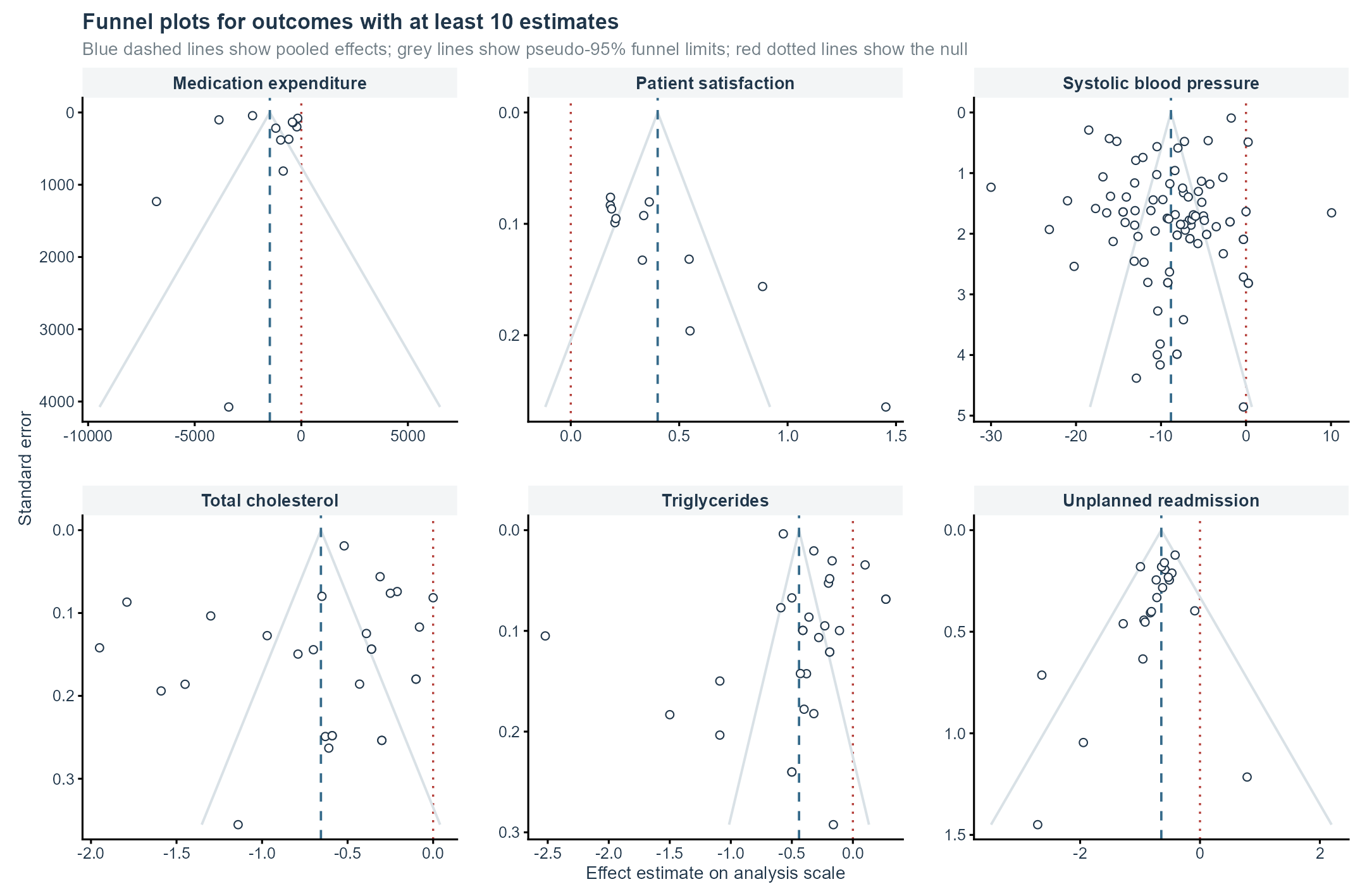

Supplementary Figure S27 | Funnel plots for outcomes with at least 10 estimates, part 3. Panels use outcome-specific axes. Blue dashed lines show pooled effects, grey lines pseudo-95% funnel limits, and red dotted lines the null. These diagnostic displays are not proof of publication bias.

### Appendix 1 | Database search strategies and execution record

Reproducibility note: The search counts and PRISMA flow derive from the final search run completed through July 20, 2026. The database-specific strategies reproduced below are the strategies used for that search. Chinese-language query strings are shown as executed. No study-design, journal-type, or other database filter was appended.

Supplementary search methods

Chinese-language journal-scope criterion: During conduct of the review, reports in Chinese were restricted to journals included in the union of the 2023 Peking University core list, the 2023-2024 CSSCI source-journal list, and the 2024 Chinese Science and Technology Core list. This criterion defined the primary evidence scope and was not used as a proxy for trial quality.

Final executed database-specific search strategies

Pharmaceutical services for six cardiometabolic diseases in China

Databases: PubMed, Ovid Embase, Cochrane Library, China National Knowledge Infrastructure (CNKI), Wanfang Data, and SinoMed

| **Framework** | **Pharmaceutical-service concept AND disease concept AND China-region concept.** |
| --- | --- |
| Disease sets | Hypertension; Diabetes Mellitus; Coronary Disease; Dyslipidemia; Heart Failure; Myocardial Infarction. |
| Main strategy | PubMed is the conceptual master. Controlled vocabulary and field syntax are translated for each database. |
| Chinese databases | Chinese disease/service synonyms are used. A China-country term is not required by default because it can reduce recall; study location should be checked during screening. |
| Filters | No study-design, journal-type, or other database filter was appended; the strategies contain only pharmaceutical-service, disease, and China-region concepts. |

### PubMed

Master strategy. MeSH terms are combined with explicit Title/Abstract synonyms to capture indexed and not-yet-indexed records.

#### Hypertension

**("Medication Therapy Management"[Mesh] OR "Pharmaceutical Services"[Mesh] OR "Pharmacists"[Mesh] OR "Medication Reconciliation"[Mesh] OR "Community Pharmacy Services"[Mesh] OR "Pharmacy Service, Hospital"[Mesh] OR pharmacist*[Title/Abstract] OR "clinical pharmacy"[Title/Abstract] OR "clinical pharmacist"[Title/Abstract] OR "clinical pharmacists"[Title/Abstract] OR "pharmaceutical care"[Title/Abstract] OR "pharmaceutical service"[Title/Abstract] OR "pharmaceutical services"[Title/Abstract] OR "pharmacy service"[Title/Abstract] OR "pharmacy services"[Title/Abstract] OR "medication therapy management"[Title/Abstract] OR "medication management"[Title/Abstract] OR "medication review"[Title/Abstract] OR "medication reviews"[Title/Abstract] OR "medicines review"[Title/Abstract] OR "medicines reviews"[Title/Abstract] OR "medication reconciliation"[Title/Abstract] OR "pharmacist intervention"[Title/Abstract] OR "pharmacist interventions"[Title/Abstract] OR "pharmacist led"[Title/Abstract] OR "pharmacy led"[Title/Abstract] OR "community pharmacy service"[Title/Abstract] OR "community pharmacy services"[Title/Abstract] OR "hospital pharmacy service"[Title/Abstract] OR "hospital pharmacy services"[Title/Abstract]) AND ("Hypertension"[Mesh] OR hypertens*[Title/Abstract] OR "high blood pressure"[Title/Abstract]) AND ("China"[Mesh] OR China[Title/Abstract] OR Chinese[Title/Abstract] OR Taiwan[Title/Abstract] OR "Hong Kong"[Title/Abstract] OR Hongkong[Title/Abstract] OR Macau[Title/Abstract] OR Macao[Title/Abstract])**

#### Diabetes Mellitus

**("Medication Therapy Management"[Mesh] OR "Pharmaceutical Services"[Mesh] OR "Pharmacists"[Mesh] OR "Medication Reconciliation"[Mesh] OR "Community Pharmacy Services"[Mesh] OR "Pharmacy Service, Hospital"[Mesh] OR pharmacist*[Title/Abstract] OR "clinical pharmacy"[Title/Abstract] OR "clinical pharmacist"[Title/Abstract] OR "clinical pharmacists"[Title/Abstract] OR "pharmaceutical care"[Title/Abstract] OR "pharmaceutical service"[Title/Abstract] OR "pharmaceutical services"[Title/Abstract] OR "pharmacy service"[Title/Abstract] OR "pharmacy services"[Title/Abstract] OR "medication therapy management"[Title/Abstract] OR "medication management"[Title/Abstract] OR "medication review"[Title/Abstract] OR "medication reviews"[Title/Abstract] OR "medicines review"[Title/Abstract] OR "medicines reviews"[Title/Abstract] OR "medication reconciliation"[Title/Abstract] OR "pharmacist intervention"[Title/Abstract] OR "pharmacist interventions"[Title/Abstract] OR "pharmacist led"[Title/Abstract] OR "pharmacy led"[Title/Abstract] OR "community pharmacy service"[Title/Abstract] OR "community pharmacy services"[Title/Abstract] OR "hospital pharmacy service"[Title/Abstract] OR "hospital pharmacy services"[Title/Abstract]) AND ("Diabetes Mellitus"[Mesh] OR diabet*[Title/Abstract] OR hyperglycemi*[Title/Abstract] OR "high blood glucose"[Title/Abstract]) AND ("China"[Mesh] OR China[Title/Abstract] OR Chinese[Title/Abstract] OR Taiwan[Title/Abstract] OR "Hong Kong"[Title/Abstract] OR Hongkong[Title/Abstract] OR Macau[Title/Abstract] OR Macao[Title/Abstract])**

#### Coronary Disease

**("Medication Therapy Management"[Mesh] OR "Pharmaceutical Services"[Mesh] OR "Pharmacists"[Mesh] OR "Medication Reconciliation"[Mesh] OR "Community Pharmacy Services"[Mesh] OR "Pharmacy Service, Hospital"[Mesh] OR pharmacist*[Title/Abstract] OR "clinical pharmacy"[Title/Abstract] OR "clinical pharmacist"[Title/Abstract] OR "clinical pharmacists"[Title/Abstract] OR "pharmaceutical care"[Title/Abstract] OR "pharmaceutical service"[Title/Abstract] OR "pharmaceutical services"[Title/Abstract] OR "pharmacy service"[Title/Abstract] OR "pharmacy services"[Title/Abstract] OR "medication therapy management"[Title/Abstract] OR "medication management"[Title/Abstract] OR "medication review"[Title/Abstract] OR "medication reviews"[Title/Abstract] OR "medicines review"[Title/Abstract] OR "medicines reviews"[Title/Abstract] OR "medication reconciliation"[Title/Abstract] OR "pharmacist intervention"[Title/Abstract] OR "pharmacist interventions"[Title/Abstract] OR "pharmacist led"[Title/Abstract] OR "pharmacy led"[Title/Abstract] OR "community pharmacy service"[Title/Abstract] OR "community pharmacy services"[Title/Abstract] OR "hospital pharmacy service"[Title/Abstract] OR "hospital pharmacy services"[Title/Abstract]) AND ("Coronary Disease"[Mesh] OR "Coronary Artery Disease"[Mesh] OR "coronary disease"[Title/Abstract] OR "coronary artery disease"[Title/Abstract] OR "coronary heart disease"[Title/Abstract] OR "ischemic heart disease"[Title/Abstract] OR "ischaemic heart disease"[Title/Abstract]) AND ("China"[Mesh] OR China[Title/Abstract] OR Chinese[Title/Abstract] OR Taiwan[Title/Abstract] OR "Hong Kong"[Title/Abstract] OR Hongkong[Title/Abstract] OR Macau[Title/Abstract] OR Macao[Title/Abstract])**

#### Dyslipidemia

**("Medication Therapy Management"[Mesh] OR "Pharmaceutical Services"[Mesh] OR "Pharmacists"[Mesh] OR "Medication Reconciliation"[Mesh] OR "Community Pharmacy Services"[Mesh] OR "Pharmacy Service, Hospital"[Mesh] OR pharmacist*[Title/Abstract] OR "clinical pharmacy"[Title/Abstract] OR "clinical pharmacist"[Title/Abstract] OR "clinical pharmacists"[Title/Abstract] OR "pharmaceutical care"[Title/Abstract] OR "pharmaceutical service"[Title/Abstract] OR "pharmaceutical services"[Title/Abstract] OR "pharmacy service"[Title/Abstract] OR "pharmacy services"[Title/Abstract] OR "medication therapy management"[Title/Abstract] OR "medication management"[Title/Abstract] OR "medication review"[Title/Abstract] OR "medication reviews"[Title/Abstract] OR "medicines review"[Title/Abstract] OR "medicines reviews"[Title/Abstract] OR "medication reconciliation"[Title/Abstract] OR "pharmacist intervention"[Title/Abstract] OR "pharmacist interventions"[Title/Abstract] OR "pharmacist led"[Title/Abstract] OR "pharmacy led"[Title/Abstract] OR "community pharmacy service"[Title/Abstract] OR "community pharmacy services"[Title/Abstract] OR "hospital pharmacy service"[Title/Abstract] OR "hospital pharmacy services"[Title/Abstract]) AND ("Dyslipidemias"[Mesh] OR dyslipid*[Title/Abstract] OR hyperlipid*[Title/Abstract] OR hypercholester*[Title/Abstract] OR hypertriglycerid*[Title/Abstract] OR hyperlipemi*[Title/Abstract]) AND ("China"[Mesh] OR China[Title/Abstract] OR Chinese[Title/Abstract] OR Taiwan[Title/Abstract] OR "Hong Kong"[Title/Abstract] OR Hongkong[Title/Abstract] OR Macau[Title/Abstract] OR Macao[Title/Abstract])**

#### Heart Failure

**("Medication Therapy Management"[Mesh] OR "Pharmaceutical Services"[Mesh] OR "Pharmacists"[Mesh] OR "Medication Reconciliation"[Mesh] OR "Community Pharmacy Services"[Mesh] OR "Pharmacy Service, Hospital"[Mesh] OR pharmacist*[Title/Abstract] OR "clinical pharmacy"[Title/Abstract] OR "clinical pharmacist"[Title/Abstract] OR "clinical pharmacists"[Title/Abstract] OR "pharmaceutical care"[Title/Abstract] OR "pharmaceutical service"[Title/Abstract] OR "pharmaceutical services"[Title/Abstract] OR "pharmacy service"[Title/Abstract] OR "pharmacy services"[Title/Abstract] OR "medication therapy management"[Title/Abstract] OR "medication management"[Title/Abstract] OR "medication review"[Title/Abstract] OR "medication reviews"[Title/Abstract] OR "medicines review"[Title/Abstract] OR "medicines reviews"[Title/Abstract] OR "medication reconciliation"[Title/Abstract] OR "pharmacist intervention"[Title/Abstract] OR "pharmacist interventions"[Title/Abstract] OR "pharmacist led"[Title/Abstract] OR "pharmacy led"[Title/Abstract] OR "community pharmacy service"[Title/Abstract] OR "community pharmacy services"[Title/Abstract] OR "hospital pharmacy service"[Title/Abstract] OR "hospital pharmacy services"[Title/Abstract]) AND ("Heart Failure"[Mesh] OR "heart failure"[Title/Abstract] OR "cardiac failure"[Title/Abstract] OR "ventricular failure"[Title/Abstract]) AND ("China"[Mesh] OR China[Title/Abstract] OR Chinese[Title/Abstract] OR Taiwan[Title/Abstract] OR "Hong Kong"[Title/Abstract] OR Hongkong[Title/Abstract] OR Macau[Title/Abstract] OR Macao[Title/Abstract])**

#### Myocardial Infarction

**("Medication Therapy Management"[Mesh] OR "Pharmaceutical Services"[Mesh] OR "Pharmacists"[Mesh] OR "Medication Reconciliation"[Mesh] OR "Community Pharmacy Services"[Mesh] OR "Pharmacy Service, Hospital"[Mesh] OR pharmacist*[Title/Abstract] OR "clinical pharmacy"[Title/Abstract] OR "clinical pharmacist"[Title/Abstract] OR "clinical pharmacists"[Title/Abstract] OR "pharmaceutical care"[Title/Abstract] OR "pharmaceutical service"[Title/Abstract] OR "pharmaceutical services"[Title/Abstract] OR "pharmacy service"[Title/Abstract] OR "pharmacy services"[Title/Abstract] OR "medication therapy management"[Title/Abstract] OR "medication management"[Title/Abstract] OR "medication review"[Title/Abstract] OR "medication reviews"[Title/Abstract] OR "medicines review"[Title/Abstract] OR "medicines reviews"[Title/Abstract] OR "medication reconciliation"[Title/Abstract] OR "pharmacist intervention"[Title/Abstract] OR "pharmacist interventions"[Title/Abstract] OR "pharmacist led"[Title/Abstract] OR "pharmacy led"[Title/Abstract] OR "community pharmacy service"[Title/Abstract] OR "community pharmacy services"[Title/Abstract] OR "hospital pharmacy service"[Title/Abstract] OR "hospital pharmacy services"[Title/Abstract]) AND ("Myocardial Infarction"[Mesh] OR "myocardial infarction"[Title/Abstract] OR "myocardial infarct"[Title/Abstract] OR "heart attack"[Title/Abstract]) AND ("China"[Mesh] OR China[Title/Abstract] OR Chinese[Title/Abstract] OR Taiwan[Title/Abstract] OR "Hong Kong"[Title/Abstract] OR Hongkong[Title/Abstract] OR Macau[Title/Abstract] OR Macao[Title/Abstract])**

### Ovid Embase

Platform assumption: Ovid Embase. Emtree headings are exploded and combined with title, abstract, and author-keyword terms.

#### Hypertension

**(exp medication therapy management/ or exp pharmaceutical care/ or exp pharmacist/ or exp medication review/ or exp medication reconciliation/ or exp community pharmacy/ or exp hospital pharmacy/ or (pharmacist* or clinical pharmacy or clinical pharmacist* or pharmaceutical care or pharmaceutical service* or pharmacy service* or medication therapy management or medication management or medication review* or medicines review* or medication reconciliation or pharmacist intervention* or pharmacist-led or pharmacy-led or community pharmacy service* or hospital pharmacy service*).ti,ab,kw.) and (exp hypertension/ or (hypertens* or "high blood pressure").ti,ab,kw.) and (exp China/ or exp Taiwan/ or Hong Kong/ or (China or Chinese or Taiwan or Hong Kong or Hongkong or Macau or Macao).ti,ab,kw.)**

#### Diabetes Mellitus

**(exp medication therapy management/ or exp pharmaceutical care/ or exp pharmacist/ or exp medication review/ or exp medication reconciliation/ or exp community pharmacy/ or exp hospital pharmacy/ or (pharmacist* or clinical pharmacy or clinical pharmacist* or pharmaceutical care or pharmaceutical service* or pharmacy service* or medication therapy management or medication management or medication review* or medicines review* or medication reconciliation or pharmacist intervention* or pharmacist-led or pharmacy-led or community pharmacy service* or hospital pharmacy service*).ti,ab,kw.) and (exp diabetes mellitus/ or (diabet* or hyperglycemi* or "high blood glucose").ti,ab,kw.) and (exp China/ or exp Taiwan/ or Hong Kong/ or (China or Chinese or Taiwan or Hong Kong or Hongkong or Macau or Macao).ti,ab,kw.)**

#### Coronary Disease

**(exp medication therapy management/ or exp pharmaceutical care/ or exp pharmacist/ or exp medication review/ or exp medication reconciliation/ or exp community pharmacy/ or exp hospital pharmacy/ or (pharmacist* or clinical pharmacy or clinical pharmacist* or pharmaceutical care or pharmaceutical service* or pharmacy service* or medication therapy management or medication management or medication review* or medicines review* or medication reconciliation or pharmacist intervention* or pharmacist-led or pharmacy-led or community pharmacy service* or hospital pharmacy service*).ti,ab,kw.) and (exp coronary artery disease/ or ("coronary disease" or "coronary artery disease" or "coronary heart disease" or "ischemic heart disease" or "ischaemic heart disease").ti,ab,kw.) and (exp China/ or exp Taiwan/ or Hong Kong/ or (China or Chinese or Taiwan or Hong Kong or Hongkong or Macau or Macao).ti,ab,kw.)**

#### Dyslipidemia

**(exp medication therapy management/ or exp pharmaceutical care/ or exp pharmacist/ or exp medication review/ or exp medication reconciliation/ or exp community pharmacy/ or exp hospital pharmacy/ or (pharmacist* or clinical pharmacy or clinical pharmacist* or pharmaceutical care or pharmaceutical service* or pharmacy service* or medication therapy management or medication management or medication review* or medicines review* or medication reconciliation or pharmacist intervention* or pharmacist-led or pharmacy-led or community pharmacy service* or hospital pharmacy service*).ti,ab,kw.) and (exp dyslipidemia/ or (dyslipid* or hyperlipid* or hypercholester* or hypertriglycerid* or hyperlipemi*).ti,ab,kw.) and (exp China/ or exp Taiwan/ or Hong Kong/ or (China or Chinese or Taiwan or Hong Kong or Hongkong or Macau or Macao).ti,ab,kw.)**

#### Heart Failure

**(exp medication therapy management/ or exp pharmaceutical care/ or exp pharmacist/ or exp medication review/ or exp medication reconciliation/ or exp community pharmacy/ or exp hospital pharmacy/ or (pharmacist* or clinical pharmacy or clinical pharmacist* or pharmaceutical care or pharmaceutical service* or pharmacy service* or medication therapy management or medication management or medication review* or medicines review* or medication reconciliation or pharmacist intervention* or pharmacist-led or pharmacy-led or community pharmacy service* or hospital pharmacy service*).ti,ab,kw.) and (exp heart failure/ or ("heart failure" or "cardiac failure" or "ventricular failure").ti,ab,kw.) and (exp China/ or exp Taiwan/ or Hong Kong/ or (China or Chinese or Taiwan or Hong Kong or Hongkong or Macau or Macao).ti,ab,kw.)**

#### Myocardial Infarction

**(exp medication therapy management/ or exp pharmaceutical care/ or exp pharmacist/ or exp medication review/ or exp medication reconciliation/ or exp community pharmacy/ or exp hospital pharmacy/ or (pharmacist* or clinical pharmacy or clinical pharmacist* or pharmaceutical care or pharmaceutical service* or pharmacy service* or medication therapy management or medication management or medication review* or medicines review* or medication reconciliation or pharmacist intervention* or pharmacist-led or pharmacy-led or community pharmacy service* or hospital pharmacy service*).ti,ab,kw.) and (exp myocardial infarction/ or ("myocardial infarction" or "myocardial infarct" or "heart attack").ti,ab,kw.) and (exp China/ or exp Taiwan/ or Hong Kong/ or (China or Chinese or Taiwan or Hong Kong or Hongkong or Macau or Macao).ti,ab,kw.)**

### Cochrane Library

The Cochrane Library/Cochrane Central Register of Controlled Trials (CENTRAL) base strategy is reproduced without an added study-design filter; CENTRAL functions as a trials source, but no RCT module is appended here.

#### Hypertension

**([mh "Medication Therapy Management"] OR [mh "Pharmaceutical Services"] OR [mh "Pharmacists"] OR [mh "Medication Reconciliation"] OR [mh "Community Pharmacy Services"] OR [mh "Pharmacy Service, Hospital"] OR (pharmacist* OR "clinical pharmacy" OR "clinical pharmacist" OR "clinical pharmacists" OR "pharmaceutical care" OR "pharmaceutical service" OR "pharmaceutical services" OR "pharmacy service" OR "pharmacy services" OR "medication therapy management" OR "medication management" OR "medication review" OR "medication reviews" OR "medicines review" OR "medicines reviews" OR "medication reconciliation" OR "pharmacist intervention" OR "pharmacist interventions" OR "pharmacist led" OR "pharmacy led" OR "community pharmacy service" OR "community pharmacy services" OR "hospital pharmacy service" OR "hospital pharmacy services"):ti,ab,kw) AND ([mh "Hypertension"] OR (hypertens* OR "high blood pressure"):ti,ab,kw) AND ([mh "China"] OR [mh "Taiwan"] OR (China OR Chinese OR Taiwan OR "Hong Kong" OR Hongkong OR Macau OR Macao):ti,ab,kw)**

#### Diabetes Mellitus

**([mh "Medication Therapy Management"] OR [mh "Pharmaceutical Services"] OR [mh "Pharmacists"] OR [mh "Medication Reconciliation"] OR [mh "Community Pharmacy Services"] OR [mh "Pharmacy Service, Hospital"] OR (pharmacist* OR "clinical pharmacy" OR "clinical pharmacist" OR "clinical pharmacists" OR "pharmaceutical care" OR "pharmaceutical service" OR "pharmaceutical services" OR "pharmacy service" OR "pharmacy services" OR "medication therapy management" OR "medication management" OR "medication review" OR "medication reviews" OR "medicines review" OR "medicines reviews" OR "medication reconciliation" OR "pharmacist intervention" OR "pharmacist interventions" OR "pharmacist led" OR "pharmacy led" OR "community pharmacy service" OR "community pharmacy services" OR "hospital pharmacy service" OR "hospital pharmacy services"):ti,ab,kw) AND ([mh "Diabetes Mellitus"] OR (diabet* OR hyperglycemi* OR "high blood glucose"):ti,ab,kw) AND ([mh "China"] OR [mh "Taiwan"] OR (China OR Chinese OR Taiwan OR "Hong Kong" OR Hongkong OR Macau OR Macao):ti,ab,kw)**

#### Coronary Disease

**([mh "Medication Therapy Management"] OR [mh "Pharmaceutical Services"] OR [mh "Pharmacists"] OR [mh "Medication Reconciliation"] OR [mh "Community Pharmacy Services"] OR [mh "Pharmacy Service, Hospital"] OR (pharmacist* OR "clinical pharmacy" OR "clinical pharmacist" OR "clinical pharmacists" OR "pharmaceutical care" OR "pharmaceutical service" OR "pharmaceutical services" OR "pharmacy service" OR "pharmacy services" OR "medication therapy management" OR "medication management" OR "medication review" OR "medication reviews" OR "medicines review" OR "medicines reviews" OR "medication reconciliation" OR "pharmacist intervention" OR "pharmacist interventions" OR "pharmacist led" OR "pharmacy led" OR "community pharmacy service" OR "community pharmacy services" OR "hospital pharmacy service" OR "hospital pharmacy services"):ti,ab,kw) AND ([mh "Coronary Disease"] OR [mh "Coronary Artery Disease"] OR ("coronary disease" OR "coronary artery disease" OR "coronary heart disease" OR "ischemic heart disease" OR "ischaemic heart disease"):ti,ab,kw) AND ([mh "China"] OR [mh "Taiwan"] OR (China OR Chinese OR Taiwan OR "Hong Kong" OR Hongkong OR Macau OR Macao):ti,ab,kw)**

#### Dyslipidemia

**([mh "Medication Therapy Management"] OR [mh "Pharmaceutical Services"] OR [mh "Pharmacists"] OR [mh "Medication Reconciliation"] OR [mh "Community Pharmacy Services"] OR [mh "Pharmacy Service, Hospital"] OR (pharmacist* OR "clinical pharmacy" OR "clinical pharmacist" OR "clinical pharmacists" OR "pharmaceutical care" OR "pharmaceutical service" OR "pharmaceutical services" OR "pharmacy service" OR "pharmacy services" OR "medication therapy management" OR "medication management" OR "medication review" OR "medication reviews" OR "medicines review" OR "medicines reviews" OR "medication reconciliation" OR "pharmacist intervention" OR "pharmacist interventions" OR "pharmacist led" OR "pharmacy led" OR "community pharmacy service" OR "community pharmacy services" OR "hospital pharmacy service" OR "hospital pharmacy services"):ti,ab,kw) AND ([mh "Dyslipidemias"] OR (dyslipid* OR hyperlipid* OR hypercholester* OR hypertriglycerid* OR hyperlipemi*):ti,ab,kw) AND ([mh "China"] OR [mh "Taiwan"] OR (China OR Chinese OR Taiwan OR "Hong Kong" OR Hongkong OR Macau OR Macao):ti,ab,kw)**

#### Heart Failure

**([mh "Medication Therapy Management"] OR [mh "Pharmaceutical Services"] OR [mh "Pharmacists"] OR [mh "Medication Reconciliation"] OR [mh "Community Pharmacy Services"] OR [mh "Pharmacy Service, Hospital"] OR (pharmacist* OR "clinical pharmacy" OR "clinical pharmacist" OR "clinical pharmacists" OR "pharmaceutical care" OR "pharmaceutical service" OR "pharmaceutical services" OR "pharmacy service" OR "pharmacy services" OR "medication therapy management" OR "medication management" OR "medication review" OR "medication reviews" OR "medicines review" OR "medicines reviews" OR "medication reconciliation" OR "pharmacist intervention" OR "pharmacist interventions" OR "pharmacist led" OR "pharmacy led" OR "community pharmacy service" OR "community pharmacy services" OR "hospital pharmacy service" OR "hospital pharmacy services"):ti,ab,kw) AND ([mh "Heart Failure"] OR ("heart failure" OR "cardiac failure" OR "ventricular failure"):ti,ab,kw) AND ([mh "China"] OR [mh "Taiwan"] OR (China OR Chinese OR Taiwan OR "Hong Kong" OR Hongkong OR Macau OR Macao):ti,ab,kw)**

#### Myocardial Infarction

**([mh "Medication Therapy Management"] OR [mh "Pharmaceutical Services"] OR [mh "Pharmacists"] OR [mh "Medication Reconciliation"] OR [mh "Community Pharmacy Services"] OR [mh "Pharmacy Service, Hospital"] OR (pharmacist* OR "clinical pharmacy" OR "clinical pharmacist" OR "clinical pharmacists" OR "pharmaceutical care" OR "pharmaceutical service" OR "pharmaceutical services" OR "pharmacy service" OR "pharmacy services" OR "medication therapy management" OR "medication management" OR "medication review" OR "medication reviews" OR "medicines review" OR "medicines reviews" OR "medication reconciliation" OR "pharmacist intervention" OR "pharmacist interventions" OR "pharmacist led" OR "pharmacy led" OR "community pharmacy service" OR "community pharmacy services" OR "hospital pharmacy service" OR "hospital pharmacy services"):ti,ab,kw) AND ([mh "Myocardial Infarction"] OR ("myocardial infarction" OR "myocardial infarct" OR "heart attack"):ti,ab,kw) AND ([mh "China"] OR [mh "Taiwan"] OR (China OR Chinese OR Taiwan OR "Hong Kong" OR Hongkong OR Macau OR Macao):ti,ab,kw)**

### CNKI

Use Professional/Advanced Search. SU denotes the subject field. The default strategy omits a China term to avoid excluding Chinese studies that do not state the country in the subject metadata.

#### Hypertension

(SU='药学服务' OR SU='药学干预' OR SU='药学监护' OR SU='药物治疗管理' OR SU='药物治疗管理服务' OR SU='用药管理' OR SU='用药教育' OR SU='用药指导' OR SU='用药咨询' OR SU='用药重整' OR SU='药物重整' OR SU='药师干预' OR SU='临床药师' OR SU='社区药师' OR SU='家庭药师' OR SU='药师主导' OR SU='药师参与' OR SU='处方审核') AND (SU='高血压' OR SU='高血压病' OR SU='原发性高血压')

#### Diabetes Mellitus

(SU='药学服务' OR SU='药学干预' OR SU='药学监护' OR SU='药物治疗管理' OR SU='药物治疗管理服务' OR SU='用药管理' OR SU='用药教育' OR SU='用药指导' OR SU='用药咨询' OR SU='用药重整' OR SU='药物重整' OR SU='药师干预' OR SU='临床药师' OR SU='社区药师' OR SU='家庭药师' OR SU='药师主导' OR SU='药师参与' OR SU='处方审核') AND (SU='糖尿病' OR SU='2型糖尿病' OR SU='Ⅱ型糖尿病' OR SU='II型糖尿病')

#### Coronary Disease

(SU='药学服务' OR SU='药学干预' OR SU='药学监护' OR SU='药物治疗管理' OR SU='药物治疗管理服务' OR SU='用药管理' OR SU='用药教育' OR SU='用药指导' OR SU='用药咨询' OR SU='用药重整' OR SU='药物重整' OR SU='药师干预' OR SU='临床药师' OR SU='社区药师' OR SU='家庭药师' OR SU='药师主导' OR SU='药师参与' OR SU='处方审核') AND (SU='冠心病' OR SU='冠状动脉粥样硬化性心脏病' OR SU='冠状动脉疾病' OR SU='缺血性心脏病')

#### Dyslipidemia

(SU='药学服务' OR SU='药学干预' OR SU='药学监护' OR SU='药物治疗管理' OR SU='药物治疗管理服务' OR SU='用药管理' OR SU='用药教育' OR SU='用药指导' OR SU='用药咨询' OR SU='用药重整' OR SU='药物重整' OR SU='药师干预' OR SU='临床药师' OR SU='社区药师' OR SU='家庭药师' OR SU='药师主导' OR SU='药师参与' OR SU='处方审核') AND (SU='血脂异常' OR SU='高脂血症' OR SU='高胆固醇血症' OR SU='高甘油三酯血症' OR SU='高脂蛋白血症')

#### Heart Failure

(SU='药学服务' OR SU='药学干预' OR SU='药学监护' OR SU='药物治疗管理' OR SU='药物治疗管理服务' OR SU='用药管理' OR SU='用药教育' OR SU='用药指导' OR SU='用药咨询' OR SU='用药重整' OR SU='药物重整' OR SU='药师干预' OR SU='临床药师' OR SU='社区药师' OR SU='家庭药师' OR SU='药师主导' OR SU='药师参与' OR SU='处方审核') AND (SU='心力衰竭' OR SU='心衰' OR SU='慢性心力衰竭' OR SU='急性心力衰竭' OR SU='充血性心力衰竭')

#### Myocardial Infarction

(SU='药学服务' OR SU='药学干预' OR SU='药学监护' OR SU='药物治疗管理' OR SU='药物治疗管理服务' OR SU='用药管理' OR SU='用药教育' OR SU='用药指导' OR SU='用药咨询' OR SU='用药重整' OR SU='药物重整' OR SU='药师干预' OR SU='临床药师' OR SU='社区药师' OR SU='家庭药师' OR SU='药师主导' OR SU='药师参与' OR SU='处方审核') AND (SU='心肌梗死' OR SU='心肌梗塞' OR SU='急性心肌梗死' OR SU='急性心肌梗塞')

### Wanfang Data

Use Advanced Search. Chinese service and disease synonyms are searched in the subject field.

#### Hypertension

主题:("药学服务" OR "药学干预" OR "药学监护" OR "药物治疗管理" OR "药物治疗管理服务" OR "用药管理" OR "用药教育" OR "用药指导" OR "用药咨询" OR "用药重整" OR "药物重整" OR "药师干预" OR "临床药师" OR "社区药师" OR "家庭药师" OR "药师主导" OR "药师参与" OR "处方审核") AND 主题:("高血压" OR "高血压病" OR "原发性高血压")

#### Diabetes Mellitus

主题:("药学服务" OR "药学干预" OR "药学监护" OR "药物治疗管理" OR "药物治疗管理服务" OR "用药管理" OR "用药教育" OR "用药指导" OR "用药咨询" OR "用药重整" OR "药物重整" OR "药师干预" OR "临床药师" OR "社区药师" OR "家庭药师" OR "药师主导" OR "药师参与" OR "处方审核") AND 主题:("糖尿病" OR "2型糖尿病" OR "Ⅱ型糖尿病" OR "II型糖尿病")

#### Coronary Disease

主题:("药学服务" OR "药学干预" OR "药学监护" OR "药物治疗管理" OR "药物治疗管理服务" OR "用药管理" OR "用药教育" OR "用药指导" OR "用药咨询" OR "用药重整" OR "药物重整" OR "药师干预" OR "临床药师" OR "社区药师" OR "家庭药师" OR "药师主导" OR "药师参与" OR "处方审核") AND 主题:("冠心病" OR "冠状动脉粥样硬化性心脏病" OR "冠状动脉疾病" OR "缺血性心脏病")

#### Dyslipidemia

主题:("药学服务" OR "药学干预" OR "药学监护" OR "药物治疗管理" OR "药物治疗管理服务" OR "用药管理" OR "用药教育" OR "用药指导" OR "用药咨询" OR "用药重整" OR "药物重整" OR "药师干预" OR "临床药师" OR "社区药师" OR "家庭药师" OR "药师主导" OR "药师参与" OR "处方审核") AND 主题:("血脂异常" OR "高脂血症" OR "高胆固醇血症" OR "高甘油三酯血症" OR "高脂蛋白血症")

#### Heart Failure

主题:("药学服务" OR "药学干预" OR "药学监护" OR "药物治疗管理" OR "药物治疗管理服务" OR "用药管理" OR "用药教育" OR "用药指导" OR "用药咨询" OR "用药重整" OR "药物重整" OR "药师干预" OR "临床药师" OR "社区药师" OR "家庭药师" OR "药师主导" OR "药师参与" OR "处方审核") AND 主题:("心力衰竭" OR "心衰" OR "慢性心力衰竭" OR "急性心力衰竭" OR "充血性心力衰竭")

#### Myocardial Infarction

主题:("药学服务" OR "药学干预" OR "药学监护" OR "药物治疗管理" OR "药物治疗管理服务" OR "用药管理" OR "用药教育" OR "用药指导" OR "用药咨询" OR "用药重整" OR "药物重整" OR "药师干预" OR "临床药师" OR "社区药师" OR "家庭药师" OR "药师主导" OR "药师参与" OR "处方审核") AND 主题:("心肌梗死" OR "心肌梗塞" OR "急性心肌梗死" OR "急性心肌梗塞")

### SinoMed

Use the Chinese advanced-search interface. The "Smart Common Fields" option searches title, abstract, keywords, and subject headings with synonym expansion. Parentheses are explicit to preserve Boolean precedence.

#### Hypertension

(("药学服务"[常用字段:智能] OR "药学干预"[常用字段:智能] OR "药学监护"[常用字段:智能] OR "药物治疗管理"[常用字段:智能] OR "药物治疗管理服务"[常用字段:智能] OR "用药管理"[常用字段:智能] OR "用药教育"[常用字段:智能] OR "用药指导"[常用字段:智能] OR "用药咨询"[常用字段:智能] OR "用药重整"[常用字段:智能] OR "药物重整"[常用字段:智能] OR "药师干预"[常用字段:智能] OR "临床药师"[常用字段:智能] OR "社区药师"[常用字段:智能] OR "家庭药师"[常用字段:智能] OR "药师主导"[常用字段:智能] OR "药师参与"[常用字段:智能] OR "处方审核"[常用字段:智能]) AND ("高血压"[常用字段:智能] OR "高血压病"[常用字段:智能] OR "原发性高血压"[常用字段:智能])) AND ("1"[期刊类型] OR "2"[期刊类型])

#### Diabetes Mellitus

(("药学服务"[常用字段:智能] OR "药学干预"[常用字段:智能] OR "药学监护"[常用字段:智能] OR "药物治疗管理"[常用字段:智能] OR "药物治疗管理服务"[常用字段:智能] OR "用药管理"[常用字段:智能] OR "用药教育"[常用字段:智能] OR "用药指导"[常用字段:智能] OR "用药咨询"[常用字段:智能] OR "用药重整"[常用字段:智能] OR "药物重整"[常用字段:智能] OR "药师干预"[常用字段:智能] OR "临床药师"[常用字段:智能] OR "社区药师"[常用字段:智能] OR "家庭药师"[常用字段:智能] OR "药师主导"[常用字段:智能] OR "药师参与"[常用字段:智能] OR "处方审核"[常用字段:智能]) AND ("糖尿病"[常用字段:智能] OR "2型糖尿病"[常用字段:智能] OR "Ⅱ型糖尿病"[常用字段:智能] OR "II型糖尿病"[常用字段:智能])) AND ("1"[期刊类型] OR "2"[期刊类型])

#### Coronary Disease

(("药学服务"[常用字段:智能] OR "药学干预"[常用字段:智能] OR "药学监护"[常用字段:智能] OR "药物治疗管理"[常用字段:智能] OR "药物治疗管理服务"[常用字段:智能] OR "用药管理"[常用字段:智能] OR "用药教育"[常用字段:智能] OR "用药指导"[常用字段:智能] OR "用药咨询"[常用字段:智能] OR "用药重整"[常用字段:智能] OR "药物重整"[常用字段:智能] OR "药师干预"[常用字段:智能] OR "临床药师"[常用字段:智能] OR "社区药师"[常用字段:智能] OR "家庭药师"[常用字段:智能] OR "药师主导"[常用字段:智能] OR "药师参与"[常用字段:智能] OR "处方审核"[常用字段:智能]) AND ("冠心病"[常用字段:智能] OR "冠状动脉粥样硬化性心脏病"[常用字段:智能] OR "冠状动脉疾病"[常用字段:智能] OR "缺血性心脏病"[常用字段:智能])) AND ("1"[期刊类型] OR "2"[期刊类型])

#### Dyslipidemia

(("药学服务"[常用字段:智能] OR "药学干预"[常用字段:智能] OR "药学监护"[常用字段:智能] OR "药物治疗管理"[常用字段:智能] OR "药物治疗管理服务"[常用字段:智能] OR "用药管理"[常用字段:智能] OR "用药教育"[常用字段:智能] OR "用药指导"[常用字段:智能] OR "用药咨询"[常用字段:智能] OR "用药重整"[常用字段:智能] OR "药物重整"[常用字段:智能] OR "药师干预"[常用字段:智能] OR "临床药师"[常用字段:智能] OR "社区药师"[常用字段:智能] OR "家庭药师"[常用字段:智能] OR "药师主导"[常用字段:智能] OR "药师参与"[常用字段:智能] OR "处方审核"[常用字段:智能]) AND ("血脂异常"[常用字段:智能] OR "高脂血症"[常用字段:智能] OR "高胆固醇血症"[常用字段:智能] OR "高甘油三酯血症"[常用字段:智能] OR "高脂蛋白血症"[常用字段:智能])) AND ("1"[期刊类型] OR "2"[期刊类型])

#### Heart Failure

(("药学服务"[常用字段:智能] OR "药学干预"[常用字段:智能] OR "药学监护"[常用字段:智能] OR "药物治疗管理"[常用字段:智能] OR "药物治疗管理服务"[常用字段:智能] OR "用药管理"[常用字段:智能] OR "用药教育"[常用字段:智能] OR "用药指导"[常用字段:智能] OR "用药咨询"[常用字段:智能] OR "用药重整"[常用字段:智能] OR "药物重整"[常用字段:智能] OR "药师干预"[常用字段:智能] OR "临床药师"[常用字段:智能] OR "社区药师"[常用字段:智能] OR "家庭药师"[常用字段:智能] OR "药师主导"[常用字段:智能] OR "药师参与"[常用字段:智能] OR "处方审核"[常用字段:智能]) AND ("心力衰竭"[常用字段:智能] OR "心衰"[常用字段:智能] OR "慢性心力衰竭"[常用字段:智能] OR "急性心力衰竭"[常用字段:智能] OR "充血性心力衰竭"[常用字段:智能])) AND ("1"[期刊类型] OR "2"[期刊类型])

#### Myocardial Infarction

(("药学服务"[常用字段:智能] OR "药学干预"[常用字段:智能] OR "药学监护"[常用字段:智能] OR "药物治疗管理"[常用字段:智能] OR "药物治疗管理服务"[常用字段:智能] OR "用药管理"[常用字段:智能] OR "用药教育"[常用字段:智能] OR "用药指导"[常用字段:智能] OR "用药咨询"[常用字段:智能] OR "用药重整"[常用字段:智能] OR "药物重整"[常用字段:智能] OR "药师干预"[常用字段:智能] OR "临床药师"[常用字段:智能] OR "社区药师"[常用字段:智能] OR "家庭药师"[常用字段:智能] OR "药师主导"[常用字段:智能] OR "药师参与"[常用字段:智能] OR "处方审核"[常用字段:智能]) AND ("心肌梗死"[常用字段:智能] OR "心肌梗塞"[常用字段:智能] OR "急性心肌梗死"[常用字段:智能] OR "急性心肌梗塞"[常用字段:智能])) AND ("1"[期刊类型] OR "2"[期刊类型])

### Platform and syntax notes

| **Database** | **Implementation note** | **Reference** |
| --- | --- | --- |
| PubMed | MeSH headings are paired with [Title/Abstract] keywords. Field tags turn off automatic term mapping for the tagged term, so synonyms are stated explicitly. | https://pubmed.ncbi.nlm.nih.gov/help/ |
| Ovid Embase | The strategy assumes Ovid Embase, using exploded Emtree headings (/exp) and title/abstract/keyword fields (.ti,ab,kw.). | https://tools.ovid.com/embase/nazarbayev/ |
| Cochrane Library | The strategy uses Search Manager syntax with [mh "..."] and :ti,ab,kw. CENTRAL is already a controlled-trials register. | https://www.cochranelibrary.com/advanced-search/search-manager |
| SinoMed | 常用字段 includes title, abstract, keywords, and subject headings; 智能检索 expands synonyms and controlled terms. | https://www.sinomed.ac.cn/help/content/2.1.3.html |

### Appendix 2 | PRISMA 2020 checklist

| **Section and Topic** | **Item #** | **Checklist item** | **Location where item is reported** |
| --- | --- | --- | --- |
| TITLE | TITLE | TITLE |  |
| Title | 1 | Identify the report as a systematic review. | Title page |
| ABSTRACT | ABSTRACT | ABSTRACT |  |
| Abstract | 2 | See the PRISMA 2020 for Abstracts checklist. | Abstract |
| INTRODUCTION | INTRODUCTION | INTRODUCTION |  |
| Rationale | 3 | Describe the rationale for the review in the context of existing knowledge. | Introduction |
| Objectives | 4 | Provide an explicit statement of the objective(s) or question(s) the review addresses. | Introduction, final paragraph |
| METHODS | METHODS | METHODS |  |
| Eligibility criteria | 5 | Specify the inclusion and exclusion criteria for the review and how studies were grouped for the syntheses. | Methods: Search strategy and eligibility |
| Information sources | 6 | Specify all databases, registers, websites, organisations, reference lists and other sources searched or consulted to identify studies. Specify the date when each source was last searched or consulted. | Methods: Search strategy; Appendix 1 |
| Search strategy | 7 | Present the full search strategies for all databases, registers and websites, including any filters and limits used. | Appendix 1 |
| Selection process | 8 | Specify the methods used to decide whether a study met the inclusion criteria of the review, including how many reviewers screened each record and each report retrieved, whether they worked independently, and if applicable, details of automation tools used in the process. | Methods: Study selection, extraction, and classification |
| Data collection process | 9 | Specify the methods used to collect data from reports, including how many reviewers collected data from each report, whether they worked independently, any processes for obtaining or confirming data from study investigators, and if applicable, details of automation tools used in the process. | Methods: Study selection, extraction, and classification |
| Data items | 10a | List and define all outcomes for which data were sought. Specify whether all results that were compatible with each outcome domain in each study were sought (e.g. for all measures, time points, analyses), and if not, the methods used to decide which results to collect. | Methods: Outcomes and intervention reporting |
| Data items | 10b | List and define all other variables for which data were sought (e.g. participant and intervention characteristics, funding sources). Describe any assumptions made about any missing or unclear information. | Methods: Study selection, extraction, and classification |
| Study risk of bias assessment | 11 | Specify the methods used to assess risk of bias in the included studies, including details of the tool(s) used, how many reviewers assessed each study and whether they worked independently, and if applicable, details of automation tools used in the process. | Methods: Risk of bias and certainty |
| Effect measures | 12 | Specify for each outcome the effect measure(s) (e.g. risk ratio, mean difference) used in the synthesis or presentation of results. | Methods: Statistical analysis |
| Synthesis methods | 13a | Describe the processes used to decide which studies were eligible for each synthesis (e.g. tabulating the study intervention characteristics and comparing against the planned groups for each synthesis (item #5)). | Methods: Outcomes and intervention reporting |
| Synthesis methods | 13b | Describe any methods required to prepare the data for presentation or synthesis, such as handling of missing summary statistics, or data conversions. | Methods: Statistical analysis and expenditure analysis |
| Synthesis methods | 13c | Describe any methods used to tabulate or visually display results of individual studies and syntheses. | Main Tables 2-5 and Figures 2-6; Supplementary Figures S5-S27 |
| Synthesis methods | 13d | Describe any methods used to synthesize results and provide a rationale for the choice(s). If meta-analysis was performed, describe the model(s), method(s) to identify the presence and extent of statistical heterogeneity, and software package(s) used. | Methods: Statistical analysis; Supplementary Methods: Software environment |
| Synthesis methods | 13e | Describe any methods used to explore possible causes of heterogeneity among study results (e.g. subgroup analysis, meta-regression). | Methods: Statistical analysis; Outcome-specific and robustness analyses |
| Synthesis methods | 13f | Describe any sensitivity analyses conducted to assess robustness of the synthesized results. | Methods: Statistical analysis; Supplementary Tables S5-S7 and S12A-S13A; Supplementary Figures S1 and S4 |
| Reporting bias assessment | 14 | Describe any methods used to assess risk of bias due to missing results in a synthesis (arising from reporting biases). | Methods: Outcome-specific and robustness analyses; Supplementary Table S11C |
| Certainty assessment | 15 | Describe any methods used to assess certainty (or confidence) in the body of evidence for an outcome. | Methods: Risk of bias and certainty |
| RESULTS | RESULTS | RESULTS |  |
| Study selection | 16a | Describe the results of the search and selection process, from the number of records identified in the search to the number of studies included in the review, ideally using a flow diagram. | Results; Figure 1 |
| Study selection | 16b | Cite studies that might appear to meet the inclusion criteria, but which were excluded, and explain why they were excluded. | Not reported; a study-level list of apparently eligible but excluded studies is not provided. |
| Study characteristics | 17 | Cite each included study and present its characteristics. | Results; Table 1; Supplementary Table S8A |
| Risk of bias in studies | 18 | Present assessments of risk of bias for each included study. | Results; Supplementary Tables S10A-S10B |
| Results of individual studies | 19 | For all outcomes, present, for each study: (a) summary statistics for each group (where appropriate) and (b) an effect estimate and its precision (e.g. confidence/credible interval), ideally using structured tables or plots. | Main Figure 2; Supplementary Figures S5-S24; source-data files |
| Results of syntheses | 20a | For each synthesis, briefly summarise the characteristics and risk of bias among contributing studies. | Results; Table 1; Supplementary Tables S3, S6A, and S10A-S10B |
| Results of syntheses | 20b | Present results of all statistical syntheses conducted. If meta-analysis was done, present for each the summary estimate and its precision (e.g. confidence/credible interval) and measures of statistical heterogeneity. If comparing groups, describe the direction of the effect. | Results; Table 2 |
| Results of syntheses | 20c | Present results of all investigations of possible causes of heterogeneity among study results. | Results; Supplementary Tables S2A-S2B, S11A-S11B, S11E, and S12A-S12B; Supplementary Figures S2-S4 |
| Results of syntheses | 20d | Present results of all sensitivity analyses conducted to assess the robustness of the synthesized results. | Results; Table 3; Supplementary Tables S5-S7, S12A-S13A, and S21A; Supplementary Figures S1 and S4 |
| Reporting biases | 21 | Present assessments of risk of bias due to missing results (arising from reporting biases) for each synthesis assessed. | Results; Supplementary Tables S11C and S21B; Supplementary Figures S25-S27 |
| Certainty of evidence | 22 | Present assessments of certainty (or confidence) in the body of evidence for each outcome assessed. | Results; Table 4; Supplementary Table S3 |
| DISCUSSION | DISCUSSION | DISCUSSION |  |
| Discussion | 23a | Provide a general interpretation of the results in the context of other evidence. | Discussion |
| Discussion | 23b | Discuss any limitations of the evidence included in the review. | Discussion |
| Discussion | 23c | Discuss any limitations of the review processes used. | Discussion |
| Discussion | 23d | Discuss implications of the results for practice, policy, and future research. | Discussion; final paragraphs |
| OTHER INFORMATION | OTHER INFORMATION | OTHER INFORMATION |  |
| Registration and protocol | 24a | Provide registration information for the review, including register name and registration number, or state that the review was not registered. | Title page and Methods: Protocol and reporting |
| Registration and protocol | 24b | Indicate where the review protocol can be accessed, or state that a protocol was not prepared. | PROSPERO CRD42023414180 |
| Registration and protocol | 24c | Describe and explain any amendments to information provided at registration or in the protocol. | Methods: Protocol and reporting; Supplementary Table S16 |
| Support | 25 | Describe sources of financial or non-financial support for the review, and the role of the funders or sponsors in the review. | Role of the funding source |
| Competing interests | 26 | Declare any competing interests of review authors. | Declaration of interests |
| Availability of data, code and other materials | 27 | Report which of the following are publicly available and where they can be found: template data collection forms; data extracted from included studies; data used for all analyses; analytic code; any other materials used in the review. | Data sharing |

From: Page MJ, McKenzie JE, Bossuyt PM, Boutron I, Hoffmann TC, Mulrow CD, et al. The PRISMA 2020 statement: an updated guideline for reporting systematic reviews. BMJ 2021;372:n71. doi: 10.1136/bmj.n71

For more information, visit: http://www.prisma-statement.org/
